# Computation and resource efficient genome-wide association analysis for large-scale imaging studies

**DOI:** 10.1101/2025.11.11.25340011

**Authors:** Zhiwen Jiang, Jason Stein, Tengfei Li, Ethan Fang, Yun Li, Patrick Sullivan, Hongtu Zhu

## Abstract

Imaging genetics links genetic variations to brain structures and functions, but the computational challenges posed by high-dimensional imaging and genetic data are significant. In voxel-level genome-wide association studies, we introduce a Representation learning-based Voxel-level Genetic Analysis (RVGA) framework that reduces computational time and storage burden by over 200 times. RVGA enhances statistical power by denoising images and shares minimal datasets of summary statistics for associations across the whole genome of the entire image for secondary analyses. Additionally, it introduces a unified estimator for voxel heritability, genetic correlations between voxels, and cross-trait genetic correlations between voxels and non-imaging phenotypes. Applying RVGA to hippocampus shape and white matter microstructure in the UK Biobank (n = 53,454) reveals 39 and 275 novel loci, respectively. We identify heterogeneity in genetic architecture across images and subregions that share genetic bases with 14 brain-related phenotypes, such as the genetic correlation between the hippocampus and educational attainment, and between the anterior corona radiata and schizophrenia. RVGA replicates known genetic associations and uncovers new discoveries.

## Introduction

Imaging genetics elucidates how genetic variations influence the human brain and links these variations to brain-related disorders such as Alzheimer’s disease [1–6]. This field enhances our understanding of the pathophysiological pathways underlying many brain disorders, potentially leading to more precise and personalized treatments. Modern neuroimaging technologies, such as functional magnetic resonance imaging (fMRI), enable us to capture the human brain at high resolution, with 10^4^-10^6^ measures per subject. The measure unit can be either voxel (3D volume) or vertex (surface). For simplicity, we use “voxel” to represent both units where no ambiguity arises. However, most existing imaging genome-wide association studies (GWAS) do not fully exploit the rich voxel-level imaging signals and are limited to a few hundred image-derived phenotypes (IDPs), which are generated by aggregating local signals, potentially discarding significant local variations [4, 7–13]. Recent studies have begun to address this issue by employing representation learning to capture essential information from the entire image [14–16] and applying the resulting low-dimensional representations (LDRs) in GWAS. However, challenges remain in interpreting the significant loci associated with LDR, reconstructing voxel-level results, and sharing GWAS summary statistics for a large number of IDPs.

Voxel-level GWAS (VGWAS) can potentially solve all the limitations, but it faces significant computationally challenges [17, 18]. First, high-resolution VGWAS generates vast amounts of data, requiring immense computational power and storage capacity, which leads to long processing time and substantial resource demands. Second, sharing VGWAS summary statistics at high resolution is particularly difficult, as their size can be 100 times larger than the NHGRI-EBI GWAS catalog, which contains over 70,000 GWAS datasets. This limits the practical utility and broad dissemination of informative datasets for downstream analyses. Third, correcting for multiple comparisons in hypothesis testing across highly correlated voxels is challenging [19–21]. Fourth, noise in imaging data can obscure meaningful genetic associations, reducing statistical power and reliability. This challenge is compounded by the need to accurately identify the genetic architectures underlying complex brain functions and structures, which requires sophisticated statistical methods and large-scale data integration. In contrast, current VGWAS methods, such as considering multiple genetic markers simultaneously [22], screening candidate markers [23, 24], or applying rank-reduction [25–27] and Bayesian techniques [28–30], often overlook the whole-genome summary statistics necessary for secondary analyses, such as heritability and genetic correlations.

We introduce the Representation learning-based Voxel-level Genetic Analysis (RVGA) framework to address the aforementioned challenges (Fig. 1). First, RVGA enhances both computational and statistical efficiency in VGWAS by decomposing raw images into smooth images and purely random noise, and constructing LDRs to capture essential imaging signals. Second, it proposes a storage-efficient system for sharing summary statistics, enabling all voxel-level secondary analyses. Third, RVGA introduces a unified estimator for voxel heritability, genetic correlations between voxels, and cross-trait genetic correlations between voxels and other phenotypes, using the shared data and a population-matched linkage disequilibrium (LD) matrix.

**Fig. 1.**
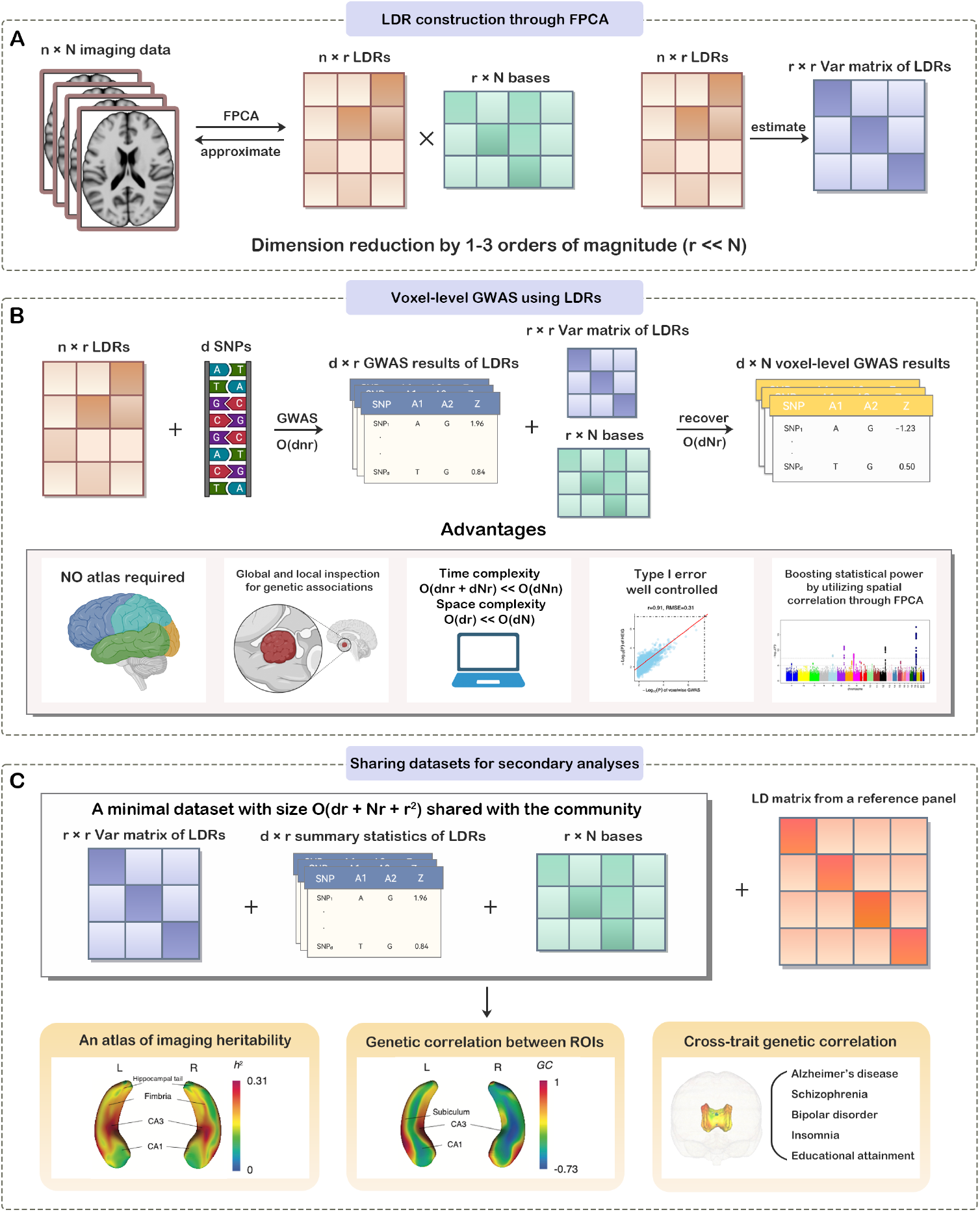
An overview of RVGA. **(A)** LDRs and functional bases are derived from imaging data using FPCA. The variance-covariance matrix of LDR is estimated by sample covariance. The number of LDRs (*r*) is 1-3 orders of magnitude less than the number of voxels (*N*) because of strong spatial correlation, resulting in substantial dimension reduction. **(B)** RVGA conducts VGWAS in two steps. First, LDRs are treated as traits to perform GWAS using external software. Next, VGWAS results are recovered by using the triplets: summary statistics of LDRs, the bases and the variance-covariance matrix of LDRs. Suppose *d* SNPs and *n* subjects in GWAS. The complexity is *O*(*dnr* + *dNr*) in time and *O*(*dr*) in space, which is drastically less than *O*(*dNn*) and *O*(*dN*) for scanning each voxel separately. Five advantages of RVGA over traditional approaches are highlighted. **(C)** Using the shared minimal dataset consisting of the triplets, as well as an LD matrix (and its inverse) estimated from a reference panel, RVGA generates an atlas of imaging heritability, estimates genetic correlations between ROIs, and investigates shared genetic bases between images and complex disorders through cross-trait genetic correlation analysis.

We analyzed hippocampus shape and white matter (WM) microstructure from 53,454 unrelated subjects of European ancestry in the UK Biobank (UKB), splitting into discovery (n = 33,324) and replication (n = 20,130). By performing GWAS on 7.8 million common variants across 30,000 hippocampus vertices and 32,217 WM voxels, we identified 39 and 275 novel loci, respectively. We reduced the total file size of summary statistics by 229 times (from 12,611 GB to 55 GB) and shared the data with community. Additionally, we generated atlases of heritability, genetic correlations within regions of interest (ROIs), and cross-trait genetic correlations between ROIs and 14 brain-related phenotypes. Novel genetic correlations were identified between WM tracts and schizophrenia, depression, bipolar disorder, and major depressive disorder. In summary, RVGA surpasses traditional voxel-level imaging genetic frameworks in computation and resource efficiency, facilitates the sharing of VGWAS summary statistics, and pinpoints genetic influences on the human brain and other organs.

## Results

### Overview of methods

We introduce RVGA, a framework to perform VGWAS and downstream analyses on large-scale imaging genetic datasets. RVGA uses functional principal component analysis (FPCA) to separate smooth imaging signals from white noise and to derive LDRs from images [31, 32]. These LDRs are used in GWAS to produce whole-genome summary statistics, from which RVGA reconstructs voxel-level summary statistics, creating a comprehensive atlas of genetic associations without the need to store voxel-level data. RVGA offers substantial time and storage savings by 1–3 orders of magnitude. This efficiency not only facilitates data sharing but also enables detailed examination of association patterns across ROIs, enhancing the investigation of genetic architecture at very high resolution.

Suppose we have biomedical images for *n* genetically unrelated subjects (no common ancestor for three generations). Let *Y*_*i*_(*v*) = *X*_*i*_(*v*) + *ϵ*_*i*_(*v*) represent the imaging data for subject *i*, with *N* common voxels indexed by *v*, where *X*_*i*_(*v*) are the underlying images and *ϵ*_*i*_(*v*) are independent white noise terms. The covariance function between two voxels, *u* and *v, C*(*u, v*) = Cov(*X*_*i*_(*u*), *X*_*i*_(*v*)), has a spectral decomposition 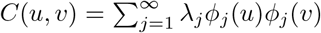, where *λ*_1_ ⩾ *λ*_2_ ⩾ · · · ⩾ 0 are the eigenvalues, and *ϕ*_1_, *ϕ*_2_, … are the corresponding functional bases.

To estimate *C*, we first smooth each raw image *Y*_*i*_(·) using a local linear estimator [33] with a Gaussian kernel. Each voxel is represented by a weighted average of itself and its neighboring voxels (Methods, Supplementary Note). We then perform spectral decomposition on the sample covariance of the smoothed images to obtain eigen pairs 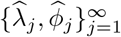. By extracting the top *r* bases, 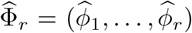, where 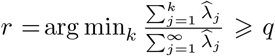 and *q* represents a proportion of variance (⩾ 80%), we project the raw image *Y*_*i*_(·) onto the functional subspace spanned by 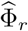, yielding LDRs as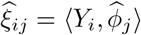. The underlying image can be approximated as 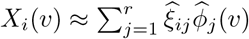. Thus, the dimensionality of the imaging data *Y* is reduced from *n* × *N* to *n* × *r*, with the LDRs 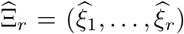, as shown in Fig. 1A. Due to the strong spatial correlation within images, the eigenvalues decay rapidly, making *r* 1–3 orders of magnitude smaller than *N*, while still preserving significant information from the original images (Fig. S1).

In GWAS, the marginal genetic effect of a continuous trait is estimated by projecting the trait vector onto the space spanned by the single nucleotide polymorphism (SNP) vector, assuming that all covariate effects have been removed. Similarly, in RVGA, the LDRs are treated as traits for GWAS analysis, projected onto each SNP vector, and the bases 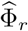 are used to recover the summary statistics for all voxels (Methods, Fig. 1B). With *d* SNPs, the computational burden of RVGA is only *O*(*dnr*) for LDR-level GWAS and *O*(*dNr*) for recovering VGWAS summary statistics. In contrast, directly performing VGWAS is computationally inefficient, with a complexity of *O*(*dNn*). In addition to improving computational efficiency, decomposing raw images into underlying smooth images and white noise via the FPCA framework enhances statistical power in GWAS.

Importantly, sharing the triplets—the summary statistics of LDRs, the functional bases, and the variance-covariance matrix of LDRs—is sufficient for all secondary analyses (Fig. 1C). This reduces the storage requirement from *O*(*dN*) for all VGWAS summary statistics to *O*(*dr* + *Nr* + *r*^2^). Next, we introduce a unified estimator to compute voxel SNP heritability, genetic correlations between voxels, and cross-trait genetic correlations with non-imaging phenotypes. Unlike regular definition of heritability, which is the ratio of genetic variance to phenotypic variance, Var(*Y* (·)), voxel heritability is defined as the ratio of genetic variance to the underlying image variance, Var(*X*(·)) (Methods). The estimator is accurate regardless of noise levels. Moreover, with variance analytically derived, it requires only the shared triplets and a populationmatched LD matrix (and its inverse). It is far more computationally and statistically efficient than current methods and does not impose assumptions about the distribution or genetic architecture of the effects, making it robust to model mis-specification.

### Simulation studies

Our simulation studies incorporated factors reflecting real imaging genetic studies: potential confounding effects in images, a polynomial decay rate of image eigenvalues (1.8 for slow decay, 2.5 for fast decay), with or without white noise (noise percentage = 50%, 20%, 0%), white noise following Gaussian or Rayleigh distribution, voxel heritability (3%, 10%, 30%), polygenicity (1%, 20% of causal variants), and data truncation levels (75%, 80%, up to 95% of variance preserved by the top LDRs). Refer to the Online Methods section for more details.

#### The relationship between number of LDRs and amount of preserved variance

We observed that the amount of variance preserved by the LDRs depends on the decay rate of eigenvalues and noise percentages, with more LDRs required for slower decay rates and lower noise percentages (Fig. S2). A slower decay rate indicates reduced spatial correlation, necessitating more LDRs. Noise-free images containing 100% of real imaging signals require more LDRs to capture local variations. Additionally, the number of LDRs remained stable across varying heritability and polygenicity.

#### The accuracy of RVGA GWAS summary statistics

We evaluated the accuracy of RVGA GWAS summary statistics compared to VGWAS, using the root-mean-squared-error (RMSE) of z-scores across all voxel-variant pairs. For noise-free images, RMSE of z-scores decreased almost linearly with an increasing number of LDRs (Fig. S3). For noisy images with Gaussian noise, the most significant RMSE improvement occurred when transitioning from preserving 95% of variance to using raw data (RMSE = 0), and the improvement increased with higher noise level. Additionally, noise increased RMSE at all truncation levels. For noisy images with Rayleigh noise, the pattern was similar to the Gaussian case (Fig. S4). Higher-heritability cases experienced larger bias, but the bias was generally unrelated to polygenicity. Taken together, the white noise is a major source of bias in summary statistics, and preserving less imaging signals slightly reduces accuracy.

#### The type I error rate and statistical power

We evaluated type I error by the proportion of null tests with *p*-value less than 10^*−*2^, 10^*−*3^, and 10^*−*4^. It was well controlled for RVGA regardless of noise percentage, eigenvalue decay rate, and the proportion of variance preserved (Figs. S5-S7).

We quantified statistical power by the proportion of causal variants identified at the significance level of 10^*−*4^. There was substantial power gain compared to doing GWAS on raw data for noisy images (Extended Data Fig. 1). The power for RVGA was unaffected by noise levels and eigenvalue decay rate. In contrast, doing GWAS on raw data was sensitive to noise. The power gain doesn’t mean no genetic signal is lost, but rather that the genetic signal loss is minor compared to the noise reduction. Another evidence was that the power of noisy images was similar to the power of noisefree images for RVGA. As expected, higher heritability and smaller polygenicity (i.e., larger effect size) yielded higher power. We observed slight power inflation in noise-free images with a slow eigenvalue decay rate, which is attributed to the relative smoothness between genetic and non-genetic effects. We also repeated the same investigation for images with Rayleigh noise, the pattern was similar to the Gaussian case (Fig. S8).

### The heritability and genetic correlation estimator

To investigate the robustness of the estimator to various genetic architectures, we varied the coupling strength between genetic effect sizes and minor allele frequency (MAF)/LD using the LDAK model [34]. We also considered different LD matrix types, using either imputed genotype data or genotype array data, with multiple regularization levels. For example, {90%, 85%} denotes preserving 90% of the variance in the LD matrix and 85% in its inverse, achieved by performing eigen-decomposition on each LD block.

The heritability estimator was robust to data truncation and polygenicity but sensitive to LD regularization under certain genetic architectures. Specifically, when effect sizes were strongly correlated with both MAF and LD (Extended Data Fig. 2B, Fig. S9), restrictive LD regularization led to large mean absolute error (MAE), especially in high-heritability scenarios. This effect was more pronounced in imputed genotype data due to more complex LD between variants. The estimator remained stable in other cases. Using {98%, 95%} for imputed genotype data (Extended Data Figs. 2 and 3) and {85%, 80%} for genotype array data (Figs. S9 and S10) produced unbiased estimates with low MAE, and is thus recommended for real data analysis.

We further assessed the impact of white noise on heritability estimation. Applying the estimator to each voxel without noise removal, we observed that noise-laden images resulted in significantly downward-biased estimates, while noise-free images maintained unbiased estimations (Extended Data Fig. 4, Fig. S11). However, the estimator was unbiased with noise removal as previously shown. This highlights that RVGA can preserve additive genetic effects while denoising images.

Genetic correlation estimates between voxels showed distinct patterns. Data truncation level and true heritability were the main factors affecting MAE, while polygenicity and genetic architecture had negligible effects (Extended Data Fig. 5, Fig. S12). Preserving more imaging signals improved MAE and reduced bias. LD regularization controlled the standard error of the estimates (Extended Data Fig. 6, Fig. S13). Similar patterns were observed for cross-trait genetic correlation estimates (Extended Data Figs. 7 and 8, Figs. S14 and S15), where we recommend using relatively restrictive LD regularization, such as {75%, 70%} for genotype array data and {90%, 85%} for imputed genotype data, to achieve smaller standard error.

We observed high consistency between the mean standard error and empirical standard error of the estimator (Fig. S16), validating the analytically derived variance.

### Voxel-level GWAS for brain imaging data in UKB

We applied RVGA to analyze hippocampus shape and WM microstructure from 53,454 unrelated European subjects in UKB (Table S1), and 33,324 of them were used in discovery. The shape feature, measured by radial distance from the medial model (i.e., a reference for evaluating shape) at each vertex, characterizes morphometric changes perpendicular to the surface. WM microstructure is assessed by fractional anisotropy (FA) for each voxel, indicating the restriction of water diffusion along WM tracts. The hippocampus shape is represented as a 3D mesh surface with 30,000 vertices, while the atlas-defined WM tract regions range in size from 88 voxels (inferior fronto-occipital fasciculus) to 3,503 voxels (superior longitudinal fasciculus), with a total of 32,217 voxels. To further demonstrate RVGA’s efficiency in whole-cerebral cortex analysis without relying on any atlas to segment into ROIs, we additionally analyzed cortical surface curvature with 59,412 vertices from 28,183 unrelated European subjects (Supplementary Note).

We adaptively generated LDRs for each WM tract and hippocampus in the left and right hemispheres (each with 15,000 vertices), extracting the top principal components (PCs) that contributed at least 80% of the variance (Methods). The eigenvalues of the hippocampus showed a steeper decay compared to those of WM tracts (Fig. S17). We constructed 49 LDRs (0.16% of 30,000) to capture 90% of hippocampal signals, whereas 1,034 LDRs (3.2% of 32,217) were needed for WM tracts to preserve 80% of the signals (Table S2). To show that the LDRs captured most of imaging signals, we reconstructed images using the LDRs and bases, calculating a correlation of 0.97 (*sd* = 0.03) for the hippocampus and 0.88 (*sd* = 0.03) for WM tracts with the raw images.

Using imputed genotype data of 7.8 million SNPs (MAF *>* 0.01), we conducted GWAS on the LDRs. The shared summary statistics for in total 1,083 LDRs were 55 GB after processing (Methods), reducing storage burden by 229 times compared to 12,611 GB required to store 62,217 GWAS datasets (the total number of voxels and vertices analyzed) in gzip format. VGWAS summary statistics were reconstructed by saving only significant associations. Specifically, the effective number for each ROI was used to adjust for multiple hypothesis testing (Methods, Table S2). For the hippocampus (resp. WM tracts), the effective number was 10.1 (resp. 261.8), resulting in a Bonferroni *p*-value threshold of 5 × 10^*−*8^*/*10.1 = 4.94 ×10^*−*9^ (resp. 1.91 × 10^*−*10^).

We excluded SNPs associated with a small number of voxels by using the wild bootstrap approach [35] (Methods and Supplementary Note), since the loci derived from these associations are likely due to local perturbations in images. The remaining voxel-variant associations within each ROI were combined into loci using the Peaks algorithm [8]. All significant voxel-variant pairs within a locus were close in genetic distance, with the longest distance from the most significant association being less than 0.25 cM. Compared with combining all voxel-variant associations (815 loci for WM tracts and 134 loci for hippocampus), 37.0% of loci were excluded. For WM tracts, we ended up with 526 loci, with 251 (47.7%) overlapping with significant loci identified in previous studies using the same UKB data and IDP-based approaches [8, 10] (*<* 0.25 cM), and 49.7% of known loci in these two studies (251 out of 505) were replicated by our study (Fig. 2A, Table S3). For the hippocampus, we identified 72 loci, with 33 (45.8%) overlapping with associations in the NHGRI-EBI GWAS catalog and our previous study [12] (Fig. 2B, Table S4). In total, we identified 275 novel loci for WM tracts and 39 novel loci for the hippocampus through voxelwise inspection.

**Fig. 2.**
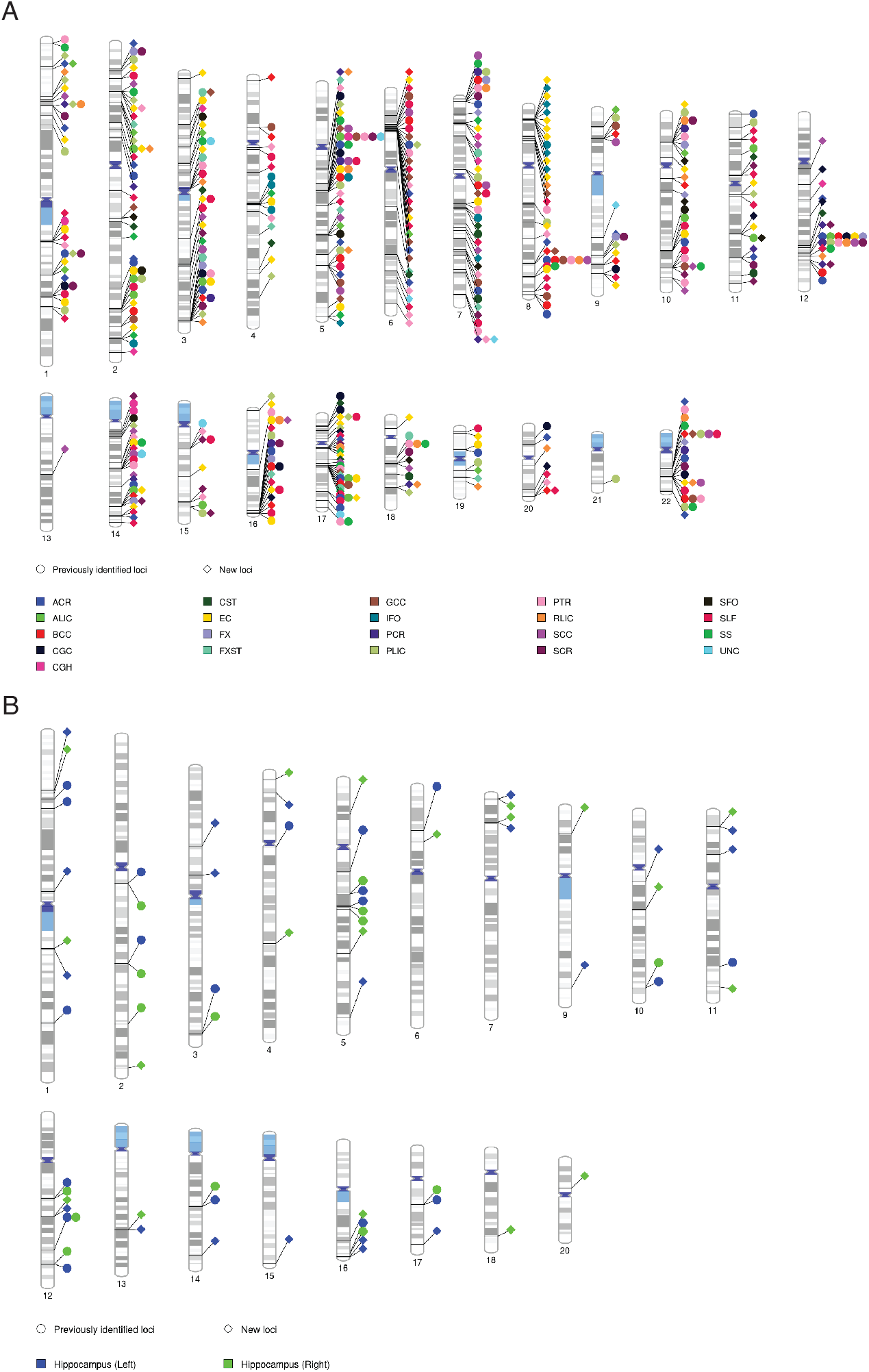
The identified genetic loci for WM microstructure and the hippocampus shape. Each point represents a locus by grouping significant voxel-variant pairs within an ROI such that the longest distance from any variant to the most significant variant was less than 0.25 cM. Each voxel-variant pair can be included in one and only one locus. A previous locus was replicated by our study if the most significant variant in that locus was within 0.25 cM from any of the most significant variants in our loci. We have harmonized the definition of loci in previous studies and the current study before comparison. **(A)** Ideogram of loci influencing WM tracts with the *p*-value threshold 1.91 *×* 10^*−*10^, including 275 novel loci and 251 previously identified loci [8, 10]. **(B)** Ideogram of loci influencing hippocampus with the *p*-value threshold 4.94 *×* 10^*−*9^, including 39 novel loci and 33 previously identified loci from our previous study [12] and all hippocampus-related studies on NHGRI-EBI GWAS catalog.

We discovered numerous colocalizations with other brain structural and functional measurements, as well as with brain-related phenotypes. At 3q24, the index SNP (i.e., the most significant SNP in a locus) rs2279829 was associated with external capsule (novel) and superior longitudinal fasciculus (previously known) [10] (Fig. 3A and B). This locus was also linked to insomnia [36]. A novel locus at 13q31.1 was associated with the left hippocampal tail (Fig. 3E and F) and showed colocalization with smoking [37–39], educational attainment [36, 40], and major depressive disorder [41]. Both these two loci can be stringently replicated using an independent dataset (see the next section). We observed that the index SNP z-score atlases from RVGA were nearly identical to those from VGWAS (Fig. 3C vs. D, G vs. H).

**Fig. 3.**
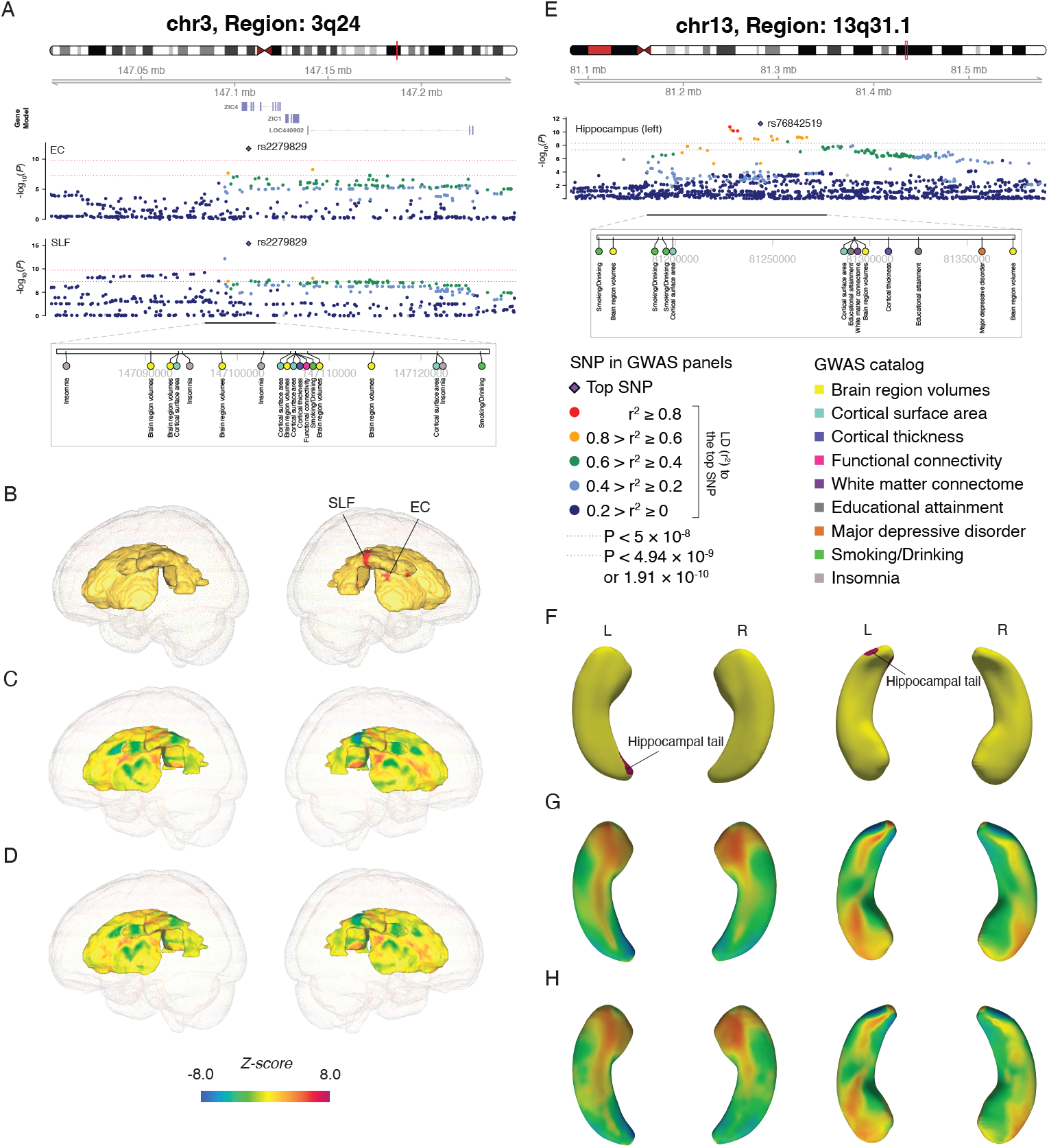
Selected genetic loci that were associated with subregions in WM tracts/hippocampus, brain structural and functional traits, and brain-related phenotypes. **(A)** At 3q24, we identified a locus with index variant rs2279829 that was simultaneously associated with external capsule (EC, novel) and superior longitudinal fasciculus (SLF, previously known). **(B)** Significant subregions associated with rs2279829 (*P <* 1.91 *×* 10^*−*10^) are highlighted in red. **(C-D)** Atlases of z-scores for rs2279829 from RVGA and VGWAS, respectively. **(E)** At 13q31.1, we discovered a locus linked to vertices in left hippocampal tail. This locus was novel to the hippocampus, while it displayed connections with brain structural and connectivity measurements as well as smoking initiation, educational attainment, and major depressive disorder. **(F)** Significant subregions in the left hippocampal tail associated with rs76842519 (*P <* 4.94 *×* 10^*−*9^) are highlighted in red. **(G-H)** Atlases of z-scores for rs76842519 (left hippocampus) and rs36188842 (right hippocampus, the most significant SNP in the locus) from RVGA and VGWAS, respectively.

### Validating RVGA GWAS results

We validated the RVGA results through several complementary analyses. First, we wanted to show that RVGA is aligned well with VGWAS for significant associations. We performed association analyses on 10 randomly selected variants that showed significant associations (*P <* 5 ×10^*−*8^) in RVGA, along with 90 additional randomly selected variants. We evaluated these variants across all vertices in the left hippocampus and all voxels in the superior fronto-occipital fasciculus using both RVGA and VGWAS (Extended Data Figs. 9 and 10). Across all 100 variants, the mean correlation coefficient of genetic effect was 0.90 (*sd* = 0.05) and the mean correlation coefficient of z-score was 0.84 (*sd* = 0.07) for the hippocampus, while the mean correlation coefficient of genetic effect was 0.84 (*sd* = 0.06) and the mean correlation coefficient of z-score was 0.84 (*sd* = 0.07) for the superior fronto-occipital fasciculus. If focusing on the first 10 variants, the counterparts were improved to 0.98 (*sd* = 0.01) and 0.95 (*sd* = 0.01) for the hippocampus, and improved to 0.95 (*sd* = 0.04) and 0.95 (*sd* = 0.02) for the superior fronto-occipital fasciculus. Moreover, more significant associations were identified by RVGA relative to VGWAS. Overall, RVGA aligned with and for most cases outperformed VGWAS in identifying significant associations, while the noise removal caused the relatively low alignment for insignificant associations (Discussion).

Additionally, we wanted to evaluate RVGA summary statistics across the whole genome. We performed GWAS for the 460,000 genotyped SNPs on 100 randomly selected points from the above two ROIs, comparing RVGA z-scores to those from VGWAS. Focusing on SNPs with *p*-values less than 0.05 in both RVGA and VGWAS, we observed a mean correlation coefficient of 0.99 (*sd* = 0.005) and a mean RMSE of 0.34 (*sd* = 0.09) for the left hippocampus. For the superior fronto-occipital fasciculus, the mean correlation and mean RMSE were 0.98 (*sd* = 0.002) and 0.44 (*sd* = 0.03), respectively. Considering all SNPs, we observed similar genomic inflation factors (*λ*_*GC*_) of 1.05 (*sd* = 0.01) for RVGA and 1.04 (*sd* = 0.02) for VGWAS for the left hippocampus, and 1.05 (*sd* = 0.01) for RVGA and 1.04 (*sd* = 0.01) for VGWAS for the superior fronto-occipital fasciculus.

We further replicated our findings using an independent dataset of 20,130 unrelated European subjects from UKB (Methods and Supplementary Note). We considered all SNPs in the significant loci from the discovery phase. Because of strong LD among SNPs in a locus, any SNPs being significant in the replication study indicated the locus was replicated. For WM microstructure (Table S5), we first evaluated alignment of genetic effect size by matching significant voxel-variant pairs in the discovery (*P <* 1.91 × 10^*−*10^) and in the replication (*P <* 0.05*/*526). The correlation of effect size estimates was 0.96, with 99.3% of effects showing a consistent direction. Then we evaluated loci-level replication. Out of the 526 loci, 513 (97.5%) were replicated 8 (Table S3), and 369 (70.2%) were replicated additionally considering the effective number (*P <* 0.05*/*526*/*261.8). For the hippocampus (Table S6), the correlation of effect size estimates was 0.98, with 99.8% of effects showing a consistent direction. All of the 72 loci were replicated (*P <* 0.05*/*72, Table S4), and 63 (87.5%) were replicated at the more stringent threshold (*P <* 0.05*/*72*/*10.1). Extensive sensitivity analyses, including comparing PCA and FPCA, varying numbers of LDRs, and adjusting for additional covariates, are detailed in the Supplementary Note.

### Heritability and genetic correlation in brain images

We estimated voxel heritability and genetic correlations for each ROI using the shared triplets and the LD matrix of 1,160,000 HapMap3 SNPs with regularization of {98%, 95%} (Methods). All ROIs exhibited moderate heritability (Table S7). The mean heritability for the hippocampus was 18.0% (*se* = 1.9%). WM tracts showed a mean heritability of 23.2% (*se* = 1.9%), ranging from 12.7% for the corticospinal tract to 30.2% for the inferior fronto-occipital fasciculus.

Voxelwise heritability estimates revealed significant variations across subregions. Heritability was high (23%) in the CA3 and presubiculum of the hippocampus, while the hippocampal tail, CA1, and subiculum exhibited lower heritability (10%) (Fig. 4A). In WM tracts, regions of the retrolenticular part of the internal capsule and the superior corona radiata showed high heritability (37%), which decreased to 20% in nearby regions (Fig. 4A).

**Fig. 4.**
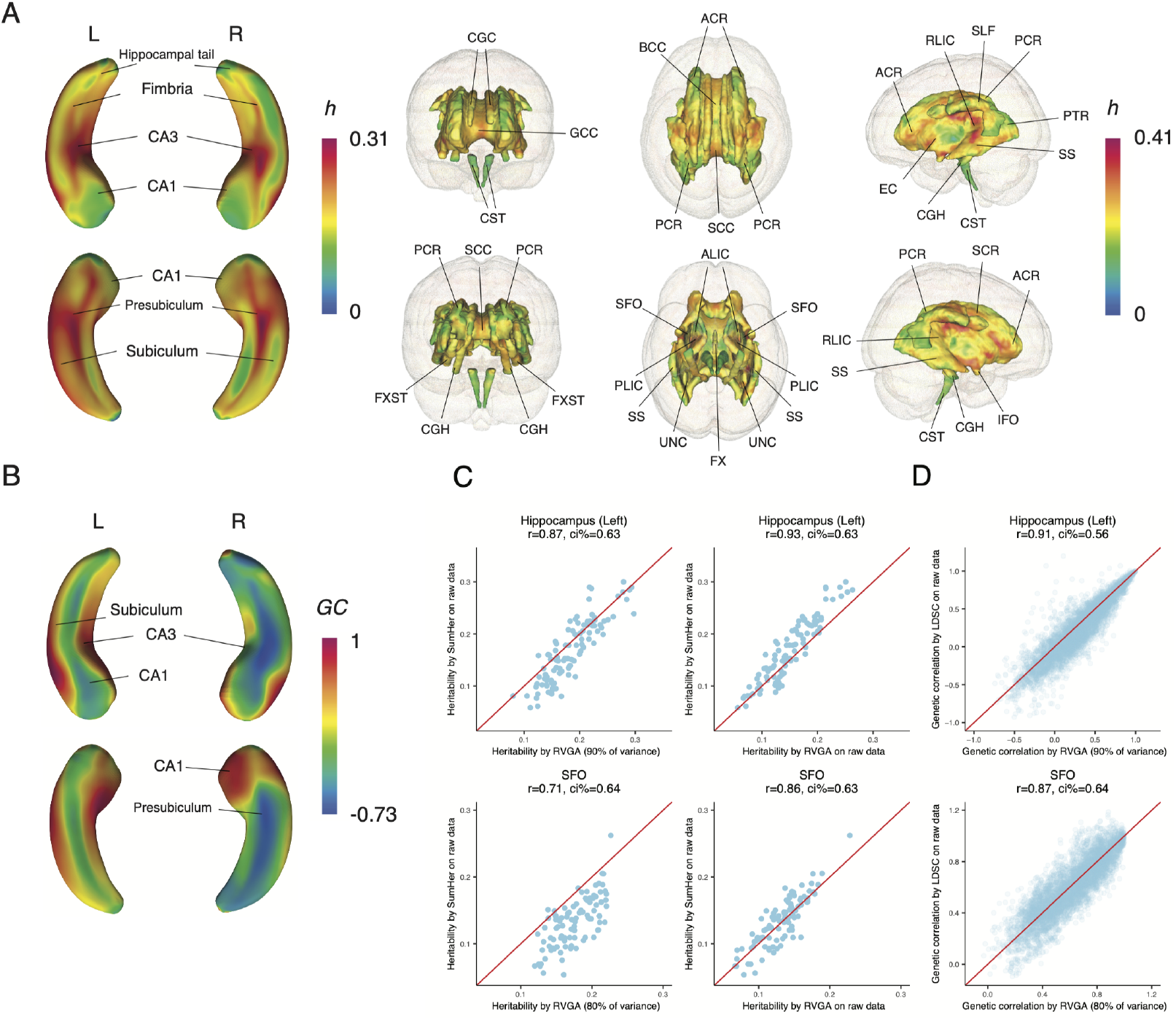
Heritability and genetic correlation estimates in brain images. **(A)** The voxelwise heritability of the hippocampus shape and WM microstructure. Refer to Table S1 for the full name of WM tracts. **(B)** The genetic correlations between the most heritable vertices and other vertices within the left and right hippocampus, respectively. The most heritable vertex is at the subiculum for both the left and right hippocampus. **(C)** A comparison of RVGA estimates to SumHer estimates on 100 randomly selected vertices (voxels) on the left hippocampus (the superior fronto-occipital fasciculus). Left column: RVGA implemented FPCA to construct LDRs for a certain proportion of variance, then estimated vertex (voxel) heritability by using LDR summary statistics. Right column: RVGA directly estimated heritability by using summary statistics of each vertex (voxel). **(D)** A comparison of genetic correlation estimates between RVGA and LDSC. In each plot, “r” refers to Pearson correlation coefficient, and “ci %” refers to relative confidence interval width, defined as the mean standard error of RVGA estimates to that of SumHer (LDSC) estimates. SumHer (LDSC) was conducted using the summary statistics estimated from VGWAS. We used the “BLD-LDAK” model in SumHer and LD scores estimated by using 1000 Genome data in LDSC.

Genetic correlation estimates indicate shared genetic architecture between subregions (Table S8). Nearby voxels had genetic correlations close to one, but this was not always true for distant voxels. For example, the subiculum of the left hippocampus had a highly positive genetic correlation with CA3 (90%) but a negative correlation with CA1 (− 50%). In the right hippocampus, the genetic correlation between the subiculum and CA1 ranged from − 45% to 80% (Fig. 4B).

Comparing RVGA heritability estimates with those from SumHer [42] for 100 randomly selected points, we found a correlation coefficient of 0.87 for the left hippocampus and 0.71 for the superior fronto-occipital fasciculus (Fig. 4C, left column). The relative confidence interval (CI) width between RVGA and SumHer was 0.63 and 0.64, respectively. The two methods were highly consistent when estimating heritability using RVGA directly on each point (Fig. 4C, right column). Therefore, RVGA did not overestimate heritability but rather enhanced genetic influence by extracting underlying imaging signals through FPCA (Supplementary Note).

The RVGA genetic correlation estimates closely matched those from LDSC, with correlation coefficients of 0.91 and 0.87, and relative CI widths of 0.56 and 0.64, respectively. RVGA achieved statistically efficient estimates by reducing noise through FPCA and utilizing complete-sample-overlap information. Extensive sensitivity analyses are detailed in the Supplementary Note.

### Cross-trait genetic correlation between brain images and brain-related phenotypes

We gathered summary statistics for 11 complex brain disorders and three brain-related phenotypes (Table S9), generating atlases of genetic correlations between ROIs and phenotypes (Methods). We applied regularization of {90%, 85%} on the LD matrix in the analysis.

To summarize the overall significance of the genetic correlations between ROIs and phenotypes, we used the Cauchy combination strategy [43] to meta-analyze the *p*-values of all points in an ROI. We controlled the false discovery rate separately at 0.05 for the hippocampus (28 ROI-phenotype pairs, Table S10) and WM tracts (294 ROI-phenotype pairs, Table S11), yielding no significant results for the hippocampus and 51 significant results for WM tracts (Fig. 5A). Of these, 19 (37.3%) corroborated findings from a previous study [10]. Specifically, we replicated widespread genetic correlations for FA of WM tracts with educational attainment, cognitive performance, and intelligence. Additionally, we replicated significant pairs involving the anterior limb of the internal capsule and fornix/stria terminalis with depression, and the superior longitudinal fasciculus with schizophrenia.

**Fig. 5.**
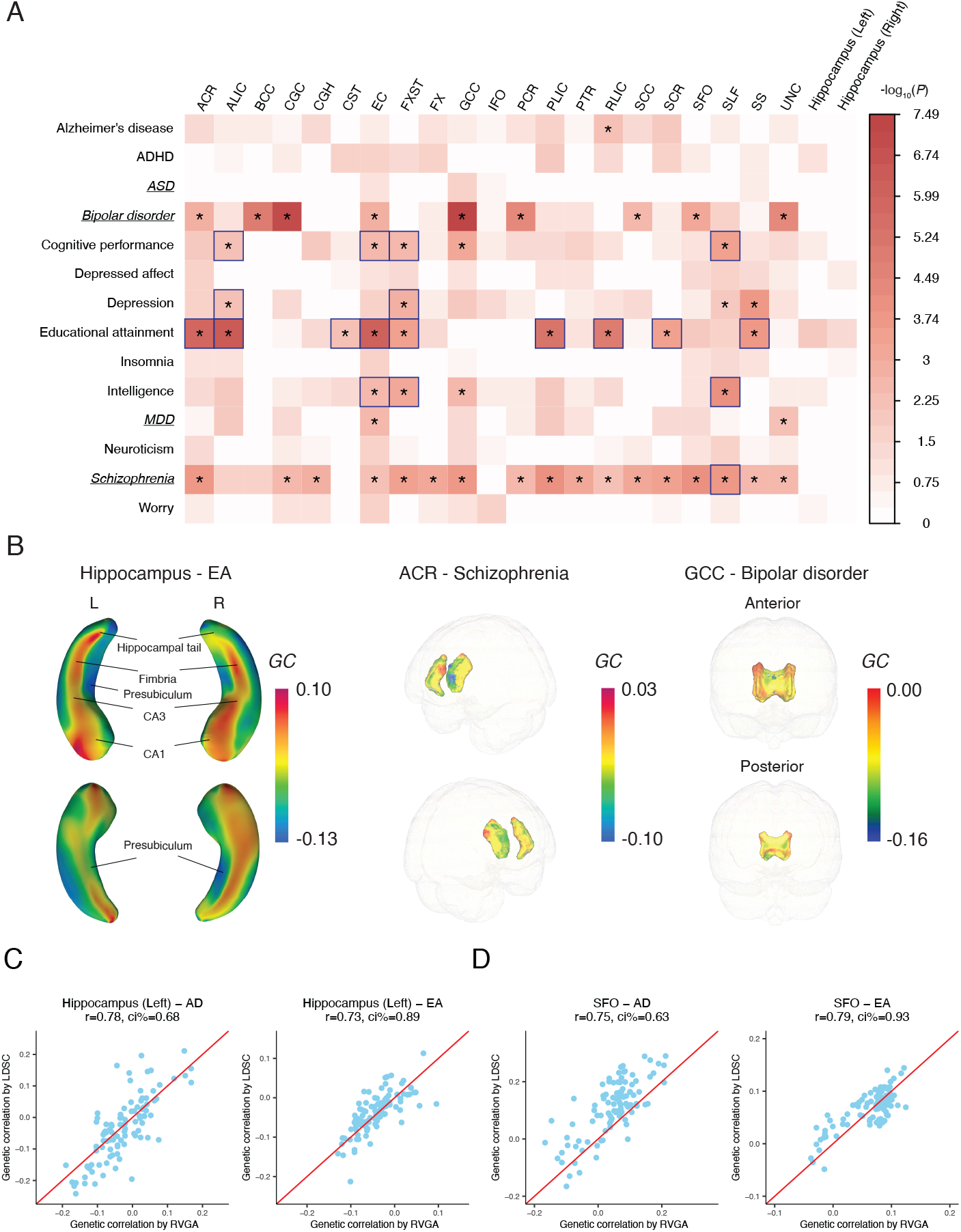
Highlighted discoveries from cross-trait genetic correlation analysis between brain images and brain-related phenotypes. **(A)** The overall strength of genetic correlation between ROIs and brain-related phenotypes, which is measured through a *p*-value generated by meta-analyzing voxelwise genetic correlation *p*-values using the Cauchy combination strategy [43]. The asterisks denote significant results by controlling the false discovery rate at 0.05 considering all ROI-phenotype pairs. We conducted multiple hypothesis correction separately for the hippocampus and WM tracts. The boxes denote replicates for the previous study [10]. Italic and underscored phenotypes indicate no evidence of sample overlap with the UKB cohort, while other phenotypes suggest potential sample overlap. Refer to Table S1 for the full name of WM tracts. **(B)** Selected cross-trait genetic correlations between the ROIs and brain-related phenotypes. EA: educational attainment. **(C)** and **(D)** A comparison of RVGA cross-trait genetic correlation estimates and LDSC counterparts on 100 randomly selected vertices (voxels) on the left hippocampus (the superior fronto-occipital fasciculus). LDSC was conducted using the summary statistics estimated from VGWAS and LD scores estimated by using 1000 Genome data. In each plot, “r” refers to Pearson correlation coefficient, and “ci %” refers to relative confidence interval width, defined as the mean standard error of RVGA estimates to that of LDSC estimates.

Inspecting genetic correlations between subregions and phenotypes (Fig. 5B), we found, for example, that the hippocampal tail, fimbria, CA3, and CA1 had positive correlations (6% ~ 10%, *se* = 3.8%) with educational attainment, while the presubiculum exhibited a negative correlation (− 13%, two-sided *z* test, *P* = 0.001 *<* 0.05*/*10.1). Inconsistent patterns were observed between hemispheres, such as the right anterior corona radiata showing a negative correlation (− 9%, *P* = 4.3 × 10^*−*5^ *<* 0.05*/*261.8) with schizophrenia. Moreover, the genu of the corpus callosum was negatively correlated with bipolar disorder, with varying levels across subregions (− 16% ~ 0, *se* = 2.6%).

We validated our findings by comparing RVGA genetic correlation estimates with those from LDSC [44] for 100 randomly selected points. For the left hippocampus and Alzheimer’s disease/educational attainment, the correlation coefficients were 0.78 and 0.73, with relative CI widths of 0.68 and 0.89, respectively (Fig. 5C). For the superior fronto-occipital fasciculus and the same traits, the coefficients were 0.75 and 0.79, and relative CI widths were 0.63 and 0.93, respectively (Fig. 5D). Extensive sensitivity analyses are detailed in the Supplementary Note.

### Computational cost

To assess computational efficiency, we benchmarked RVGA using data from the left hippocampus (15,000 vertices and 32,021 subjects) and cortical surface curvature (59,412 vertices and 15,752 subjects), along with 7.8 million variants, on computational clusters equipped with four 2.30 GHz CPUs running in parallel (Table 1). For the hippocampus, most steps took less than 30 minutes and used under 25 GB of memory. The most time consuming steps were LDR GWAS and wild bootstrap, taking 4.7 and hours, respectively. RVGA demonstrated high efficiency, even for high-resolution images of the whole cerebral cortex. For example, reconstructing whole-genome summary statistics for 59,412 vertices took 32.3 hours, while estimating vertex heritability and genetic correlations between all vertex pairs required only 25 minutes.

**Table 1.** Benchmarking the computational efficiency for RVGA. The first dataset includes the left hippocampus shape data with 15, 000 vertices and 32, 021 subjects. The second dataset includes the cortical surface curvature data with 59.412 vertices and 15, 752 subjects. A total of 7.8 million common SNPs and 4 CPUs with 2.30 GHz were used in the experiment.

|  | Time (hours) | Memory (GB) | Output (GB) |
| --- | --- | --- | --- |
| FPCA<sup>1</sup> | 1.7 | 9.7 | 0.2 |
| Constructing 25 LDRs | 0.02 | 12.0 | 0.04 |
| LDR GWAS | 4.7 | 20.0 | 17.0 |
| Processing LDR summary statistics<sup>2</sup> | 0.04 | 9.5 | 2.2 |
| Voxel-level summary statistics recovery<sup>3</sup> | 1.2 | 22.1 | 0.05 |
| Heritability and genetic correlation | 0.03 | 5.5 | 1.7 |
| Cross-trait genetic correlation | 0.02 | 3.4 | 0.002 |
| Wild bootstrap<sup>7</sup> | 5.5 | 30.0 | 0.01 |
| FPCA<sup>4</sup> | 2.5 | 25.5 | 3.5 |
| Constructing 1750 LDRs | 0.3 | 20.0 | 0.3 |
| LDR GWAS<sup>5</sup> | 40.8 | 20.0 | 411.2 |
| Processing LDR summary statistics<sup>2</sup> | 0.6 | 20.0 | 87.2 |
| Voxel-level summary statistics recovery<sup>6</sup> | 32.3 | 50.0 | 0.06 |
| Heritability and genetic correlation | 0.4 | 20.0 | 27.0 |
| Cross-trait genetic correlation | 0.3 | 20.0 | 0.005 |
| Wild bootstrap<sup>7</sup> | 25.2 | 60.0 | 0.01 |
| LD matrix construction<sup>8</sup> | 6.5 | 4.2 | 2.8 |

<sup>1</sup>Estimating top 3,000 components.
<sup>2</sup>Removing duplicate SNPs, strand-ambiguous SNPs, SNPs with small sample size, missing or infinite values, and indels, resulting in 6.6 million SNPs. Saving the data in HDF5 format of data type float32.
<sup>3</sup>Saving significant associations ( $P < 4.94 \times 10^{-9}$ ).
<sup>4</sup>Estimating all 15,752 components.
<sup>5</sup>Conducting GWAS for 200 LDRs in a batch.
<sup>6</sup>Saving significant associations ( $P < 3.15 \times 10^{-11}$ ).
<sup>7</sup>Using 150,000 independent SNPs and computing 50 bootstrap samples.
<sup>8</sup>Using 42,000 subjects with 1,160,000 HapMap3 SNPs from UKB imputed genotype data.

## Discussion

We introduced the novel RVGA framework. RVGA’s major contributions include the ability to demonstrate genetic effects on the human brain at a substantially higher resolution than IDP-based methods, with straightforward potential for extension to other organs using biomedical images. This enables users to precisely identify regions significant associated with genetic variants, inspect heritability and genetic correlation variations, and evaluate genetic connections between organs and complex disorders.

RVGA utilizes FPCA to construct LDRs for images, conducts GWAS on LDRs, and employs functional bases to reconstruct voxel-level summary statistics. This approach significantly reduces computational time, cost, and storage space by orders of magnitude compared to performing GWAS on each individual voxel. Additionally, RVGA optimizes data storage by saving only the summary statistics of LDRs, the functional bases, and the variance-covariance matrix of LDRs. This standard enables the sharing of highly informative voxel-level summary statistics with the community for secondary analyses.

In real data analysis, RVGA identified 598 genetic loci, 585 (97.8%) of which can be replicated and 432 (72.2%) can be replicated at more stringent thresholds. Notably, we discovered novel local genetic connections, such as CA1 and presubiculum with educational attainment, subregions in the anterior corona radiata with schizophrenia, and the genu of the corpus callosum with bipolar disorder. More importantly, we reduced data size for voxel-level secondary analyses by 229 times.

We demonstrated RVGA’s robustness and effectiveness through extensive simulations, real data, and sensitivity analyses. When comparing association z-scores between RVGA and VGWAS, we observed that RVGA showed strong alignment with VGWAS for significant associations, although the alignment was weaker for insignificant ones. That is because RVGA effectively smooths images while slightly shrinking each voxel’s value toward zero. In general, this shrinkage leads to a significant reduction in standard error estimates and a small decrease in the absolute effect sizes. For associations with large effect sizes, this reduction can yield smaller *p*-values. For insignificant associations, standard error estimates are still reduced, but changes in absolute effect sizes are random, as these values are near zero, resulting in weaker alignment. It is the reason why RVGA can boost statistical power while protecting type I error rate.

Although top LDRs can capture majority of imaging signals, it is not necessary that no genetic variability is lost. Nevertheless, we have shown that RVGA is more powerful than VGWAS to detect true associations, indicating more noise is removed relative to genetic signals. Additionally, the voxel heritability estimator based on the top LDRs is unbiased under strong noise. Overall, selecting an appropriate number of LDRs can enhance statistical power and computational efficiency. We recommend preserving 80%-90% of imaging signals and ensuring the correlation between the raw and reconstructed images is 0.85-0.95 to balance bias, variance, and computational cost.

There are several limitations and potential future directions for RVGA. First, the primary source of bias in RVGA is model mis-specification - it is not able to correctly specify and capture all covariate/genetic effects and imaging signals. Second, RVGA currently restricts GWAS to unrelated subjects and common variants. Future work should address relatedness among subjects [45] and incorporate approaches for whole-genome/exome sequencing data [46]. Third, RVGA is computationally intractable for images with millions of voxels due to the increasing memory demand for FPCA, necessitating more efficient representation learning approaches.

Nevertheless, RVGA advances imaging genetic studies to the next level. Beyond traditional biomedical images, RVGA is well-suited to assess other data types, such as microscopy of 3D brains [47], and transcriptomic and spatial cell-type atlases [48].

## Data Availability

The raw data is from UK Biobank at https://www.ukbiobank.ac.uk;
The shared data generated by using HEIG software is at https://zenodo.org/records/13787684.

https://github.com/Zhiwen-Owen-Jiang/heig

## Methods

### Math notations

Table 2 demonstrates key math notations used in the article.

**Table 2.** Math notations used in the article.

| Notation | Meaning | Notation | Meaning |
| --- | --- | --- | --- |
| $Y(\cdot)$ | Raw image | $N$ | Number of voxels/vertices |
| $X(\cdot)$ | Underlying image | $d$ | Number of variants |
| $\alpha(\cdot)$ | Fixed covariate effect | $n$ | Sample size |
| $\beta(\cdot)$ | Fixed genetic effect | $r$ | Number of LDRs |
| $\eta(\cdot)$ | Image without covariate and genetic effects | $\phi, \Phi$ | Functional base |
| $\epsilon(\cdot)$ | White noise | $\lambda$ | Image eigenvalue |
| $W$ | Covariate matrix | $u, v$ | Voxel/vertex index |
| $Z$ | Genotype matrix | $\xi, \Xi$ | Low-dimensional representation |
| $R$ | LD matrix | $H$ | kernel bandwidth |
| $\Omega$ | LD matrix inverse | $Q(\cdot, \cdot)$ | Genetic variance/covariance |
| $h(\cdot)$ | Heritability | $GC(\cdot, \cdot)$ | Genetic correlation |
| $b_{kj}$ | Projection of $\beta_k(\cdot)$ on $\phi_j(\cdot)$ ($j$-th base) | $\sigma^2$ | Variance of white noise |
| $\theta_{ij}$ | Projection of $\eta_i(\cdot)$ on $\phi_j(\cdot)$ ($j$-th base) | $a$ | Polynomial decay rate of image eigenvalues |
| $\gamma$ | Strength of genetic effect size affected by LD | $\alpha$ | Strength of genetic effect size affected by MAF |

### A varying coefficient model for imaging genetics

In the RVGA framework, the imaging data is assumed to be indexed on a common compact set *ℐ* ⊂ ℝ ^3^ for all subjects, capable of capturing curves (1D), surfaces (2D) and volumes (3D) in ℝ^3^. All subjects have *N* common voxels in the image after appropriate preprocessing. A varying coefficient model for subject *i* ∈ {1, …, *n*} at voxel *v* is expressed as:

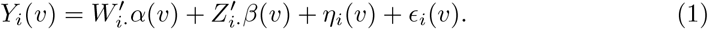

Here, *Y*_*i*_(·) represents an *N* × 1 vector of the image, *W*_*i·*_ is a *p* × 1 vector of covariates, including the intercept, and *α*(·) is a *p* × *N* matrix of fixed coefficients. *Z*_*i·*_ is a *d* × 1 vector of the genetic profile, and *β*(·) is a *d* × *N* matrix of fixed genetic effects. The unexplained image signals are captured by *η*_*i*_(·) and *ϵ*_*i*_(·). The term *η*_*i*_(·) is an *N* × 1 vector capturing relatively smooth noise component while the term *ϵ*_*i*_(·), also an *N*× 1 vector, captures purely random white noise.

The genotype matrix *Z* across all subjects with dimension *n* × *d* is assumed to be normalized using the formula 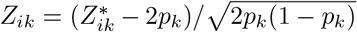. Here, 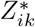 is the number of copies of the reference allele for the *i*-th subject and *k*-th SNP, and *p*_*k*_ is the frequency of the reference allele in the population. The genetic profile *Z*_*i·*_ follows a multivariate normal distribution *𝒩* (0, *R*), where 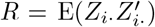 is a positive-definite LD matrix of *d* SNPs. Both *α*_*l*_(·), *l* ∈ {1, …, *p*}, and *β*_*k*_(·),*k* ∈ {1, …, *d*}, are fixed smooth functions. The residual term *η*_*i*_(·) is modeled as a stochastic process, represented by *η*_*i*_(·) ∼ SP(0, Σ_*η*_(·, ·)). The most common case is Gaussian process; however, the methodology of FPCA does not rely on the Gaussian assumption. For any *u, v* ∈ *ℐ*, Cov(*η*_*i*_(*u*), *η*_*i*_(*v*)) = Σ_*η*_(*u, v*) capturing spatial relationship. The white noise *ϵ*_*i*_(*v*) is assumed to be independent and identically distributed (i.i.d.) across all voxels and subjects. The distribution of *ϵ*_*i*_(*v*) can be normal, or other skewed distributions. The covariance of raw image is 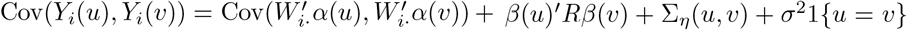, where 1{·} is the indicator function. Finally, it is assumed that 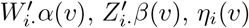, and *ϵ*_*i*_(*v*) are independent of each other.

Our varying coeffcient model can be regarded as a combination of standard voxel-wise linear regression models and random field theory. At each voxel *v*, the model decomposes the residual term into individual curve variations (*η*_*i*_(*v*)) and white noise (*ϵ*_*i*_(*v*)). The random field theory is applicable after smoothing *Y*_*i*_(*v*) or reducing *ϵ*_*i*_(*v*) to zero. Compared to voxel-wise linear models, varying coefficient models provide a unified framework for modeling different voxels. We have extensively discussed several key features (e.g., spatial smoothness) of varying coefficient models in Huang et al [24]. We have used the model to fit almost all MRI imaging phenotypes (e.g., cortical and subcortical structures) used in imaging genetics [49–55], some of which are presented in this paper.

### Representation learning with functional principal component analysis

We apply FPCA to decompose the image *Y* into smooth imaging signals and white noise. We first smooth the images and then compute a set of smooth functional bases to construct LDRs. In contrast, traditional PCA is less effective at removing noise, as it directly computes bases from raw images. We discussed the differences between FPCA and PCA in dimension reduction and noise removal, and compared RVGA to two related representation learning approaches in the Supplementary Note.

A local linear estimator [33] with a Gaussian kernel is used to smooth raw images by approximating the smooth functions underlying the raw images *Y*. Essentially, each voxel is represented by a weighted average of itself and neighboring voxels. The Gaussian kernel with a specified bandwidth (a hyperparameter) determines the weights and the number of voxels included. We initialize the bandwidth as *H* = *N*^*−*1*/*(*dim*+4)^, where *dim* is the dimension of the image (i.e., 2D or 3D), and use generalized cross-validation (GCV) [32] to select the optimal one from 0.5*H, H*, 2*H*, 3*H*, 5*H*, and 10*H*. Given the weights, the estimation is equivalent to solving a weighted least squares problem and, therefore, has a closed-form solution. The alternatives include local constant and local quadratic estimators. However, the former suffers from bias and poor boundary behavior, while the latter increases computational burden and estimate variance.

Let 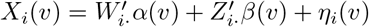 denote the real image without white noise, where we assume ∫_*ℐ*_ E(*X* ^2^) *<* ∞ and let the covariance function be *C*(*u, v*) = Cov(*X*_*i*_(*u*), *X*_*i*_(*v*)), which is positive semidefinite. The covariance function admits a spectral decomposition in terms of non-negative eigenvalues *λ*_*j*_ that

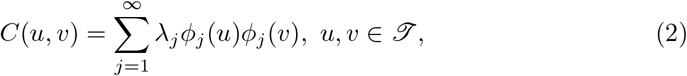

where *λ*_1_ ⩾ *λ*_2_ ⩾ · · · ⩾ 0, and the functions *ϕ*_1_, *ϕ*_2_, … form an orthonormal basis for the space of all square-integrable functions on *ℐ*.

Denote the sample covariance of the smoothed images as 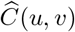. The spectral decomposition is applied to estimate eigenvalues 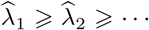, and functional basis 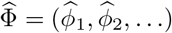 such that 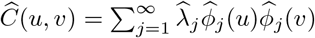 (we equivalently use singular value decomposition (SVD) in practice). Each base is an *N* × 1 vector. We project *Y*_*i*_ onto the subspace spanned by the first *r* bases and generate LDRs 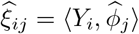, where *j*∈ {1, …, *r*}, 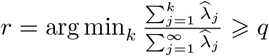 and *q* is a proportion of variance (⩾ 80%). The underlying imaging data can be approximately recovered by 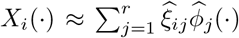.

As a guardrail, we suggest the correlation between the raw and reconstructed images greater than 0.85. A higher correlation, ideally 0.90-0.95, is preferable if computationally feasible. For large imaging datasets of high resolution, it may be computationally intractable to compute all *λ*’s (PCs) before selecting LDRs. We propose to compute only the top PCs and dynamically select LDRs in such cases (refer to “Subtlety of FPCA” section in Supplementary Note).

### Voxel-level GWAS using low-dimensional representations

In this section, we first present the association analysis for each voxel-variant pair, then move on to efficient reconstruction using LDR summary statistics. The covariate effects are first removed from the imaging and genetic data using a projection matrix *I*− *M*_*W*_ = *I* − *W* (*W*^*′*^*W*)^*−*1^*W*^*′*^, where *I* is the identity matrix, and *W* is an *n* × *p* matrix including the intercept and is assumed to be full rank. Denoting the resulting imaging data as 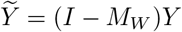, the genetic matrix as 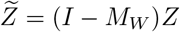. We normalize 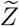 to have a variance of one. A univariate varying coefficient model for VGWAS is

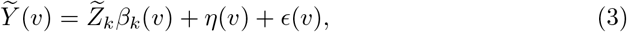

where 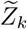 is an *n* × 1 genotype vector for the *k*-th SNP, and *β*_*k*_(*v*) is its fixed effect for voxel *v*. The terms *η*(*v*) and *ϵ*(*v*) are *n* × 1 vectors. Note that 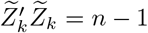 after normalization. For each fixed *v*, model (3) is essentially a linear model. Therefore, the voxelwise summary statistics can be estimated using the ordinary least square method that

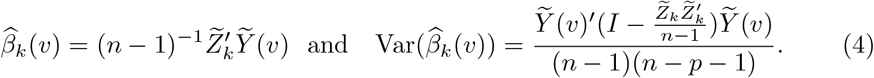

We use the Wald test to evaluate associations,

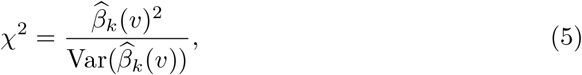

which follows a 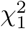 distribution under the null hypothesis that the true marginal effect *β*_*k*_(*v*) = 0, assuming *η*(·) and *ϵ*(·) are Gaussian. When the normal assumption is violated, the test statistic asymptotically follows a *χ*^2^ distribution as long as the sample size is large and the variant is common.

We use LDR summary statistics to reconstruct 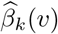 and 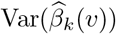. Denote 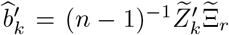 as the genetic effects of SNP *k* for all *r* LDRs, and 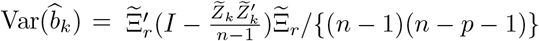 as the estimated variance-covariance matrix of LDR genetic effects, where 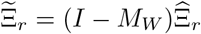. Recall that the real imaging data can be approximated by 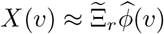, where 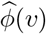 is the *v*-th column of 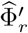 We have

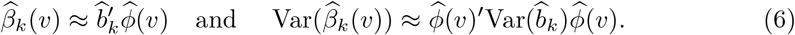

In practice, the genotype matrix is not normalized prior to GWAS, and the actual sample size *n*_*k*_ may differ across SNPs due to different missing patterns in genetic profiles. Consequently, the summary statistics and the variance have a more general form that

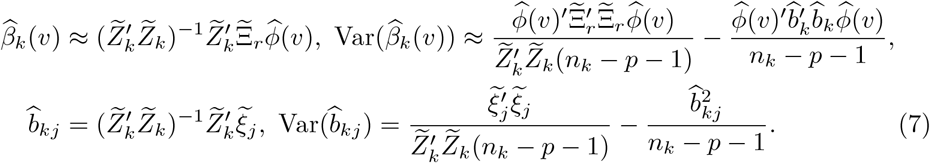

An additional step is to estimate 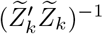 from the summary statistics of LDRs that

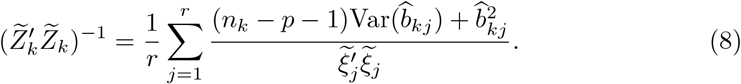

In summary, we only need to compute and store the summary statistics of LDRs, the variance-covariance matrix of the adjusted LDRs 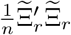, and the corresponding bases 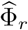 to recover the VGWAS results. This reduces the storage burden from *O*(*dN*) to *O*(*dr* + *Nr* + *r*^2^), and the computational burden from *O*(*dNn*) to *O*(*dNr* + *dnr*), achieving an average computation reduction by *N/r* times.

### Post-GWAS screening based on cluster size

The post-GWAS screening is based on evaluating cluster size, defined as the number of associated voxels for a SNP. The key idea is that reliable associations for a SNP should not be restricted to a small number of voxels. We compute the null distribution of cluster size using the wild bootstrap approach [35]. SNPs with a cluster size less than the quantile at 1 − 0.05/(number of loci * effective number) are excluded. The remaining voxel-variant associations are aggregated into loci and reported. Refer to the Supplementary Note for the detailed algorithm of wild bootstrap.

### Heritability and genetic correlation analysis in images

In heritability and (cross-trait) genetic correlation sections, we ignore the covariate term in model (1) for the ease of illustration, assuming all covarite effects have been removed. Let the real imaging data without white noise be 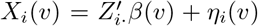, where *Z*_*i·*_ and *η*_*i*_(*v*) are random. The genetic covariance between *u, v* ∈ *ℐ*^*·*^ is

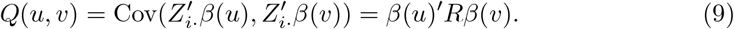

The heritability for voxel *v* is defined as

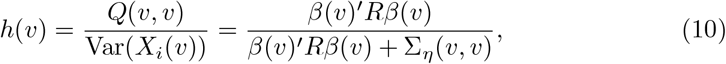

and the genetic correlation between a pair of voxels *u, v* is defined as

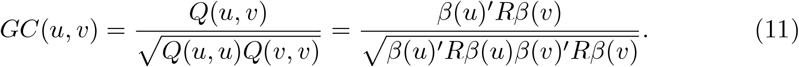

Note the variance of white noise *Σ* is not incorporated in the definition of voxel heritability in (10) because it is introduced by scanner instability rather than by environmental factors. If including *Σ*^2^, the same group of subjects scanned by two image scanners with different levels of white noise would produce inconsistent heritability. Although we cannot observe the real imaging data, the FPCA procedure can reduce the variance of white noise to *O*(*r/N*) ≈ *O*(1*/N*), which is negligible compared with *β*(*v*)^*′*^*Rβ* and Σ_*η*_(*v, v*) (assuming they are *O*(1)). If we were to directly perform analysis on each voxel in the raw imaging data, the heritability estimate would always be downward biased (Fig. 4C, Extended Data Fig. 4, Fig. S11, and refer to the Supplementary Note for details).

Following the work by Wang et al. [56], the genetic covariance for *u, v*∈ *ℐ* is estimated using the method of moments that

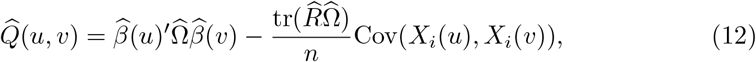

where Ω = *R*^*−*1^, and tr(·) is the trace of matrix. Check the Supplementary Note for derivation. Both 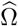 and 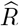 can be estimated from reference panels with matched population ancestry. It is recommended to use two different panels without sample overlap to estimate *R* and Ω for accuracy of tr(*R*Ω). Cov(*X*_*i*_(*u*), *X*_*i*_(*v*)) can be estimated using the variance-covariance matrix of LDRs and the functional bases. A plug-in estimator for heritability is then constructed by

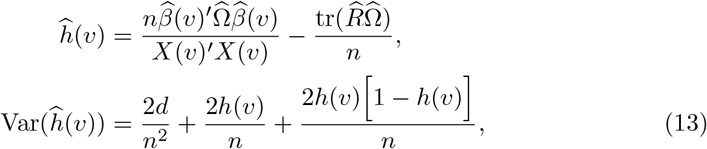

and the plug-in estimator for genetic correlation is

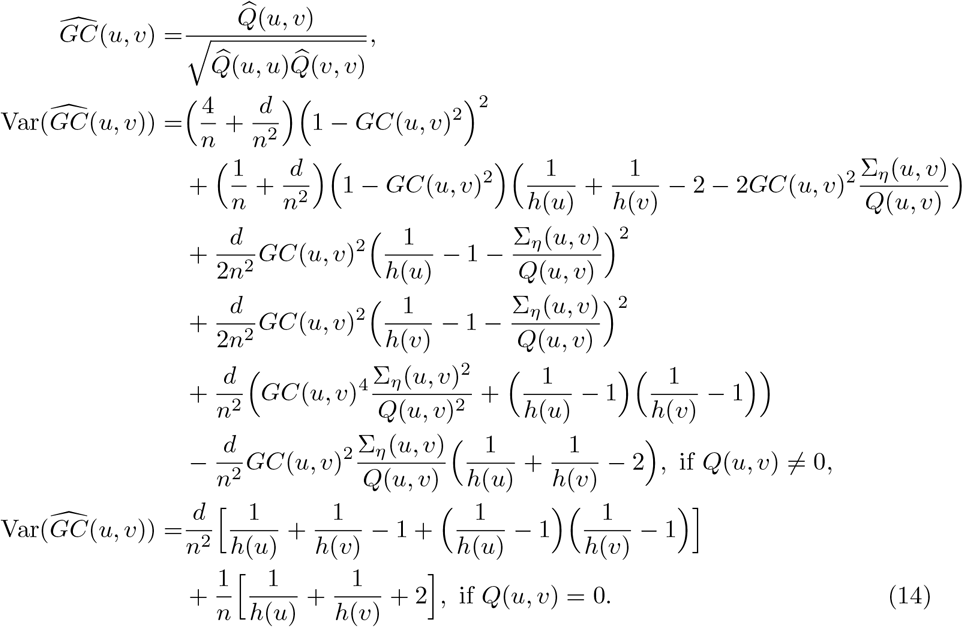

The derivation of the variance is in the Supplementary Note. With the estimate and standard error, we have a *χ*^2^ statistic that asymptotically follows *χ*^2^ under the null hypothesis that the true value is 0 [56, 57].

These estimators perfectly adapt to our framework as both 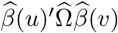 and *X*(*u*)^*′*^*X*(*v*) can be first estimated at LDR level and then use the bases to project to voxel level, thus the high efficiency. The estimators do not depend on the normal distribution assumption nor an infinitesimal model or specific structure on genetic effects [58]. Therefore, the estimators are expected to be robust to model mis-specification [56, 59]. Our heritability estimator is related to generalized random effects (GRE) model [59] and we compared them in the Supplementary Note.

### Cross-trait genetic correlation analysis between images and non-imaging phenotypes

Suppose that we have summary statistics of a non-imaging phenotype *U* from the identical population, which can be continuous or binary. We aim to evaluate the genetic correlation between *U* and *X*(*v*), *v* ∈ *ℐ*, i.e. the genetic correlation between *U* and each voxel in *X*. Let 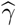 denote the summary statistics of *U* measured on a common set of SNPs as *X*(*v*). Suppose that there are *n*_2_ unrelated subjects for *U*, and *n*_0_ among them are overlapped with the subjects in *X*(*v*). We estimate the genetic covariance and genetic correlation by

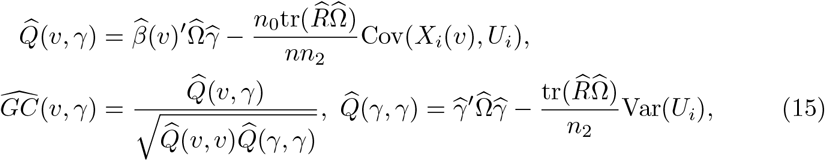

where the subject *i* was recruited in both studies. Note that in practice, it is unlikely to know the overlapping sample size *n*_0_ and Cov(*X*_*i*_(*v*), *U*_*i*_) from summary statistics. However, if there is no evidence of subject overlap in two studies, then *n*_0_ = 0, and we have the variance of 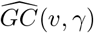 in an analytical form [56] that

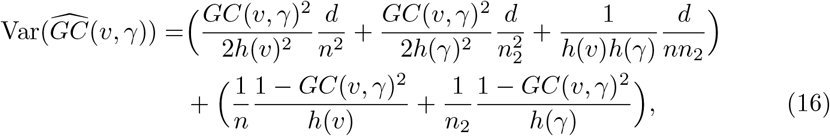

where 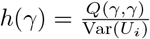 is the heritability of *U*. If sample overlap exists between two studies, we implement cross-trait LDSC [44] within RVGA and use the intercept to estimate *n*_0_Cov(*X*_*i*_(*v*), *U*_*i*_), as LDSC is exactly a special case of our estimator (Supplementary Note). Specifically, we implement cross-trait LDSC for each pair of LDR 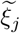 and *U*, obtaining an estimate for 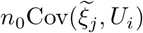. We then use the bases to project 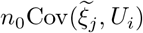 onto the voxel level. LD scores are automatically estimated when constructing an LD matrix in RVGA (see the section below).

The analytical variance of 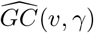 is too complicated to derive in this case, and the unknown *n*_0_ and Cov(*X*_*i*_(*v*), *U*_*i*_) make it impossible to evaluate. We resort to LD block-wise jackknife with one LD block left out each time to empirically estimate the variance, for which we evenly split the whole genome into approximately 200 blocks of adjacent SNPs.

### Estimation of LD matrix and LD scores from external datasets

The LD matrix *R* is a *d* × *d* positive-semidefinite matrix that measures the correlation among SNPs. The whole genome, excluding the sex chromosomes, can be divided into 1, 703 approximately independent LD blocks (based on the genome of white subjects, GRCh37) [60]. Consequently, we only take into account the correlation among SNPs within each block, assuming that there is no inter-block correlation. In the current version of RVGA, we used only common variants with an MAF greater than 0.01 from the genotype array or HapMap3 SNPs, mainly because it is adequate to capture the majority of heritability.

We noticed that regularization on the LD matrix is crucial for accurate and robust estimation. We employ eigen-decomposition and only preserve top eigenvalues. The specific proportion of variance to preserve depends on the type of SNPs included in the LD matrix. More details about LD matrix and LD score estimation are included n the Supplementary Note.

### *p*-Value threshold of multiple hypothesis testing across voxels

To determine the *p*-value threshold for multiple hypothesis testing across all voxels in GWAS, heritability and genetic correlation analysis, we resort to the effective number of independent voxels [61], defined as

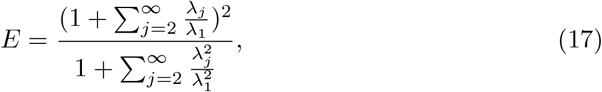

where *λ*_*j*_ are eigenvalues of Cov(*X*(·)) which are computed in FPCA. For GWAS, we use a Bonferroni threshold of genome-wide significance 5 × 10^*−*8^*/E*. And for heritability and genetic correlation, the Bonferroni threshold is 0.05*/E*.

### The pipeline of RVGA

The procedure of using RVGA to conduct voxel-level GWAS and secondary analysis includes the following steps. Refer to “The pipeline of RVGA” section in the Supplementary Note for more details.

#### Image loading

Images can be in NIFTI, CIFTI, FreeSurfer morphometry data, or text file formats. An additional image or text file for coordinate information is required. Non-imaging phenotypes can also be analyzed using RVGA.

#### FPCA

Raw images are initially subjected to kernel smoothing. The optimal bandwidth for smoothing is selected adaptively from a candidate list based on the GCV score. The images are then processed with `IncrementalPCA` to compute functional bases and eigenvalues. RVGA generates a table summarizing the number of LDRs required to preserve various proportions of image variance, offering guidance on selecting an appropriate number of LDRs.

#### LDR construction

Raw images, bases, covariates, and the number of LDRs are input. RVGA constructs the designated number of LDRs and computes the variance-covariance matrix of covariate-effect-removed LDRs. RVGA prints a table of mean correlation between raw and reconstructed images using varying numbers of LDRs.

#### LDR GWAS

LDR GWAS can be internally conducted.

#### GWAS summary statistics processing

RVGA processes raw LDR summary statistics and saves in a single file for fast voxel-level GWAS reconstruction and secondary analyses. Specifically, RVGA removes SNPs from LDR or non-imaging phenotype GWAS summary statistics if they exhibit any of the following characteristics: 1) a duplicated rsID; 2) an ambiguous strand; 3) an effective sample size less than 0.67 times the 90th percentile of the sample size; 4) multiple alleles; or 5) a missing or infinite z-score.

#### Voxel-level GWAS reconstruction

RVGA is flexible in reconstructing voxel-level summary statistics for different scenarios: 1) scanning the whole genome and all voxels, and saving only significant associations that pass a provided threshold; 2) conducting analysis for the whole genome and a subset of voxels; 3) conducting analysis for selected variants or a genome segment across all voxels.

#### Post-GWAS screening

RVGA computes an empirical null distribution of cluster size, defined as the number of associated voxels for a SNP, using the wild bootstrap approach (refer to the Supplementary Note for specific algorithm). Users can manually exclude SNPs with a small cluster size based on the quantile at level 1 − 0.05/(number of loci * effective number). The remaining voxel-variant associations can be aggregated into loci and reported.

#### LD matrix estimation

RVGA estimates the LD matrix and its inverse from a pair of PLINK bfiles based on a specified regularization level.

#### Heritability and (cross-trait) genetic correlation analysis

The inputs include processed LDR summary statistics, processed non-imaging phenotype summary statistics (optional), the LD matrix and its inverse, bases, and the variance-covariance matrix of covariate-effect-removed LDRs.

### Computational complexity

For an imaging-genetic study with *N* total voxels, *r* LDRs, *p* covariates, *n* subjects, *n*_1_ external subjects, *m* LD blocks, and *d* SNPs, the computational complexities are as follows:

1. Adaptively selecting the optimal bandwidth and estimating a sparse smoothing matrix using the local-linear method take the maximum *O*(*N* ^2^ + *nN*) time and *O*(*N* ^2^ + *nN*) memory.
2. Randomized SVD for the smoothed imaging data takes the maximum *O*(*nN* min {*n, N*}) time and *O*(*N* ^2^ + *nN*) memory.
3. Constructing LDRs takes *O*(*Nnr*) time; computing the variance-covariance matrix of LDRs takes *O*(*r*^2^*n* + *rpn* + *p*^3^ + *r*^2^*p*) time. Storing the data (including the top *r* bases) takes *O*(*nr* + *Nr* + *r*^2^) storage space.
4. Computing summary statistics for LDRs takes *O*(*dnr*) time and *O*(*dr*) storage space.
5. Assume each block in the LD matrix has *O*(*d/m*) SNPs, then estimating the whole LD matrix takes *O*(*n*_1_*d*^2^*/m*) time and *O*(*d*^2^*/m*) storage space.
6. Computing heritability and genetic correlation takes *O*(*rd*^2^*/m* + *N* ^2^*r*) time. Specifically, 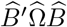 takes *O*(*rd*^2^*/m*+*r*^2^*d*) time, 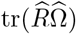 takes *O*(*d*^2^*/m*) time, and mapping the results on the low-dimensional space to the original space takes *O*(*Nr*^2^ + *N* ^2^*r*) time. Saving heritability and genetic correlation estimates takes *O*(*N*) and *O*(*N* ^2^) storage space, respectively.

Cumulatively, the proposed method takes the maximum *O*(*nN* min {*n, N*}+ *dnr* + *n*_1_*d*^2^*/m*) time, *O*(*N* ^2^ + *nN*) memory, and *O*(*N* ^2^ + *dr* + *d*^2^*/m*) storage space. In the “The pipeline of RVGA” section in the Supplementary Note, we outline the input and output of each step in the pipeline, along with general strategies to enhance computational efficiency.

### Imaging acquisition and feature generation

We obtained structural MRI (sMRI) and diffusion MRI (dMRI) imaging data through UK Biobank application 22783. The UKB team has already preprocessed and quality controlled the data before releasing. For detailed information on image acquisition and preprocessing, refer to the protocol (https://biobank.ctsu.ox.ac.uk/crystal/crystal/docs/brain_mri.pdf). Using FMRIB Software Library (FSL v5.0.9) (https://fsl.fmrib.ox.ac.uk/), sMRI were linearly registered into a strandard brain space (MNI152). We segmented the left and right hippocampus from sMRI and computed radial distances for each vertex. The radial distance represents the Euclidean distance between each vertex and the medial axis, the geometric center of the isoparametric curve. Both left and right hippocampus images had a resolution of 100 × 150 × 1 (15, 000 vertices). The pipeline is available at https://www.nitrc.org/frs/?group_id=1461. The detailed steps can be found in the Supplementary Note of ref [12].

Regarding dMRI features, each individual fractional anisotropy (FA) image was generated by fitting diffusion tensor imaging (DTI) models using the FSL software. Our analysis employed the ENIGMA-DTI pipeline, a standardized set of procedures for processing DTI data (http://enigma.ini.usc.edu/protocols/dti-protocols/). We first linearly registered each FA image to the ENIGMA FA template, which was set at a 1×1×1 mm^3^ spatial resolution in the MNI152 standard space. We then applied nonlinear registration techniques to further refine the alignment of the FA images to the standard space and masked the registered FA images using the template brain mask. We manually checked the registration performance and removed those with low quality. The ENIGMA skeleton, representing the major WM pathways, was projected onto the registered images. We finally extracted 21 predefined WM tracts according to the JHU ICBM-DTI-81 WM atlas. Tract resolutions ranged from 88 (inferior fronto-occipital fasciculus) to 3, 503 voxels (superior longitudinal fasciculus). More details for implementing the pipeline can be found in ref [10].

### Real data analysis

We collected the UKB phases 1 to 3 sMRI and dMRI data for 35,000 subjects of European ancestry. To remove related subjects, we computed genetic relatedness matrix using the genotype array data of autosomes with 460,000 common SNPs (MAF *>* 0.01) through GCTA [62] (v1.93.2beta, https://yanglab.westlake.edu.cn/software/gcta/#Overview). We removed one of a pair of related subjects with the threshold 0.05 (`--grm-cutoff` 0.05), which excluded the fourth and more related relatives. We further excluded subjects with excessive heterozygosity (Field ID 22027), discrepancies between reported and genetic gender (Field ID 22001), potential sex chromosome anomalies (Field ID 22019). The final sample size was 33,324.

The LDRs were constructed to capture more than 80% of the variance for each WM tract. For hippocampus, we captured 90% of variance due to rapid decay of eigenvalues. The LDRs were adjusted for the intercept, age, sex, age^2^, age × sex, age^2^ × sex, assessment center (Data Field 54), and 40 genetic PCs to estimate the variance-covariance matrix.

Imputed genotype data was used in LDR GWAS. During quality control, we removed SNPs with 1) an MAF less than 0.01; 2) a *p*-value of Hardy–Weinberg equilibrium (HWE) test less than 10^*−*7^; 3) a missing call rate greater than 10%; and 4) an imputation score less than 0.9, resulting in approximately 7.8 million autosomal SNPs.

After processing LDR GWAS summary statistics, we reconstructed voxel-level summary statistics and saved only significant signals. The significance threshold was determined by the effective number (Methods, Table S2). For hippocampus, we considered left and right hemispheres altogether and the significance threshold was 4.94 × 10^*−*9^. We considered all WM tracts altogether and the threshold was 1.91 × 10^*−*10^. We extracted 150,000 independent SNPs with a pairwise LD less than 0.1 from 460,000 genotyped SNPs in UKB and computed a null distribution of cluster size using the wild bootstrap approach with 50 bootstrap samples, resulting 7,500,000 points (Methods and Supplementary Note). SNPs with a cluster size less than the quantile at 1 − 0.05/(number of loci * effective number) were excluded. Note the number of loci can be determined by aggregating all voxel-variant associations before screening.

We applied the Peaks algorithms [8] to group the remaining voxel-level associations within each ROI. Specifically, a locus is defined as a collection of significant voxel-variant pairs such that the distance from any variant in the locus to the most significant variant is less than 0.25 cM. Any voxel-variant pair can be included in one and only one ocus. A locus in previous studies was replicated by our study if the most significant variant in that locus was within 0.25 cM from any of the most significant variants n our loci. For WM tracts, we compared with previous studies using UKB WM FA traits [8, 10]. For hippocampus, we checked our previous study [12] and all the hippocampus-related associations published on NHGRI-EBI GWAS catalog (up to April 2024).

We estimated LD matrices including genotyped SNPs or imputed HapMap3 SNPs. For genetyped SNPs, two datasets of unrelated white subjects from UKB were extracted, each with a sample size of 8, 400 and no sample overlap. SNPs in the major histocompatibility complex (MHC, GRCh37: chr6 28.5M - 33.5M) and/or those with an MAF less than 0.01 and/or a *p*-value of HWE test less than 10^*−*7^ and/or a missing call rate greater than 10% were removed, resulting in approximately 460, 000 SNPs.

The genome was partitioned into 1, 703 approximately independent LD blocks [60]. Missing values in a SNP vector were imputed by the sample mean. The LD matrix for each block was first estimated by sample correlation, then subject to eigen decomposition and 85% and 80% of variance were preserved for the LD matrix and its nverse, respectively. To enhance statistical efficiency in cross-trait genetic correlation estimates, we used a more restrictive regularization of {75%, 70%}. For imputed HapMap3 SNPs, we used two datasets each with a sample size of 42, 000. The SNPs were screened using the same procedure, resulting in 1, 160, 000 SNPs in analysis. The regularization was {98%, 95%} for heritability and genetic correlation and {90%, 85%} for cross-trait genetic correlation.

We gathered summary statistics for 11 complex brain disorders, including Alzheimer’s disease [63], schizophrenia [64], insomnia [36], autism spectrum disorder (ASD) [65], bipolar disorder (BD) [66], major depressive disorder (MDD) [67], attention-deficit hyperactivity disorder (ADHD) [68], neuroticism, neuroticism subclusters (depressed affect and worry), and depression [69]. Additionally, we included three brain-related phenotypes: educational attainment [40], cognitive performance 40], and intelligence [70]. Detailed information for each dataset is provided in Table S9. Most summary statistics were generated through meta-analysis on multiple cohorts and consortiums. For those involving UKB cohorts, we accounted for potential sample overlap in RVGA.

### Validation of the RVGA voxel-level GWAS results

We compared voxel-level summary statistics produced by RVGA to those from VGWAS (PLINK2) using correlation and RMSE of beta coefficients and z-scores. In the first phase, we randomly selected 100 variants for analysis in the left hippocampus and another 100 variants for the superior fronto-occipital fasciculus, where the first 10 variants in each ROI were randomly selected from significant loci. In the second phase, we randomly selected 100 points from the above two ROIs and scanned the whole genome using 460,000 genotyped SNPs.

We collected UKB phases 4 to 6 sMRI and dMRI data, including 20,130 unreated subjects of European ancestry, for replication. The imaging and genotype data were processed using the same pipelines and thresholds as before. We independently estimated functional bases while adjusted for the identical list of covariates. Specifically, we extracted all SNPs in significant loci from the discovery phase and evaluated them in the replication study. Any SNPs in a locus being significant in the replication study indicated the locus was replicated. We used two thresholds in replication: 0.05/(number of loci) and 0.05/(number of loci * effective number). Refer to the Supplementary Note for a brief review of replication studies in literature, and more details about the replication study.

### Evaluation of the RVGA heritability and genetic correlation estimates

We compared RVGA heritability estimates to those from SumHer [42]. We downloaded pre-computed taggings of white subjects produced by using UKB genotyped SNPs and HapMap3 SNPs, respectively, corresponding to the “BLD-LDAK” model (https://dougspeed.com/pre-computed-tagging-files/). RVGA genetic correlation and cross-trait genetic correlation estimates were compared to those from LDSC [44]. For genotyped SNPs, we estimated LD scores using a genotype array dataset of sample size 8, 400 through the LDSC command line tool (https://github.com/bulik/ldsc?tab=readme-ov-file#ldsc-ld-score-v101), setting LD window as 10, 000 Kb (`--ld-wind-kb 10000`). For HapMap3 SNPs, we downloaded pre-computed LD scores from the 1000 Genome data. SumHer and LDSC were directly applied to each voxel using the summary statistics from VGWAS. In cross-trait genetic correlation analysis, if the cohort in non-imaging phenotypes had no evidence of sample overlap with the UKB cohort, we employed the RVGA estimator without correction, and constrained the intercept in cross-trait LDSC as 0 (`--intercept-gencov 0,0`). Otherwise, we employed the RVGA estimator considering sample overlap and did not constrain the LDSC intercept. We calculated Pearson correlation coefficient and relative confidence interval width, defined as the mean standard error of RVGA estimates to that of SumHer/LDSC estimates across 100 points.

### Simulation studies

We employed real genotype array data or imputed genotype data (HapMap3 SNPs) from UKB in the simulation. We randomly extracted *n* = 10, 000 unrelated white subjects and all common variants (MAF *>* 0.01) on chromosome 10 (23, 203 genotyped SNPs) or on chromosome 19 (21, 576 imputed SNPs). We standardized each SNP by the formula 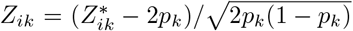, where 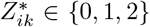 is the number of reference alleles and *p*_*k*_ is the sample MAF.

To simulate imaging data, we employed a varying coefficient model

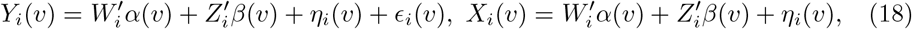

where *i* ∈ {1, …, *n*} and 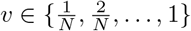. Moreover, *W*_*i*_ is a 2 × 1 covariate vector, where the first covariate was sampled from *Bernoulli*(0.5) and the second one was sampled from *𝒩* (0, 0.01). Each fixed covariate effect 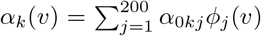, where *α*_0*kj*_ ~ *𝒩* (0, (*j* + 3)^*−*3^) for *k* = 1, 2.

A set of SNPs *𝒞* were randomly chosen as causal SNPs. For *k* ∈ *𝒞*, 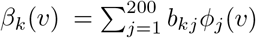, with 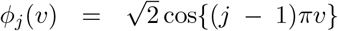 and 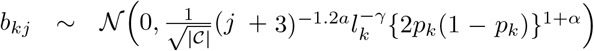, where |*𝒞*| is the number of causal SNPs; *a >* 1 is a polynomial decay rate; *γ* ∈ {1, 0} specifies whether effect sizes are related to the LD weights 1*/l*_*k*_; *α* ∈ {−1, −0.25} indicates strong/weak inverse relationship between MAF and effect sizes. For *k* ∉ C, *β*_*k*_(·) = 0. The underlying imaging data without white noise was decomposed as 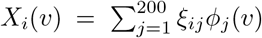 with *ξ*_*ij*_ *~ 𝒩* (0, *λ*_*j*_), where *λ*_*j*_ = 2(*j* + 2)^*−a*^. The non-genetic effect 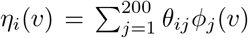, where *θ*_*ij*_ ~ 𝒩(0, max *λ*_*j*_ Var(*Wα*_0*j*_ + *Zb*_*j*_), 0). We adjusted the genetic variance of each voxel to achieve a fixed heritability level. Finally, we generated *ϵ*_*i*_(*v*) following *𝒩* (0, *σ*^2^) (normal noise) or Rayleigh distribution with parameter *Σ*, where *Σ*^2^ was adaptively set for a specific noise percentage *w* = *σ*^2^*/*(Var(*X*(·)) + *σ*^2^).

For assessing type I error rate, we used the reduced model without genetic effect 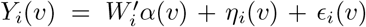 and set *N* = 200, *n* = 10, 000, *a* = {1.8, 2.5}, *w* = {50%, 20%, 0}, and the proportion of variance to keep with LDRs *q* = {0.75, 0.80, 0.85, 0.90, 0.95}. For each scenario, 1, 000 replicates were generated.

We investigated the following scenarios for statistical power: *N* = 200, *n* = 10, 000, |*𝒞*|*/d* = {1%, 20%} (proportion of causal SNPs or polygenicity), *h* = {0.03, 0.1, 0.3}, *α* = −1, *γ* = 0, *a* = {1.8, 2.5}, *w* = {50%, 20%, 0}, noise following *𝒩* (0, *Σ*^2^) or Rayleigh distribution with scale *Σ*, and the proportion of variance to keep with LDRs *q* = {0.75, 0.80, 0.85, 0.90, 0.95}. For each scenario, 100 replicates were generated.

For evaluating heritability and genetic correlation estimates, we considered the following scenarios: *N* = 200, *n* = 10, 000, |*𝒞*| */d* = {1%, 20%}, *h* = {0.03, 0.1, 0.3} (*h* = 0.03 only for heritability as it is too small to get stable genetic correlation estimates), *α* = {−1, − 0.25}, *γ* = {1, 0}, *a* = 1.8, *w* = 20%, normal noise, and the proportion of variance to keep with LDRs *prop* = {0.75, 0.80, 0.85, 0.90, 0.95}. For each scenario, 100 replicates were generated.

We further simulated a single trait *U* to evaluate cross-trait genetic correlation estimates. The true genetic effect *β*_*U*_ was simulated by 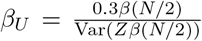, and the observed data *U* = *Z*_*U*_ *β*_*U*_ + *ϵ*_*U*_ was generated, where *Z*_*U*_ was extracted from the real data with no sample overlap with *Z* and *ϵ*_*U*_ ~ *𝒩* (0, Var(*Z*_*U*_ *β*_*U*_ (*v*))*I*). That indicates the heritability of *U* was fixed at 0.5.

The LD matrix and its inverse were independently generated each using 10, 000 unrelated white subjects. For genotyped SNPs, four regularization levels for LD matrix and its inverse were assessed: {0.90, 0.85}, {0.85, 0.80}, {0.80, 0.75}, and {0.75, 0.70}. For imputed SNPs, the setup was {0.98, 0.95}, {0.95, 0.90}, {0.90, 0.85}, and {0.85, 0.80}.

The type I error was evaluated by the proportion of null tests with a *p*-value less than 10^*−*2^, 10^*−*3^, and 10^*−*4^. The statistical power was quantified by the proportion of causal variants identified at level 10^*−*4^. The distance between RVGA summary statistics and those from VGWAS was evaluated by RMSE. For heritability and (crosstrait) genetic correlation analysis, we employed MAE to compare the estimates with the ground truth. We additionally compared the average estimate to the average true value across all voxels.

## Supplementary information

Supplementary Note, Figures, and Tables can be accessed online.

## Acknowledgments

We thank research assistants (X. Qian, S. Huang, P. Guan, J. Chen, M. Hu, X. Qi, B. Tang, and V. Pathare) in the UNC BIGS2 team for downloading and preprocessing raw images. We thank the individuals represented in the UKB study for their participation and the research teams for their work in collecting, processing and disseminating these datasets for analysis. We thank University of North Carolina at Chapel Hill and the Research Computing groups for providing computational resources and support that have contributed to the research results. This research has been conducted using the UK Biobank resource (application number 22783), subject to a data transfer agreement. We thank S. Gao from Yunnan University (China) for helping polish the figures. We used imaging icons from BioRender.com in Figure 1.

## Declarations

### Ethics approval

The UKB has obtained ethics approval from the Northwest Multi-Centre Research Ethics Committee (MREC, approval number: 11/NW/0382), and obtained written informed consent from all participants prior to the study.

### Consent to participate

Not applicable

### Consent for publication

Not applicable

### Availability of data and materials

The triplets of summary statistics as well as LD matrix are shared at Zenodo [71], https://doi.org/10.5281/zenodo.13787684. Raw structural MRI and diffusion MRI imaging data was obtained from UK Biobank application 22783, https://www.ukbiobank.ac.uk. NHGRI-EBI GWAS catalog, https://www.ebi.ac.uk/gwas/. Links to summary statistics: schizophrenia, https://figshare.com/articles/dataset/scz2022/19426775?file=34517828; Alzheimer’s disease, https://ctg.cncr.nl/documents/p1651/PGCALZ2sumstatsExcluding23andMe.txt.gz; insomnia, https://ctg.cncr.nl/documents/p1651/insomnia_ukb2b_EUR_sumstats_20190311_with_chrX_mac_100.txt.gz; educational attainment, https://thessgac.com/papers/3; cognitive performance, https://thessgac.com/papers/3; autism spectrum disorder, https://pgc.unc.edu/for-researchers/download-results/; bipolar disorder https://pgc.unc.edu/for-researchers/download-results/; major depressive disorder, https://pgc.unc.edu/for-researchers/download-results/; attention deficit hyperactivity disorder, https://pgc.unc.edu/for-researchers/download-results/; intelligence, https://ctg.cncr.nl/documents/p1651/SavageJansen_IntMeta_sumstats.zip; neuroticism, https://ctg.cncr.nl/documents/p1651/sumstats_neuroticism_ctg_format.txt.gz; depression, https://ctg.cncr.nl/documents/p1651/sumstats_depression_ctg_format.txt.gz; depression affect subcluster, https://ctg.cncr.nl/documents/p1651/sumstats_depressed_affect_ctg_format.txt.gz; worry subcluster, https://ctg.cncr.nl/documents/p1651/sumstats_worry_ctg_format.txt.gz.

### Code availability

RVGA is a component in Highly Efficient Imaging Genetics (HEIG) toolbox which is a comprehensive solution for voxel-level imaging genetic analysis. HEIG (v1.2.1) is an open-source Python software, https://github.com/Zhiwen-Owen-Jiang/HEIG with tutorial, https://github.com/Zhiwen-Owen-Jiang/HEIG/wiki and example data, https://doi.org/10.5281/zenodo.14214075; The example code for reproducing the main results, https://github.com/Zhiwen-Owen-Jiang/HEIG/tree/pub/misc/code_manuscript/code.md; LDSC (v1.0.1), https://github.com/bulik/ldsc; SumHer, https://dougspeed.com/sumher/; GCTA (v1.93.2beta), https://yanglab.westlake.edu.cn/software/gcta/#Overview; PLINK2 (v2.00a3LM), https://www.cog-genomics.org/plink/2.0/); Peaks algorithm (v1.0), https://github.com/wnfldchen/peaks.

### Authors’ contributions

Z.J. and H.Z. proposed the concept and designed the methodology. H.Z. supervised the project. Z.J. performed the statistical analysis, visualized the results, developed and tested the software. T.L. designed the pipeline for preprocessing raw images. Z.J. drafted the initial manuscript. Z.J., J.S., P.S., E.F., Y.L., and H.Z. suggested revision ideas and revised the manuscript. All authors critically reviewed the manuscript and approved the final version.

## Extended Data

**Extended Data Fig.1.**
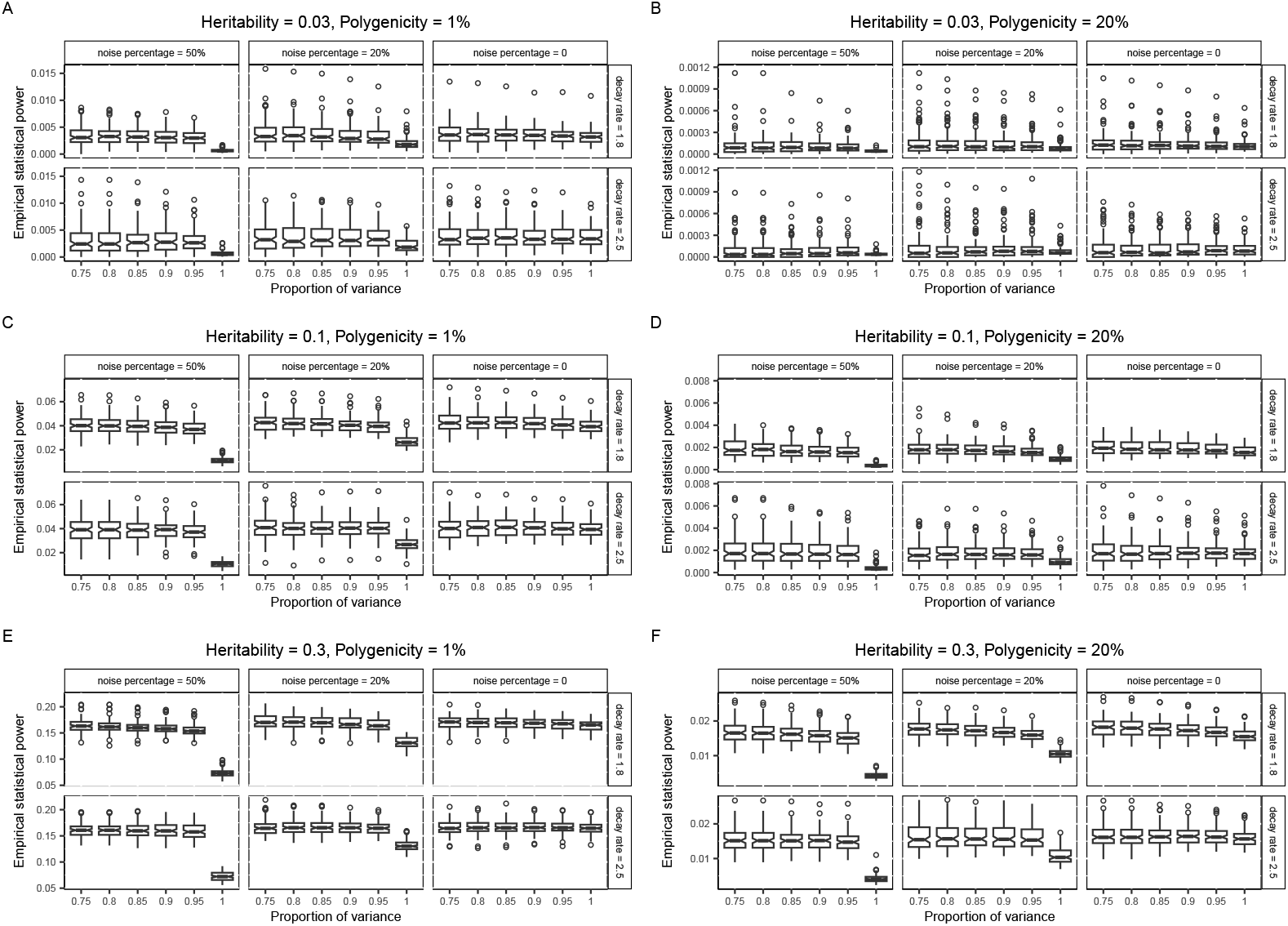
The empirical statistical power in simulation studies (Gaussian noise). The power is evaluated by the proportion of causal variants being significant at level 10^*−*4^. We used the same genotype data of 10,000 subjects and 21,576 common variants across all simulation setups. There are 216 causal variants for the polygenicity = 1% cases and 4,315 causal variants for the polygenicity = 20% cases. These two numbers are fixed, but the specific causal variants for each replicate are randomly selected. Each voxel in a replicate is affected by the same list of causal variants, but the true genetic effects are varying across voxels. Each point in a box plot represents the average empirical power across 200 voxels for a replicate and there are 100 replicates in each box plot. For example, if the empirical power is 0.01 for the polygenicity = 20% case, on average 4, 315 *×* 0.01 = 43.15 causal variants are detected for a voxel. The line in the box shows the median and the hinge represents an approximate 95% confidence interval around the median. Whiskers represent 1.5 times the interquartile range (IQR). The proportion of variance indicates the amount of imaging signals preserved by LDRs. **(A)** The scenario of low heritability and low polygenicity. **(B)** The scenario of low heritability and high polygenicity. **(C)** The scenario of medium heritability and low polygenicity. **(D)** The scenario of medium heritability and high polygenicity. **(E)** The scenario of high heritability and low polygenicity. **(F)** The scenario of high heritability and high polygenicity.

**Extended Data Fig.2.**
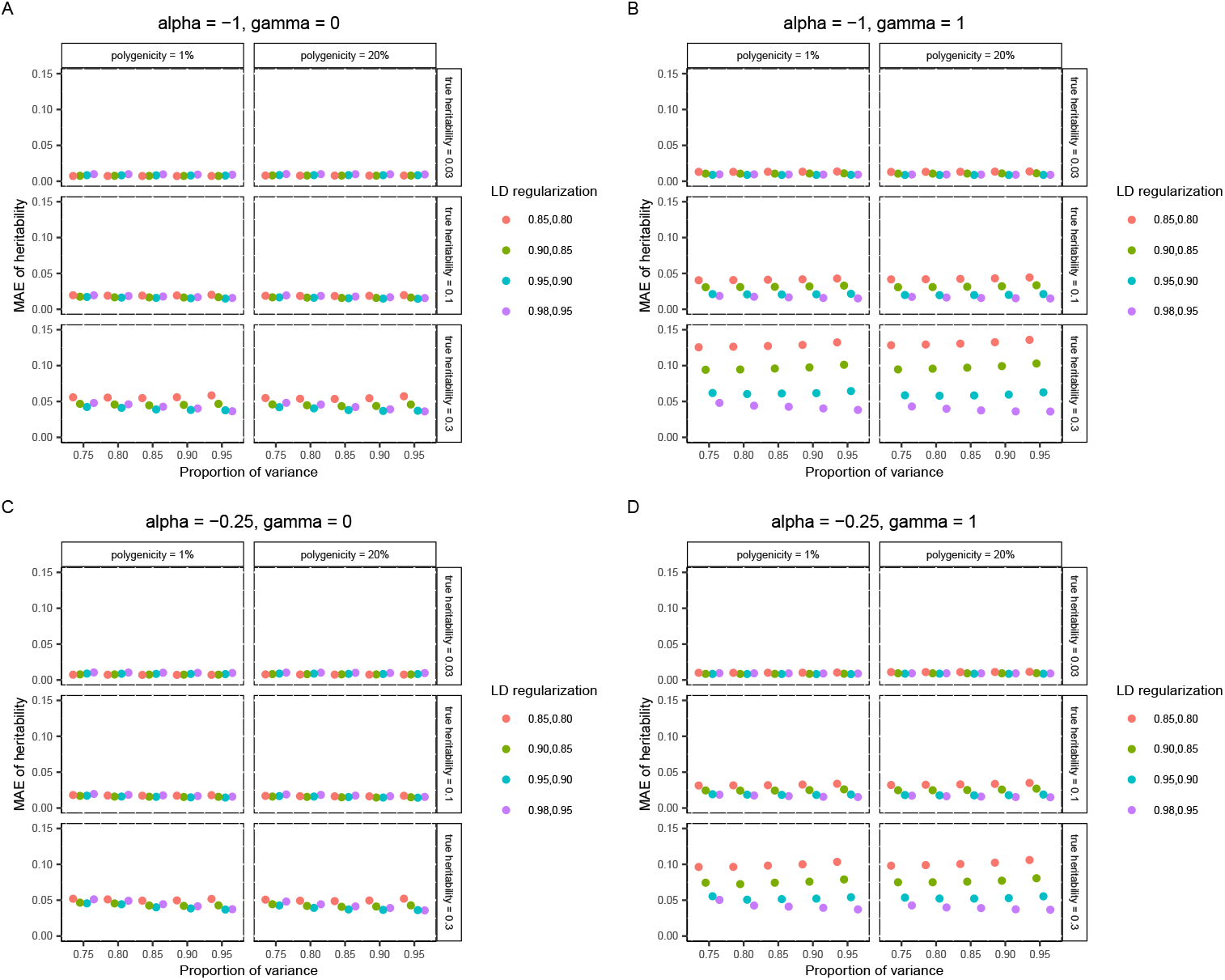
The mean absolute error of heritability estimates across all voxels in simulation studies (imputed SNPs). The proportion of variance indicates the amount of signals preserved by LDRs. LD regularization indicates the regularization imposed on the LD matrix and its inverse. **(A)** The genetic effects have a strong correlation with MAF but no dependence on the LD. **(B)** The genetic effects have a strong correlation with MAF and strong dependence on the LD. **(C)** The genetic effects have a weak correlation with MAF and no dependence on the LD. **(D)** The genetic effects have a weak correlation with MAF but strong dependence on the LD.

**Extended Data Fig.3.**
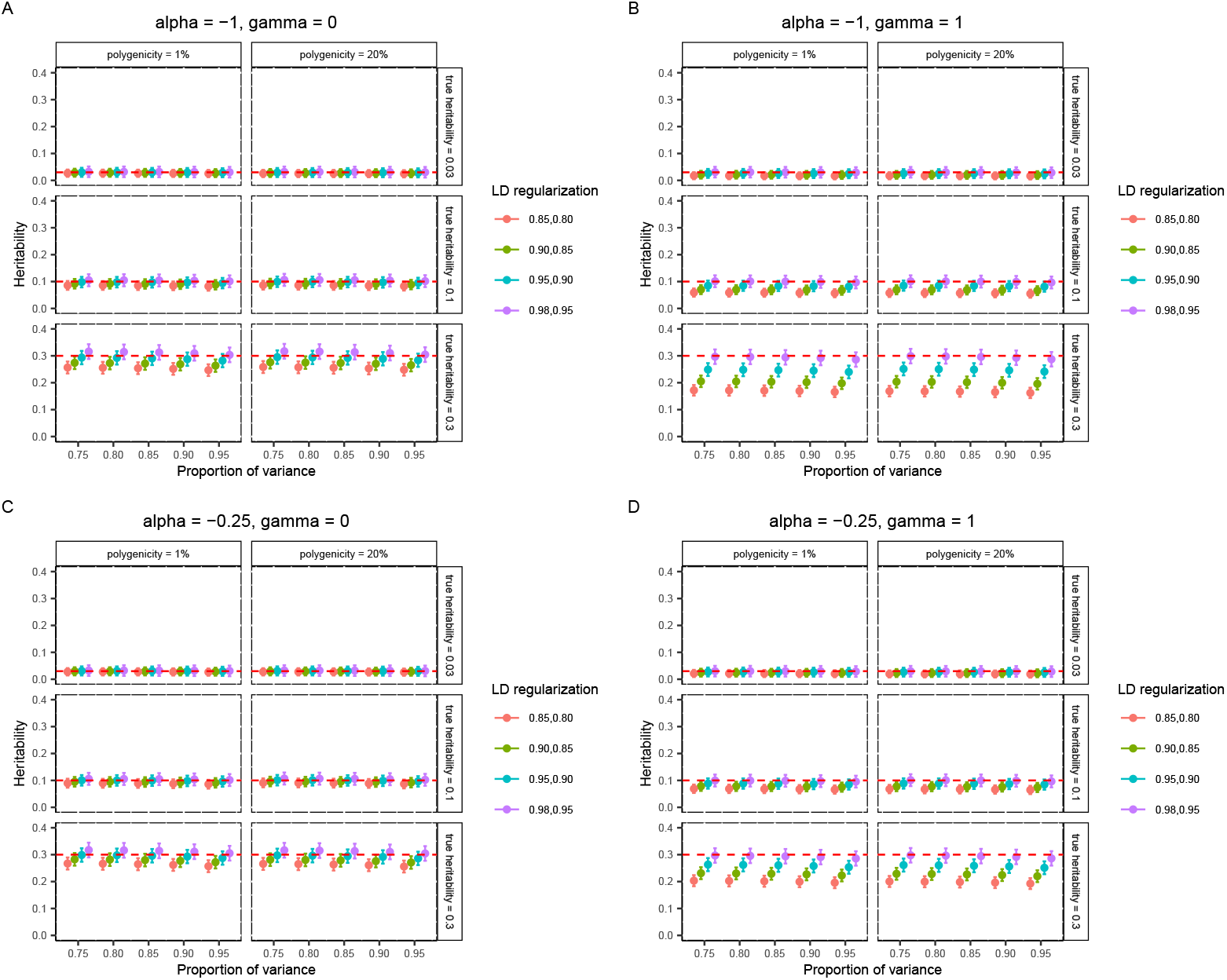
The mean and standard error of heritability estimates in simulation studies (imputed SNPs). The red dashed lines denote the true heritability of each voxel. The dots show the mean heritability estimates across all voxels. The error bars show the 95% confidence intervals constructed by mean estimates *±*1.96 mean standard error estimates across all voxels. The proportion of variance indicates the amount of signals preserved by LDRs. LD regularization indicates the regularization imposed on the LD matrix and its inverse. **(A)** The genetic effects have a strong correlation with MAF but no dependence on the LD. **(B)** The genetic effects have a strong correlation with MAF and strong dependence on the LD. **(C)** The genetic effects have a weak correlation with MAF and no dependence on the LD. **(D)** The genetic effects have a weak correlation with MAF but strong dependence on the LD.

**Extended Data Fig.4.**
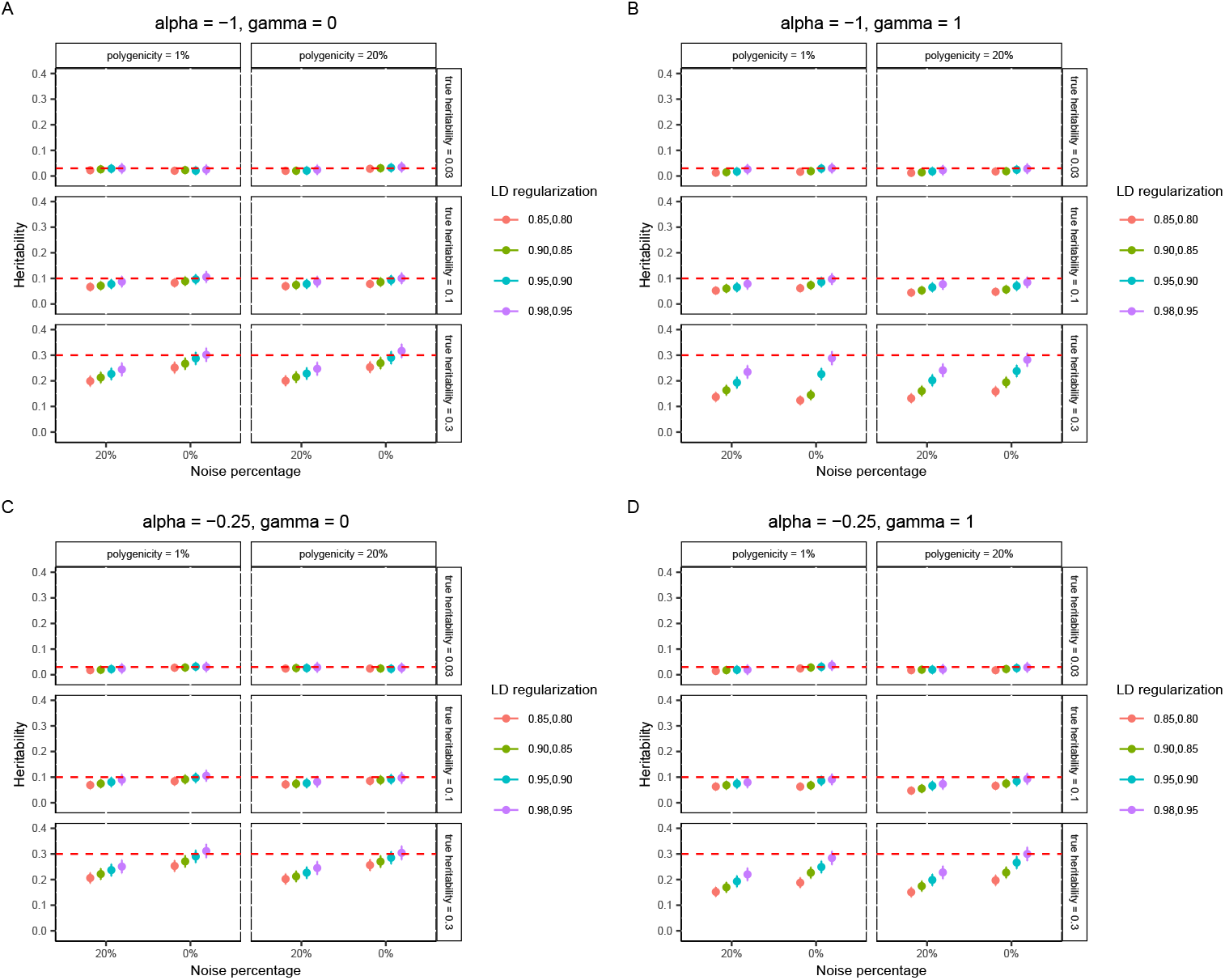
The mean and standard error of heritability estimates comparing images with and without noise (imputed SNPs). RVGA heritability estimator was applied to each voxel individually. The red dashed lines denote the true heritability of each voxel. The dots show the mean heritability estimates across all voxels. The error bars show the 95% confidence intervals constructed by mean estimates *±* 1.96 mean standard error estimates across all voxels. LD regularization indicates the regularization imposed on the LD matrix and its inverse. **(A)** The genetic effects have a strong correlation with MAF but no dependence on the LD. **(B)** The genetic effects have a strong correlation with MAF and strong dependence on the LD. **(C)** The genetic effects have a weak correlation with MAF and no dependence on the LD. **(D)** The genetic effects have a weak correlation with MAF but strong dependence on the LD.

**Extended Data Fig.5.**
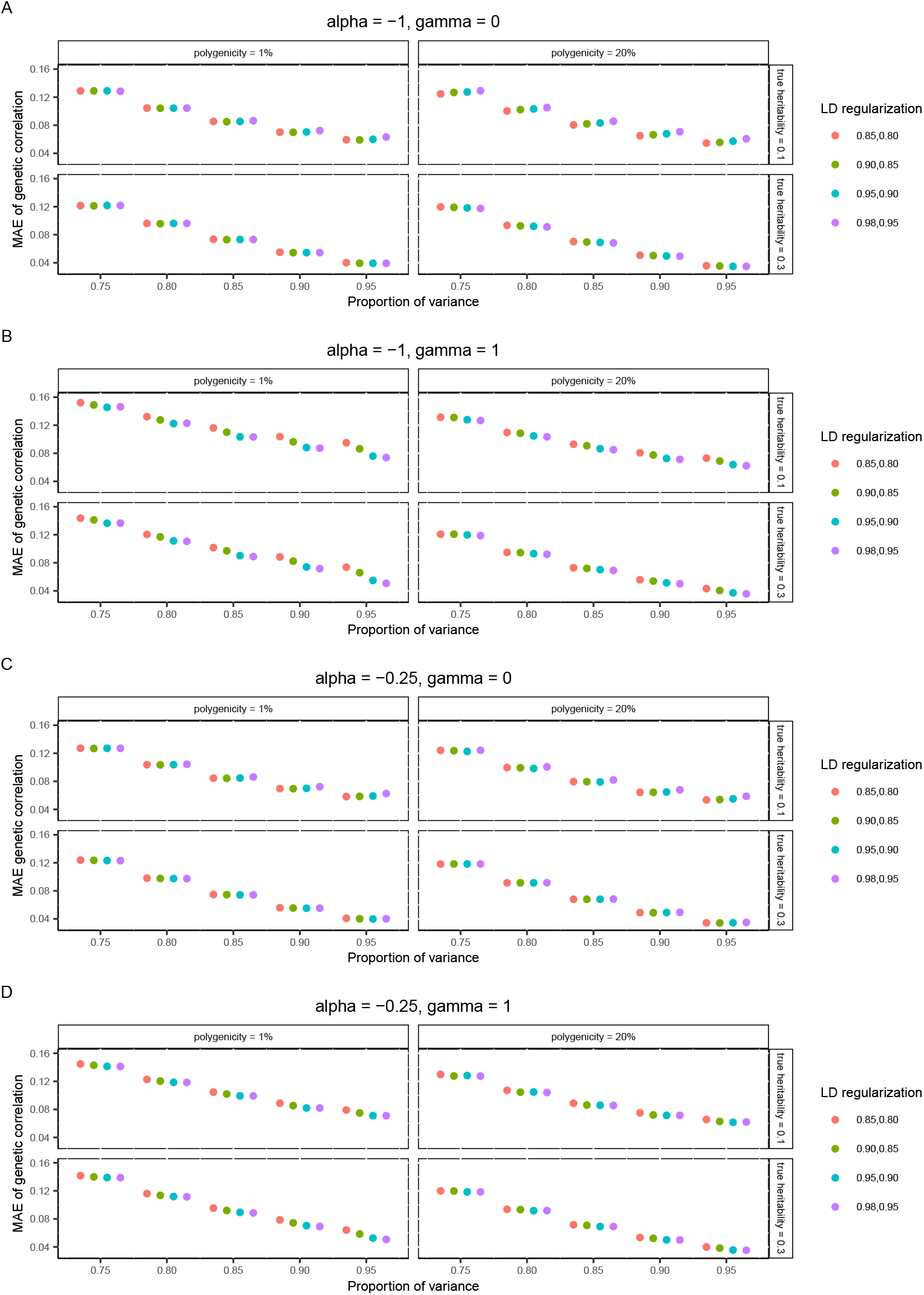
The mean absolute error of genetic correlation estimates across all voxel pairs in simulation studies (imputed SNPs). The proportion of variance indicates the amount of signals preserved by LDRs. LD regularization indicates the regularization imposed on the LD matrix and its inverse. **(A)** The genetic effects have a strong correlation with MAF but no dependence on the LD. **(B)** The genetic effects have a strong correlation with MAF and strong dependence on the LD. **(C)** The genetic effects have a weak correlation with MAF and no dependence on the LD. **(D)** The genetic effects have a weak correlation with MAF but strong dependence on the LD.

**Extended Data Fig.6.**
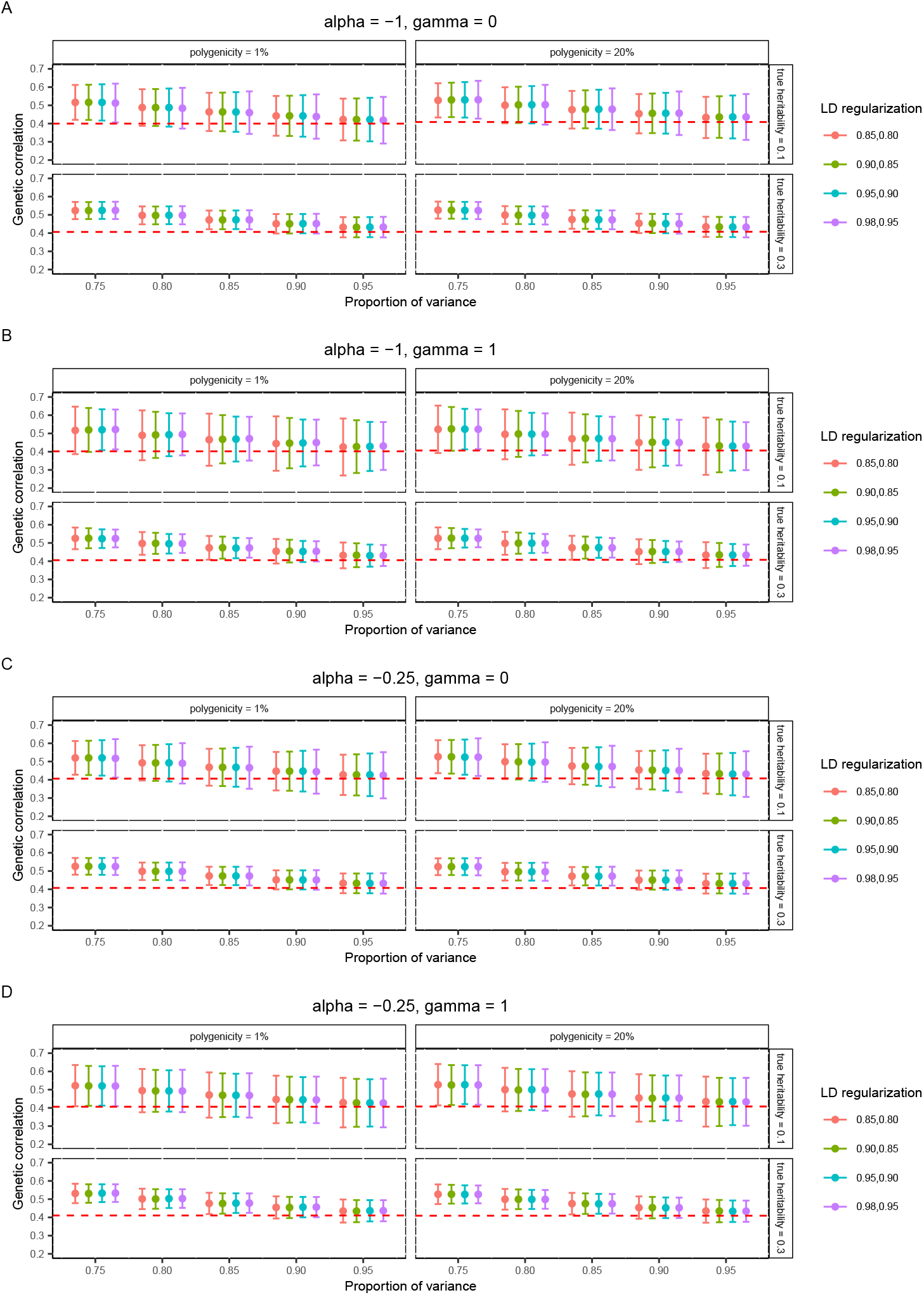
The mean and standard error of genetic correlation estimates in simulation studies (imputed SNPs). The red dashed lines denote the true mean genetic correlation across all voxel pairs. The dots show the mean genetic correlation estimates across all voxel pairs. The error bars show the 95% confidence intervals constructed by mean estimates *±*1.96 mean standard error estimates across all voxels. The proportion of variance indicates the amount of signals preserved by LDRs. LD regularization indicates the regularization imposed on the LD matrix and its inverse. **(A)** The genetic effects have a strong correlation with MAF but no dependence on the LD. **(B)** The genetic effects have a strong correlation with MAF and strong dependence on the LD. **(C)** The genetic effects have a weak correlation with MAF and no dependence on the LD. **(D)** The genetic effects have a weak correlation with MAF but strong dependence on the LD.

**Extended Data Fig.7.**
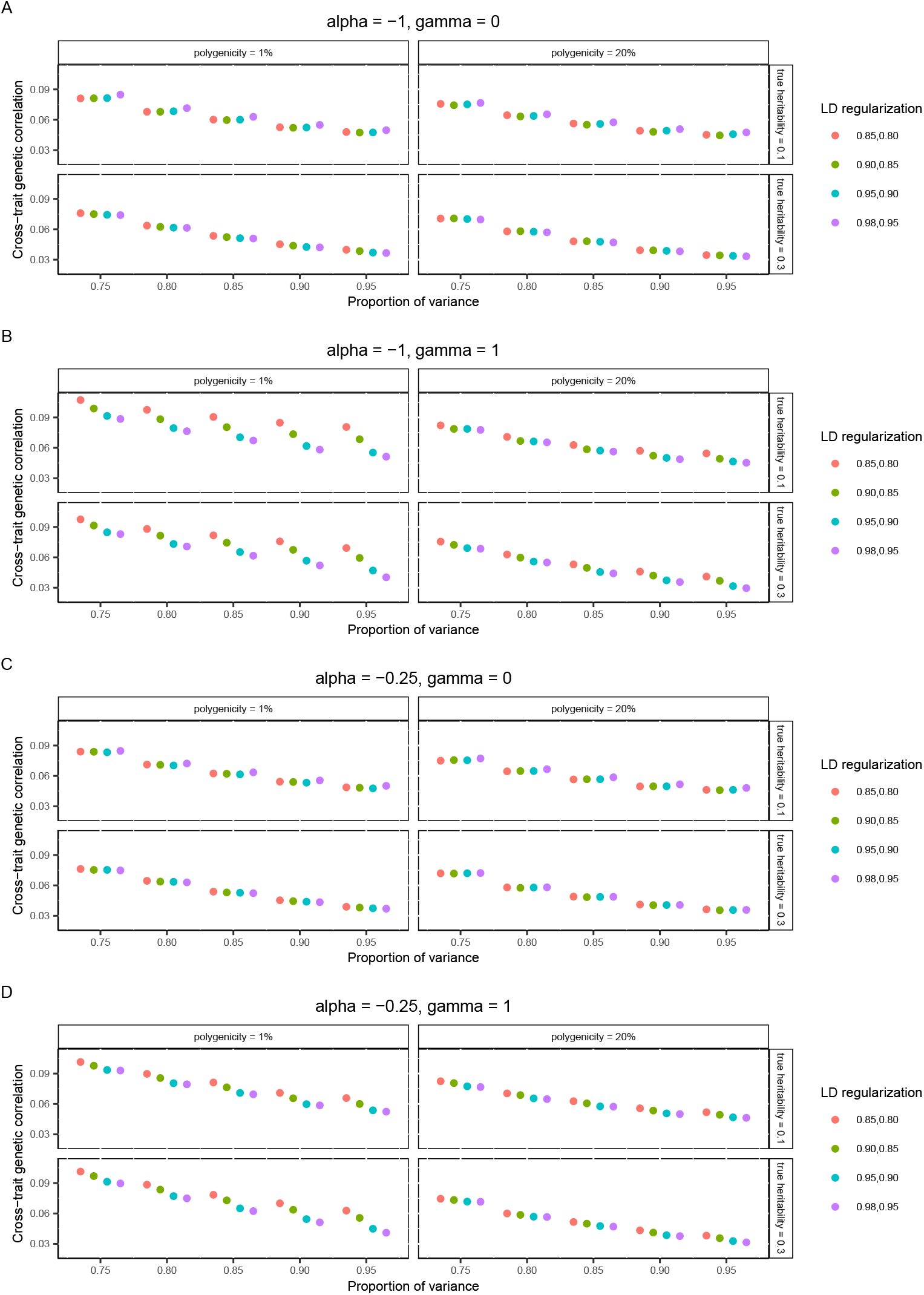
The mean absolute error of cross-trait genetic correlation estimates across all voxels in simulation studies (imputed SNPs). The proportion of variance indicates the amount of signals preserved by LDRs. LD regularization indicates the regularization imposed on the LD matrix and its inverse. **(A)** The genetic effects have a strong correlation with MAF but no dependence on the LD. **(B)** The genetic effects have a strong correlation with MAF and strong dependence on the LD. **(C)** The genetic effects have a weak correlation with MAF and no dependence on the LD. **(D)** The genetic effects have a weak correlation with MAF but strong dependence on the LD.

**Extended Data Fig.8.**
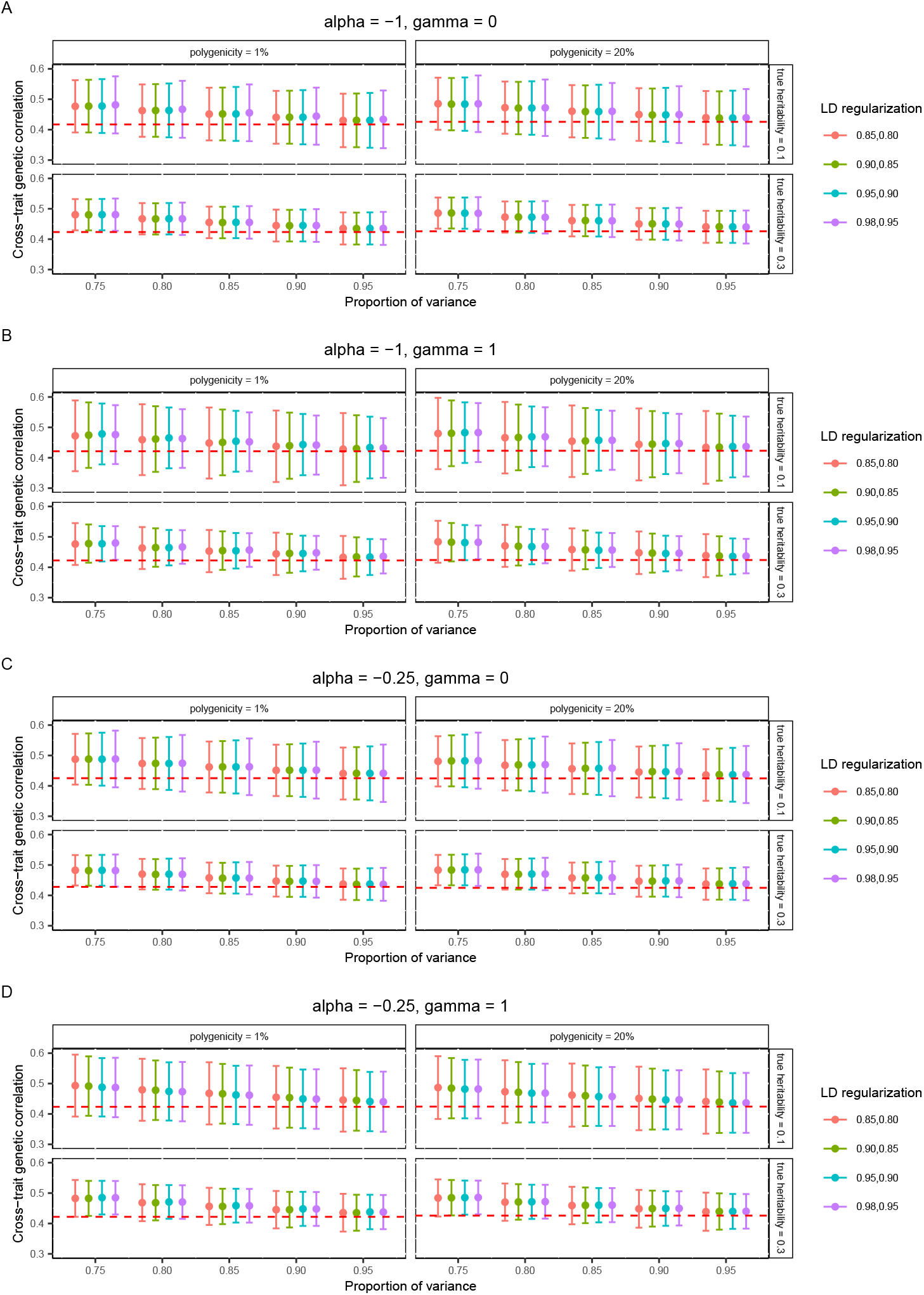
The mean and standard error of cross-trait genetic correlation estimates in simulation studies (imputed SNPs). The red dashed lines denote the true mean cross-trait genetic correlation across all voxels. The dots show the mean cross-trait genetic correlation estimates across all voxels. The error bars show the 95% confidence intervals constructed by mean estimates *±*1.96 mean standard error estimates across all voxels. The proportion of variance indicates the amount of signals preserved by LDRs. LD regularization indicates the regularization imposed on the LD matrix and its inverse. **(A)** The genetic effects have a strong correlation with MAF but no dependence on the LD. **(B)** The genetic effects have a strong correlation with MAF and strong dependence on the LD. **(C)** The genetic effects have a weak correlation with MAF and no dependence on the LD. **(D)** The genetic effects have a weak correlation with MAF but strong dependence on the LD.

**Extended Data Fig.9.**
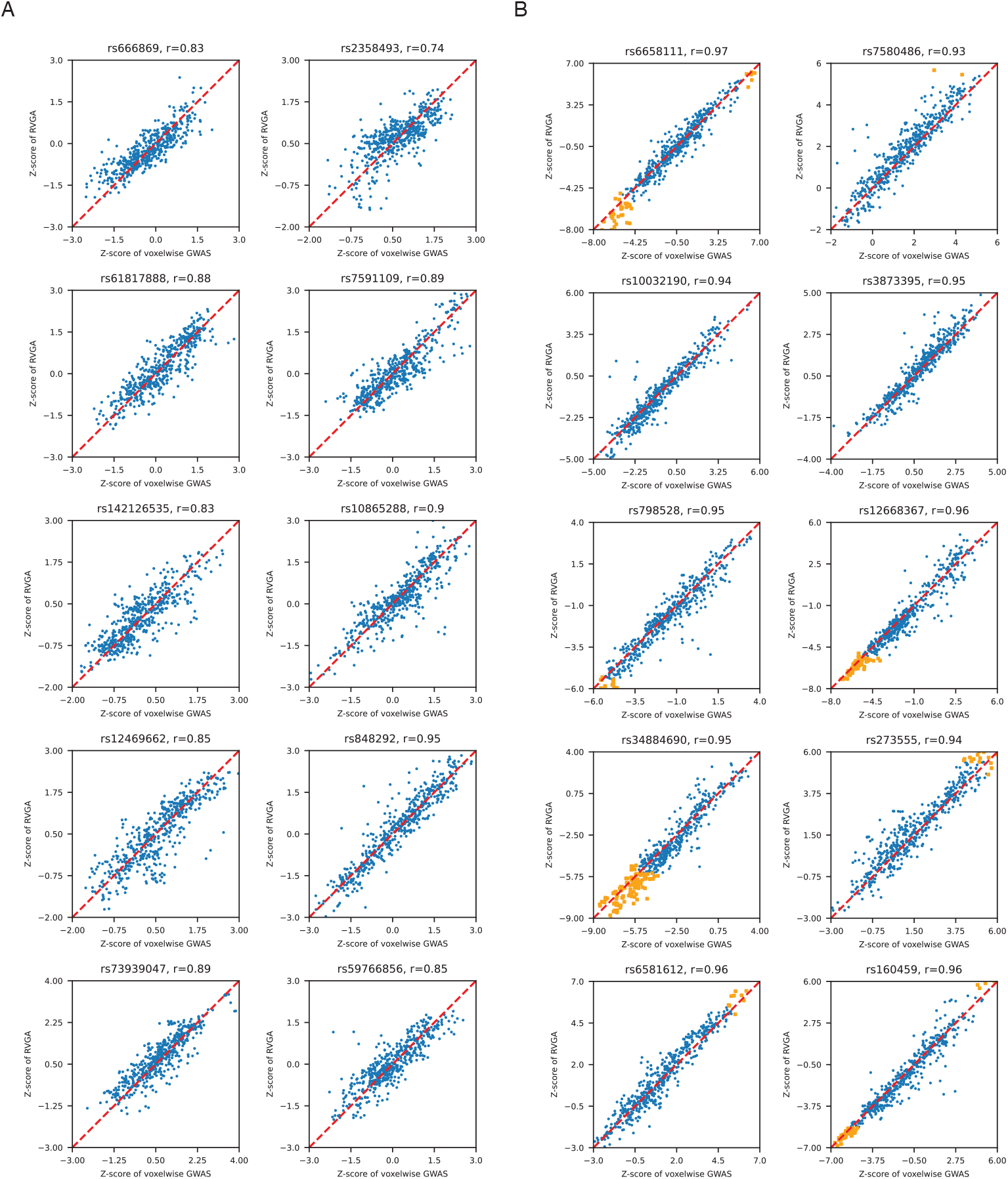
A Comparison of RVGA recovered vertex-wise z-scores and those directly estimated using VGWAS for the left hippocampus shape. Each subplot is a randomly selected variant. For the ease of visualization, we display 500 randomly selected vertices. “r” refers to Pearson correlation coefficient, which was calculated using all 15,000 vertices. A, Z-scores of randomly selected SNPs. B, Z-scores of SNPs randomly selected from significant loci. Nominally significant associations (*P <* 5 *×* 10^*−*8^) are highlighted.

**Extended Data Fig.10.**
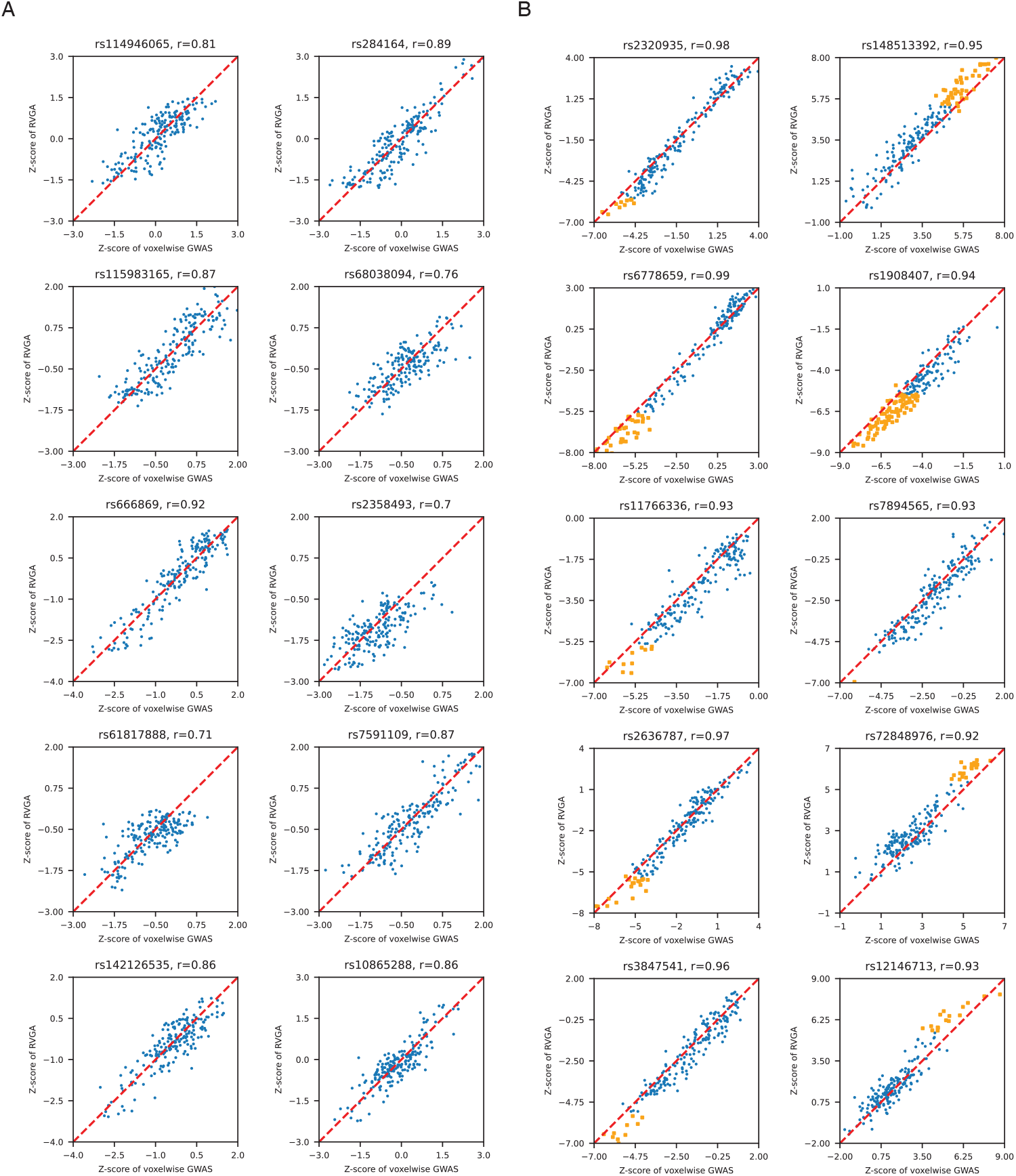
A Comparison of RVGA recovered voxel-wise genetic z-scores and those directly estimated using VGWAS for the superior fronto-occipital fasciculus. Each subplot is a randomly selected variant. We display all 193 voxels. “r” refers to Pearson correlation coefficient. A, Z-scores of randomly selected SNPs. B, Z-scores of SNPs randomly selected from significant loci. Nominally significant associations (*P <* 5 *×* 10^*−*8^) are highlighted.

