## Supplementary Materials for "Computation and resource efficient genome-wide association analysis for large-scale imaging studies"

**2 Supplementary Figures** **35**

**3 Supplementary Tables** **80**

### 1 Supplementary Note

#### 1.1 Vertex-level genome-wide association analysis for the cortical surface curvature

We collected T1-weighted structural MRI data from phases 1 to 3 of the UK Biobank (UKB). For the discovery phase of the genome-wide association analysis (GWAS), we utilized unrelated white subjects from phase 3 (n = 15,752), while the remaining dataset (n = 12,431) was used for replication. Cortical surface modeling was based on FreeSurfer analyses previously conducted by the UKB team, employing the `recon-all` preprocessing workflows. For each subject, maps of cortical thickness, local surface area, sulcal depth, and curvature were generated on the cortical surface. The cortical surfaces were then aligned using the `ciftify` package, which applied surface-based alignment of the cortical mesh using the MSM-Sulc algorithm [1]. This was followed by resampling to a common 32k mesh in standard MNI space with the medial wall excluded, resulting in 59,412 vertices in both hemispheres. To demonstrate RVGA's ability to efficiently perform vertex-wise analysis across the whole cerebral cortex, we randomly selected cortical surface curvature as an example.

In the discovery study, we conducted FPCA to compute all possible 15,752 PCs and constructed 1,750 LDRs, capturing approximately 80% of the variance with a correlation of 0.85 between the raw and reconstructed images. The effective number of independent vertices was 1585.3. We performed GWAS on the LDRs for 7.8 million common variants, followed by reconstructing vertex-level summary statistics using RVGA. At the Bonferroni-corrected threshold ($P < 5 \times 10^{-8}/1585.3 = 3.15 \times 10^{-11}$), we identified 1,431 significant loci by aggregating vertex-variant associations through the Peaks algorithm [2]. We further conducted cluster analysis on the 8,000 unique SNPs in these loci and excluded SNPs with a cluster size less than the quantile of the cluster size null distribution at $1 - 0.05/1,431$, resulting in 4,025 SNPs (Method). We used a less stringent threshold here due to a relatively small sample size. We aggregated the vertex-variant associations of these SNPs and ended up with 35 robust loci (Table S12).

We subsequently conducted a replication study and generated 1,750 LDRs. We extracted all significant variants ($P < 3.15 \times 10^{-11}$) from the discovery study and computed their summary statistics across all vertices. We first selected significant vertex-variant pairs in the replication study ($P < 0.0014 = 0.05/35$) and matched them with those in the discovery. The correlation coefficient of effect size estimates was 0.98, with 99.7% of effect size estimates showing consistent direction. We then aggregated vertex-variant associations into loci (Table S13) and all 35 loci from the discovery phase were replicated (Table S12). Using a more stringent threshold considering the number of loci and the effective number of independent vertices, all 35 loci from the discovery phase were replicated ($P < 0.05/35/1585.3$, Table S12). These loci were visualized in Fig. S18.

We examined the ROIs that contained at least one significant vertex-variant association ($P < 3.15 \times 10^{-11}$) based on the Desikan-Killiany-Tourville (DKT) atlas (Fig. S19). Most of the ROIs in the inferior regions were associated with genetic markers, while fewer associations were identified in the frontal area. Additionally, we calculated the percentage of vertices associated with at least one SNP for each ROI (Fig. S20). Notably, the postcentral, precentral, cuneus, and fusiform regions were the most enriched ROIs in both the left and right hemispheres. Specifically, 24% of the postcentral region was associated with SNPs.

We computed vertex heritability and annotated significant regions ($P < 0.05/1585.3$, Fig. S21) that were distributed across the entire brain. The postcentral, medial orbitofrontal, parahippocampal, and lateral orbitofrontal regions were relatively more heritable ($h > 0.25$), partially overlapping with the most enriched ROIs. The genetic correlation estimates in the image displayed a mean of 0.0 ($se = 0.23$), which was unstable due to the relatively small sample size and heritability estimates.

Finally, we assessed the genetic correlations between cortical surface curvature and 14 brain-related phenotypes (Table S14). We observed that worry (49), neuroticism (27), and depressed effect (14) showed the most associated vertices with a $p$-value less than 0.001.

#### 1.2 Algorithm of wild bootstrap for computing null distribution of cluster size and post-GWAS screening

The post-GWAS screening is based on evaluating cluster size, defined as the number of associated voxels for a SNP. The key idea is that reliable associations for a SNP should not be restricted to a small number of voxels given strong spatial correlations in images. Therefore, we want to exclude some SNPs after reconstructing voxel-variant associations and before aggregating them into loci. The detailed steps are as follows:

- Prepare a genotype dataset containing independent SNPs and the same group of subjects in the discovery phase. For example, there are 150,000 independent genotyped SNPs (LD <0.1 and MAF >0.01) in UKB identified by using PLINK2 (`--indep-pairwise 50 5 0.1`) for phases 1 to 3. We also need the sames LDRs, bases, variance-covariance matrix of LDRs, and covariates as used in LDR GWAS and voxel-level summary statistics reconstruction.

- Select a threshold for significant voxel-variant associations. The default is $10^{-5}$. Then compute an empirical null distribution of cluster size across 150,000 SNPs and 50 wild bootstrap samples, resulting in 7,500,000 points. Each point means the number of associated voxels for a SNP at $10^{-5}$. Refer to the algorithm below for wild bootstrap.

- Compute the cluster size for target SNPs (those identified in voxel-level summary statistics reconstruction in discovery phase) at the same threshold $10^{-5}$.

- Exclude SNPs with a cluster size less than the quantile at level 1 - 0.05/(number of loci * effective number). Note the initial number of loci can be determined by aggregating all voxel-variant associations before screening.

- Aggregate the remaining voxel-variant associations into loci and report.

Using a threshold of $10^{-5}$ but not the original $5 \times 10^{-8}$/effect number to determine significance for voxel-variant associations in wild bootstrap can ensure the null distribution of cluster size is not all zeros. The SNPs survive at $10^{-5}$ are supposed to survive at $5 \times 10^{-8}$/effect number, because the cluster size of these SNPs at $10^{-5}$ is very big, indicating they produce robust associations. However, in some rare cases, if a SNP would be excluded at $10^{-5}$ but happens to survive at $5 \times 10^{-8}$/effect number, the SNP is very significantly associated with a small number of voxels, which indicates there might be local perturbations in images and thus such associations are not reliable.

Below we present the detailed algorithm for wild bootstrap, which can be done by `--cluster` in software HEIG developed by ourselves.

**Algorithm 1:** Wild bootstrap

**Input:** LDR residuals $\widetilde{\Xi}_r$ of $n$ subjects in the discovery phase, bases $\widetilde{\Phi}_r$, genotype data of $m$ independent SNPs $Z$, covariates $W$, number of bootstrap samples $B = 50$, significance threshold of voxel-variants associations $10^{-5}$.

**Output:** Null distribution of cluster size (number of associated voxels).

**for** $b \leftarrow 1$ **to** $B$ **do**
    **for** $i \leftarrow 1$ **to** $n$ **do**
        Randomly generate $v_i^{(b)} \sim \mathcal{N}(0, 1)$.
        Calculate bootstrap samples $\tilde{\xi}_{ij}^{(b)} = v_i^{(b)} \tilde{\xi}_{ij}$ across $j = 1, \dots, r$.
    **end**
    Denote $\widetilde{\Xi}_r^{(b)} = (\tilde{\xi}_1^{(b)}, \dots, \tilde{\xi}_r^{(b)})$ and calculate variance-covariance matrix $\frac{1}{n} \widetilde{\Xi}_r^{(b)\prime} \widetilde{\Xi}_r^{(b)}$.
    Do LDR GWAS for $\widetilde{\Xi}_r^{(b)}$ using $Z$ and $W$, resulting in LDR summary statistics.
    Reconstruct voxel-variant associations using LDR summary statistics, the variance-covariance matrix, and bases $\widetilde{\Phi}_r$, saving associations passing the threshold ($P < 10^{-5}$).
    Count the number of associated voxels for each SNP, denoting as $(c_1^{(b)}, \dots, c_m^{(b)})$.
**end**
**return** $C = (c_1^{(1)}, \dots, c_m^{(1)}, \dots, c_1^{(B)}, \dots, c_m^{(B)})$.

#### 1.3 Replication study

We collected and processed structural MRI and diffusion MRI data from phases 4 to 6 of UKB (n = 23,030). After excluding related subjects, 19,494 and 20,767 unrelated participants of European ancestry remaining for the hippocampus and white matter microstructure, respectively. We conducted FPCA, constructed LDRs, performed LDR GWAS, and reconstructed voxel-level GWAS results. For variant-level replication, we matched significant voxel-variant associations between the discovery and replication phases and evaluated correlation and direction of effect size. For locus-level replication, we extracted all SNPs in significant loci from the discovery phase and evaluated them in the replication study. Any SNPs in a locus being significant in the replication study indicates the locus is replicated by leveraging high LD among SNPs in a locus. The significant variants in replication must be in 0.25 cM from the index variant in discovery phase, according to our definition of loci. This strategy is consistent with identifying overlapping loci with previous studies, where we considered that the index variants of two loci should be within 0.25 cM.

To further justify our strategy, we compared it with similar approaches used in the literature. Below are examples of widely accepted methodologies in GWASs:

- Smith et al. [2] conducted GWAS for 3,144 brain imaging traits using 22,138 subjects in discovery and 11,086 subjects in replication. The authors defined a genetic cluster as replicating if at least one of the phenotype/variant pairs had nominal significance in their replication cohort ($P < 0.05$).

- Huerta-Chagoya et al. [3] conducted GWAS meta-analysis on type 2 diabetes (T2D) focusing on rare variants and identified 34 new variants. In replication, the authors used three independent datasets including in total 73,088 cases and 79,827 controls. By doing individual analysis first and subsequent meta-analysis, the authors considered replication when the variant showed evidence of association at $P < 0.05/34$ and consistent direction of effect with the discovery meta-analysis.

- Loya et al. [4] recently published a new efficient and scalable GWAS approach. In variant-level replication, the authors extracted significant variants from the discovery phase in UKB ($P < 5 \times 10^{-9}$), and evaluated effect size with significant variants in Biobank Japan under multiple replication thresholds ($P < 5 \times 10^{-2}$, $P < 5 \times 10^{-4}$, $P < 5 \times 10^{-6}$). In locus-level replication, they defined a credible set for a locus to contain a lead associated variant together with additional proxy variants found within a 50-kb window from the lead variant and with association significance ($P < 100P_{lead}$). They considered replication if any variant in the credible set was also found to be associated at $P < 5 \times 10^{-2}$ with the same direction of effect in the replication cohort.

We found for variant-level replication, most studies evaluated effect size correlation and direction, as we did here. For locus-level replication, Smith et al. [2] used a liberal threshold without considering numbers of traits and loci; Loya et al. [4] extracted less candidates in a credible set but used a liberal threshold for replication, even if they have tested 40 independent traits and a number of loci. Compared to these approaches, our study design is more rigorous and methodologically robust, setting a strong precedent for voxel-level GWAS studies in the future. Below are specific results:

For kernel smoothing, we observed all ROIs, except the right hippocampus, achieved the minimum generalized cross-validation (GCV) score at the same bandwidth as in the discovery phase. The effective number of independent voxels displayed a correlation coefficient of 0.994 between the discovery and replication phase. Specifically, the total effective number for the hippocampus was 10.1 in the discovery phase and 10.5 in the replication phase, while for the white matter tracts, it was 261.8 and 250.6, respectively. This strong alignment suggests a global similarity between the two independent datasets.

**Hippocampus results.** We first matched significant vertex-variant pairs in the discovery ($P < 4.94 \times 10^{-9}$) and in the replication ($P < 0.05/72$). The correlation coefficient of effect size estimates was 0.98, with 99.8% of effect size estimates showing consistent directions. Using the standard replication threshold ($P < 0.05/72$), all 72 loci were replicated (Tables S4 and S6). Applying a more stringent threshold that accounted for both the number of loci and the effective number ($P < 0.05/72/10.1$), we replicated 63 out of the 72 loci (87.5%, Table S4).

**White matter microstructure results.** We first matched significant voxel-variant pairs in the discovery ($P < 1.91 \times 10^{-10}$) and in the replication ($P < 0.05/526$). The correlation coefficient of effect size estimates was 0.96, with 99.3% of estimates showing consistent directions. Among the 526 loci identified in the discovery phase, 513 (97.5%) were replicated ($P < 0.05/526$), and 369 (70.2%) were replicated using a more stringent threshold ($P < 0.05/526/261.8$, Tables S3 and S5).

#### 1.4 FPCA vs. PCA: dimension reduction while removing white noise and recovering images

Both FPCA and PCA can be used for dimensionality reduction in images, but they differ in how the bases for image projection are computed. FPCA leverages the prior knowledge that images are essentially smooth, meaning that a voxel is highly correlated with its neighboring voxels, with this correlation decreasing as the distance increases. To exploit this property, FPCA first applies kernel smoothing to recover the underlying smooth images from the observed ones. Kernel smoothing is a non-parametric technique that combines each voxel with its nearby voxels to reduce local fluctuations. The recovery doesn't need to perfectly match the true underlying images because the goal is to estimate a set of smooth functional bases from the smoothed images. Other smoothing methods, such as smoothing splines or regression splines, could also be used, but we have not yet compared our approach with these alternatives.

To demonstrate that FPCA outperforms PCA in recovering imaging data and removing white noise, we simulated a dataset consisting of 100 images, each with 100 voxels. In this simulation, each image had a noise percentage of 0.1, meaning that 10% of the image variance was due to white noise. Fig. S22A shows a comparison between randomly selected underlying data, the raw data (which is the underlying data plus white noise), and the smoothed data obtained using kernel smoothing. After smoothing the raw data, we applied singular value decomposition (SVD) to compute a set of bases from the smoothed images. For PCA, we directly applied SVD to the raw data. We then used the bases corresponding to the top 20 singular values to reconstruct the images (Fig. S22B). The results clearly show that the image reconstructed by FPCA aligns much more closely with the underlying image, whereas the image recovered by PCA remains noisy and exhibits fluctuations.

Additionally, as shown in Fig. S1, FPCA effectively reconstructs images from real data. For the left hippocampus, the correlation between the raw images and those reconstructed with 90% of the variance retained is 0.97, while for the anterior corona radiata, the corresponding correlation is 0.95.

Why is FPCA able to remove white noise and recover the underlying data? The reason lies in the fact that FPCA projects raw images onto a set of *smooth* functional bases. Additionally, the leading bases, which capture global information, are inherently smoother than the subsequent bases, which capture more localized details. Fig. S22C compares the first basis function of FPCA with that of PCA. The FPCA basis is very smooth and free from discrete white noise, whereas the PCA basis still contains white noise. Fig. S22D illustrates the 20th basis function for both FPCA and PCA. Both show fluctuations similar to white noise, reflecting their role in capturing local signals.

As a result, FPCA can effectively decompose raw images into their underlying smoothed images and separate out the white noise, whereas PCA cannot achieve this. By using more bases in FPCA (i.e., more LDRs) to reconstruct images, more local signals are preserved. Conversely, using fewer LDRs imposes stricter regularization on the images, leading to reduced variance at each voxel. This reduction in variance can be beneficial in statistical analyses, as it may help lower the standard error of estimators.

In imaging genetics, we consider the genetic effect of a SNP $k$ across voxels as a smooth function $\beta_k(\cdot)$. The degree to which $\beta_k(\cdot)$ is accurately recovered in RVGA depends on the inherent properties of $\beta_k(\cdot)$, which are typically unknown. Consequently, some significant associations identified using the raw data may not be reproduced when using the recovered images. This is discussed in the "Genome-wide association analysis with varying proportion of variance" section below. Nevertheless, the strength of RVGA lies in its ability to provide an efficient and flexible approach for voxel-level analysis.

#### 1.5 FPCA vs. PCA: performance in real data analysis

We performed PCA for each ROI, bypassing the smoothing step in the FPCA process (`--skip-smoothing`). We selected the same number of LDRs as in FPCA and subsequently conducted GWAS on these LDRs. Finally, we reconstructed voxel-level significant associations, applying thresholds of $P < 4.94 \times 10^{-9}$ for the hippocampus and $P < 1.91 \times 10^{-10}$ for the white matter tracts.

We first counted the number of associations for each voxel and compared the total numbers. The correlation between the number of associations identified by FPCA and PCA was 0.9995 across ROIs. On average, FPCA identified 273,701 associations, compared to 269,489 associations identified by PCA (Table S15). This result is consistent with our simulation findings, which demonstrate that FPCA effectively reduces white noise for each voxel, thereby enhancing statistical power.

We further aggregated variant-voxel associations within each ROI using the Peaks algorithm and removed loci containing a single variant-voxel association. For white matter tracts, the correlation between the number of loci identified by FPCA and PCA was 0.9998. In total, FPCA identified 715 loci, while PCA identified 719 loci, with 687 loci (95.8%) overlapping between the two methods (Table S15).

For the hippocampus, FPCA identified 42 loci in the left hemisphere and 43 loci in the right hemisphere. In contrast, PCA identified 84 and 69 loci for the left and right hemispheres, respectively, with 41 and 32 loci overlapping with those identified by FPCA (Table S15). We compared the image reconstruction quality and observed that both FPCA and PCA achieved a correlation of 0.97 between raw and reconstructed images. We then hypothesized that such a high correlation has introduced noise, prominently in PCA, as the bases of PCA potentially capture white noise.

We observed that the top 12 bases in FPCA and PCA were highly correlated ($> 0.98$), but the correlation began to decline beyond the 15th base ($< 0.95$). Based on this observation, we reduced the number of LDRs from 25 to 10, preserving 80% of the image variance and reducing the correlation between raw and reconstructed images to 0.92. Using this updated approach, FPCA identified 40 loci and PCA identified 37 loci for the left hippocampus, with 37 loci overlapping. For the right hippocampus, FPCA identified 36 loci and PCA identified 39 loci, with 28 loci overlapping. We found that the number of loci identified by FPCA was robust across varying levels of preserved image variance. This robustness is due to FPCA's ability to capture the majority of underlying imaging signals in the top LDRs. In contrast, PCA does not differentiate between imaging signals and white noise, making it more sensitive to the level of preserved image variance.

In summary, our findings demonstrate that FPCA is more robust to varying image variance levels, highlighting the importance of noise reduction in imaging genetic analyses. Additionally, we recommend against preserving excessive image variance or achieving overly high correlation coefficients between raw and reconstructed images. As a general guideline, preserving 80%-90% of image variance and achieving a correlation coefficient 0.85-0.95 is optimal.

#### 1.6 Related representation learning methods

Extracting LDRs from biomedical images falls under the category of representation learning [5], which aims to effectively encode high-dimensional, sparse, graph-structured data into low-dimensional, dense vectors. We briefly compare RVGA with the approaches proposed by Wen et al. [6] and Patel et al. [7], among others.

Wen's approach utilizes stochastic orthogonally projective non-negative matrix factorization (sopNMF) to generate LDRs, which is an extension of PCA. Compared to RVGA, it directly processes raw images without denoising. Additionally, the bases $\Phi \in \mathbb{R}^{N \times r}$ (we use the same notation as RVGA) are subject to the constraints $\Phi \geqslant 0$ and $\Phi'\Phi = I$, where $I$ is the identity matrix. Instead of adaptively selecting LDRs, Wen et al. constructed multi-scale LDRs with $r = 32, 64, 128, 256, 512, 1024$, resulting in a total of 2,003 representations (13 of which were removed during quality control) at different granularities. They mapped each voxel to a single LDR by taking the index of the maximum value in the corresponding base: $\arg\max_k \Phi_{jk}$. Overall, Wen's approach emphasizes feature learning rather than dimensionality reduction and voxel-level analysis.

Patel's approach employs a convolutional neural network (CNN)-based autoencoder to learn LDRs. The model follows a U-net structure, where the vectors in the bottleneck layer ($r = 128$) are extracted as representations. The authors proposed a perturbation-based method to link LDRs to regions in the image. Compared to RVGA, the CNN-based framework can also aggregate neighborhood information for each voxel, but it does not utilize the coordinates of voxels that reflect distance. It may be more computationally efficient than RVGA, as it represents the entire brain image with only 128 LDRs while maintaining relatively good reconstruction performance. However, similar to Wen's approach, Patel's method also emphasizes feature learning rather than voxel-level analysis.

We observed that both Wen's approach and Patel's approach split the dataset into training and test sets to avoid data leakage, but this is unnecessary for RVGA, because RVGA is essentially an unsupervised learning framework. Unlike traditional machine learning tasks where overfitting is a significant concern, RVGA projects raw images onto a subspace spanned by a set of smooth functional bases, aiming for dimensionality reduction and high reconstruction quality. By using the proportion of variance metric and image reconstruction correlation, RVGA can autonomously balance bias and computational burden without the need for external data. Applying the same subjects to generate bases and perform projections does not adversely affect any downstream tasks. In this sense, RVGA more effectively utilizes all available subjects.

#### 1.7 "Overestimation" of RVGA heritability estimator

We evaluated the RVGA heritability estimator using 100 randomly selected voxels (vertices) from the superior fronto-occipital fasciculus (the left hippocampus), considering both genotyped variants and HapMap3 variants. We estimated heritability individually for each voxel or vertex using RVGA and

SumHer. We downloaded pre-computed taggings of white subjects, produced using UKB genotyped variants and HapMap3 variants, respectively, corresponding to the "BLD-LDAK" model.

The results using genotyped variants are shown in Fig. S23, where A, B, and C correspond to the left hippocampus, and D, E, and F correspond to the superior fronto-occipital fasciculus. The estimates are highly consistent across the raw data despite the distinct methodologies (Fig. S23A, B, D, and E). The two regularization levels ({80%, 75%} and {85%, 75%}) on the LD matrix produced slightly different results. We ultimately chose {80%, 75%} as it is more accurate for estimates greater than 0.20, the median voxelwise heritability. Using this regularization, we observed that the RVGA estimates on the raw data are consistently smaller than those obtained using FPCA for a certain proportion of variance (Fig. S23C and F).

We also evaluated the performance using HapMap3 variants (Fig. S24). RVGA estimates are concordant with SumHer estimates, showing high correlation coefficients (Fig. S24A, B, D, and E). We selected {98%, 95%} as the optimal regularization for better alignment. Once again, we observed that RVGA estimates on the raw data are smaller than those obtained using FPCA for a certain proportion of variance (Fig. S24C and F). The optimal regularization also matches the best one in simulation studies.

The above results align with our definition of voxel heritability, where the denominator is the underlying image variance $\mathrm{Var}\{X(\cdot)\}$ rather than the raw image variance $\mathrm{Var}\{Y(\cdot)\}$. The "overestimation" of RVGA based on FPCA observed in the sensitivity analyses below occurs because FPCA removes white noise. Therefore, RVGA is not only efficient but also effectively aggregates imaging signals. There is no problem of "overestimation" for RVGA heritability estimator.

#### 1.8 Subtlety of FPCA

##### 1.8.1 Computing only the top $k$ eigenvalues using all subjects

In this section, we explore the scenario where we compute only the top $k$ eigenvalues using all available subjects greater than image resolution. $k$ is smaller than the image resolution, as computing all eigenvalues becomes computationally intractable for large imaging datasets, such as those with 100,000 subjects and 50,000 voxels. We will address the following questions:

- Will the distribution of eigenvalues change? This could impact the effective number of independent voxels and the number of LDRs required to preserve the proportion of variance.
- How to get accurate effective number of we only have the top $k$ eigenvalues?
- Will this affect the accuracy of the top bases?
- How can we ensure that the maximum top $k$ LDRs capture sufficient image signals?

To address these important questions, we conducted a sensitivity analysis using the smoothed retrolenticular part of the internal capsule, which contains 1,496 voxels. We set $k = 100$, 500, and 1000, but still used all subjects (n = 33,324) to compute the top $k$ PCs. We used the `IncrementalPCA` module in scipy where data can be loaded into memory in batch of size $k$. We considered the results from computing all 1,496 PCs as the ground truth.

**The distribution of eigenvalues.** Fig. S25A shows the identical top 80 eigenvalues across different $k$, which means as long as using the same subjects, computing the top $k$ eigenvalues is equivalent to computing all available eigenvalues and selecting the top $k$.

Fig. S25B shows that the fewer PCs we compute, the fewer LDRs are required to capture a certain proportion of variance. This is because computing only the top $k$ PCs already truncates the imaging data, enhancing the influence of the leading PCs. Therefore, if we naively construct 41 LDRs to capture 90% of the variance (with the ground truth being 76) based on the distribution obtained from computing the top 100 PCs, the downstream analyses will be biased. The effective number of independent voxels was slightly downward biased, with values of 9.33, 10.80, 10.93, and 10.94 for $k = 100$, 500, 1000, and 1496, respectively. That means we will get false positives if only using the top eigenvalues to estimate the effective number.

**Predicting the effective number based on top eigenvalues.** Since image eigenvalues usually decay rapidly, we don't need to get all accurate eigenvalue estimates for an accurate effective number. We propose to use B-spline to predict all uncomputed eigenvalues based on the top eigenvalues. Fig. S26 shows predictions for six randomly selected ROIs. For each ROI, we computed the top 20% of eigenvalues and trained a B-spline (degree = 1) using the eigenvalue index as the predictor and log eigenvalues as the outcome. We then used the model to predict uncomputed log eigenvalues. The predicted effective numbers based on the predicted eigenvalues were very close to the true effective numbers.

As a result, as long as the sample size is greater than image resolution and 20% of image eigenvalues are computed, RVGA can accurately predict the effective number, which significantly reduces computational time and memory by at least 25 times for large imaging datasets.

**The accuracy of the top bases.** Comparing the top $k$ bases obtained from computing only the top $k$ PCs to those from computing all PCs (i.e., the ground truth), we observed that the leading bases were almost identical to the ground truth (Fig. S25C). However, the bases toward the tail became increasingly unstable, showing low correlation coefficients with the ground truth. Specifically, for $k = 100$, the alignment began to decrease at the 81st PC (Fig. S25C). Although the top 81 LDRs were able to capture more than 90% of the variance according to the eigenvalue distribution of the ground truth, caution is still warranted if $k$ is significantly less than the number of voxels.

**Ensuring that the maximum top $k$ LDRs capture sufficient image signals.** To ensure that the LDRs constructed by the top bases effectively capture sufficient image signals, RVGA evaluates the correlation between the raw and the reconstructed images during the LDR construction process. In our real data analysis, the correlation coefficient was 0.97 for the hippocampus with 90% of variance captured, and 0.88 for WM tracts with 80% of variance captured. It is reasonable that the correlation is not perfect because white noise is removed in the reconstructed images. A correlation around 0.9 (0.85-0.95) can be used as an empirical threshold to ensure the quality of reconstruction.

##### 1.8.2 Smaller sample size than image resolution

In the main article, since our sample size (n = 33,324) was greater than image resolution (max = 15,000), we can estimate the full spectrum of image eigenvalues. Here, we additionally conducted analysis for cortical surface curvature for which the sample size (n = 15,752) was much smaller than the resolution (N = 59,412). The maximum number of eigenvalues we can compute is $\min\{n, N\} = 15,752$. We are interested in the distribution of these eigenvalues compared to the top $15,752$ eigenvalues computed from using a dataset with larger sample size than the image resolution.

we conducted a sensitivity analysis using the smoothed retrolenticular part of the internal capsule, which contains 1,496 voxels. We set $n$ = 30, 100, and 300, mimicking the scenarios where we have only 30, 100, and 300 subjects for images with 1,496 voxels. We observed the distributions of eigenvalues were different from that computed using all 33,324 subjects (Fig. S25D). The effective numbers were downward biased based on the computed $n$ eigenvalues: 8.56 (n = 30), 9.54 (n = 100), 10.34 (n = 300), 10.94 (n = 33,324). Unfortunately, we cannot predict the true effective number based on eigenvalues from a limited sample size. That essentially means we do not have enough information to distinguish voxels in a higher-dimensional space. However, we can separate the whole image to small pieces and compute effective numbers for each piece and finally aggregate all the local effective numbers. This approximation will slightly overestimate the effective number, producing conservative association results.

#### 1.9 LD matrix estimation

To efficiently store the constructed LD matrix and avoid performing eigen-decomposition for each analysis, we store the weighted eigenvectors for each block in a binary format. Specifically, let $R_k$ denote the $k$-th LD block, which undergoes eigen-decomposition as $R_k = Q_k \Lambda_k Q_k'$. By extracting a certain proportion of eigenvalues, we obtain the dimension-reduced matrices $Q_k^*$ and $\Lambda_k^*$. We store $Q_k^* \sqrt{\Lambda_k^*}$ for the LD matrix and $Q_k^* (\sqrt{\Lambda_k^*})^{-1}$ for the inverse LD matrix, where $\sqrt{\cdot}$ is taken element-wise for $\Lambda_k^*$. This strategy reduces the file size by an order of magnitude. For an LD matrix with approximately 460,000 SNPs, the total file size is 0.36 GB, with 80% of the variance in each LD block preserved.

#### 1.10 LD score estimation

RVGA incorporates the intercept of cross-trait LDSC to adjust for bias due to sample overlap. LDSC is internally implemented in RVGA if the user enables the sample overlap flag (`--overlap`). Since the unified heritability and genetic correlation estimator relies on a block-diagonal LD matrix, it is natural to derive LD scores from these mutually independent LD blocks. Specifically, let $l_k(u) = 1 + \sum_{m \in \mathcal{S}} r_{km}^2$, where $l_k$ is the LD score for SNP $k$, $\mathcal{S}$ is a set of SNPs in the same LD block as SNP $k$, and $r_{mk}$ is the sample correlation between SNP $k$ and SNP $m$. We adjusted the raw correlation coefficient using the formula $r_{km,adj}^2 = r_{km}^2 - \frac{1-r_{km}^2}{n_1-2}$, where $n_1$ is the sample size of the external dataset. In contrast, LDSC [9] uses a fixed window (1 cM) around the index variant for the sum of $r^2$. The major difference in LD scores between these two strategies may occur at the boundaries of LD blocks.

We used genotype array data from chromosome 1 to compare the similarity between the two strategies. We removed SNPs with a minor allele frequency (MAF) less than 0.01 and/or a $p$-value from the Hardy–Weinberg equilibrium test less than $10^{-7}$, resulting in $36,578$ SNPs. We partitioned chromosome 1 into 133 mutually independent LD blocks and collected $9,200$ unrelated subjects of European ancestry to first estimate the LD blocks and then estimate LD scores within each block. For LDSC, the fixed window was set to $10,000$ Kb (`--ld-wind-kb 10000.0`). We observed an exceptionally high correlation coefficient of 0.98 between LD scores estimated using RVGA and LDSC (Fig. S27). We verified that most of the LD scores underestimated by RVGA were at the boundaries of LD blocks, and the number of these SNPs is very small (less than 0.1%), which would not affect the analysis results.

#### 1.11 The pipeline of RVGA

RVGA is an end-to-end pipeline for conducting voxel-level GWAS in large-scale imaging studies with support for multi-threading computation. RVGA belongs to a toolbox for imaging genetics entitled "Highly Efficient Imaging Genetics" (HEIG).

**Image loading.** Images can be in NIFTI, CIFTI, FreeSurfer morphometry data, or text file formats. An additional image or text file for coordinates is required. For NIFTI, this can be a mask in NIFTI format or one of NIFTI images to load; for CIFTI, it is a GIFTI file; for FreeSurfer morphometry data, it is a FreeSurfer surface mesh file; and for images in text file format, it is also a text file. Images can be stored in multiple directories, with the subject ID included in the file name following this naming convention: `<id><suffix>`. RVGA can read all images matching the provided suffix and extract the subject ID. The loaded images, along with metadata, will be stored in a single file in HDF5 format.

RVGA also supports to analyze non-imaging data, which can be loaded as a text file. Users are also required to provide coordinate information for the phenotypes. However, this coordinate file will not be utilized in the analysis if kernel smoothing is skipped by specifying the `--skip-smoothing` option in the FPCA step. This means the coordinate file can be any arbitrary file, provided its number of rows matches the number of phenotypes.

**FPCA.** Raw images are first subjected to kernel smoothing. The optimal bandwidth is selected from an adaptive candidate list. Then, the smoothed images are processed using `IncrementalPCA` to compute the top $k$ principal components (PCs), where $k$ is either specified by the user or adaptively selected by RVGA. During each step, images are loaded into memory in batches, and the smoothed images are saved to disk and reloaded into memory in batches as well. The bases, eigenvalues, and the effective number of independent voxels are output for user reference. RVGA prints a table indicating the number of LDRs required to preserve different proportions of image variance.

**LDR construction.** Raw images, bases, the number of LDRs, and covariates are input. RVGA constructs the LDRs and computes the variance-covariance matrix of covariate-effect-removed LDRs. It is important to ensure that the subjects included in the variance-covariance matrix are also included in the LDR GWAS, as otherwise, the downstream analyses may be biased. RVGA prints a table showing the mean correlation between reconstructed and raw images using varying numbers of LDRs.

**LDR GWAS.** LDR GWAS analysis can be internally conducted in software HEIG.

**LDR GWAS summary statistics processing.** RVGA removes SNPs from LDR GWAS summary statistics if they exhibit any of the following characteristics: 1) a duplicated rsID; 2) an ambiguous strand; 3) an effective sample size less than 0.67 times the 90th percentile of the sample size; 4) multiple alleles; or 5) a missing or infinite z-score. The genetic effect estimates and z-scores across all LDRs are saved in blocks of size (`n_snps`, 20) in a HDF5 file. The metadata for the SNPs are saved in a single text file. The summary statistics of non-imaging phenotypes can also be processed in this module.

**Voxel-level GWAS.** RVGA is flexible in reconstructing voxel-level summary statistics for different scenarios: 1) scanning the whole genome and all voxels, and saving only significant associations that pass a provided threshold; 2) conducting analysis for the whole genome and a subset of voxels; 3) conducting analysis for selected variants or a genome segment across all voxels. During computation, summary statistics are loaded into memory in batches.

**Cluster analysis and post-GWAS screening.** Some voxel-variant associations might be false positives caused by local segmentation errors, registration biases, or other types of image noise. RVGA effectively excludes SNPs by evaluating the cluster size, defined as the number of associated voxels at a certain threshold (e.g. $10^{-5}$). RVGA computes the null distribution of cluster size by the wild bootstrap approach. Users can filter SNPs with a cluster size less than the quantile at 1 - 0.05/(number of loci * effective number).

**LD matrix estimation.** RVGA estimates the LD matrix and its inverse from a pair of PLINK2 bfiles, using a specified regularization level.

**Heritability and (cross-trait) genetic correlation analysis.** RVGA performs heritability and genetic correlation analysis within images, as well as cross-trait genetic correlation analysis between imaging and non-imaging phenotypes. The inputs include processed LDR summary statistics, processed non-imaging phenotype summary statistics (optional), the LD matrix and its inverse, bases, and the variance-covariance matrix of LDRs. The LD blocks and summary statistics are loaded into memory in small batches.

#### 1.12 Sensitivity analysis

##### 1.12.1 Genome-wide association analysis with varying proportions of variance

In the main article, we retained 80% and 90% of the variance for the white matter (WM) microstructure and for the hippocampus shape, respectively. Our investigation into the GWAS summary statistics involved varying the proportion of variance preserved by LDRs from 80%, 85%, to 90%, and also comparing the results to those obtained using raw data. This process generated 10, 15, and 25 LDRs for the left hippocampus, and 19, 26, and 39 LDRs for the superior fronto-occipital fasciculus. We used 100 randomly selected voxels (vertices) from the superior fronto-occipital fasciculus (the left hippocampus) and 1,160,000 HapMap3 variants. The threshold of significance was set at $5 \times 10^{-8}$.

We first conducted GWAS and applied the Peaks algorithm [2] to aggregate associations for each voxel or vertex separately (Fig. S28A for the left hippocampus and Fig. S29A for the superior fronto-occipital fasciculus). The comparison was based on total loci in all voxels or vertices.

Using the raw data, we identified 111 loci in 100 vertices for the left hippocampus, and 83 of them (75%) overlapped with those identified by preserving 90% of the variance. The overlap decreased as fewer imaging signals were preserved: it was 79 for 85% of the variance and 69 for 80% of the variance. However, preserving fewer imaging signals increased the total number of significant loci, with 130, 128, and 139 loci identified for 90%, 85%, and 80% of the variance, respectively. The alignment of loci between the three RVGA setups was much greater. For instance, 110 loci were co-identified by preserving 90% and 85% of the variance.

We observed a similar pattern in the superior fronto-occipital fasciculus, where we identified 156 loci in 100 voxels, with 119 of them (76%) overlapping with those identified by preserving 80% of the variance. In contrast, increasing imaging signals did not necessarily improve alignment, as 115 and 117 loci overlapped with those identified by preserving 85% and 90% of the variance, respectively. Nevertheless, preserving fewer imaging signals increased the total number of loci, with 202, 210, and 242 loci identified for 90%, 85%, and 80% of the variance, respectively.

In the second phase, we further aggregated loci across vertices or voxels (Fig. S28B for the left hippocampus and Fig. S29B for the superior fronto-occipital fasciculus). For the left hippocampus, 19 loci were identified by preserving 90% of the variance, and 12 of them overlapped with those identified using the raw data (17 loci). Preserving different proportions of variance generated nearly identical numbers of loci (16 and 18 for 85% and 80% of the variance, respectively), and they largely overlapped with each other. For the superior fronto-occipital fasciculus, all 21 loci identified using the raw data were also discovered when preserving 80% of the variance (25 loci).

In general, preserving varying proportions of variance produces slightly different numbers of significant loci. RVGA was stable across these setups (i.e., 80%, 85%, and 90% of variance). When comparing loci in each voxel or vertex separately, RVGA boosts statistical power by removing white noise and combining information from the neighborhood. Some local signals might be lost in the smoothing process, leading RVGA to overlook several significant loci identified in the raw data. However, a large proportion of loci (12 out of 17 for the left hippocampus and 21 out of 21 for the superior fronto-occipital fasciculus) were effectively captured by RVGA.

##### 1.12.2 Genome-wide association analysis adjusting for more covariates

In the main article, we adjusted for age, sex, age$^2$, age $\times$ sex, age$^2 \times$ sex, assessment center (Data Field 54), and 40 genetic PCs in the LDR GWAS. Here, we additionally adjusted for head size (Field ID 25000), scan position X (Field ID 25756), scan position Y (Field ID 25757), scan position Z (Field ID 25758), scan table position (Field ID 25759), mean rfMRI head motion (Field ID 25741), and mean tfMRI head motion (Field ID 25742), as well as scan position X squared, scan position Z squared, mean rfMRI head motion squared, and mean tfMRI head motion squared.

We conducted LDR GWAS with the new list of covariates for the left hippocampus (90% of variance, 25 LDRs) and the superior fronto-occipital fasciculus (80% of variance, 19 LDRs). We then recovered voxel-level GWAS results using RVGA, extracted significant associations, aggregated them within each ROI, and finally removed suspicious loci with one a single voxel-variant association. For the left hippocampus, we identified 41 significant loci ($P < 4.94 \times 10^{-9}$), 39 of which overlapped with the results using the current list of covariates (42 loci). For the superior fronto-occipital fasciculus, we identified 14 loci ($P < 1.91 \times 10^{-10}$), 12 of which were overlapping with those using the current list of covariates (13 loci). In conclusion, our main results were not affected by confounding effects.

###### 1.12.3 Heritability and genetic correlation analysis with varying proportions of variance and regularization levels

Using the 100 randomly selected voxels (vertices) in the superior fronto-occipital fasciculus (the left hippocampus), we conducted a sensitivity analysis for the heritability and (cross-trait) genetic correlation estimators in RVGA, comparing them with SumHer for heritability and with LDSC for genetic correlation. Specifically, we retained 80% and 90% of imaging signals and applied different LD matrices and regularization levels. If the LD matrix contained genotyped SNPs, we used regularization levels of $\{70\%, 65\%\}$, $\{75\%, 70\%\}$, $\{80\%, 75\%\}$, $\{85\%, 80\%\}$, and $\{90\%, 85\%\}$. If the LD matrix contained imputed HapMap3 SNPs, we used $\{90\%, 85\%\}$, $\{95\%, 90\%\}$, and $\{98\%, 95\%\}$. The imputed SNPs contain more complex LD patterns, and more SNPs are in high LD with each other. Less restrictive regularization is needed to preserve more details in the LD matrix.

For genotyped SNPs, we downloaded the pre-computed tagging of the genotype version for the "BLD-LDAK" model from the LDAK website (`https://dougspeed.com/pre-computed-tagging-files/`), and we manually computed LD scores using 9,200 unrelated European subjects in UKB (LDSC command line tool, `--ld-wind-kb 10000`). For imputed SNPs, we downloaded the pre-computed tagging of the HapMap3 version for the "BLD-LDAK" model from the same website and LD scores generated using 1000 Genomes data from the LDSC website (`https://github.com/bulik/ldsc`). Both SumHer and LDSC were directly applied to summary statistics from VGWAS.

Preserving more imaging signals yielded a higher correlation coefficient for heritability estimates but did not significantly increase the standard error (Figs. S30, S31, S32, and S33). On average, preserving 80% and 90% of imaging signals achieved correlation coefficients of 0.7-0.8 and 0.8-0.9, respectively. Using either genotyped SNPs (Figs. S30 and S32) or imputed SNPs (Figs. S31 and S33) in RVGA produced results consistent with SumHer estimates. Different regularization levels on the LD matrix mainly impacted the standard error but had little effect on the correlation. However, overly restrictive levels (e.g., $\{90\%, 85\%\}$ for imputed SNPs) might cause downward bias. Finally, RVGA estimates were much more statistically efficient than SumHer, with the confidence interval (CI) width ratio being 0.4-0.7.

Preserving more signals and applying more restrictive regularization on the LD matrix had little impact on the correlation coefficient for genetic correlation, whether using genotyped SNPs (Figs. S34 and S36) or imputed SNPs (Figs. S35 and S37). The correlation was 0.8-0.9 in almost all cases. The width ratio of the confidence interval varied from 0.5 to 0.75. Including an additional 10% of imaging signals may increase the correlation coefficient by 0.03, at the cost of a higher CI width ratio by 0.04. The standard error slightly increased with less restrictive regularization, but it remained much more efficient than LDSC.

For cross-trait genetic correlation, we investigated the correlation with Alzheimer's disease (Figs. S38, S39, S42, and S43), and educational attainment (Figs. S40, S41, S44, and S45). In general, more restrictive regularization on the LD matrix effectively controlled the standard error, but preserving more imaging signals may or may not increase the correlation coefficient with LDSC. The performance highly depended on the absolute level of genetic correlation. However, RVGA and LDSC had better agreement on educational attainment (Figs. S40, S41, S44, and S45). For most cases, the correlation between RVGA estimates and LDSC estimates was at least 0.75, and the width ratio of the confidence interval varied from 0.7 to 1.2.

Employing more LDRs for heritability and genetic correlation analysis is advisable. To enhance accuracy, it is recommended to preserve more signals while simultaneously applying more restrictive regularization on the LD matrix.

#### 1.13 The theory of heritability and genetic correlation estimator

In this section, we provide theoretical results for the estimator of heritability and genetic correlation in RVGA. We consider the varying coefficient model for subject $i$ and index $v$, $i \in \{1, \ldots, n\}$, $v \in \{1, \ldots, N\}$,

$$Y_i(v) = W_{i\cdot}'\alpha(v) + Z_{i\cdot}'\beta(v) + \eta_i(v) + \epsilon_i(v). \tag{1}$$

Here, $Y_i(\cdot)$ represents an $N \times 1$ vector of the image, $W_{i\cdot}$ is a $p \times 1$ vector of covariates, including the intercept, and $\alpha(\cdot)$ is a $p \times N$ matrix of fixed coefficients. $Z_{i\cdot}$ is a $d \times 1$ vector of the genetic profile, and $\beta(\cdot)$ is a $d \times N$ matrix of fixed genetic effects. The term $\eta_i(\cdot)$ is an $N \times 1$ vector capturing imaging signals unexplained by covariates and genotypes, such as those affected by environmental factors. The measurement error (i.e. white noise) is denoted by $\epsilon_i(\cdot)$ represented as an $N \times 1$ vector.

We assume that each element of the $n \times d$ genotype matrix $Z$ has been normalized by the formula $Z_{ik} = (Z_{ik}^* - 2p_k)/\sqrt{2p_k(1-p_k)}$, where $Z_{ik}^*$ is the number of copies of the reference allele for the $i$-th subject and $k$-th SNP and $p_k$ is the frequency of the reference allele in the population. The genetic profile $Z_{i\cdot} \sim \mathcal{N}(0, R)$, where $R = \mathrm{E}(Z_{i\cdot}Z_{i\cdot}')$ is a positive definite LD matrix of $d$ SNPs. The residual term for spacial correlation $\eta_i(v)$ follows a Gaussian process $\eta_i(v) \sim \mathrm{GP}(0, \Sigma_\eta(v,v))$, and $\mathrm{Cov}(\eta_i(u), \eta_i(v)) = \Sigma_\eta(u,v)$. The measurement error $\epsilon_i(v)$ is i.i.d. across all $v$ and $i$, following a normal distribution $\epsilon_i(v) \sim \mathcal{N}(0, \sigma^2)$ and $\mathrm{Cov}(\epsilon_i(u), \epsilon_i(v)) = \sigma^2 1\{u = v\}$, where $1\{\cdot\}$ is the indicator function. Finally, we assume that all $Z_{i\cdot}'\beta(v)$, $\eta_i(v)$, and $\epsilon_i(v)$ are independent with each other.

**Proposition 1.** *Define marginal genetic effect estimates obtained in VGWAS (ignoring covariates) based on model (1) for $u, v \in \{1, \ldots, N\}$ as*

$$\widehat{\beta}(u) = \frac{Z'Y(u)}{n} \quad \textit{and} \quad \widehat{\beta}(v) = \frac{Z'Y(v)}{n}.$$

*Then for $\Omega = R^{-1}$, we have a moment estimator of genetic covariance $Q(u,v) = \beta(u)'R\beta(v)$*

$$\widehat{Q}(u,v) = \widehat{\beta}(u)'\Omega\widehat{\beta}(v) - \frac{\mathrm{tr}(R\Omega)}{n}\mathrm{Cov}(Y_i(u), Y_i(v)). \quad (2)$$

*Proof.* We derive the moment estimator by evaluating $\mathrm{E}(\widehat{\beta}'(u)\Omega\widehat{\beta}(v))$. Specifically,

$$\begin{aligned} n^2\mathrm{E}(\widehat{\beta}'(u)\Omega\widehat{\beta}(v)) =& \mathrm{E}\Big(Y(u)'Z\Omega Z'Y(v)\Big) \\ =& \mathrm{E}\Big(\beta(u)'Z'Z\Omega Z'Z\beta(v)\Big) + \mathrm{E}\Big(\beta(u)'Z'Z\Omega Z\eta(v)\Big) \\ &+ \mathrm{E}\Big(\eta(u)'Z\Omega Z'Z\beta(v)\Big) + \mathrm{E}\Big(\eta(u)'Z\Omega Z\eta(v)\Big) + \mathrm{E}\Big(\epsilon(u)'Z\Omega Z\epsilon(v)\Big). \end{aligned}$$

Using the moment results for Wishart distribution in Proposition 2, we have

$$\mathrm{E}(\widehat{\beta}(u)'\Omega\widehat{\beta}(v)) = (1 + \frac{1}{n})\beta(u)'R\beta(v) + \frac{\mathrm{tr}(R\Omega)}{n}\mathrm{Cov}(Y_i(u), Y_i(v)).$$

Ignoring $\frac{1}{n}\beta(u)'R\beta(v)$ for simplicity, we finally have $\widehat{Q}(u,v) = \widehat{\beta}(u)'\Omega\widehat{\beta}(v) - \frac{\mathrm{tr}(R\Omega)}{n}\mathrm{Cov}(Y_i(u), Y_i(v))$. □

When $u = v$, $\mathrm{Cov}(Y_i(v), Y_i(v)) = \beta(v)'R\beta(v) + \Sigma_\eta(v,v) + \sigma^2$, which introduces white noise into the estimator (2). However, white noise is not included in the definition of genetic variance in imaging genetics, as it is produced by the imaging scanner rather than the environment in which the subject lives. This fact indicates that techniques such as FPCA must be used to remove the white noise before conducting analysis. The effectiveness of separating white noise from imaging signals is primarily due to the independence of white noise across voxels. A rigorous proof is more technically involved and is beyond the scope of this article. Readers interested in this topic can refer to the literature on functional data analysis. To this end, we rewrite the estimator (2) in a version without white noise:

$$\widehat{Q}(u,v) = \widehat{\beta}(u)'\Omega\widehat{\beta}(v) - \frac{\mathrm{tr}(R\Omega)}{n}\mathrm{Cov}(X_i(u), X_i(v)),$$

where $X_i(v) = Y_i(v) - \epsilon_i(v)$.

##### 1.13.1 Variance of the heritability estimator

Recall we estimate the heritability using a plug-in estimator

$$\widehat{h}(v) = \frac{\widehat{Q}(v,v)}{\mathrm{Var}(X_i(v))},$$

where $\mathrm{Var}(X_i(v)) = \widehat{Q}(v,v) + \widehat{\Sigma}_\eta(v,v)$. Following the results in Lemma 2 in [10], we immediately have

$$\begin{aligned} \mathrm{Var}(\widehat{Q}(v,v)) &= \Big\{\frac{2d}{n^2}\Big[Q(v,v) + \Sigma_\eta(v,v)\Big]^2 + \frac{4Q(v,v)}{n}\Big[Q(v,v) + \Sigma_\eta(v,v)\Big] + \frac{4}{n}Q(v,v)^2\Big\}\Big\{1 + O(\frac{1}{n})\Big\}, \\ \mathrm{Var}(\widehat{\Sigma}_\eta(v,v)) &= \Big\{\frac{2d}{n^2}\Big[Q(v,v) + \Sigma_\eta(v,v)\Big]^2 + \frac{2}{n}\Sigma_\eta(v,v)^2 + \frac{2}{n}Q(v,v)^2\Big\}\Big\{1 + O(\frac{1}{n})\Big\}, \\ \mathrm{Cov}(\widehat{Q}(v,v), \widehat{\Sigma}_\eta(v,v)) &= \Big\{\frac{-2d}{n^2}\Big[Q(v,v) + \Sigma_\eta(v,v)\Big]^2 - \frac{4}{n}Q(v,v)^2\Big\}\Big\{1 + O(\frac{1}{n})\Big\}. \end{aligned}$$

Let $f(x,y) = \frac{x}{x+y}$, then the gradient of $f$ is given by

$$\frac{\partial f}{\partial x} = \frac{y}{(x+y)^2} \quad \text{and} \quad \frac{\partial f}{\partial y} = \frac{-x}{(x+y)^2}.$$

Using the Delta method, we have

$$\mathrm{Var}(\widehat{h}(v)) = \Big\{\frac{2d}{n^2} + \frac{2h(v)}{n} + \frac{2h(v)\Big[1-h(v)\Big]}{n}\Big\}\Big\{1 + O(\frac{1}{n})\Big\}.$$

##### 1.13.2 Variance of the genetic correlation estimator

We first compute the general form of covariance between two genetic correlation estimators

$$\begin{aligned}
&\mathrm{Cov}(\widehat{Q}(u,v), \widehat{Q}(a,b)) \\
=&\mathrm{Cov}(\widehat{\beta}(u)'\widehat{\beta}(v), \widehat{\beta}(a)'\widehat{\beta}(b)) + \frac{d^2}{n^4}\mathrm{Cov}(X(u)'X(v), X(a)'X(b)) \\
&- \frac{d}{n^2}\mathrm{Cov}(\widehat{\beta}(u)'\widehat{\beta}(v), X(a)'X(b)) - \frac{d}{n^2}\mathrm{Cov}(\widehat{\beta}(a)'\widehat{\beta}(b), X(u)'X(v)),
\end{aligned} \tag{3}$$

for $u, v, a, b \in \{1, \ldots, N\}$. Conditional on $Z$, and using the law of total covariance, we have

$$\begin{aligned}
&n^4\mathrm{Cov}(\widehat{\beta}(u)'\widehat{\beta}(v), \widehat{\beta}(a)'\widehat{\beta}(b)) \\
=&\mathrm{E}\Big(\mathrm{Cov}\Big[X(u)'ZZ'X(v), X(a)'ZZ'X(b)|Z\Big]\Big) + \mathrm{Cov}\Big(\mathrm{E}\Big[X(u)'ZZ'X(v)|Z\Big], \mathrm{E}\Big[X(a)'ZZ'X(b)|Z\Big]\Big) \\
=&\mathrm{E}\Big(\beta(u)'(Z'Z)^3\beta(a)\Big)\Sigma_\eta(v,b) + \mathrm{E}\Big(\beta(u)'(Z'Z)^3\beta(b)\Big)\Sigma_\eta(v,a) + \mathrm{E}\Big(\beta(v)'(Z'Z)^3\beta(a)\Big)\Sigma_\eta(u,b) \\
&+ \mathrm{E}\Big(\beta(v)'(Z'Z)^3\beta(b)\Big)\Sigma_\eta(u,a) + \mathrm{E}\Big(tr(Z'ZZ'Z)\Big)\Big[\Sigma_\eta(u,a)\Sigma_\eta(v,b) + \Sigma_\eta(u,b)\Sigma_\eta(v,a)\Big] \\
&+ \mathrm{E}\Big(\beta(u)'(Z'Z)^2\beta(v)\beta(a)'(Z'Z)^2\beta(b)\Big) - \mathrm{E}\Big(\beta(u)'(Z'Z)^2\beta(v)\Big)\mathrm{E}\Big(\beta(a)'(Z'Z)^2\beta(b)\Big) \\
&+ \mathrm{E}\Big(\beta(u)'(Z'Z)^2\beta(v)tr(Z'Z)\Big)\Sigma_\eta(a,b) - \mathrm{E}\Big(\beta(u)'(Z'Z)^2\beta(v)\Big)\mathrm{E}\Big(tr(Z'Z)\Big)\Sigma_\eta(a,b) \\
&+ \mathrm{E}\Big(\beta(a)'(Z'Z)^2\beta(b)tr(Z'Z)\Big)\Sigma_\eta(u,v) - \mathrm{E}\Big(\beta(a)'(Z'Z)^2\beta(b)\Big)\mathrm{E}\Big(tr(Z'Z)\Big)\Sigma_\eta(u,v) \\
&+ \mathrm{E}\Big(tr(Z'Z)^2\Big)\Sigma_\eta(u,v)\Sigma_\eta(a,b) - \mathrm{E}\Big(tr(Z'Z)\Big)^2\Sigma_\eta(u,v)\Sigma_\eta(a,b).
\end{aligned} \tag{4}$$

Similarly,

$$\begin{aligned}
&\mathrm{Cov}\Big(\widehat{\beta}(u)'\widehat{\beta}(v), X(a)'X(b)\Big) \\
=&\mathrm{E}\Big\{\mathrm{Cov}\Big[\widehat{\beta}(u)'\widehat{\beta}(v), X(a)'X(b)|Z\Big]\Big\} + \mathrm{Cov}\Big\{\mathrm{E}\Big[\widehat{\beta}(u)'\widehat{\beta}(v)|Z\Big], \mathrm{E}\Big[X(a)'X(b)|Z\Big]\Big\} \\
=&\frac{1}{n^2}\Big\{\mathrm{E}\Big(\beta(u)'(Z'Z)^2\beta(a)\Big)\Sigma_\eta(v,b) + \mathrm{E}\Big(\beta(u)'(Z'Z)^2\beta(b)\Big)\Sigma_\eta(v,a) \\
&+ \mathrm{E}\Big(\beta(v)'(Z'Z)^2\beta(a)\Big)\Sigma_\eta(u,b) + \mathrm{E}\Big(\beta(v)'(Z'Z)^2\beta(b)\Big)\Sigma_\eta(u,a) \\
&+ \mathrm{E}\Big[tr(Z'Z)\Big]\Big[\Sigma_\eta(u,a)\Sigma_\eta(v,b) + \Sigma_\eta(u,b)\Sigma_\eta(v,a)\Big]\Big\}
\end{aligned}$$

$$+\frac{1}{n^2}\Big\{\mathrm{E}\Big[\beta(u)'(Z'Z)^2\beta(v)\beta(a)'Z'Z\beta(b)\Big]-\mathrm{E}\Big[\beta(u)'(Z'Z)^2\beta(v)\Big]\mathrm{E}\Big[\beta(a)'Z'Z\beta(b)\Big]$$
$$+\mathrm{E}\Big[tr(Z'Z)\beta(a)'Z'Z\beta(b)\Big]\Sigma_\eta(u,v)-\mathrm{E}\Big[tr(Z'Z)\Big]\mathrm{E}\Big[\beta(a)'Z'Z\beta(b)\Big]\Sigma_\eta(u,v)\Big\}. \quad (5)$$

Following the Proposition S1 in [10], we have similar but more general moment results for Wishart distribution. Let $Q_k(u,v)=\beta(u)'R^k\beta(v)$ for nonnegative integers $k$, and let $W=Z'Z$.

**Proposition 2.** *For $u,v,a,b\in\{1,\dots,N\}$, we have*

$$\begin{aligned}
\mathrm{E}\Big(\beta(u)'W^3\beta(v)\Big) &= nQ_1(u,v)\mathrm{tr}(R)^2+2n(n+1)Q_2(u,v)\mathrm{tr}(R)\\
&\quad+n(n+1)Q_1(u,v)\mathrm{tr}(R^2)+(n^3+3n^2+4n)Q_3(u,v)\\
\mathrm{E}\Big(\mathrm{tr}(W^2)\Big) &= n\mathrm{tr}(R)^2+n(n+1)\mathrm{tr}(R^2)\\
\mathrm{E}\Big(\beta(u)'W^2\beta(v)\beta(a)'W^2\beta(b)\Big) &= \Big[n^4+2n^3+3n^2+2n\Big]Q_2(u,v)Q_2(a,b)\\
&\quad+\Big[2n^3+4n^2+2n\Big]Q_2(u,a)Q_2(v,b)+\Big[2n^3+4n^2+2n\Big]Q_2(u,b)Q_2(v,a)\\
&\quad+\Big[n^3+3n^2+4n\Big]Q_1(v,a)Q_3(u,b)+\Big[4n^2+4n\Big]Q_1(a,b)Q_3(u,v)\\
&\quad+\Big[n^3+3n^2+4n\Big]Q_1(v,b)Q_3(u,a)+\Big[n^3+3n^2+4n\Big]Q_1(u,a)Q_3(v,b)\\
&\quad+\Big[4n^2+4n\Big]Q_1(u,v)Q_3(a,b)+\Big[n^3+3n^2+4n\Big]Q_1(u,b)Q_3(v,a)\\
&\quad+\Big[n^3+n^2+2n\Big]Q_1(a,b)Q_2(u,v)\mathrm{tr}(R)+\Big[2n^2+2n\Big]Q_1(v,b)Q_2(u,a)\mathrm{tr}(R)\\
&\quad+\Big[2n^2+2n\Big]Q_1(v,a)Q_2(u,b)\mathrm{tr}(R)+\Big[n^3+n^2+2n\Big]Q_1(u,v)Q_2(a,b)\mathrm{tr}(R)\\
&\quad+\Big[2n^2+2n\Big]Q_1(u,a)Q_2(v,b)\mathrm{tr}(R)+\Big[2n^2+2n\Big]Q_1(u,b)Q_2(v,a)\mathrm{tr}(R)\\
&\quad+n^2Q_1(a,b)Q_1(u,v)\mathrm{tr}(R)^2+nQ_1(u,a)Q_1(v,b)\mathrm{tr}(R)^2\\
&\quad+nQ_1(u,b)Q_1(v,a)\mathrm{tr}(R)^2+2nQ_1(a,b)Q_1(u,v)\mathrm{tr}(R^2)\\
&\quad+\Big[n^2+n\Big]Q_1(u,a)Q_1(v,b)\mathrm{tr}(R^2)+\Big[n^2+n\Big]Q_1(u,b)Q_1(v,a)\mathrm{tr}(R^2)\\
\mathrm{E}\Big(\beta(u)'W^2\beta(v)\Big) &= nQ_1(u,v)\mathrm{tr}(R)+n(n+1)Q_2(u,v)\\
\mathrm{E}\Big(\beta(u)'W\beta(v)\Big) &= nQ_1(u,v)\\
\mathrm{E}\Big(\beta(u)'W^2\beta(v)\beta(a)'W\beta(b)\Big) &= n^3Q_2(u,v)Q_1(a,b)\\
&\quad+n^2\Big[Q_2(u,b)Q_1(v,a)+Q_2(u,a)Q_1(v,b)\Big]\\
&\quad+n^2\Big[Q_2(v,a)Q_1(u,b)+Q_2(v,b)Q_1(u,a)\Big]\\
&\quad+n^2\Big[Q_2(u,v)Q_1(a,b)+Q_1(u,v)Q_1(a,b)\mathrm{tr}(R)\Big]\\
&\quad+n\Big[Q_2(u,b)Q_1(v,a)+Q_2(u,a)Q_1(v,b)\\
&\quad+2Q_1(u,v)Q_2(a,b)+Q_1(u,a)Q_1(v,b)\mathrm{tr}(R)+Q_1(u,b)Q_1(v,a)\mathrm{tr}(R)\\
&\quad+Q_2(v,a)Q_1(u,b)+Q_2(v,b)Q_1(u,a)\Big]\\
\mathrm{E}\Big(\beta(u)'W\beta(v)\beta(a)'W\beta(b)\Big) &= n^2Q_1(u,v)Q_1(a,b)+nQ_1(u,a)Q_1(v,b)+nQ_1(u,b)Q_1(v,a)
\end{aligned}$$

$$\begin{aligned}
\mathrm{E}\Big(\beta(u)'W^2\beta(v)\mathrm{tr}(W)\Big) &= n(n^2+n+2)Q_2(u,v)\mathrm{tr}(R) + 4n(n+1)Q_3(u,v) \\
&\quad + n^2Q_1(u,v)\mathrm{tr}(R)^2 + 2nQ_1(u,v)\mathrm{tr}(R^2) \\
\mathrm{E}\Big(\beta(u)'W\beta(v)\mathrm{tr}(W)\Big) &= n^2Q_1(u,v)\mathrm{tr}(R) + 2nQ_2(u,v) \\
\mathrm{E}\Big(\mathrm{tr}(W)^2\Big) &= n^2\mathrm{tr}(R)^2 + 2n\mathrm{tr}(R^2) \\
\mathrm{E}\Big(\mathrm{tr}(W)\Big) &= n\mathrm{tr}(R)
\end{aligned}$$

*Proof.* Following [10], we present a detailed proof for $\mathrm{E}\Big(\beta(u)'W^2\beta(v)\beta(a)'W^2\beta(b)\Big)$ and other terms can be proved in a similar way. Recall $W = Z'Z \sim \mathrm{Wishart}(n, R)$. Let $S_k$ be the symmetric group on $k$ elements. Then each permutation $\pi \in S_k$ can be expressed as a product of disjoint cycle decompositions, $\pi = C_1, \ldots, G_{m(\pi)}$, where $C_j = (c_{1j}, \ldots, c_{k_j j})$, $k_1 + \cdots + k_{m(\pi)} = k$, and all of the $c_{ij} \in \{1, \ldots, k\}$ are distinct. Let $H_1, \ldots, H_k$ be $d \times d$ symmetric matrices and define the polynomial

$$r_\pi(R)(H_1, \ldots, H_k) = \prod_{j=1}^{m(\pi)} \mathrm{tr}\Big(\prod_{i=1}^{k_j} RH_{c_{ij}}\Big). \tag{6}$$

We have the moment result for each term in the polynomial [12, 11]

$$\mathrm{E}\Big(\mathrm{tr}(WH_1)\cdots\mathrm{tr}(WH_k)\Big) = \sum_{\pi\in S_k} 2^{k-m(\pi)} n^{m(\pi)} r_\pi(R)(H_1, \ldots, H_k). \tag{7}$$

For $u \in \{1, \ldots, N\}$, define $H_{ui} = \frac{1}{2}[\beta(u)e_i' + e_i\beta(u)']$, where $e_i \in \mathbb{R}^d$ and $\sum_{i=1}^d e_ie_i' = I$, we expand the target expectation using (6)

$$\begin{aligned}
&\mathrm{E}\Big(\beta(u)'W^2\beta(v)\beta(a)'W^2\beta(b)\Big) \\
=&\mathrm{E}\Big(\sum_{i=1}^d \beta(u)'We_ie_i'W\beta(v) \sum_{j=1}^d \beta(u)'We_je_j'W\beta(v)\Big) \\
=&\sum_{i,j} \mathrm{E}\Big(\mathrm{tr}(WH_{ui})\mathrm{tr}(WH_{vi})\mathrm{tr}(WH_{aj})\mathrm{tr}(WH_{bj})\Big) \\
=&\sum_{i,j} n^4\mathrm{tr}(RH_{ui})\mathrm{tr}(RH_{vi})\mathrm{tr}(RH_{aj})\mathrm{tr}(RH_{bj}) + \sum_{i,j} 2n^3\mathrm{tr}(RH_{ui})\mathrm{tr}(RH_{vi})\mathrm{tr}(RH_{aj}RH_{bj}) \\
&+ \sum_{i,j} 2n^3\mathrm{tr}(RH_{ui})\mathrm{tr}(RH_{bj})\mathrm{tr}(RH_{vi}RH_{aj}) + \sum_{i,j} 4n^2\mathrm{tr}(RH_{ui})\mathrm{tr}(RH_{vi}RH_{aj}RH_{bj}) \\
&+ \sum_{i,j} 4n^2\mathrm{tr}(RH_{ui})\mathrm{tr}(RH_{vi}RH_{bj}RH_{aj}) + \sum_{i,j} 2n^3\mathrm{tr}(RH_{ui})\mathrm{tr}(RH_{aj})\mathrm{tr}(RH_{vi}RH_{bj}) \\
&+ \sum_{i,j} 2n^3\mathrm{tr}(RH_{aj})\mathrm{tr}(RH_{bj})\mathrm{tr}(RH_{ui}RH_{vi}) + \sum_{i,j} 4n^2\mathrm{tr}(RH_{ui}RH_{vi})\mathrm{tr}(RH_{aj}RH_{bj}) \\
&+ \sum_{i,j} 4n^2\mathrm{tr}(RH_{ui}RH_{vi}RH_{aj})\mathrm{tr}(RH_{bj}) + \sum_{i,j} 8n\mathrm{tr}(RH_{ui}RH_{vi}RH_{aj}RH_{bj}) \\
&+ \sum_{i,j} 4n^2\mathrm{tr}(RH_{ui}RH_{vi}RH_{bj})\mathrm{tr}(RH_{aj}) + \sum_{i,j} 8n\mathrm{tr}(RH_{ui}RH_{vi}RH_{bj}RH_{aj})
\end{aligned}$$

$$
\begin{aligned}
&+\sum_{i,j} 4n^2 \mathrm{tr}(RH_{ui}RH_{aj}RH_{vi})\mathrm{tr}(RH_{bj}) + \sum_{i,j} 8n\mathrm{tr}(RH_{ui}RH_{aj}RH_{bj}RH_{vi}) \\
&+\sum_{i,j} 2n^3 \mathrm{tr}(RH_{ui}RH_{aj})\mathrm{tr}(RH_{vi})\mathrm{tr}(RH_{bj}) + \sum_{i,j} 4n^2\mathrm{tr}(RH_{ui}RH_{aj}RH_{bj})\mathrm{tr}(RH_{vi}) \\
&+\sum_{i,j} 4n^2 \mathrm{tr}(RH_{ui}RH_{aj})\mathrm{tr}(RH_{vi}RH_{bj}) + \sum_{i,j} 8n\mathrm{tr}(RH_{ui}RH_{aj}RH_{vi}RH_{bj}) \\
&+\sum_{i,j} 8n \mathrm{tr}(RH_{ui}RH_{bj}RH_{aj}RH_{vi}) + \sum_{i,j} 4n^2\mathrm{tr}(RH_{ui}RH_{bj}RH_{vi})\mathrm{tr}(RH_{aj}) \\
&+\sum_{i,j} 4n^2 \mathrm{tr}(RH_{ui}RH_{bj}RH_{aj})\mathrm{tr}(RH_{vi}) + \sum_{i,j} 2n^3\mathrm{tr}(RH_{ui}RH_{bj})\mathrm{tr}(RH_{vi})\mathrm{tr}(RH_{aj}) \\
&+\sum_{i,j} 8n \mathrm{tr}(RH_{ui}RH_{bj}RH_{vi}RH_{aj}) + \sum_{i,j} 4n^2\mathrm{tr}(RH_{ui}RH_{bj})\mathrm{tr}(RH_{vi}RH_{aj})
\end{aligned}
$$

We compute the result of each term using (7)

$$
\begin{aligned}
&\sum_{i,j} n^4 \mathrm{tr}(RH_{ui})\mathrm{tr}(RH_{vi})\mathrm{tr}(RH_{aj})\mathrm{tr}(RH_{bj}) = n^4 Q_2(u,v)Q_2(a,b) \\
&\sum_{i,j} 2n^3 \mathrm{tr}(RH_{ui})\mathrm{tr}(RH_{vi})\mathrm{tr}(RH_{aj}RH_{bj}) = n^3\Big[Q_2(u,v)Q_2(a,b) + Q_1(a,b)Q_2(u,v)\mathrm{tr}(R)\Big] \\
&\sum_{i,j} 2n^3 \mathrm{tr}(RH_{ui})\mathrm{tr}(RH_{bj})\mathrm{tr}(RH_{vi}RH_{aj}) = n^3\Big[Q_2(u,a)Q_2(v,b) + Q_1(v,a)Q_3(u,b)\Big]
\end{aligned}
$$

$$
\begin{aligned}
&\sum_{i,j} 4n^2 \mathrm{tr}(RH_{ui})\mathrm{tr}(RH_{vi}RH_{aj}RH_{bj}) \\
=&\frac{n^2}{2}\Big[Q_2(u,a)Q_2(v,b) + Q_2(u,b)Q_2(v,a) + Q_2(u,a)Q_1(v,b)\mathrm{tr}(R) \\
&+ Q_2(u,b)Q_1(v,a)\mathrm{tr}(R) + 2Q_1(a,b)Q_3(u,v) + Q_1(v,b)Q_3(u,a) + Q_1(v,a)Q_3(u,b)\Big]
\end{aligned}
$$

$$
\begin{aligned}
&\sum_{i,j} 4n^2 \mathrm{tr}(RH_{ui})\mathrm{tr}(RH_{vi}RH_{bj}RH_{aj}) \\
=&\frac{n^2}{2}\Big[Q_2(u,b)Q_2(v,a) + Q_2(u,a)Q_2(v,b) + Q_2(u,b)Q_1(v,a)\mathrm{tr}(R) \\
&+ Q_2(u,a)Q_1(v,b)\mathrm{tr}(R) + 2Q_1(a,b)Q_3(u,v) + Q_1(v,a)Q_3(u,b) + Q_1(v,b)Q_3(u,a)\Big]
\end{aligned}
$$

$$
\sum_{i,j} 2n^3 \mathrm{tr}(RH_{ui})\mathrm{tr}(RH_{aj})\mathrm{tr}(RH_{vi}RH_{bj}) = n^3\Big[Q_2(v,a)Q_2(u,b) + Q_1(v,b)Q_3(u,a)\Big]
$$

$$
\sum_{i,j} 2n^3 \mathrm{tr}(RH_{aj})\mathrm{tr}(RH_{bj})\mathrm{tr}(RH_{ui}RH_{vi}) = n^3\Big[Q_2(a,b)Q_2(u,v) + Q_2(a,b)Q_1(u,v)\mathrm{tr}(R)\Big]
$$

$$\sum_{i,j} 4n^2 \mathrm{tr}(RH_{ui}RH_{vi})\mathrm{tr}(RH_{aj}RH_{bj})$$
$$= n^2 \Big[ Q_2(a,b)Q_2(u,v) + Q_1(a,b)Q_2(u,v)\mathrm{tr}(R) + Q_2(a,b)Q_1(u,v)\mathrm{tr}(R) + Q_1(a,b)Q_1(u,v)\mathrm{tr}(R)^2 \Big]$$

$$\sum_{i,j} 4n^2 \mathrm{tr}(RH_{ui}RH_{vi}RH_{aj})\mathrm{tr}(RH_{bj})$$
$$= \frac{n^2}{2} \Big[ Q_2(u,b)Q_2(v,a) + Q_2(v,b)Q_2(u,a) + Q_1(u,a)Q_3(v,b) + Q_1(v,a)Q_3(u,b)$$
$$+ 2Q_1(u,v)Q_3(a,b) + Q_2(u,b)Q_1(v,a)\mathrm{tr}(R) + Q_2(v,b)Q_1(u,a)\mathrm{tr}(R) \Big]$$

$$\sum_{i,j} 8n \mathrm{tr}(RH_{ui}RH_{vi}RH_{aj}RH_{bj})$$
$$= \frac{n}{2} \Big[ 2Q_2(u,b)Q_2(v,a) + 2Q_1(a,b)Q_3(u,v) + 2Q_1(u,v)Q_3(a,b) + Q_1(v,a)Q_3(u,b) + Q_1(u,b)Q_3(v,a)$$
$$+ Q_1(u,v)Q_1(a,b)\mathrm{tr}(R^2) + Q_1(a,b)Q_2(u,v)\mathrm{tr}(R) + Q_1(u,v)Q_2(a,b)\mathrm{tr}(R) + Q_1(u,b)Q_1(v,a)\mathrm{tr}(R)^2$$
$$+ 2Q_1(v,a)Q_2(u,b)\mathrm{tr}(R) + 2Q_1(u,b)Q_2(v,a)\mathrm{tr}(R) \Big]$$

$$\sum_{i,j} 4n^2 \mathrm{tr}(RH_{ui}RH_{vi}RH_{bj})\mathrm{tr}(RH_{aj})$$
$$= \frac{n^2}{2} \Big[ Q_2(u,a)Q_2(v,b) + Q_2(u,b)Q_2(v,a) + Q_1(u,b)Q_3(v,a) + Q_1(v,b)Q_3(u,a)$$
$$+ 2Q_1(u,v)Q_3(a,b) + Q_1(v,b)Q_2(u,a)\mathrm{tr}(R) + Q_1(u,b)Q_2(v,a)\mathrm{tr}(R) \Big]$$

$$\sum_{i,j} 8n \mathrm{tr}(RH_{ui}RH_{vi}RH_{bj}RH_{aj})$$
$$= \frac{n}{2} \Big[ 2Q_2(u,a)Q_2(v,b) + 2Q_1(a,b)Q_3(u,v) + 2Q_1(u,v)Q_3(a,b) + Q_1(v,b)Q_3(u,a) + Q_1(u,a)Q_3(v,b)$$
$$+ Q_1(u,v)Q_1(a,b)\mathrm{tr}(R^2) + Q_1(a,b)Q_2(u,v)\mathrm{tr}(R) + Q_1(u,v)Q_2(a,b)\mathrm{tr}(R) + Q_1(u,a)Q_1(v,b)\mathrm{tr}(R)^2$$
$$+ 2Q_1(v,b)Q_2(u,a)\mathrm{tr}(R) + 2Q_1(u,a)Q_2(v,b)\mathrm{tr}(R) \Big]$$

$$\sum_{i,j} 4n^2 \mathrm{tr}(RH_{ui}RH_{aj}RH_{vi})\mathrm{tr}(RH_{bj})$$
$$= \frac{n^2}{2} \Big[ Q_2(u,a)Q_2(v,b) + Q_2(u,b)Q_2(v,a) + Q_1(v,a)Q_3(u,b) + 2Q_1(u,v)Q_3(a,b)$$
$$+ Q_1(u,a)Q_3(v,b) + Q_1(u,a)Q_2(v,b)\mathrm{tr}(R) + Q_1(v,a)Q_2(u,b)\mathrm{tr}(R) \Big]$$

$$\sum_{i,j} 8n \mathrm{tr}(RH_{ui}RH_{aj}RH_{bj}RH_{vi})$$

$$
\begin{aligned}
=&\frac{n}{2}\Big[2Q_2(u,a)Q_2(v,b)+2Q_1(u,v)Q_3(a,b)+2Q_1(a,b)Q_3(u,v)+Q_1(v,b)Q_3(u,a)+Q_1(u,a)Q_3(v,b)\\
&+Q_1(u,v)Q_1(a,b)\mathrm{tr}(R^2)+Q_1(u,a)Q_1(v,b)\mathrm{tr}(R)^2+2Q_1(v,b)Q_2(u,a)\mathrm{tr}(R)+Q_1(u,v)Q_2(a,b)\mathrm{tr}(R)\\
&+Q_1(a,b)Q_2(u,v)\mathrm{tr}(R)+2Q_1(u,a)Q_2(v,b)\mathrm{tr}(R)\Big]
\end{aligned}
$$

$$
\sum_{i,j}2n^3\mathrm{tr}(RH_{ui}RH_{aj})\mathrm{tr}(RH_{vi})\mathrm{tr}(RH_{bj})=n^2\Big[Q_2(u,b)Q_2(v,a)+Q_1(u,a)Q_3(v,b)\Big]
$$

$$
\begin{aligned}
&\sum_{i,j}4n^2\mathrm{tr}(RH_{ui}RH_{aj}RH_{bj})\mathrm{tr}(RH_{vi})\\
=&\frac{n^2}{2}\Big[Q_2(v,a)Q_2(u,b)+Q_2(v,b)Q_2(u,a)+Q_2(v,a)Q_1(u,b)\mathrm{tr}(R)\\
&+Q_2(v,b)Q_1(u,a)\mathrm{tr}(R)+2Q_1(a,b)Q_3(u,v)+Q_1(u,b)Q_3(v,a)+Q_1(u,a)Q_3(v,b)\Big]
\end{aligned}
$$

$$
\begin{aligned}
&\sum_{i,j}4n^2\mathrm{tr}(RH_{ui}RH_{aj})\mathrm{tr}(RH_{vi}RH_{bj})\\
=&n^2\Big[Q_2(u,v)Q_2(a,b)+Q_1(v,b)Q_3(u,a)+Q_1(u,a)Q_3(v,b)+Q_1(u,a)Q_1(v,b)\mathrm{tr}(R^2)\Big]
\end{aligned}
$$

$$
\begin{aligned}
&\sum_{i,j}8n\mathrm{tr}(RH_{ui}RH_{aj}RH_{vi}RH_{bj})\\
=&\frac{n}{2}\Big[2Q_2(a,b)Q_2(u,v)+3Q_1(u,b)Q_3(v,a)+3Q_1(v,b)Q_3(u,a)+3Q_1(v,a)Q_3(u,b)\\
&+3Q_1(u,a)Q_3(v,b)+Q_1(u,b)Q_1(v,a)\mathrm{tr}(R^2)+Q_1(u,a)Q_1(v,b)\mathrm{tr}(R^2)\Big]
\end{aligned}
$$

$$
\begin{aligned}
&\sum_{i,j}8n\mathrm{tr}(RH_{ui}RH_{bj}RH_{aj}RH_{vi})\\
&=\frac{n}{2}\Big[2Q_2(u,b)Q_2(v,a)+2Q_1(u,v)Q_3(a,b)+2Q_1(a,b)Q_3(u,v)+Q_1(v,a)Q_3(u,b)+Q_1(u,b)Q_3(v,a)\\
&+Q_1(u,v)Q_1(a,b)\mathrm{tr}(R^2)+Q_1(u,b)Q_1(v,a)\mathrm{tr}(R)^2+2Q_1(v,a)Q_2(u,b)\mathrm{tr}(R)+Q_1(u,v)Q_2(a,b)\mathrm{tr}(R)\\
&+Q_1(a,b)Q_2(u,v)\mathrm{tr}(R)+2Q_1(u,b)Q_2(v,a)\mathrm{tr}(R)\Big]
\end{aligned}
$$

$$
\begin{aligned}
&\sum_{i,j}4n^2\mathrm{tr}(RH_{ui}RH_{bj}RH_{vi})\mathrm{tr}(RH_{aj})\\
=&\frac{n^2}{2}\Big[Q_2(u,b)Q_2(v,a)+Q_2(u,a)Q_2(v,b)+Q_1(v,b)Q_3(u,a)+2Q_1(u,v)Q_3(a,b)\\
&+Q_1(u,b)Q_3(v,a)+Q_1(u,b)Q_2(v,a)\mathrm{tr}(R)+Q_1(v,b)Q_2(u,a)\mathrm{tr}(R)\Big]
\end{aligned}
$$

$$\sum_{i,j} 4n^2 \mathrm{tr}(RH_{ui}RH_{bj}RH_{aj})\mathrm{tr}(RH_{vi})$$
$$= \frac{n^2}{2}\Big[Q_2(u,a)Q_2(v,b) + Q_2(u,b)Q_2(v,a) + 2Q_1(a,b)Q_3(u,v) + Q_1(a,u)Q_3(v,b)$$
$$+ Q_1(u,b)Q_3(v,a) + Q_1(u,a)Q_2(v,b)\mathrm{tr}(R) + Q_1(u,b)Q_2(v,a)\mathrm{tr}(R)\Big]$$

$$\sum_{i,j} 2n^3 \mathrm{tr}(RH_{ui}RH_{bj})\mathrm{tr}(RH_{vi})\mathrm{tr}(RH_{aj}) = n^3\Big[Q_2(v,b)Q_2(u,a) + Q_1(u,b)Q_3(v,a)\Big]$$
$$\sum_{i,j} 8n \mathrm{tr}(RH_{ui}RH_{bj}RH_{vi}RH_{aj})$$
$$= \frac{n}{2}\Big[2Q_2(a,b)Q_2(u,v) + 3Q_1(u,a)Q_3(v,b) + 3Q_1(v,a)Q_3(u,b) + 3Q_1(v,b)Q_3(u,a)$$
$$+ 3Q_1(u,b)Q_3(v,a) + Q_1(u,a)Q_1(v,b)\mathrm{tr}(R^2) + Q_1(u,b)Q_1(v,a)\mathrm{tr}(R^2)\Big]$$

$$\sum_{i,j} 4n^2 \mathrm{tr}(RH_{ui}RH_{bj})\mathrm{tr}(RH_{vi}RH_{aj})$$
$$= n^2\Big[Q_2(u,v)Q_2(a,b) + Q_1(v,a)Q_3(u,b) + Q_1(u,b)Q_3(v,a) + Q_1(u,b)Q_1(v,a)\mathrm{tr}(R^2)\Big]$$

Combine the above results together, we have

$$\mathrm{E}\Big(\beta(u)'W^2\beta(v)\beta(a)'W^2\beta(b)\Big)$$
$$= \Big[n^4 + 2n^3 + 3n^2 + 2n\Big]Q_2(u,v)Q_2(a,b) + \Big[2n^3 + 4n^2 + 2n\Big]Q_2(u,a)Q_2(v,b)$$
$$+ \Big[2n^3 + 4n^2 + 2n\Big]Q_2(u,b)Q_2(v,a) + \Big[n^3 + 3n^2 + 4n\Big]Q_1(v,a)Q_3(u,b)$$
$$+ \Big[4n^2 + 4n\Big]Q_1(a,b)Q_3(u,v) + \Big[n^3 + 3n^2 + 4n\Big]Q_1(v,b)Q_3(u,a)$$
$$+ \Big[n^3 + 3n^2 + 4n\Big]Q_1(u,a)Q_3(v,b) + \Big[4n^2 + 4n\Big]Q_1(u,v)Q_3(a,b) + \Big[n^3 + 3n^2 + 4n\Big]Q_1(u,b)Q_3(v,a)$$
$$+ \Big[n^3 + n^2 + 2n\Big]Q_1(a,b)Q_2(u,v)\mathrm{tr}(R) + \Big[2n^2 + 2n\Big]Q_1(v,b)Q_2(u,a)\mathrm{tr}(R)$$
$$+ \Big[2n^2 + 2n\Big]Q_1(v,a)Q_2(u,b)\mathrm{tr}(R) + \Big[n^3 + n^2 + 2n\Big]Q_1(u,v)Q_2(a,b)\mathrm{tr}(R)$$
$$+ \Big[2n^2 + 2n\Big]Q_1(u,a)Q_2(v,b)\mathrm{tr}(R) + \Big[2n^2 + 2n\Big]Q_1(u,b)Q_2(v,a)\mathrm{tr}(R)$$
$$+ n^2Q_1(a,b)Q_1(u,v)\mathrm{tr}(R)^2 + nQ_1(u,a)Q_1(v,b)\mathrm{tr}(R)^2 + nQ_1(u,b)Q_1(v,a)\mathrm{tr}(R)^2$$
$$+ 2nQ_1(a,b)Q_1(u,v)\mathrm{tr}(R^2) + \Big[n^2 + n\Big]Q_1(u,a)Q_1(v,b)\mathrm{tr}(R^2) + \Big[n^2 + n\Big]Q_1(u,b)Q_1(v,a)\mathrm{tr}(R^2),$$

which completes the proof. □

Without loss of generalizability, let $R = I$, then $Q_k(u,v) = Q(u,v) = \beta(u)'\beta(v)$. Plug in the moments into (4) and (5) we have

$$\mathrm{Cov}(\widehat{\beta}(u)'\widehat{\beta}(v), \widehat{\beta}(a)'\widehat{\beta}(b))$$

$$
\begin{aligned}
=&\frac{1}{n^4}\Big\{\Big[d^2n+3dn(n+1)+n(n^2+3n+4)\Big]\Big[Q(u,a)\Sigma_\eta(v,b)+Q(u,b)\Sigma_\eta(v,a)\\
&+Q(v,a)\Sigma_\eta(u,b)+Q(v,b)\Sigma_\eta(u,a)\Big]+\Big[d^2n+dn(n+1)\Big]\Big[\Sigma_\eta(u,a)\Sigma_\eta(v,b)+\Sigma_\eta(u,b)\Sigma_\eta(v,a)\Big]\\
&+\Big[10n^2+10n\Big]Q(u,v)Q(a,b)+\Big[4n^3+10n^2+10n\Big]Q(u,a)Q(v,b)+\Big[4n^3+10n^2+10n\Big]Q(u,b)Q(v,a)\\
&+6dnQ(u,v)Q(a,b)+5d\Big[n^2+n\Big]Q(u,a)Q(v,b)+5d\Big[n^2+n\Big]Q(u,b)Q(v,a)\\
&+d^2nQ(u,a)Q(v,b)+d^2nQ(u,b)Q(v,a)\\
&+\Big[4dn+4n(n+1)\Big]Q(u,v)\Sigma_\eta(a,b)+\Big[4dn+4n(n+1)\Big]Q(a,b)\Sigma_\eta(u,v)+2dn\Sigma_\eta(a,b)\Sigma_\eta(u,v)\Big\}
\end{aligned}
\tag{8}
$$

$$
\begin{aligned}
&\mathrm{Cov}(\widehat{\beta}(u)'\widehat{\beta}(v),X(a)'X(b))\\
=&\frac{1}{n^2}\Big\{\Big[dn+n(n+1)\Big]\Big[Q(u,a)\Sigma_\eta(v,b)+Q(u,b)\Sigma_\eta(v,a)+Q(v,a)\Sigma_\eta(u,b)+Q(v,b)\Sigma_\eta(u,a)\Big]\\
&+dn\Big[\Sigma_\eta(u,a)\Sigma_\eta(v,b)+\Sigma_\eta(u,b)\Sigma_\eta(v,a)\Big]\\
&+2nQ(u,v)Q(a,b)+2\Big[n^2+n\Big]Q(u,b)Q(v,a)+2\Big[n^2+n\Big]Q(u,a)Q(v,b)\\
&+dnQ(u,a)Q(v,b)+dnQ(u,b)Q(v,a)+2nQ(a,b)\Sigma_\eta(u,v)\Big\}.
\end{aligned}
\tag{9}
$$

And it is straightforward to get

$$
\begin{aligned}
&\mathrm{Cov}(X(u)'X(v),X(a)'X(b))\\
=&n\Big[\Sigma_\eta(u,a)\Sigma_\eta(v,b)+\Sigma_\eta(u,a)Q(v,b)+\Sigma_\eta(v,b)Q(u,a)+Q(u,a)Q(v,b)\\
&+\Sigma_\eta(u,b)\Sigma_\eta(v,a)+\Sigma_\eta(u,b)Q(v,a)+\Sigma_\eta(v,a)Q(u,b)+Q(u,b)Q(v,a)\Big]
\end{aligned}
\tag{10}
$$

Plug (8),(9) and (10) into (3), we have

$$
\begin{aligned}
&\mathrm{Cov}(\widehat{Q}(u,v),\widehat{Q}(a,b))\\
=&\mathrm{Cov}(\widehat{\beta}(u)'\widehat{\beta}(v),\widehat{\beta}(a)'\widehat{\beta}(b))-\frac{d}{n^2}\mathrm{Cov}(\widehat{\beta}(u)'\widehat{\beta}(v),X(a)'X(b))-\frac{d}{n^2}\mathrm{Cov}(\widehat{\beta}(a)'\widehat{\beta}(b),X(u)'X(v))\\
&+\frac{d^2}{n^4}\mathrm{Cov}(X(u)'X(v),X(a)'X(b))\\
=&\frac{1}{n^4}\Big\{\Big[dn(n+1)+n(n^2+3n+4)\Big]\Big[Q(u,a)\Sigma_\eta(v,b)+Q(u,b)\Sigma_\eta(v,a)+Q(v,a)\Sigma_\eta(u,b)+Q(v,b)\Sigma_\eta(u,a)\Big]\\
&+dn(n+1)\Big[\Sigma_\eta(u,a)\Sigma_\eta(v,b)+\Sigma_\eta(u,b)\Sigma_\eta(v,a)\Big]\\
&+\Big[10n^2+10n\Big]Q(u,v)Q(a,b)+\Big[4n^3+10n^2+10n\Big]Q(u,a)Q(v,b)+\Big[4n^3+10n^2+10n\Big]Q(u,b)Q(v,a)\\
&+2dnQ(u,v)Q(a,b)+d\Big[n^2+n\Big]Q(u,a)Q(v,b)+d\Big[n^2+n\Big]Q(u,b)Q(v,a)\\
&+\Big[2dn+4n(n+1)\Big]Q(u,v)\Sigma_\eta(a,b)+\Big[2dn+4n(n+1)\Big]Q(a,b)\Sigma_\eta(u,v)+2dn\Sigma_\eta(a,b)\Sigma_\eta(u,v)\Big\}.
\end{aligned}
\tag{11}
$$

Plug different $u$, $v$, $a$, $b$ into (11), we have,

$$
\begin{aligned}
&\mathrm{Var}(\widehat{Q}(u,u))\\
=&\frac{1}{n^4}\Big\{4\Big[dn(n+2)+n(n+2)(n+3)\Big]Q(u,u)\Sigma_\eta(u,u)+2dn(n+2)\Sigma_\eta(u,u)^2\\
&+\Big[2dn(n+2)+4n(n+2)(n+3)+n(n+1)(4n+6)\Big]Q(u,u)^2\\
=&\frac{1}{n^4}\Big\{4(dn^2+n^3)Q(u,u)\Sigma_\eta(u,u)+2dn^2\Sigma_\eta(u,u)^2+2(dn^2+4n^3)Q(u,u)^2\Big\}\Big(1+O(\frac{1}{n})\Big)
\end{aligned}
$$

$$
\begin{aligned}
&\mathrm{Var}(\widehat{Q}(v,v))\\
=&\frac{1}{n^4}\Big\{4\Big[dn(n+2)+n(n+2)(n+3)\Big]Q(v,v)\Sigma_\eta(v,v)+2dn(n+2)\Sigma_\eta(v,v)^2\\
&+\Big[2dn(n+2)+4n(n+2)(n+3)+n(n+1)(4n+6)\Big]Q(v,v)^2\\
=&\frac{1}{n^4}\Big\{4(dn^2+n^3)Q(v,v)\Sigma_\eta(v,v)+2dn^2\Sigma_\eta(v,v)^2+2(dn^2+4n^3)Q(v,v)^2\Big\}\Big(1+O(\frac{1}{n})\Big)
\end{aligned}
$$

$$
\begin{aligned}
&\mathrm{Var}(\widehat{Q}(u,v))\\
=&\frac{1}{n^4}\Big\{\Big[dn(n+1)+n(n^2+3n+4)\Big]\Big[Q(u,u)\Sigma_\eta(v,v)+2Q(u,v)\Sigma_\eta(u,v)+Q(v,v)\Sigma_\eta(u,u)\Big]\\
&+dn(n+1)\Big[\Sigma_\eta(u,u)\Sigma_\eta(v,v)+\Sigma_\eta(u,v)^2\Big]\\
&+\Big[4n^3+10n^2+10n\Big]Q(u,u)Q(v,v)+\Big[4n^3+20n^2+20n\Big]Q(u,v)^2\\
&+2dnQ(u,v)^2+d\Big[n^2+n\Big]Q(u,u)Q(v,v)+d\Big[n^2+n\Big]Q(u,v)^2\\
&+\Big[2dn+4n(n+1)\Big]Q(u,v)\Sigma_\eta(u,v)+\Big[2dn+4n(n+1)\Big]Q(u,v)\Sigma_\eta(u,v)+2dn\Sigma_\eta(u,v)^2\Big\}\\
=&\frac{1}{n^4}\Big\{(dn^2+n^3)\Big[Q(u,u)\Sigma_\eta(v,v)+2Q(u,v)\Sigma_\eta(u,v)+Q(v,v)\Sigma_\eta(u,u)\Big]\\
&+dn^2\Big[\Sigma_\eta(u,u)\Sigma_\eta(v,v)+\Sigma_\eta(u,v)^2\Big]+(dn^2+4n^3)\Big[Q(u,u)Q(v,v)+Q(u,v)^2\Big]\Big\}\Big(1+O(\frac{1}{n})\Big)
\end{aligned}
$$

$$
\begin{aligned}
&\mathrm{Cov}(\widehat{Q}(u,v),\widehat{Q}(u,u))\\
=&\frac{1}{n^4}\Big\{2\Big[dn(n+2)+n(n^2+5n+6)\Big]\Big[Q(u,u)\Sigma_\eta(u,v)+Q(u,v)\Sigma_\eta(u,u)\Big]\\
&+2dn(n+2)\Sigma_\eta(u,u)\Sigma_\eta(u,v)+\Big[8n^3+30n^2+30n\Big]Q(u,u)Q(u,v)+2d\Big[n^2+2n\Big]Q(u,u)Q(u,v)\Big\}\\
=&\frac{1}{n^4}\Big\{2(dn^2+n^3)\Big[Q(u,u)\Sigma_\eta(u,v)+Q(u,v)\Sigma_\eta(u,u)\Big]+2dn^2\Sigma_\eta(u,u)\Sigma_\eta(u,v)\\
&+2(dn^2+4n^3)Q(u,u)Q(u,v)\Big\}\Big(1+O(\frac{1}{n})\Big)
\end{aligned}
$$

$$
\begin{aligned}
&\mathrm{Cov}(\widehat{Q}(u,v),\widehat{Q}(v,v))\\
=&\frac{1}{n^4}\Big\{2\Big[dn(n+2)+n(n^2+5n+6)\Big]\Big[Q(v,v)\Sigma_\eta(u,v)+Q(u,v)\Sigma_\eta(v,v)\Big]
\end{aligned}
$$

$$
\begin{aligned}
&+ 2dn(n+2)\Sigma_\eta(v,v)\Sigma_\eta(u,v) + \Big[8n^3 + 30n^2 + 30n\Big] Q(v,v)Q(u,v) + 2d\Big[n^2+2n\Big]Q(v,v)Q(u,v)\Big\}\\
=&\frac{1}{n^4}\Big\{2(dn^2+n^3)\Big[Q(v,v)\Sigma_\eta(u,v) + Q(u,v)\Sigma_\eta(v,v)\Big] + 2dn^2\Sigma_\eta(v,v)\Sigma_\eta(u,v)\\
&+ 2(dn^2+4n^3)Q(v,v)Q(u,v)\Big\}\Big(1+O(\frac{1}{n})\Big)
\end{aligned}
$$

$$
\begin{aligned}
&\mathrm{Cov}(\widehat{Q}(u,u),\widehat{Q}(v,v))\\
=&\frac{1}{n^4}\Big\{4\Big[dn(n+1) + n(n^2+3n+4)\Big]Q(u,v)\Sigma_\eta(u,v) + 2dn(n+1)\Sigma_\eta(u,v)^2\\
&+\Big[10n^2+10n\Big]Q(u,u)Q(v,v) + \Big[8n^3+20n^2+20n\Big]Q(u,v)^2 + 2dnQ(u,u)Q(v,v) + 2d\Big[n^2+n\Big]Q(u,v)^2\\
&+\Big[2dn + 4n(n+1)\Big]Q(u,u)\Sigma_\eta(v,v) + \Big[2dn+4n(n+1)\Big]Q(v,v)\Sigma_\eta(u,u) + 2dn\Sigma_\eta(v,v)\Sigma_\eta(u,u)\Big\}\\
=&\frac{1}{n^4}\Big\{4(dn^2+n^3)Q(u,v)\Sigma_\eta(u,v) + 2dn^2\Sigma_\eta(u,v)^2 + 2(dn^2+4n^3)Q(u,v)^2\Big\}\Big(1+O(\frac{1}{n})\Big).
\end{aligned}
$$

We want to compute $\mathrm{Var}(\widehat{GC}(u,v)) = \frac{\widehat{Q}(u,v)}{\sqrt{\widehat{Q}(u,u)\widehat{Q}(v,v)}}$. Let $f(x,y,z) = \frac{x}{\sqrt{yz}}$, we have the gradient

$$
\frac{\partial f}{\partial x} = \frac{1}{\sqrt{yz}},\ \frac{\partial f}{\partial y} = \frac{-x}{2\sqrt{y^3z}},\ \frac{\partial f}{\partial z} = \frac{-x}{2\sqrt{yz^3}},
$$

where $x = Q(u,v)$, $y = Q(u,u)$, $z = Q(v,v)$. Then we have

$$
\begin{aligned}
&\frac{\mathrm{Var}(\widehat{Q}(u,v))}{Q(u,u)Q(v,v)}\\
=&\frac{1}{n^4}\Big\{(dn^2+n^3)\Big[\frac{\Sigma_\eta(v,v)}{Q(v,v)} + 2\frac{Q(u,v)\Sigma_\eta(u,v)}{Q(u,u)Q(v,v)} + \frac{\Sigma_\eta(u,u)}{Q(u,u)}\Big]\\
&+ dn^2\Big[\frac{\Sigma_\eta(u,u)\Sigma_\eta(v,v)}{Q(u,u)Q(v,v)} + \frac{\Sigma_\eta(u,v)^2}{Q(u,u)Q(v,v)}\Big] + (dn^2+4n^3)\Big[1+\frac{Q(u,v)^2}{Q(u,u)Q(v,v)}\Big]\Big\}
\end{aligned}
$$

$$
\begin{aligned}
&\frac{\mathrm{Var}(\widehat{Q}(u,u))Q(u,v)^2}{4Q(u,u)^3Q(v,v)}\\
=&\frac{1}{n^4}\Big\{(dn^2+n^3)\frac{Q(u,v)^2\Sigma_\eta(u,u)}{Q(u,u)^2Q(v,v)} + \frac{dn^2}{2}\frac{\Sigma_\eta(u,u)^2Q(u,v)^2}{Q(u,u)^3Q(v,v)} + \frac{dn^2+4n^3}{2}\frac{Q(u,v)^2}{Q(u,u)Q(v,v)}\Big\}
\end{aligned}
$$

$$
\begin{aligned}
&\frac{\mathrm{Var}(\widehat{Q}(v,v))Q(u,v)^2}{4Q(v,v)^3Q(u,u)}\\
=&\frac{1}{n^4}\Big\{(dn^2+n^3)\frac{Q(u,v)^2\Sigma_\eta(v,v)}{Q(v,v)^2Q(u,u)} + \frac{dn^2}{2}\frac{\Sigma_\eta(v,v)^2Q(u,v)^2}{Q(v,v)^3Q(u,u)} + \frac{dn^2+4n^3}{2}\frac{Q(u,v)^2}{Q(u,u)Q(v,v)}\Big\}
\end{aligned}
$$

$$
-\frac{Q(u,v)\mathrm{Cov}(\widehat{Q}(u,u),\widehat{Q}(u,v))}{Q(u,u)^2Q(v,v)}
$$

$$
\begin{aligned}
=&-\frac{1}{n^4}\Big\{\Big[2(dn^2+n^3)\Big[\frac{Q(u,v)\Sigma_\eta(u,v)}{Q(u,u)Q(v,v)}+\frac{Q(u,v)^2\Sigma_\eta(u,u)}{Q(u,u)^2Q(v,v)}\Big]\\
&+2dn^2\frac{\Sigma_\eta(u,u)\Sigma_\eta(u,v)Q(u,v)}{Q(u,u)^2Q(v,v)}+2(dn^2+4n^3)\frac{Q(u,v)^2}{Q(u,u)Q(v,v)}\}
\end{aligned}
$$

$$
\begin{aligned}
&-\frac{Q(u,v)\mathrm{Cov}(\widehat{Q}(v,v),\widehat{Q}(u,v))}{Q(v,v)^2Q(u,u)}\\
=&-\frac{1}{n^4}\Big\{\Big[2(dn^2+n^3)\Big[\frac{Q(u,v)\Sigma_\eta(u,v)}{Q(u,u)Q(v,v)}+\frac{Q(u,v)^2\Sigma_\eta(v,v)}{Q(v,v)^2Q(u,u)}\Big]\\
&+2dn^2\frac{\Sigma_\eta(v,v)\Sigma_\eta(u,v)Q(u,v)}{Q(v,v)^2Q(u,u)}+2(dn^2+4n^3)\frac{Q(u,v)^2}{Q(u,u)Q(v,v)}\}
\end{aligned}
$$

$$
\begin{aligned}
&\frac{Q(u,v)^2\mathrm{Cov}(\widehat{Q}(u,u),\widehat{Q}(v,v))}{2Q(u,u)^2Q(v,v)^2}\\
=&\frac{1}{n^4}\Big\{2(dn^2+n^3)\frac{Q(u,v)^3\Sigma_\eta(u,v)}{Q(u,u)^2Q(v,v)^2}+dn^2\frac{Q(u,v)^2\Sigma_\eta(u,v)^2}{Q(u,u)^2Q(v,v)^2}+(4n^3+dn^2)\frac{Q(u,v)^4}{Q(u,u)^2Q(v,v)^2}\Big\}.
\end{aligned}
$$

If $Q(u,v)\neq 0$, then $\mathrm{Var}(\widehat{GC}(u,v))$ can be approximated by

$$
\begin{aligned}
&\mathrm{Var}(\widehat{GC}(u,v))\\
\approx&\frac{1}{Q(u,u)Q(v,v)}\mathrm{Var}(\widehat{Q}(u,v))+\frac{Q(u,v)^2}{4Q(u,u)^3Q(v,v)}\mathrm{Var}(\widehat{Q}(u,u))+\frac{Q(u,v)^2}{4Q(u,u)Q(v,v)^3}\mathrm{Var}(\widehat{Q}(v,v))\\
&-\frac{Q(u,v)}{Q(u,u)^2Q(v,v)}\mathrm{Cov}(\widehat{Q}(u,u),\widehat{Q}(u,v))-\frac{Q(u,v)}{Q(u,u)Q(v,v)^2}\mathrm{Cov}(\widehat{Q}(u,v),\widehat{Q}(v,v))\\
&+\frac{Q(u,v)^2}{2Q(u,u)^2Q(v,v)^2}\mathrm{Cov}(\widehat{Q}(u,u),\widehat{Q}(v,v))\\
=&(\frac{4}{n}+\frac{d}{n^2})\Big(GC(u,v)^2-1\Big)^2\\
&+\frac{d}{n^2}\Big[\frac{1}{h(u)}-1+\frac{1}{h(v)}-1+\Big(\frac{1}{h(u)}-1\Big)\Big(\frac{1}{h(v)}-1\Big)-2GC(u,v)^2\frac{\Sigma_\eta(u,v)}{Q(u,v)}+2GC(u,v)^4\frac{\Sigma_\eta(u,v)}{Q(u,v)}\\
&+\frac{1}{2}GC(u,v)^2\Big(\frac{1}{h(u)}-1\Big)^2+\frac{1}{2}GC(u,v)^2\Big(\frac{1}{h(v)}-1\Big)^2+GC(u,v)^2\frac{\Sigma_\eta(u,v)^2}{Q(u,v)^2}+GC(u,v)^4\frac{\Sigma_\eta(u,v)^2}{Q(u,v)^2}\\
&-GC(u,v)^2\Big(\frac{1}{h(u)}-1\Big)-2GC(u,v)^2\frac{\Sigma_\eta(u,v)}{Q(u,v)}\Big(\frac{1}{h(u)}-1\Big)-GC(u,v)^2\Big(\frac{1}{h(v)}-1\Big)\\
&-2GC(u,v)^2\frac{\Sigma_\eta(u,v)}{Q(u,v)}\Big(\frac{1}{h(v)}-1\Big)\Big]\\
&+\frac{1}{n}\Big[\frac{1}{h(u)}-1+\frac{1}{h(v)}-1-GC(u,v)^2\Big(\frac{1}{h(u)}-1\Big)-GC(u,v)^2\Big(\frac{1}{h(v)}-1\Big)-2GC(u,v)^2\frac{\Sigma_\eta(u,v)}{Q(u,v)}\\
&+2GC(u,v)^4\frac{\Sigma_\eta(u,v)}{Q(u,v)}\Big]\\
=&\Big(\frac{4}{n}+\frac{d}{n^2}\Big)\Big(1-GC(u,v)^2\Big)^2+\Big(\frac{1}{n}+\frac{d}{n^2}\Big)\Big(1-GC(u,v)^2\Big)\Big(\frac{1}{h(u)}+\frac{1}{h(v)}-2-2GC(u,v)^2\frac{\Sigma_\eta(u,v)}{Q(u,v)}\Big)\\
&+\frac{d}{2n^2}GC(u,v)^2\Big(\frac{1}{h(u)}-1-\frac{\Sigma_\eta(u,v)}{Q(u,v)}\Big)^2+\frac{d}{2n^2}GC(u,v)^2\Big(\frac{1}{h(v)}-1-\frac{\Sigma_\eta(u,v)}{Q(u,v)}\Big)^2
\end{aligned}
$$

$$+\frac{d}{n^2}\Big(GC(u,v)^4\frac{\Sigma_\eta(u,v)^2}{Q(u,v)^2}+\Big(\frac{1}{h(u)}-1\Big)\Big(\frac{1}{h(v)}-1\Big)\Big)-\frac{d}{n^2}GC(u,v)^2\frac{\Sigma_\eta(u,v)}{Q(u,v)}\Big(\frac{1}{h(u)}+\frac{1}{h(u)}-2\Big).$$

If $Q(u,v)=0$, then $\mathrm{Var}(\widehat{GC}(u,v))$ is reduced to

$$\mathrm{Var}(\widehat{GC}(u,v))\approx\frac{d}{n^2}\Big[\frac{1}{h(u)}+\frac{1}{h(v)}-1+\Big(\frac{1}{h(u)}-1\Big)\Big(\frac{1}{h(v)}-1\Big)\Big]+\frac{1}{n}\Big[\frac{1}{h(u)}+\frac{1}{h(v)}+2\Big].$$

##### 1.13.3 The equivalence between RVGA and LDSC

In a more general scenario, suppose we have two trait vectors $Y$ and $U$, measured on $n_1$ and $n_2$ unrelated subjects, respectively. Let $n_0$ denote the number of subjects that were recruited in both studies, and $0\leqslant n_0\leqslant\min\{n_1,n_2\}$. All subjects have their genotypes measured on $p$ common SNPs. We denote $X^{(1)}$ as an $n_1$ by $p$ random matrix for the first study and $X^{(2)}$ as an $n_2$ by $p$ random matrix for the second study. Suppose that all the columns in $X^{(1)}$ and $X^{(2)}$ have been normalized such that each element in both matrices has mean 0 and variance 1. The normalized genetic profile for any subject $i$ follows a multivariate normal distribution $\mathcal{N}(0,R)$, where $R$ is a $p$ by $p$ LD matrix quantifying the dependence between SNPs. Then we connect the trait and genotypes using a linear model

$$Y=X^{(1)}\beta+\epsilon$$
$$U=X^{(2)}\gamma+\delta$$

where we assume that $\beta$ and $\gamma$ are fixed effects. Without loss of generalization, we suppose that the first $n_0$ rows in $X^{(1)}$ and $X^{(2)}$ are from the overlapped subjects, denoted as $X_0$, and the remaining parts are denoted as $X_1$ and $X_2$, respectively. We split the trait vectors and the unobserved errors in an analogues way

$$X^{(1)}=(X_0',X_1')',\ Y=(Y_0',Y_1')',\ \epsilon=(\epsilon_0',\epsilon_1')'$$
$$X^{(2)}=(X_0',X_2')',\ U=(U_0',U_2'),\ \delta=(\delta_0',\delta_2')'$$

We have a GWAS estimate using univariate linear model for SNP $k$

$$\widehat{\beta}_k=(X_k^{(1)'}X_k^{(1)})^{-1}X_k^{(1)'}Y=\frac{1}{n_1}X_k^{(1)'}Y=\frac{1}{n_1}\Big(X_{0k}'Y_0+X_{1k}'Y_1\Big)$$
$$\widehat{\gamma}_k=(X_k^{(2)'}X_k^{(2)})^{-1}X_k^{(2)'}U=\frac{1}{n_2}X_k^{(2)'}U=\frac{1}{n_2}\Big(X_{0k}'U_0+X_{2k}'U_2\Big)\qquad(12)$$

where the subscript $k$ means the $k$-th column of a matrix. Then the following proposition holds

**Proposition 3.** *Assume that the LD matrix $R$ is known and the LD score of SNP $k$ is defined as $l_k=\sum_{m=1}^p r_{mk}^2$. Under the current assumption that $\beta$ and $\gamma$ are fixed effects, then*

$$\mathrm{E}(\widehat{\beta}_k\widehat{\gamma}_k)=(1+\frac{n_0}{n_1n_2})\beta'R_kR_k'\gamma+\frac{n_0}{n_1n_2}\mathrm{Cov}(Y_{0i},U_{0i}).\qquad(13)$$

*And if $n_1$ and $n_2$ are large and under the assumption of LDSC where*

$$\begin{pmatrix}\beta\\ \gamma\end{pmatrix} \sim \mathcal{N}\Big[0, \frac{1}{p}\begin{pmatrix}h_1^2 I_p & \rho_g I_p\\ \rho_g I_p & h_2^2 I_p\end{pmatrix}\Big], \ \begin{pmatrix}\epsilon_0\\ \delta_0\end{pmatrix} \sim \mathcal{N}\Big[0, \begin{pmatrix}(1-h_1^2)I_{n_0} & \rho_e I_{n_0}\\ \rho_e I_{n_0} & (1-h_2^2)I_{n_0}\end{pmatrix}\Big],$$

*then the z-scores $z_{\beta_k} = \widehat{\beta}_k/se(\widehat{\beta}_k)$, $z_{\gamma_k} = \widehat{\gamma}_k/se(\widehat{\gamma}_k)$ follow the equation*

$$\mathrm{E}(z_{\beta_k} z_{\gamma_k}) = \frac{\rho_g\sqrt{n_1 n_2}}{p} l_k + \frac{n_0(\rho_g + \rho_e)}{\sqrt{n_1 n_2}},$$

*which is exactly the cross-trait LDSC model.*

It is shown that the proposed method is a generalization of LDSC [8] where we removed the distribution assumption on the genetic effects, which means that any genetic architectures reflected by $\beta$ and $\gamma$ can be captured and it is robust to model misspecifications.

*Proof.* We plug (12) into the (13) and we have

$$\mathrm{E}(\widehat{\beta}_k\widehat{\gamma}_k) = \frac{1}{n_1 n_2}\mathrm{E}\Big(X_{0k}'Y_0X_{0k}U_0 + X_{0k}'Y_0X_{2k}'U_2 + X_{1k}'Y_1X_{0k}'U_0 + X_{1k}'Y_1X_{2k}'U_2\Big)$$

where,

$$\begin{aligned}
\mathrm{E}\Big(X_{0k}'Y_0X_{0k}U_0\Big) =& \mathrm{E}\Big(X_{0k}'X_0\beta X_{0k}'X_0\gamma + X_{0k}'X_0\beta X_{0k}'\epsilon_0 + X_{0k}'\delta_0X_{0k}'X_0\gamma + X_{0k}'\delta_0X_{0k}'\epsilon_0\Big)\\
=& \mathrm{E}\Big(X_{0k}'X_0\beta X_{0k}'X_0\gamma + X_{0k}'\delta_0X_{0k}'\epsilon_0\Big)\\
\mathrm{E}\Big(X_{0k}'Y_0X_{2k}'U_2\Big) =& \mathrm{E}\Big(X_{0k}'Y_0\Big)\mathrm{E}\Big(X_{2k}'U_2\Big)\\
\mathrm{E}\Big(X_{1k}'Y_1X_{0k}'U_0\Big) =& \mathrm{E}\Big(X_{1k}'Y_1\Big)\mathrm{E}\Big(X_{0k}'U_0\Big)\\
\mathrm{E}\Big(X_{1k}'Y_1X_{2k}'U_2\Big) =& \mathrm{E}\Big(X_{1k}'Y_1\Big)\mathrm{E}\Big(X_{2k}'U_2\Big)
\end{aligned} \tag{14}$$

It is easy to compute that

$$\begin{aligned}
\mathrm{E}\Big(X_{0k}'\delta_0X_{0k}'\epsilon_0\Big) =& n_0\mathrm{Cov}(\epsilon_{0i}, \delta_{0i}),\ \mathrm{E}\Big(X_{0k}'Y_0\Big) = n_0\sum_{m=1}^{p} r_{km}\beta_m,\ \mathrm{E}\Big(X_{2k}'U_2\Big) = (n_2 - n_0)\sum_{m=1}^{p} r_{km}\gamma_m\\
\mathrm{E}\Big(X_{1k}'Y_1\Big) =& (n_1 - n_0)\sum_{m=1}^{p} r_{km}\beta_m,\ \mathrm{E}\Big(X_{0k}'U_0\Big) = n_0\sum_{m=1}^{p} r_{km}\gamma_m
\end{aligned} \tag{15}$$

We present only the derivation of $\mathrm{E}\Big(X_{0k}'X_0\beta X_{0k}'X_0\gamma\Big)$ since other terms can be computed in a similar way. First expand the equation

$$\mathrm{E}\Big(X_{0k}'X_0\beta X_{0k}'X_0\gamma\Big) = \sum_{m=1}^{p}\mathrm{E}\Big(\beta_m X_{0m}'X_{0k}X_{0k}'X_{0m}\gamma_m\Big) + \sum_{m\neq m'}^{p}\mathrm{E}\Big(\beta_m X_{0m}'X_{0k}X_{0k}'X_{0m'}\gamma_{m'}\Big),$$

where $\mathrm{E}\Big(X_{0m}'X_{0k}X_{0k}'X_{0m}\Big)$ can be computed using the property of standard normal distribution and

Isserlis theorem

$$\mathrm{E}\Big(X_{0m}'X_{0k}X_{0k}'X_{0m}\Big) = \sum_{i=1}^{n_0}\mathrm{E}\Big(X_{0mi}^2X_{0ki}^2\Big) + \sum_{i\neq i'}^{n_0}\mathrm{E}\Big(X_{0mi}X_{0ki}X_{0mi'}X_{0ki'}\Big) = n_0(1+2r_{mk}^2) + n_0(n_0-1)r_{mk}^2$$

Similarly,

$$\mathrm{E}\Big(X_{0m}'X_{0k}X_{0k}'X_{0m'}\Big) = n_0(r_{mk}r_{m'k} + r_{mm'} + n_0r_{mk}r_{m'k})$$

Then we have

$$\begin{aligned}
&\mathrm{E}\Big(X_{0k}'X_0\beta X_{0k}'X_0\gamma\Big)\\
=&\sum_{m=1}^{p} n_0(1+n_0r_{mk}^2+r_{mk}^2)\beta_m\gamma_m + \sum_{m\neq m'}^{p}\beta_m\gamma_{m'}n_0(r_{mk}r_{m'k}+r_{mm'}+n_0r_{mk}r_{m'k})\\
=&n_0^2\sum_{m=1}^{p} r_{mk}\beta_m\sum_{m=1}^{p} r_{mk}\gamma_m + n_0\sum_{m=1}^{p} r_{mk}\beta_m\sum_{m=1}^{p} r_{mk}\gamma_m + n_0\beta'R\gamma \qquad (16)
\end{aligned}$$

Plug (15), (16) into (14), we have

$$\begin{aligned}
&\mathrm{E}(\widehat{\beta}_k\widehat{\gamma}_k)\\
=&\frac{1}{n_1n_2}\Big[n_0^2\sum_{m=1}^{p} r_{mk}\beta_m\sum_{m=1}^{p} r_{mk}\gamma_m + n_0\sum_{m=1}^{p} r_{mk}\beta_m\sum_{m=1}^{p} r_{mk}\gamma_m + n_0\mathrm{Cov}(Y_{0i},U_{0i})\\
&+ n_0(n_2-n_0)\sum_{m=1}^{p} r_{km}\beta_m\sum_{m=1}^{p} r_{km}\gamma_m + n_0(n_1-n_0)\sum_{m=1}^{p} r_{km}\beta_m\sum_{m=1}^{p} r_{km}\gamma_m\\
&+ (n_2-n_0)(n_1-n_0)\sum_{m=1}^{p} r_{km}\beta_m\sum_{m=1}^{p} r_{km}\gamma_m\Big]\\
=&\frac{1}{n_1n_2}\Big[(n_0^2+n_0+n_1n_2-n_0^2)\sum_{m=1}^{p} r_{mk}\beta_m\sum_{m=1}^{p} r_{mk}\gamma_m + n_0\mathrm{Cov}(Y_{0i},U_{0i})\Big]\\
=&(1+\frac{n_0}{n_1n_2})\sum_{m=1}^{p} r_{mk}\beta_m\sum_{m=1}^{p} r_{mk}\gamma_m + \frac{n_0}{n_1n_2}\mathrm{Cov}(Y_{0i},U_{0i})\\
=&(1+\frac{n_0}{n_1n_2})\beta'R_kR_k'\gamma + \frac{n_0}{n_1n_2}\mathrm{Cov}(Y_{0i},U_{0i}).
\end{aligned}$$

Finally, under the assumption of LDSC where $\beta$ and $\gamma$ are considered as random effects, we have

$$\mathrm{E}(\widehat{\beta}_k\widehat{\gamma}_k) = (1+\frac{n_0}{n_1n_2})R_k'R_k\frac{\rho_g}{p} + \frac{n_0}{n_1n_2}\mathrm{Cov}(Y_{0i},U_{0i}) = (1+\frac{n_0}{n_1n_2})\frac{\rho_g}{p}l_k + \frac{n_0}{n_1n_2}\mathrm{Cov}(Y_{0i},U_{0i}).$$

When the sample sizes are large, $\mathrm{E}(\widehat{\beta}_k\widehat{\gamma}_k) \approx \frac{\rho_g}{p}l_k + \frac{n_0}{n_1n_2}\mathrm{Cov}(Y_{0i},U_{0i})$. Since

$$\mathrm{Var}(\widehat{\beta}_k) = \frac{1}{n_1}\mathrm{Var}(Y_{0i}) = \frac{1}{n_1},\ \mathrm{Var}(\widehat{\gamma}_k) = \frac{1}{n_2}\mathrm{Var}(U_{0i}) = \frac{1}{n_2},$$

then

$$\mathrm{E}(z_{\beta_k} z_{\gamma_k}) = \frac{\rho_g \sqrt{n_1 n_2}}{p} l_k + \frac{n_0(\rho_g + \rho_e)}{\sqrt{n_1 n_2}}.$$

□

##### 1.13.4 The relationship between RVGA and the generalized random effects model

One of closely related heritability estimators for the whole genome is the generalized random effects (GRE) model [13]. In our notation, the GRE estimator is

$$h_{GRE} = \frac{n\widehat{\beta}'\widehat{\Omega}\widehat{\beta}}{n-d} - \frac{d}{n-d},$$

where $n$ is sample size, $d$ is the number of SNPs included in the study, $\widehat{\beta}$ is a vector of marginal genetic effects from GWAS, $\widehat{\Omega} = (\frac{1}{n}Z'Z)^{-1}$ is the *in-sample* LD inverse matrix. Compared to RVGA, GRE also uses a moment estimator and does not have distribution and genetic architecture assumptions on genetic effects $\beta$. GRE implicitly assumes $d < n$. To estimate the whole-genome heritability, GRE partitions the genome into chromosomes and separately estimates heritability

$$h_{GRE} = \sum_{k=1}^{22} \frac{n\widehat{\beta}_k'\widehat{\Omega}_k\widehat{\beta}_k}{n-d_k} - \frac{d_k}{n-d_k},\ d_k < n$$

where $\widehat{\beta}_k$, $\widehat{\Omega}_k$, and $d_k$ are defined accordingly for chromosome $k$.

The major difference between GRE and RVGA is the LD inverse matrix, for which GRE uses the in-sample one, but RVGA uses an estimate from a reference panel. Using in-sample LD matrix essentially requires the individual genetic data which does not align with our goal of a completely summary statistic-based method. More importantly, in-sample LD-based methods cannot be extended to genetic correlation estimation.

Using an in-sample LD matrix yields a slightly different estimator. Specifically,

$$\begin{aligned}
\mathrm{E}[\widehat{\beta}'\widehat{\Omega}\widehat{\beta}|Z] &= \mathrm{E}[\mathrm{tr}\{\widehat{\beta}'\widehat{\Omega}\widehat{\beta}\}|Z] \\
&= \mathrm{tr}\{\widehat{\Omega}\mathrm{E}[\widehat{\beta}\widehat{\beta}'|Z]\} \\
&= \mathrm{tr}\{\widehat{\Omega}\mathrm{Cov}[\widehat{\beta}|Z]\} + \mathrm{tr}\{\widehat{\Omega}\mathrm{E}[\widehat{\beta}|Z]\mathrm{E}[\widehat{\beta}|Z]'\} \\
&= \mathrm{tr}\{\frac{\sigma^2}{n}\widehat{\Omega}\widehat{R}\} + \beta'\widehat{R}\beta \\
&= \frac{d}{n}\sigma^2 + \beta'\widehat{R}\beta,
\end{aligned}$$

where $\sigma^2$ is the residual variance. Compared to derivation of RVGA in Proposition 1, GRE evaluates the conditional expectation of $\widehat{\beta}'\widehat{\Omega}\widehat{\beta}$ while RVGA evaluates the marginal expectation, which means RVGA considers the variation in the genotype matrix $Z$. To do that, we have an additional assumption for the (standard) genotype matrix $Z$ such that each row $Z_{i\cdot}$ follows the normal distribution $\mathcal{N}(0, R)$. Moreover, $\mathrm{tr}\{\widehat{\Omega}\widehat{R}\} = d$ requires $d < n$ but RVGA doesn't have such a requirement since we directly estimate $\mathrm{tr}\{\widehat{\Omega}\widehat{R}\}$ from reference panels. As the cost, the estimate variance of RVGA is slightly greater than that of GRE. Specifically, let $\rho = n/d$, we have

$$\mathrm{Var}(\widehat{h}_{GRE}) = \Big(\frac{\rho}{\rho-1}\Big)^2\Big(\frac{2(1-h)}{\rho} + 4h\Big)\Big(\frac{1-h}{n}\Big)$$
$$\mathrm{Var}(\widehat{h}) = \frac{2}{\rho n} + \frac{2h}{n} + \frac{2h(1-h)}{n}$$

when $\rho \gg 1$, $\frac{\rho}{\rho-1} \approx 1$, we have

$$\begin{aligned}&\mathrm{Var}(\widehat{h}_{GRE}) - \mathrm{Var}(\widehat{h})\\ =&\Big(\frac{2(1-h)}{\rho} + 4h\Big)\Big(\frac{1-h}{n}\Big) - \frac{2}{\rho n} - \frac{2h}{n} - \frac{2h(1-h)}{n}\\ =&\frac{2h}{n}\Big(\frac{-2}{\rho} + \frac{h}{\rho} - h\Big) < 0.\end{aligned}$$

However, since $n$ is large, the difference is almost negligible.

#### 2 Supplementary Figures

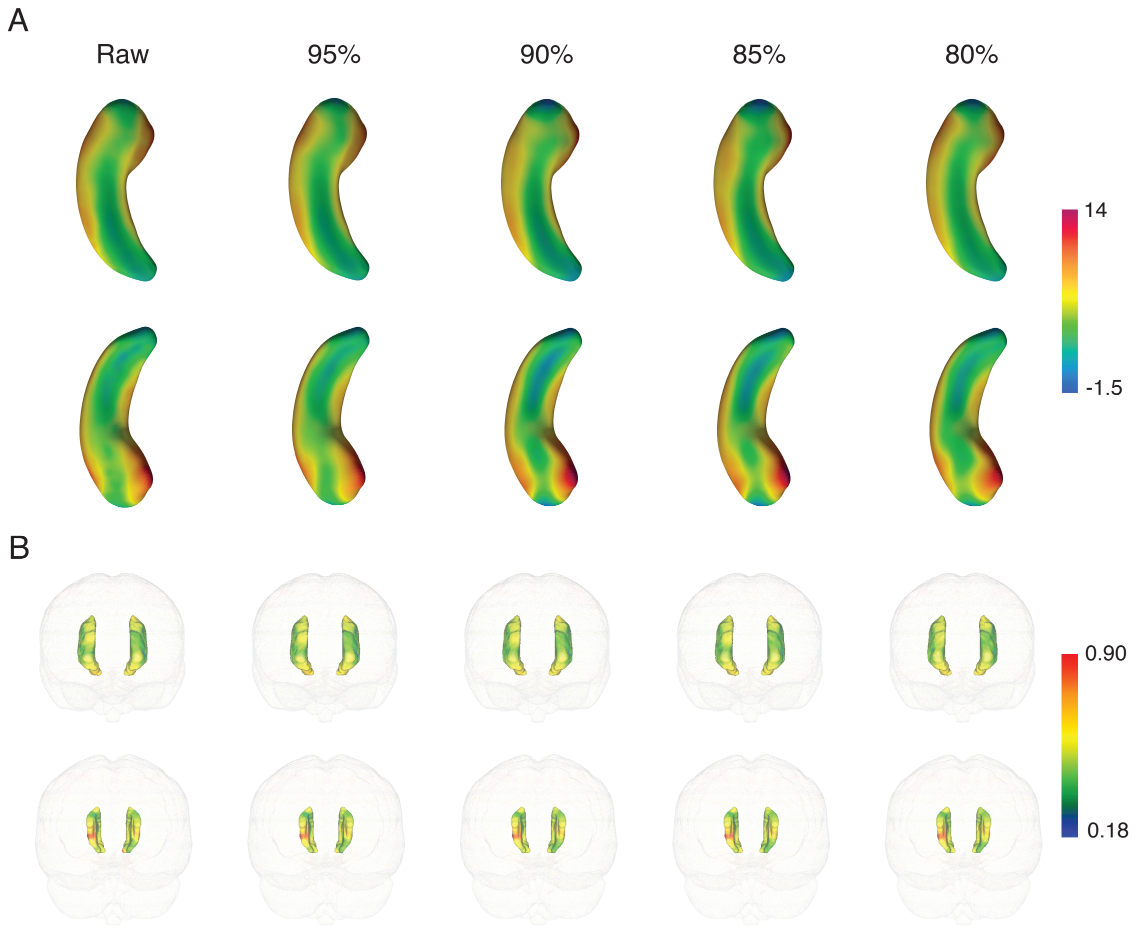

Figure S1: Image reconstruction by preserving different proportions of variance. A, The left hippocampus. The correlation coefficient between the reconstructed images and the raw images is 0.99 (95%), 0.97 (90%), 0.95 (85%), and 0.93 (80%). B, The anterior corona radiata. The correlation coefficient between the reconstructed images and the raw images is 0.98 (95%), 0.95 (90%), 0.93 (85%), and 0.91 (80%). The color bar represents radial distance for the left hippocampus and FA for the anterior corona radiata.

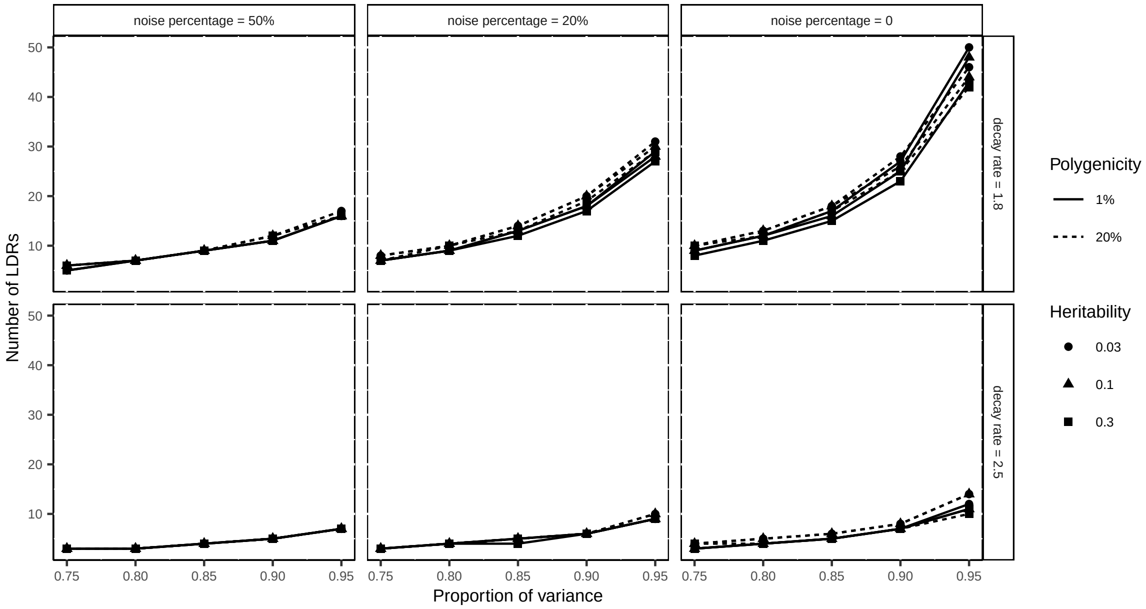

Figure S2: The number of LDRs for preserving a proportion of variance in simulation studies. Noise percentage: the proportion of image variance contributed by white noise. Decay rate: the polynomial decay rate of image eigenvalues ($\lambda$); the larger the rate, the more rapidly the eigenvalues decay. Scenarios including low/high polygenicity and low/medium/high heritability.

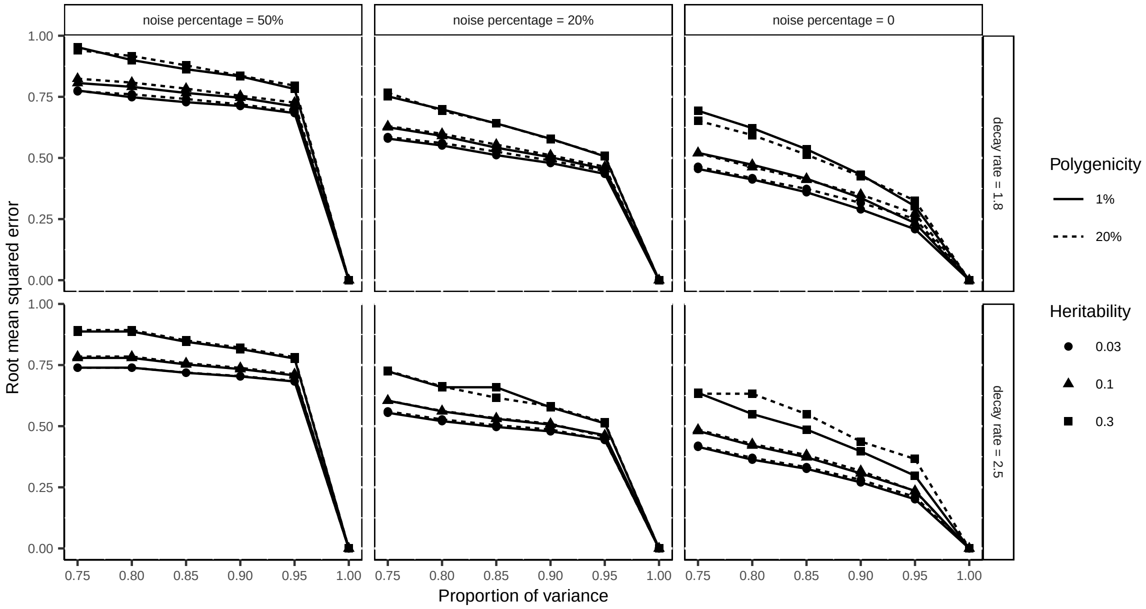

Figure S3: Root mean squared error of z-scores of GWAS summary statistics in simulation studies (Gaussian noise). Noise percentage: the proportion of image variance contributed by white noise. Decay rate: the polynomial decay rate of image eigenvalues ($\lambda$); the larger the rate, the more rapidly the eigenvalues decay. Scenarios including low/high polygenicity and low/medium/high heritability.

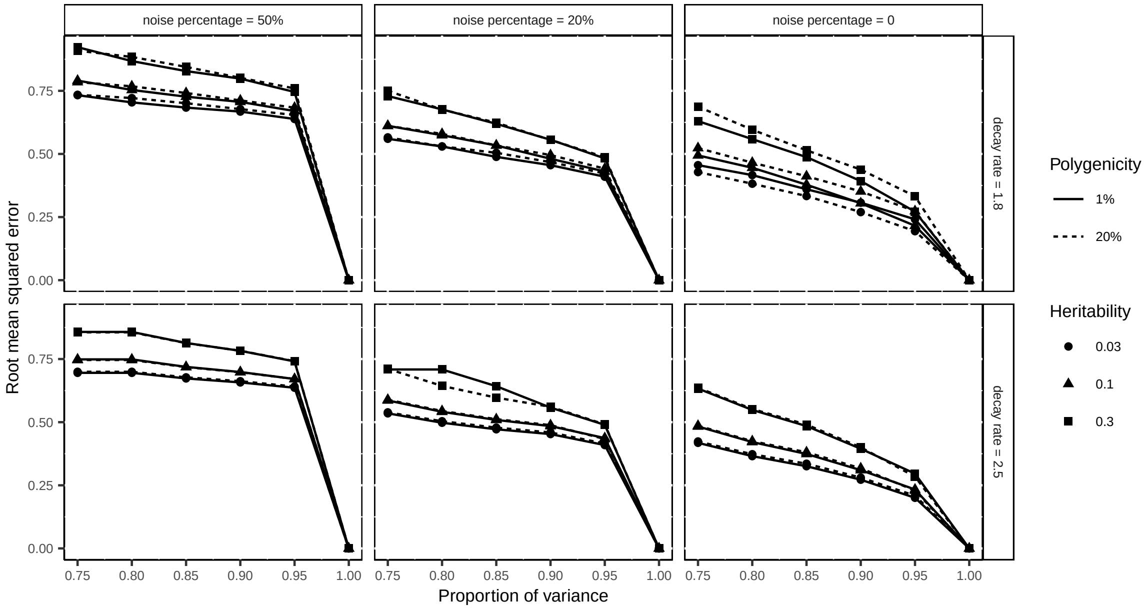

Figure S4: Root mean squared error of z-scores of GWAS summary statistics in simulation studies (Rayleigh noise). Noise percentage: the proportion of image variance contributed by white noise. Decay rate: the polynomial decay rate of image eigenvalues ($\lambda$); the larger the rate, the more rapidly the eigenvalues decay. Scenarios including low/high polygenicity and low/medium/high heritability.

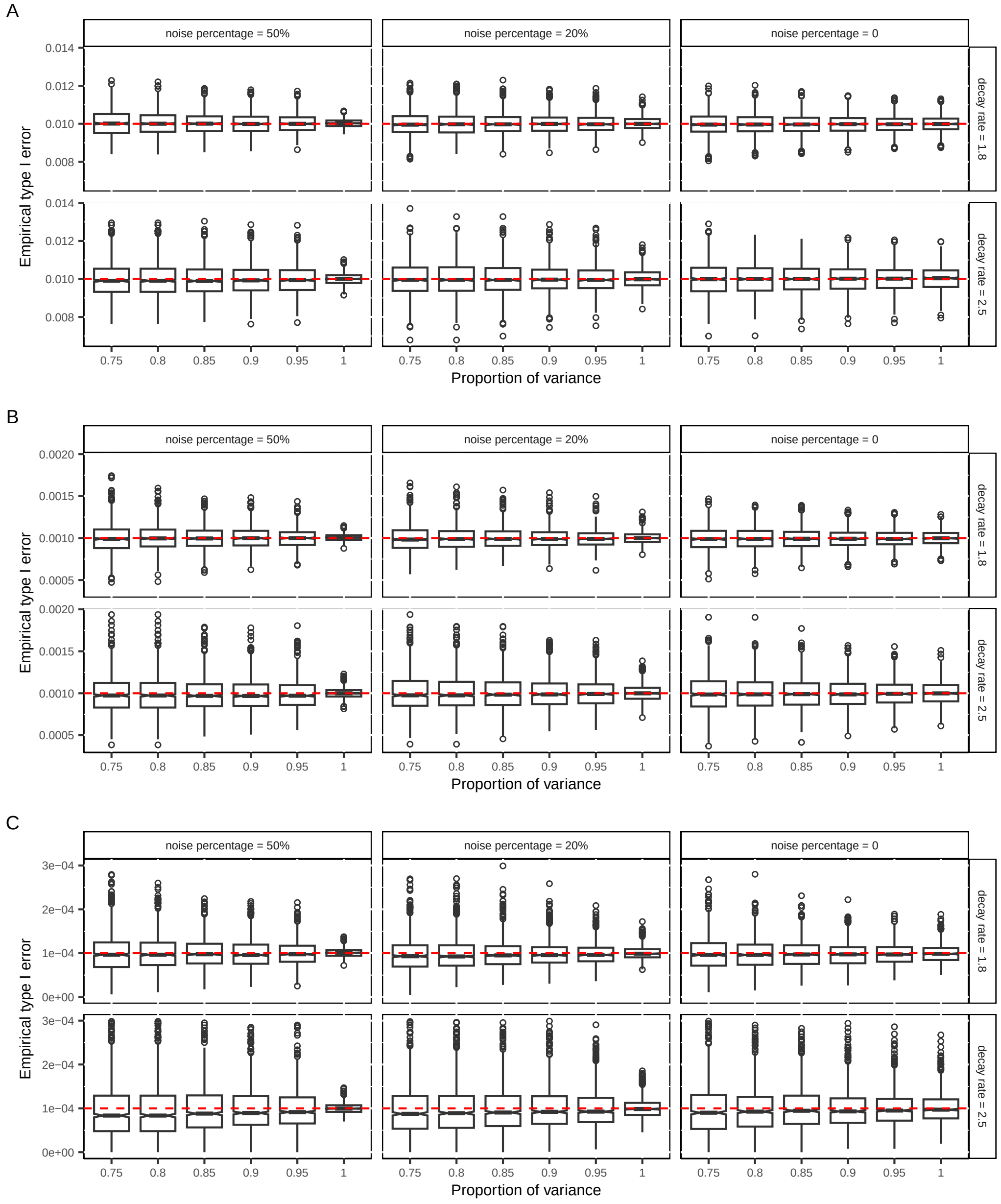

Figure S5: Empirical type I error rate for GWAS summary statistics in simulation studies (Gaussian noise). Since no genotype data was used in data generation, any significant associations are false positive. Each point represents the type I error rate across 200 voxels, and there are 1,000 replicates in each box plot. The line in the box shows the median and the hinge represents an approximate 95% confidence interval around the median. Whiskers represent 1.5 times the interquartile range (IQR). Under a noise percentage of 50%, using raw data (proportion of variance = 100%) resulted in much smaller variance. This is because the strong white noise disrupted the spatial correlation within the images, making each image resemble 200 independent phenotypes ($200 \times 1,000$ effective replicates). The similar reason applies to the slightly higher variance for preserving 75% of variance, where the number of effective replicates is smaller. Noise percentage: the proportion of image variance contributed by white noise. Decay rate: the polynomial decay rate of image eigenvalues ($\lambda$); the larger the rate, the more rapidly the eigenvalues decay. The red line represents significant level. A, Level $10^{-2}$. B, Level $10^{-3}$. C, Level $10^{-4}$.

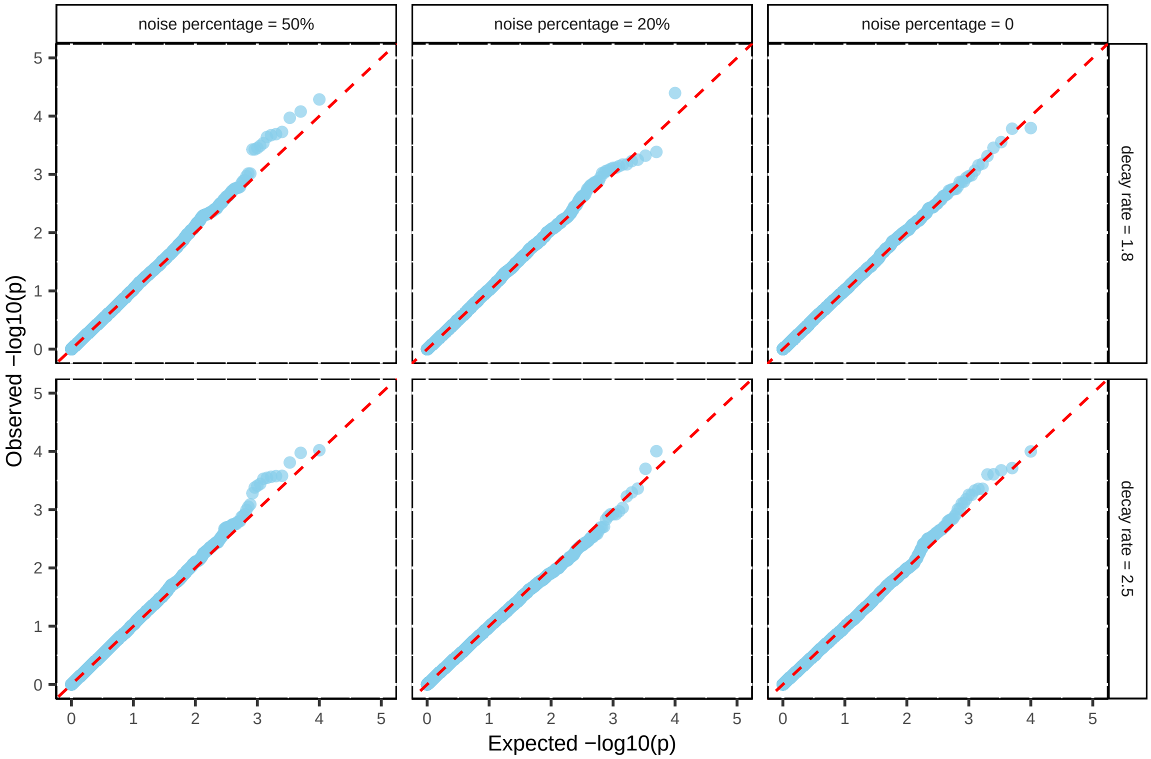

Figure S6: QQ plot of $p$-values in type I error simulation. There are 10,000 $p$-values in each plot representing 10,000 SNPs. Each plot is from a randomly selected voxel with 80% of variance retained.

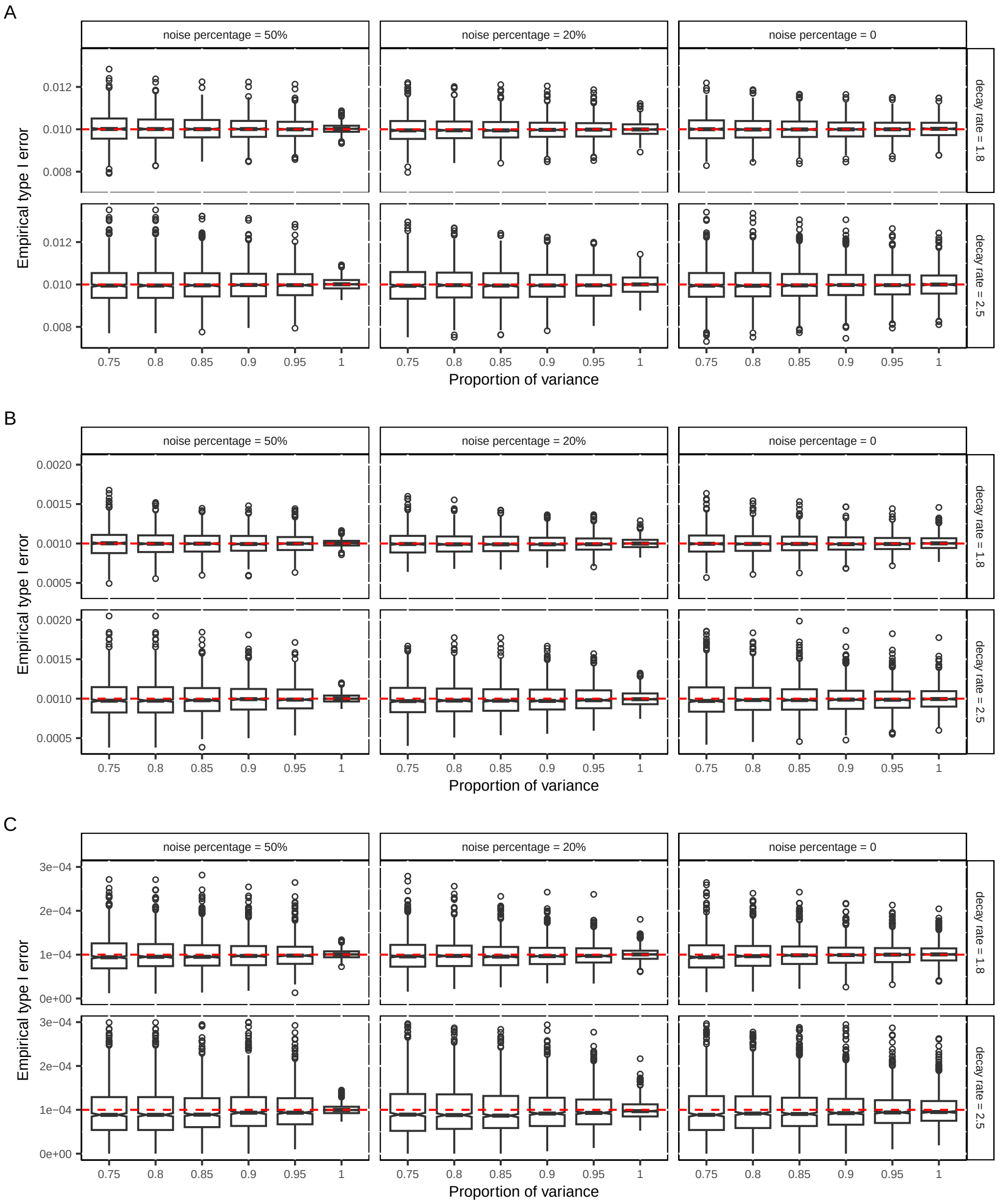

Figure S7: Empirical type I error rate for GWAS summary statistics in simulation studies (Rayleigh noise). Since no genotype data was used in data generation, any significant associations are false positive. Each point represents the type I error rate across 200 voxels, and there are 1,000 replicates in each box plot. The line in the box shows the median and the hinge represents an approximate 95% confidence interval around the median. Whiskers represent 1.5 times the interquartile range (IQR). Under a noise percentage of 50%, using raw data (proportion of variance = 100%) resulted in much smaller variance. This is because the strong white noise disrupted the spatial correlation within the images, making each image resemble 200 independent phenotypes ($200 \times 1,000$ effective replicates). The similar reason applies to the slightly higher variance for preserving 75% of variance, where the number of effective replicates is smaller. Noise percentage: the proportion of image variance contributed by white noise. Decay rate: the polynomial decay rate of image eigenvalues ($\lambda$); the larger the rate, the more rapidly the eigenvalues decay. The red line represents significant level. A, Level $10^{-2}$. B, Level $10^{-3}$. C, Level $10^{-4}$.

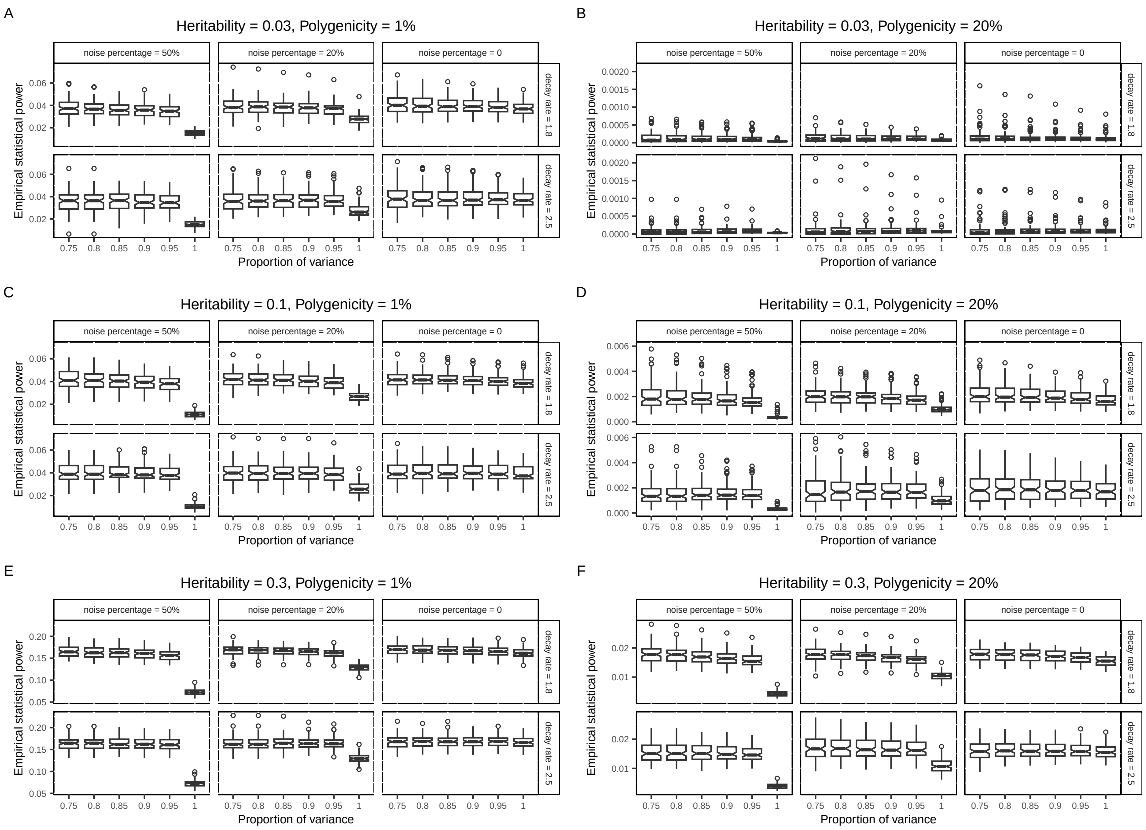

Figure S8: The empirical statistical power in simulation studies (Rayleigh noise). The power is evaluated by the proportion of causal variants being significant at level $10^{-4}$. We used the same genotype data of 10,000 subjects and 21,576 common variants across all simulation setups. There are 216 causal variants for the polygenicity = 1% cases and 4,315 causal variants for the polygenicity = 20% cases. These two numbers are fixed, but the specific causal variants for each replicate are randomly selected. Each voxel in a replicate is affected by the same list of causal variants, but the true genetic effects are varying across voxels. Each point in a box plot represents the average empirical power across 200 voxels for a replicate and there are 100 replicates in each box plot. For example, if the empirical power is 0.01 for the polygenicity = 20% case, on average $4,315 \times 0.01 = 43.15$ causal variants are detected for a voxel. The line in the box shows the median and the hinge represents an approximate 95% confidence interval around the median. Whiskers represent 1.5 times the interquartile range (IQR). The proportion of variance indicates the amount of imaging signals preserved by LDRs. **(A)** The scenario of low heritability and low polygenicity. **(B)** The scenario of low heritability and high polygenicity. **(C)** The scenario of medium heritability and low polygenicity. **(D)** The scenario of medium heritability and high polygenicity. **(E)** The scenario of high heritability and low polygenicity. **(F)** The scenario of high heritability and high polygenicity.

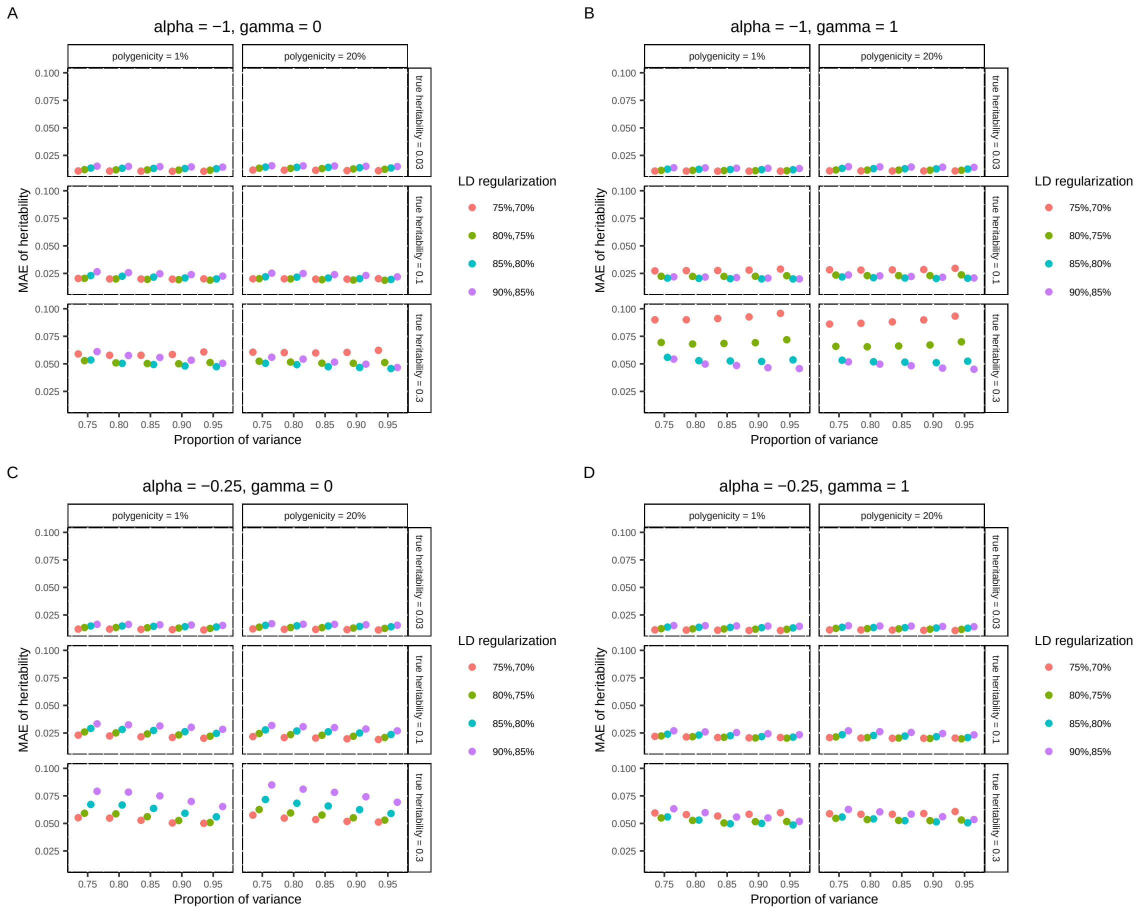

Figure S9: The mean absolute error of heritability estimates across all voxels in simulation studies (genotyped SNPs). The proportion of variance indicates the amount of signals preserved by LDRs. LD regularization indicates the regularization imposed on the LD matrix and its inverse. A, The genetic effects have a strong correlation with MAF but no dependence on the LD. B, The genetic effects have a strong correlation with MAF and strong dependence on the LD. C, The genetic effects have a weak correlation with MAF and no dependence on the LD. D, The genetic effects have a weak correlation with MAF but strong dependence on the LD.

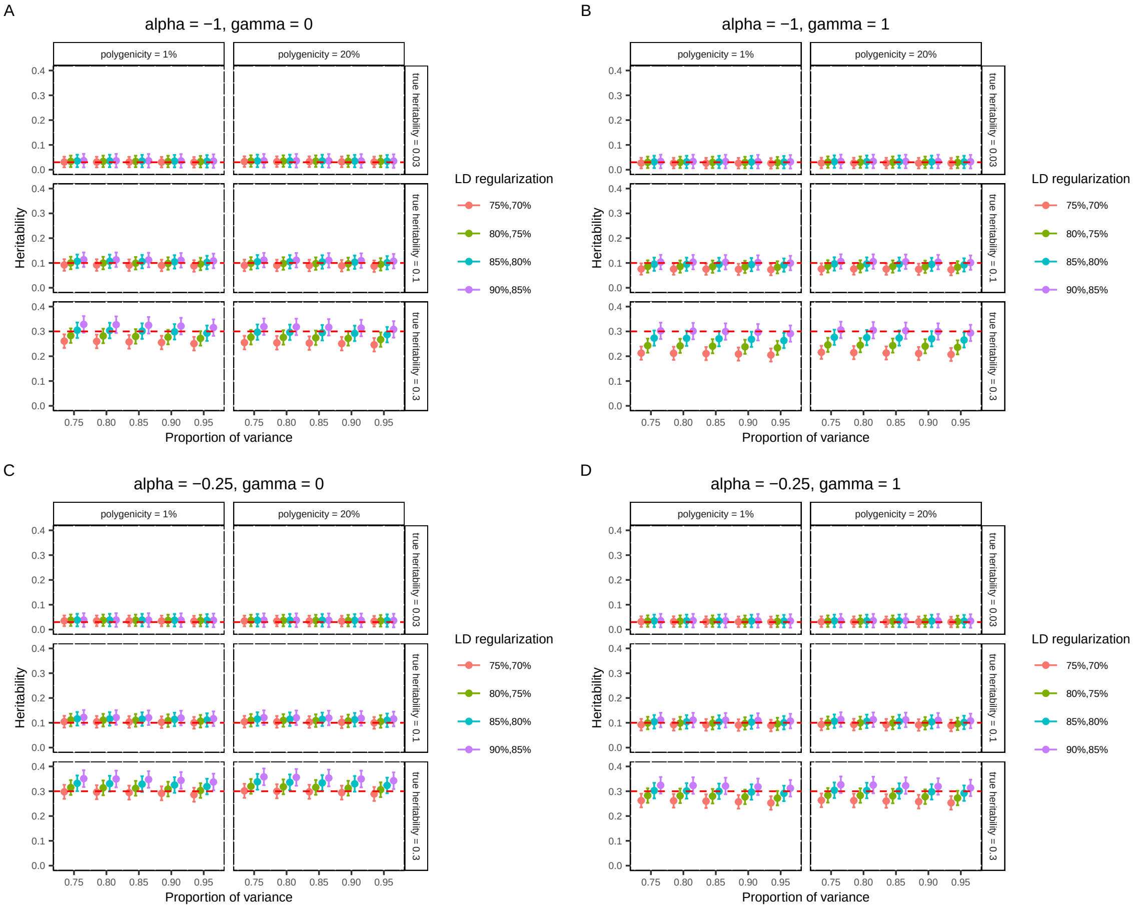

Figure S10: The mean and standard error of heritability estimates in simulation studies (genotyped SNPs). The red dashed lines denote the true heritability of each voxel. The dots show the mean heritability estimates across all voxels. The error bars show the 95% confidence intervals constructed by mean estimates $\pm 1.96$ mean standard error estimates across all voxels. The proportion of variance indicates the amount of signals preserved by LDRs. LD regularization indicates the regularization imposed on the LD matrix and its inverse. A, The genetic effects have a strong correlation with MAF but no dependence on the LD. B, The genetic effects have a strong correlation with MAF and strong dependence on the LD. C, The genetic effects have a weak correlation with MAF and no dependence on the LD. D, The genetic effects have a weak correlation with MAF but strong dependence on the LD.

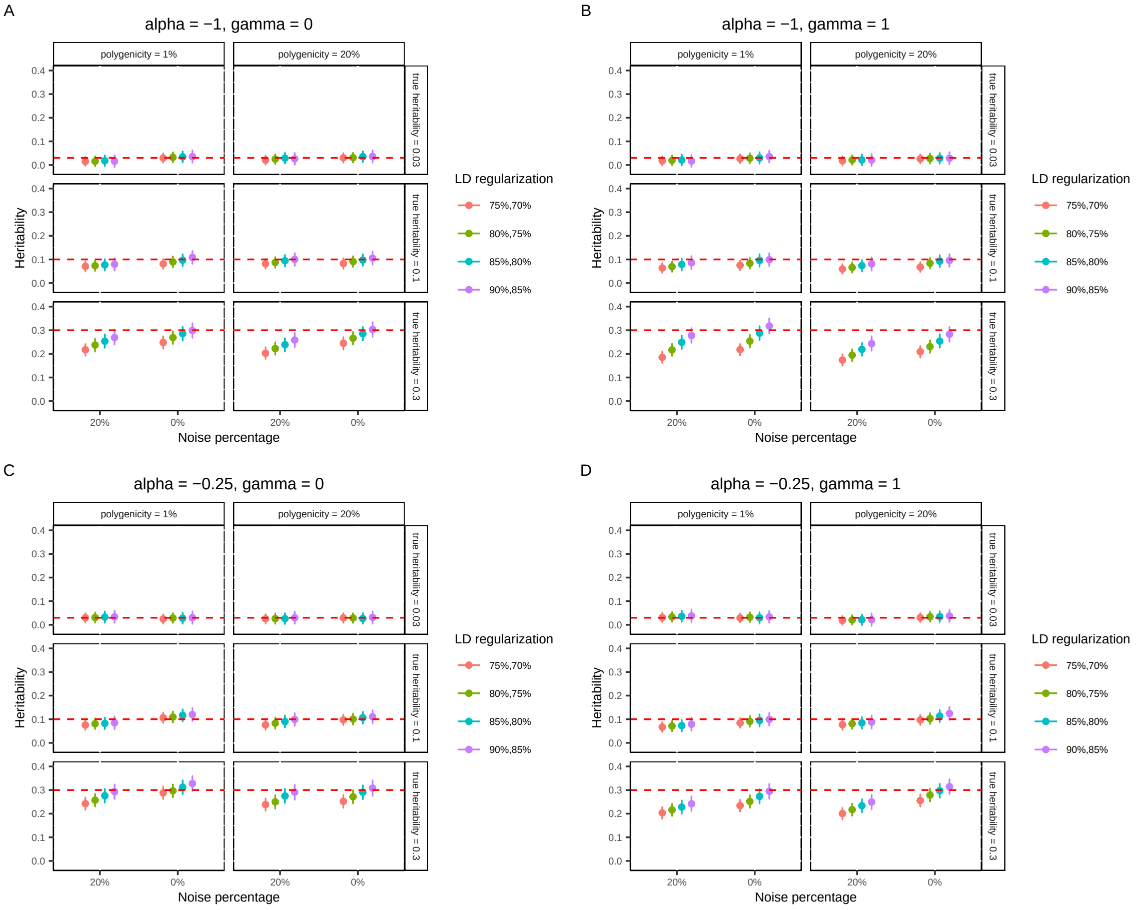

Figure S11: The mean and standard error of heritability estimates comparing images with and without noise (genotyped SNPs). RVGA heritability estimator was applied to each voxel individually. The red dashed lines denote the true heritability of each voxel. The dots show the mean heritability estimates across all voxels. The error bars show the 95% confidence intervals constructed by mean estimates $\pm 1.96$ mean standard error estimates across all voxels. LD regularization indicates the regularization imposed on the LD matrix and its inverse. **(A)** The genetic effects have a strong correlation with MAF but no dependence on the LD. **(B)** The genetic effects have a strong correlation with MAF and strong dependence on the LD. **(C)** The genetic effects have a weak correlation with MAF and no dependence on the LD. **(D)** The genetic effects have a weak correlation with MAF but strong dependence on the LD.

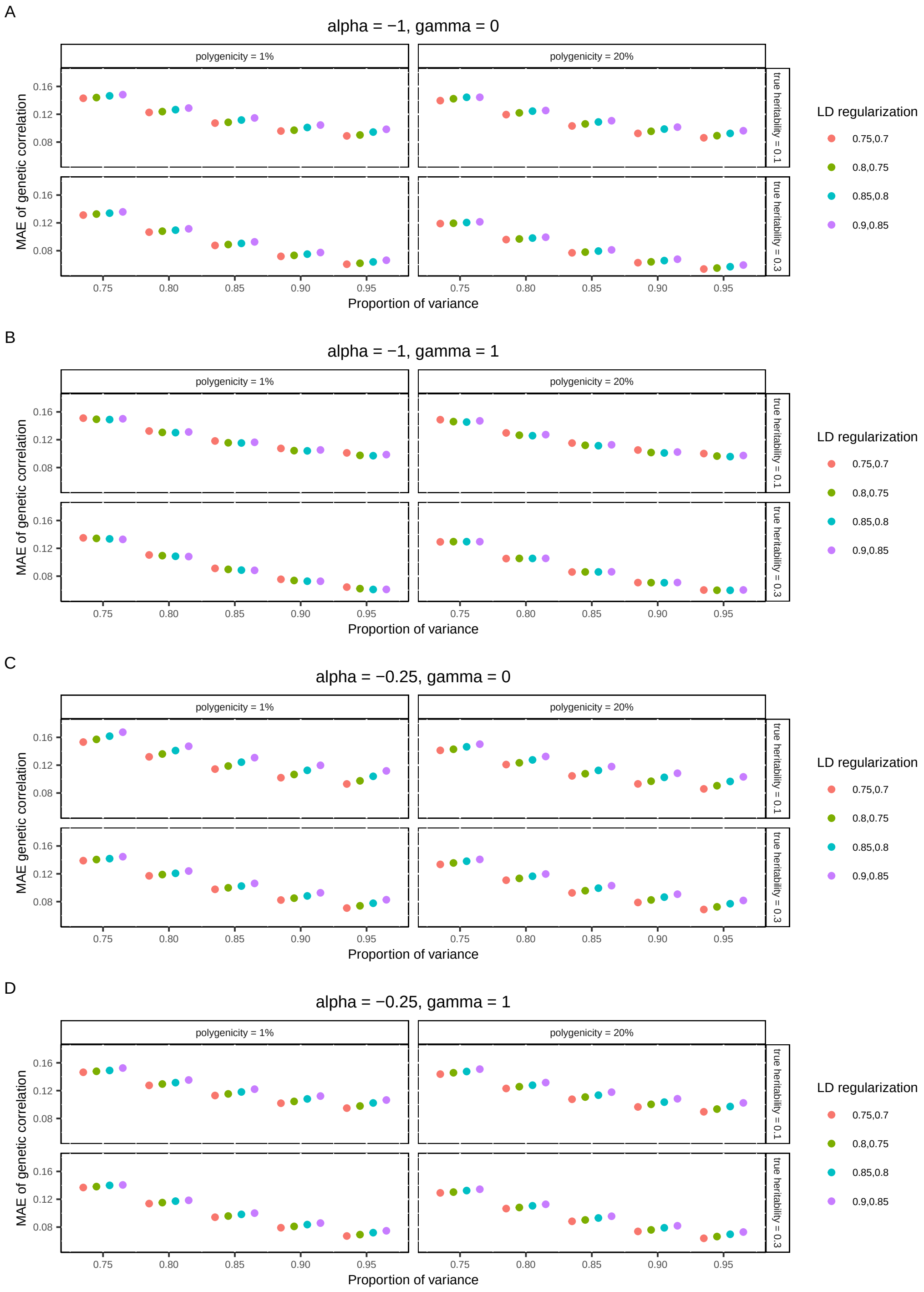

Figure S12: The mean absolute error of genetic correlation estimates across all voxel pairs in simulation studies (genotyped SNPs). The proportion of variance indicates the amount of signals preserved by LDRs. LD regularization indicates the regularization imposed on the LD matrix and its inverse. A, The genetic effects have a strong correlation with MAF but no dependence on the LD. B, The genetic effects have a strong correlation with MAF and strong dependence on the LD. C, The genetic effects have a weak correlation with MAF and no dependence on the LD. D, The genetic effects have a weak correlation with MAF but strong dependence on the LD.

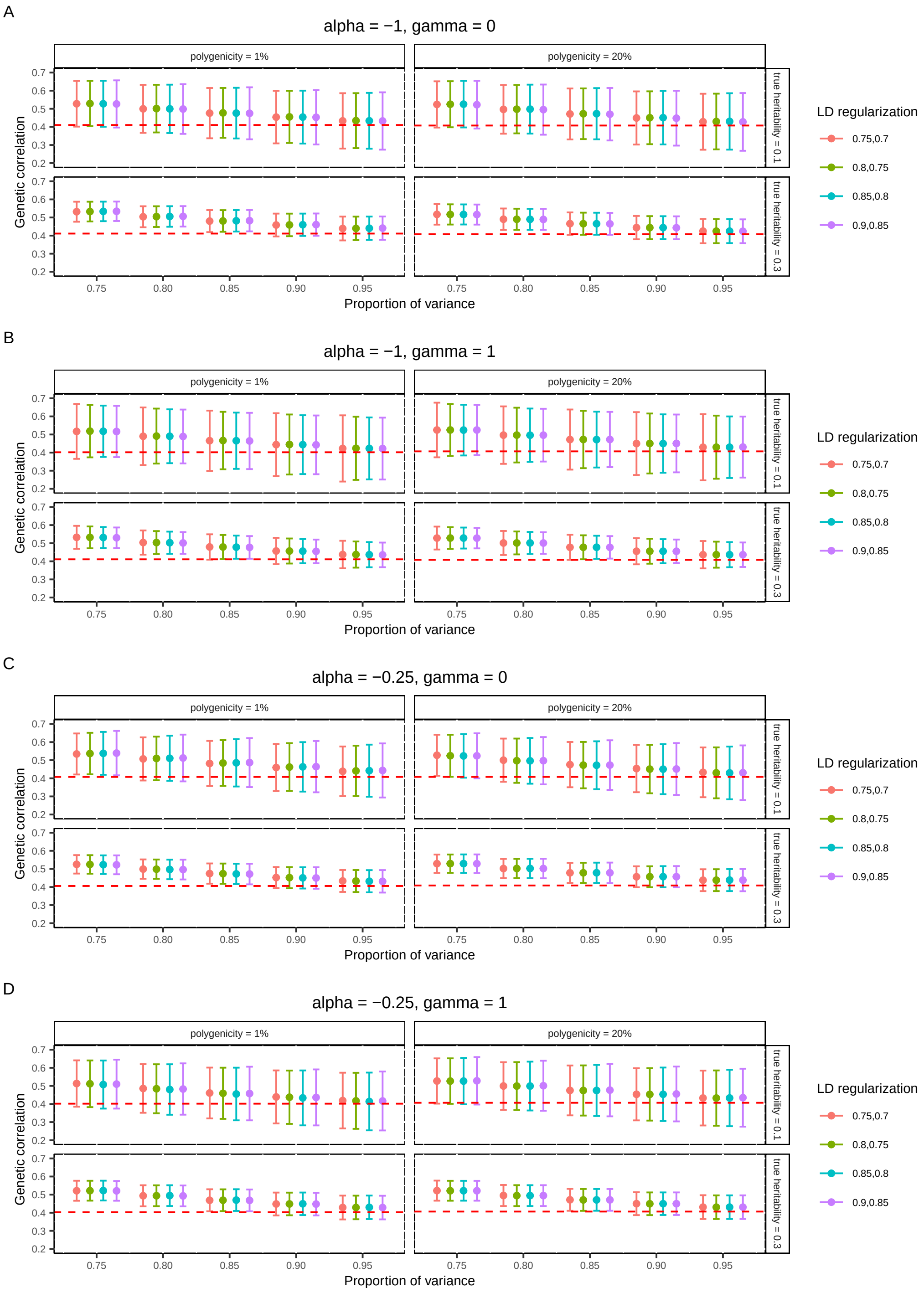

Figure S13: The mean and standard error of genetic correlation estimates in simulation studies (genotyped SNPs). The red dashed lines denote the true mean genetic correlation across all voxel pairs. The dots show the mean heritability estimates across all voxels. The error bars show the 95% confidence intervals constructed by mean estimates $\pm 1.96$ mean standard error estimates across all voxel pairs. The proportion of variance indicates the amount of signals preserved by LDRs. LD regularization indicates the regularization imposed on the LD matrix and its inverse. A, The genetic effects have a strong correlation with MAF but no dependence on the LD. B, The genetic effects have a strong correlation with MAF and strong dependence on the LD. C, The genetic effects have a weak correlation with MAF and no dependence on the LD. D, The genetic effects have a weak correlation with MAF but strong dependence on the LD.

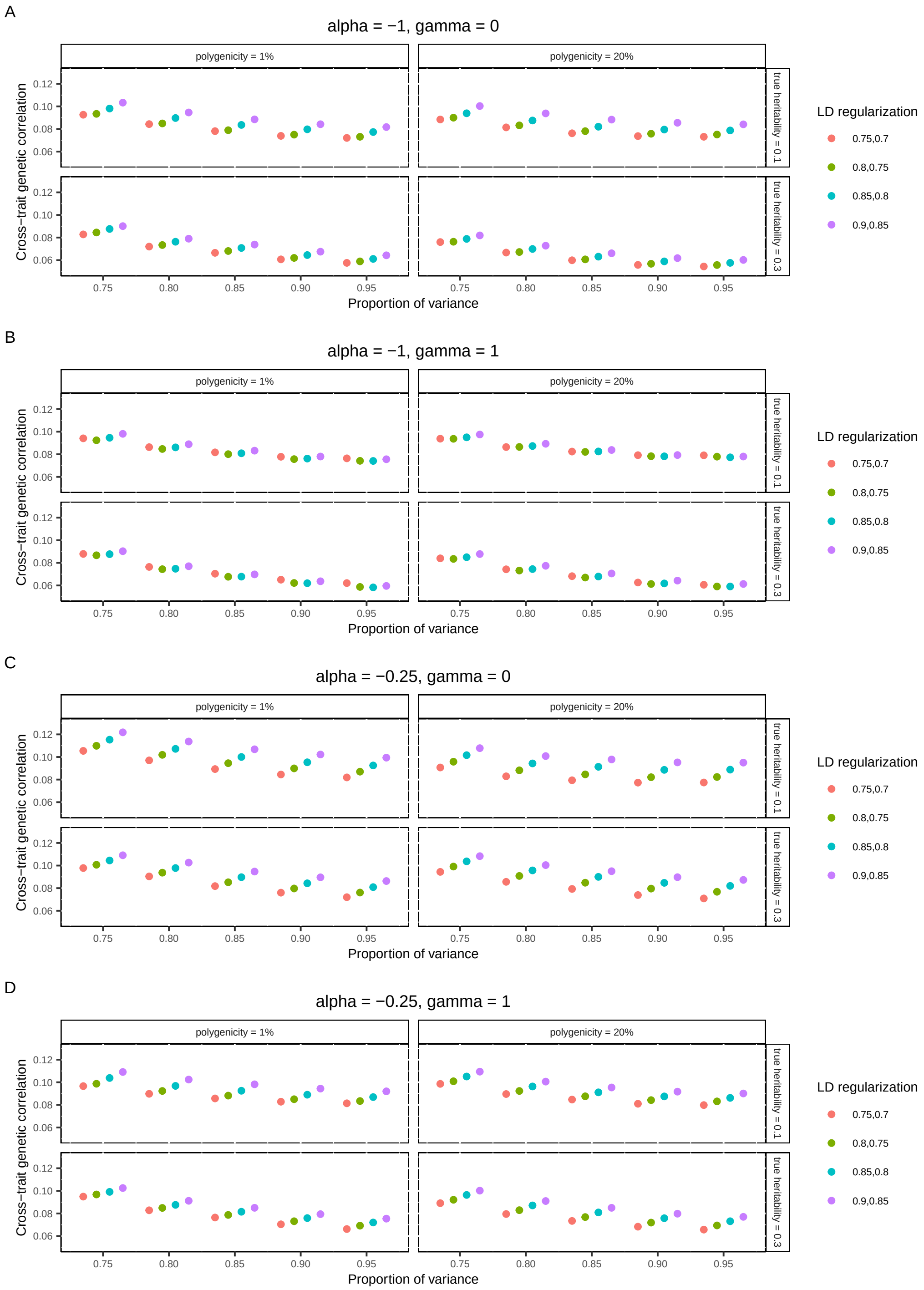

Figure S14: The mean absolute error of cross-trait genetic correlation estimates across all voxels in simulation studies (genotyped SNPs). The proportion of variance indicates the amount of signals preserved by LDRs. LD regularization indicates the regularization imposed on the LD matrix and its inverse. A, The genetic effects have a strong correlation with MAF but no dependence on the LD. B, The genetic effects have a strong correlation with MAF and strong dependence on the LD. C, The genetic effects have a weak correlation with MAF and no dependence on the LD. D, The genetic effects have a weak correlation with MAF but strong dependence on the LD.

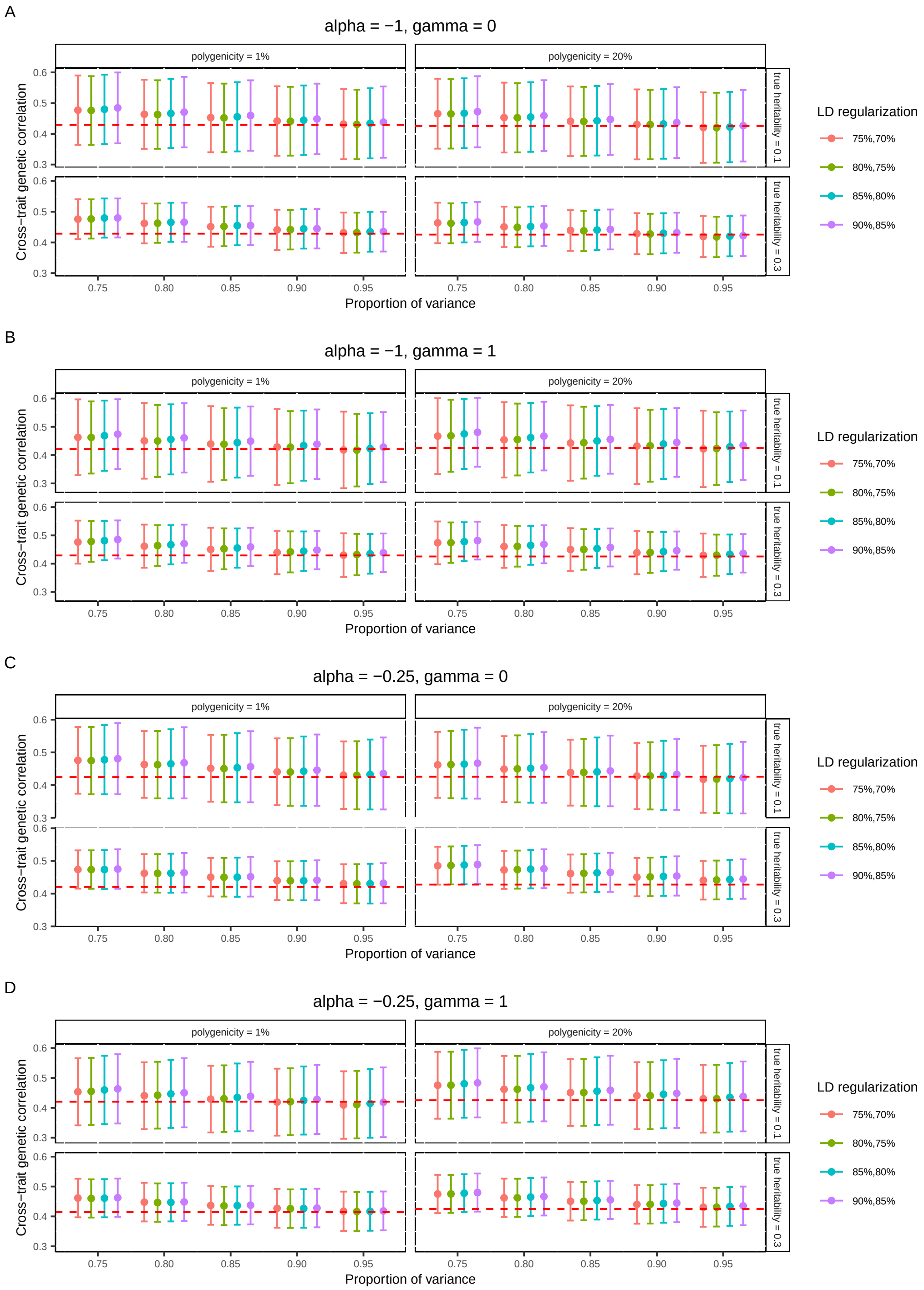

Figure S15: The mean and standard error of cross-trait genetic correlation estimates in simulation studies (genotyped SNPs). The red dashed lines denote the true mean genetic correlation across all voxel pairs. The dots show the mean heritability estimates across all voxels. The error bars show the 95% confidence intervals constructed by mean estimates ±1.96 mean standard error estimates across all voxels. The proportion of variance indicates the amount of signals preserved by LDRs. LD regularization indicates the regularization imposed on the LD matrix and its inverse. A, The genetic effects have a strong correlation with MAF but no dependence on the LD. B, The genetic effects have a strong correlation with MAF and strong dependence on the LD. C, The genetic effects have a weak correlation with MAF and no dependence on the LD. D, The genetic effects have a weak correlation with MAF but strong dependence on the LD.

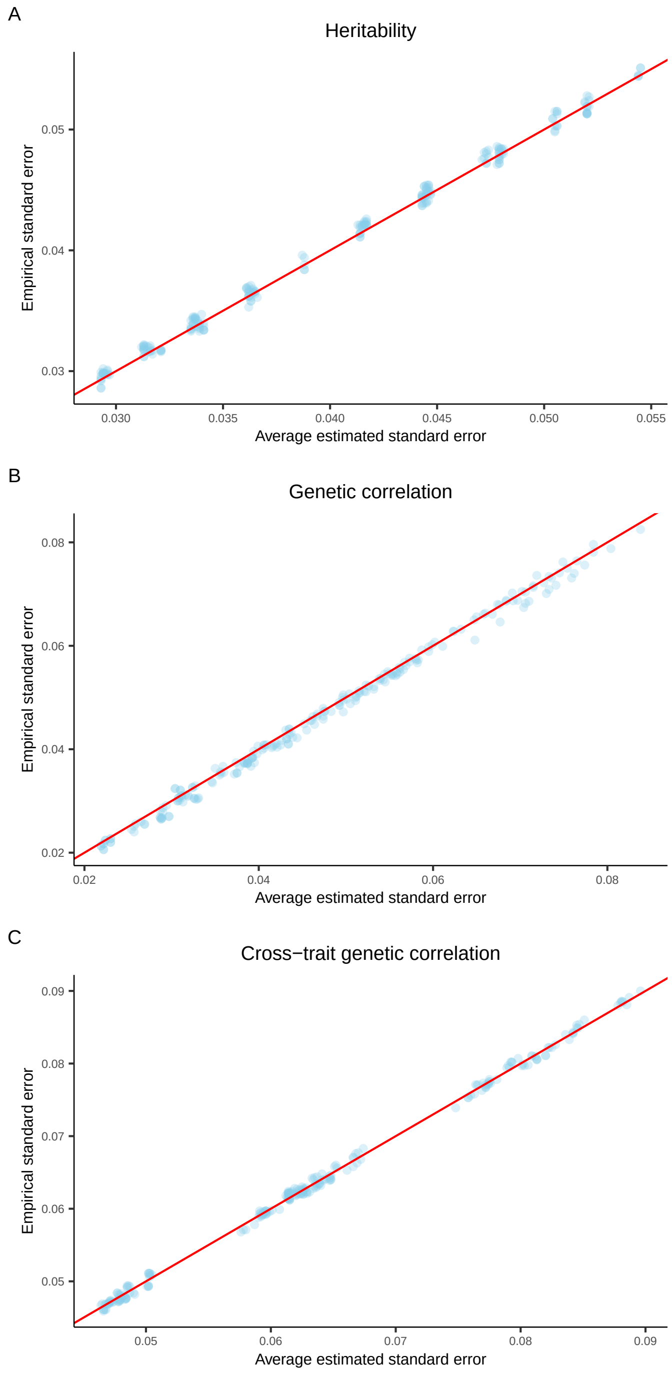

Figure S16: Comparisons of average standard error estimates and empirical standard error estimates in simulation studies. There were 100 replicates for each setup. A, Heritability. B, Genetic correlation. C, Cross-trait genetic correlation.

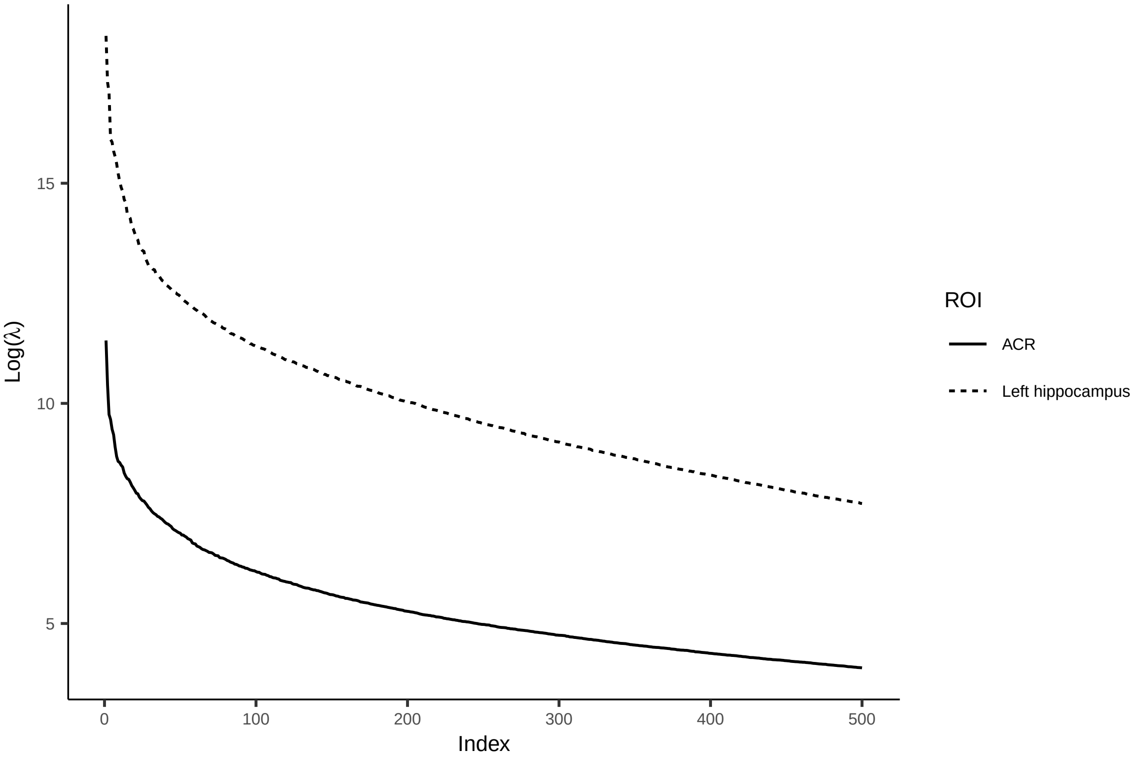

Figure S17: Decay rate of logarithmic eigenvalues for the left hippocampus and the anterior corona radiata. Only the top 500 eigenvalues are shown.

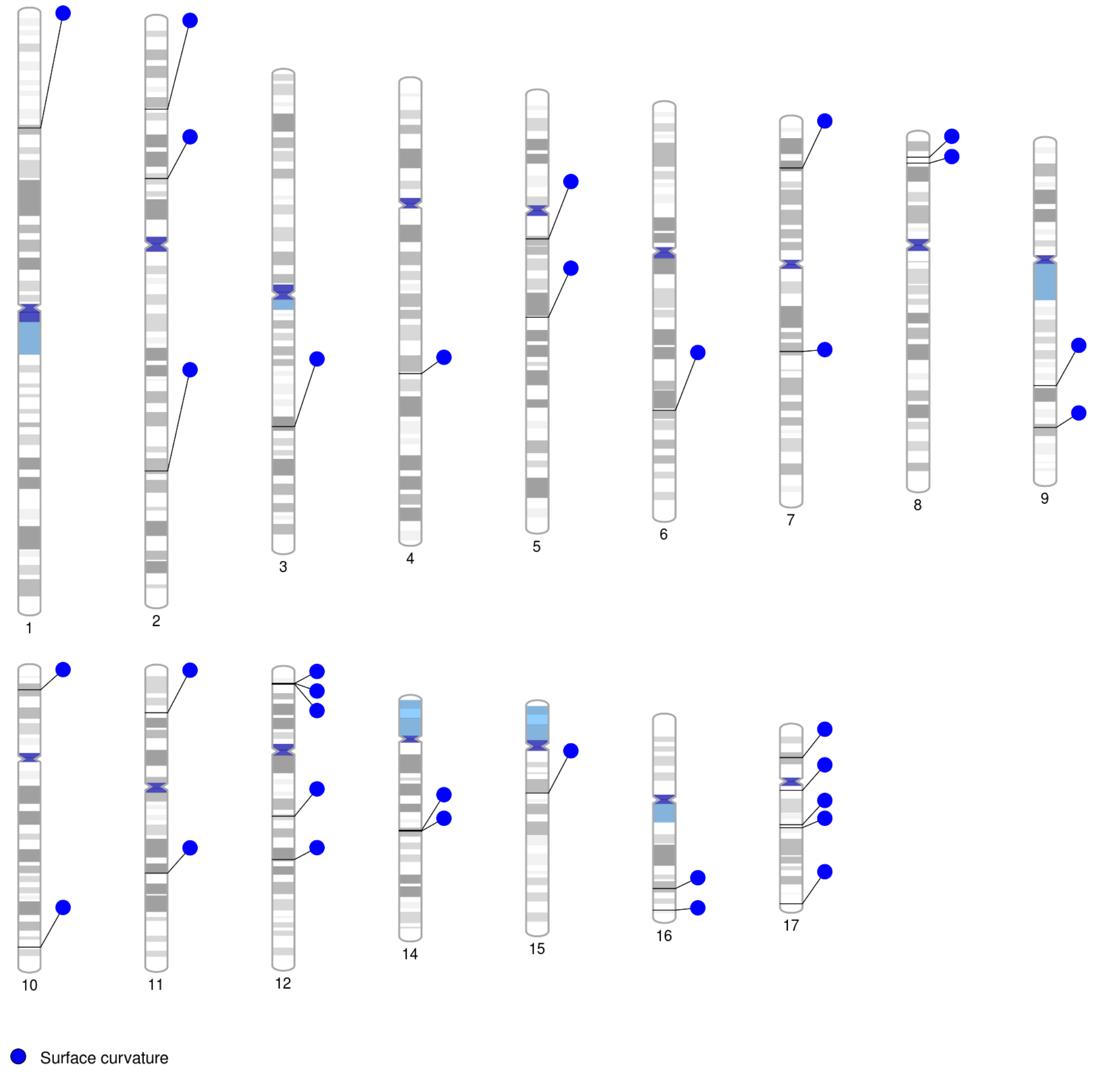

Figure S18: Significant and stringently replicated loci associated with the cortical surface curvature ($P < 3.15 \times 10^{-11}$). We used UKB phase 3 data (n = 15,752) for discovery and phase 1 and 2 data (n = 12,431) for replication. The significant vertex-variant associations were aggregated using the Peaks algorithm, producing 35 significant loci after cluster analysis, all of which were replicated ($P < 0.05/35/1585.3$).

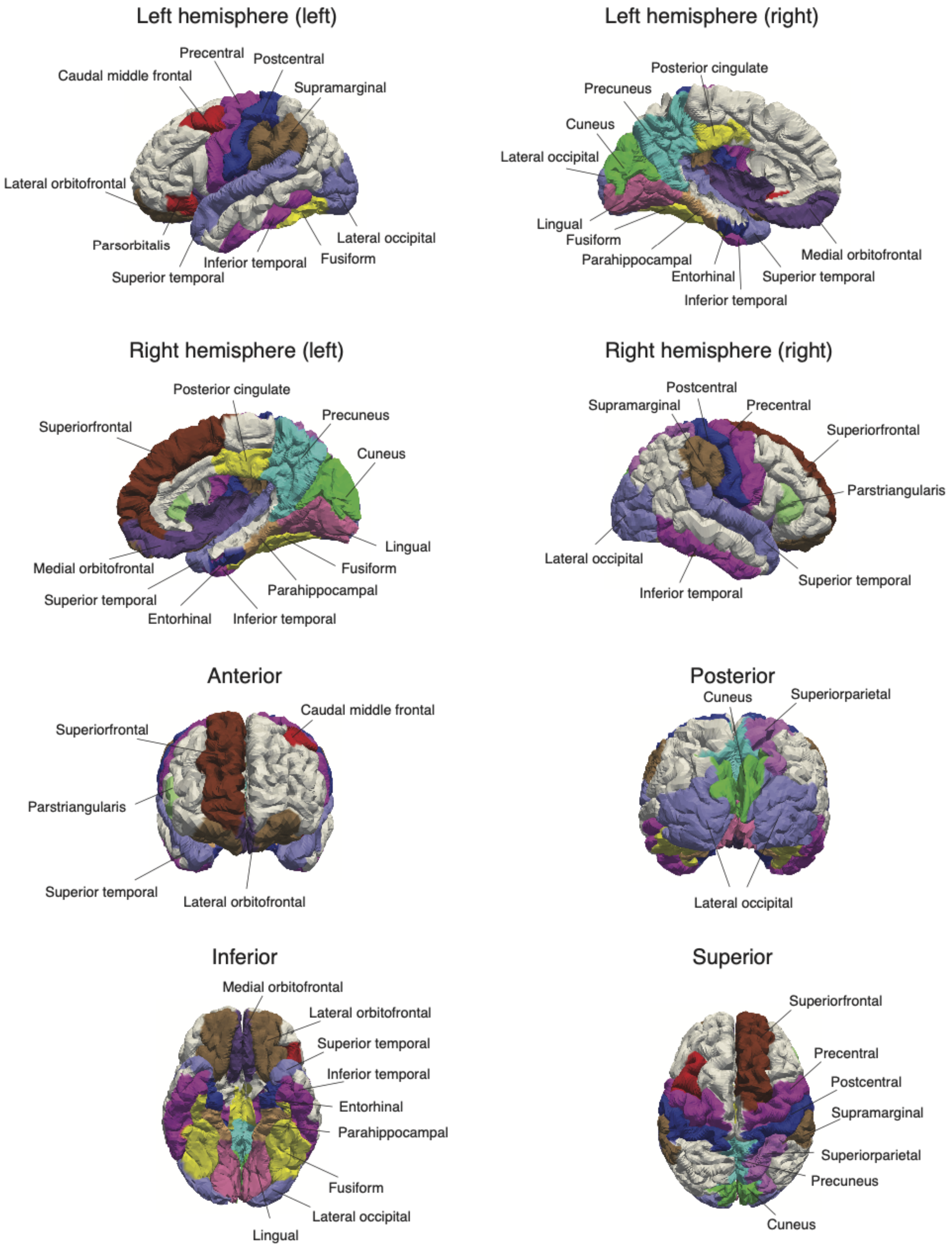

Figure S19: ROIs with at least one significant vertex-variant association ($P < 3.15 \times 10^{-11}$) for the cortical surface curvature based on the Desikan-Killiany-Tourville (DKT) atlas. The ROI names are annotated.

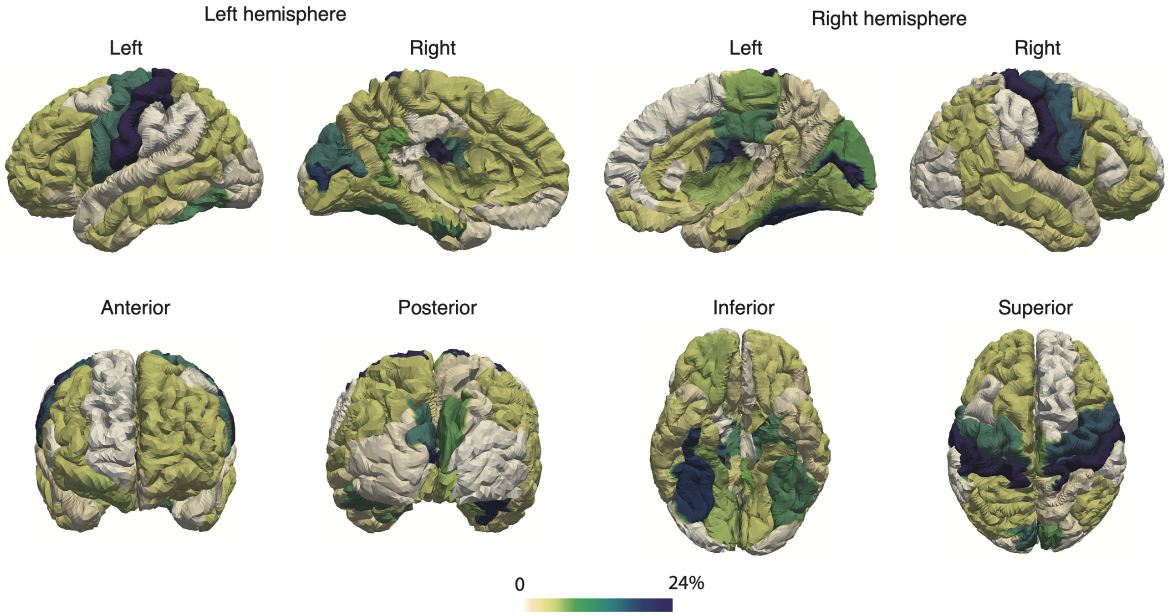

Figure S20: Percentage of vertices significantly associated with at least one genetic variant in each ROI for the cortical surface curvature.

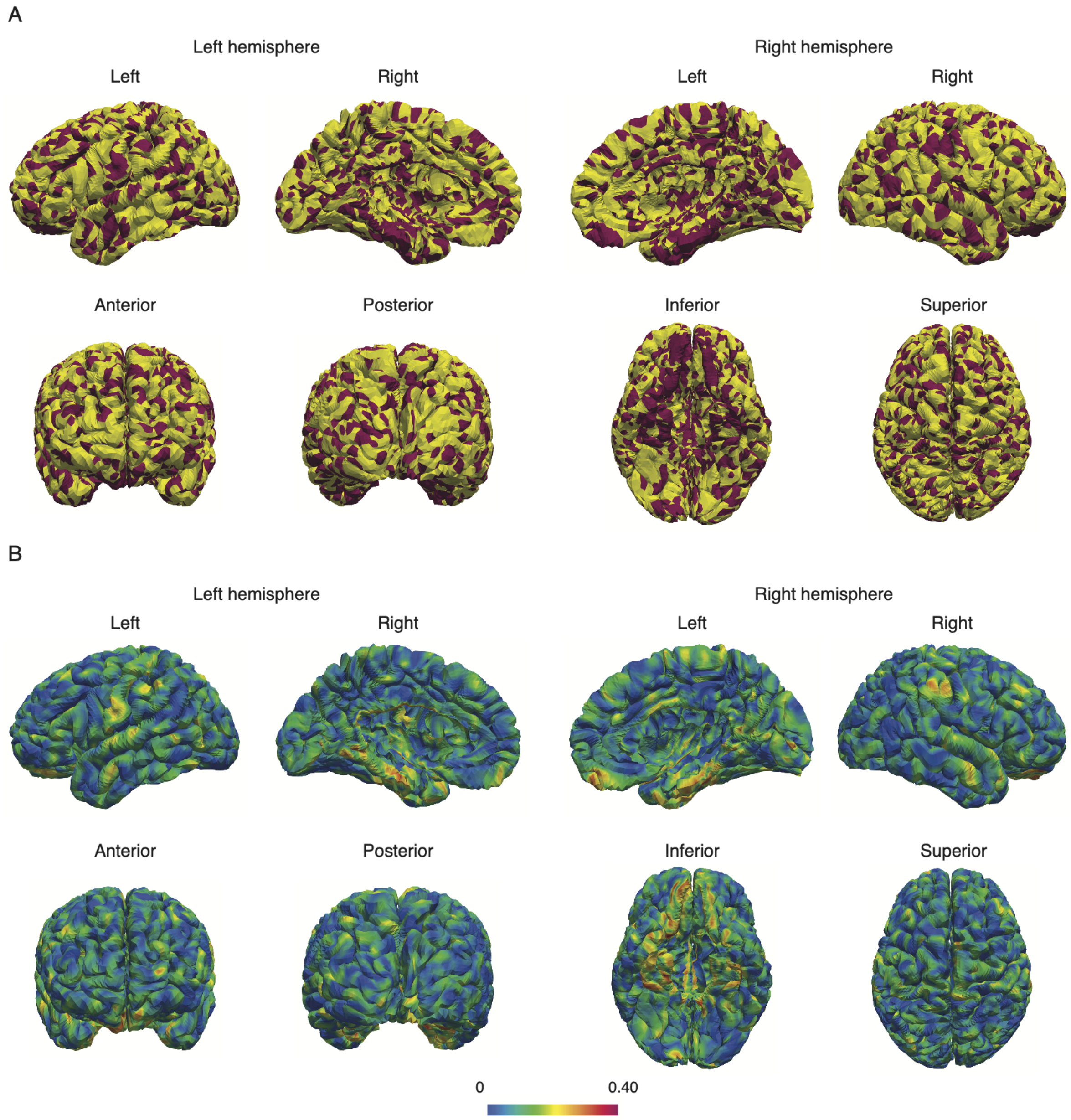

Figure S21: Vertex heritability of the cortical surface curvature. A, Highlighted regions with significant heritability estimates ($P < 0.05/1585.3$). B, An atlas of vertex heritability.

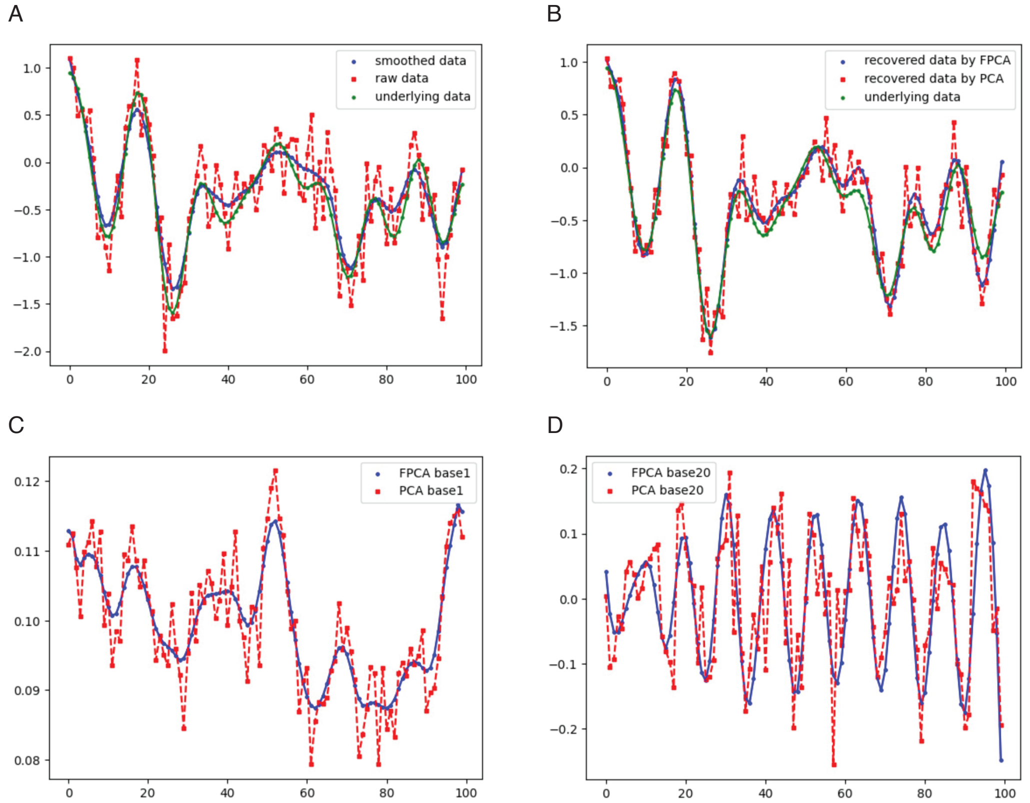

Figure S22: A comparison of FPCA and PCA for imaging data recovery and white noise removal. The simulated dataset contains 100 images each with 100 voxels and each image has a noise percentage of 0.1. A randomly selected image is shown. A, A comparison of underlying data, raw data (the underlying data plus white noise), and smoothed data. B, A comparison of recovered data by FPCA and PCA using the top 20 bases, respectively, and the underlying data. C, The first base of FPCA and PCA, respectively. D, The 20th base of of FPCA and PCA, respectively.

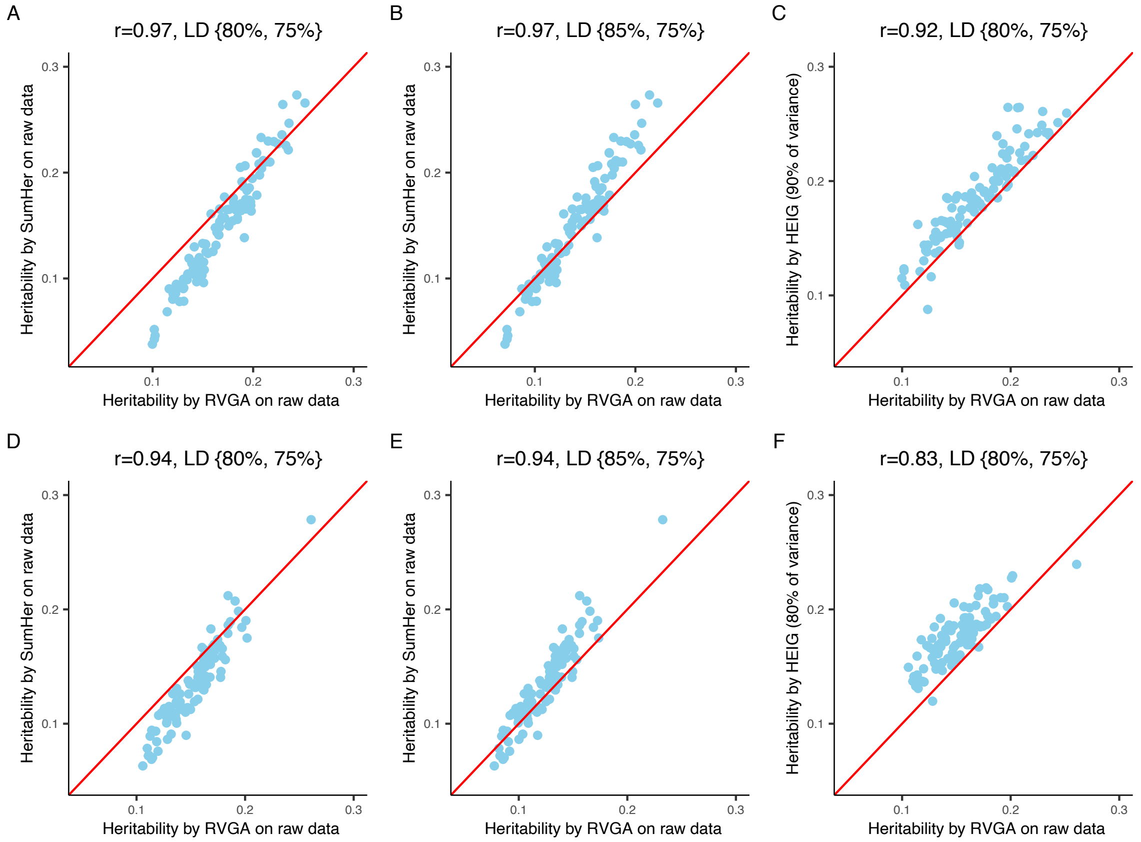

Figure S23: Illustrating the consistent heritability estimates between RVGA and SumHer on raw data using genotyped variants, and RVGA augmenting genetic signals if denoising images using FPCA. The first two columns show how we fine-tune the optimal regularization for LD matrix by comparing RVGA and SumHer estimates on raw data. The third column shows increasing heritability by comparing RVGA estimates on raw data with those on data where a certain proportion of variance is preserved. A-C, The left hippocampus. D-F, The superior fronto-occipital fasciculus.

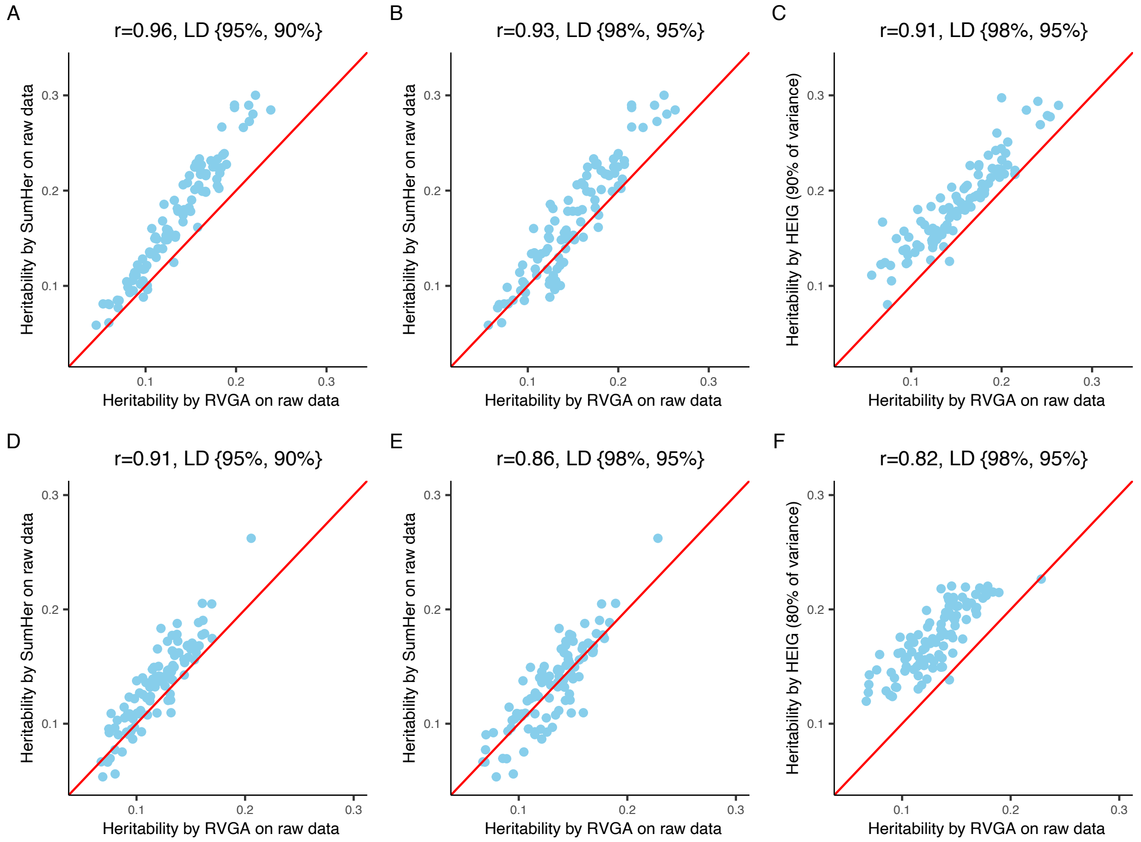

Figure S24: Illustrating the consistent heritability estimates between RVGA and SumHer on raw data using HapMap3 variants, and RVGA augmenting genetic signals if denoising images using FPCA. The first two columns show how we fine-tune the optimal regularization for LD matrix by comparing RVGA and SumHer estimates on raw data. The third column shows increasing heritability by comparing RVGA estimates on raw data with those on data where a certain proportion of variance is preserved. A-C, The left hippocampus. D-F, The superior fronto-occipital fasciculus.

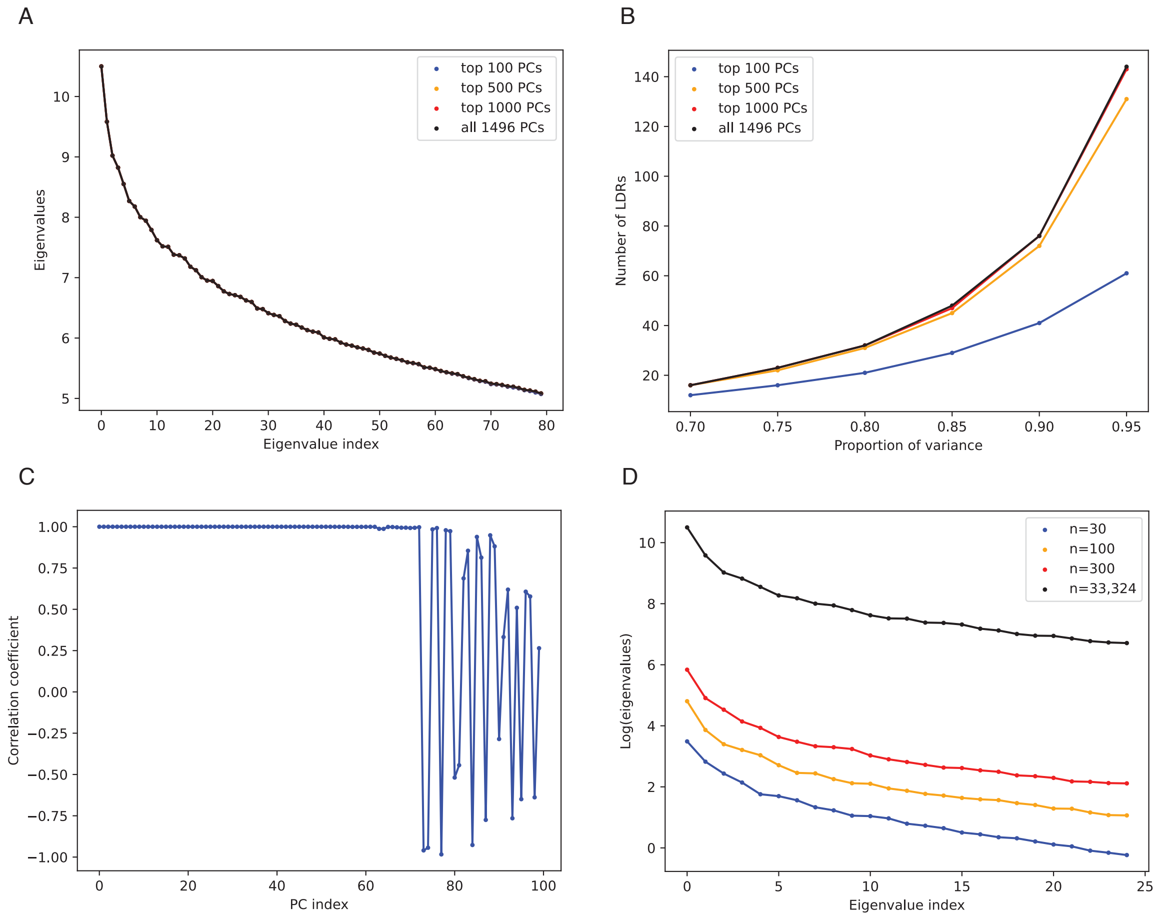

Figure S25: A comparison of computing only top $k$ PCs and all PCs in FPCA. The data is smoothed retrolenticular part of the internal capsule which contains 1,496 voxels. We used all subjects (n = 33,324) to compute the top $k$ PCs in A, B, and C, while we used only n = 30, 100, and 300 subjects in D. A, The top 80 eigenvalues from computing only the top $k$ PCs using all subjects. B, The relationship between the number of LDRs and the proportion of variance when computing only the top $k$ PCs. C, The correlation coefficients of bases from computing top 100 PCs and those from computing all PCs. D, The distributions of top 25 eigenvalues computed using only $n$ subjects (smaller than the image resolution).

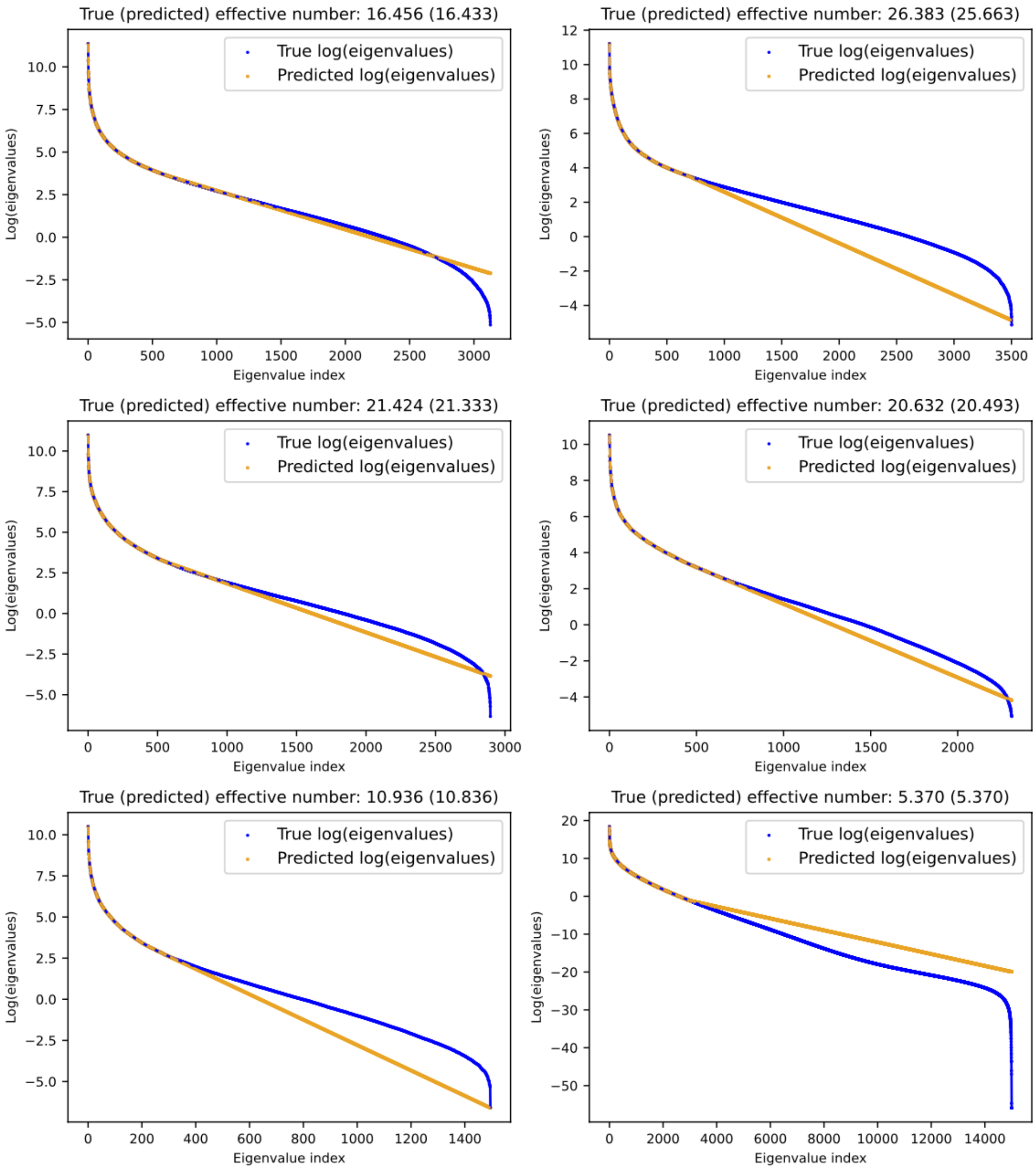

Figure S26: Prediction of the effective number using the top $k$ eigenvalues for six randomly selected ROIs. We used the top $k$ eigenvalues to train a B-spline (degree = 1) to predict uncomputed eigenvalues, where $k$ was 20% of image resolution. The predicted effective numbers using the predicted eigenvalues were very close to the true effective numbers, which were computed using all eigenvalues.

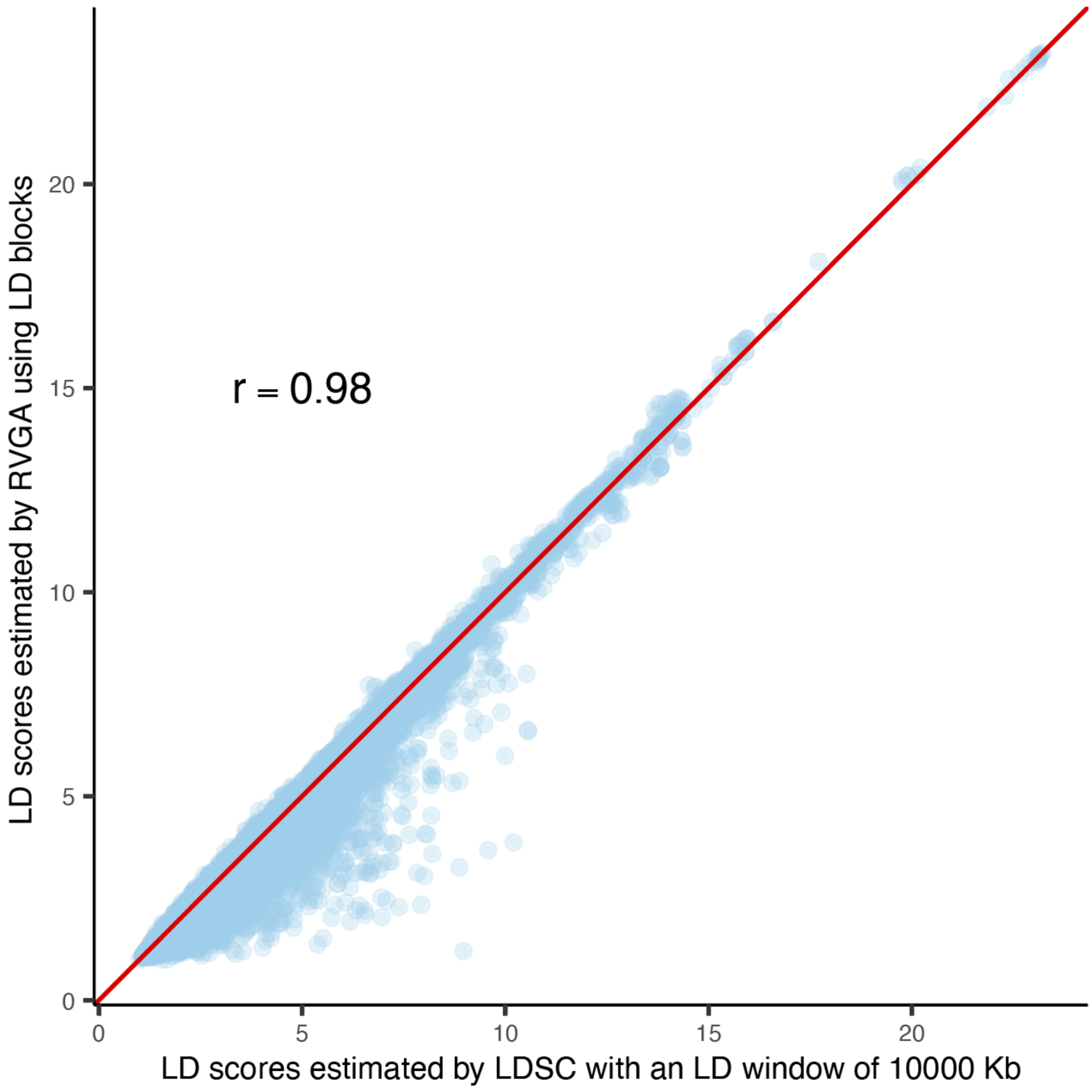

Figure S27: Comparison of LD scores estimated by RVGA and LDSC. LD scores for chromosome 1 estimated using genotype array data with 36, 578 common SNPs (MAF > 0.01) and 9, 200 unrelated subjects with European ancestry in UKB. RVGA calculates LD scores within mutually independent LD blocks, whereas LDSC uses a fixed window of 10, 000 Kb around the index SNP to calculate LD scores. More details include in the Methods and Supplementary Note. r: Pearson correlation coefficient.

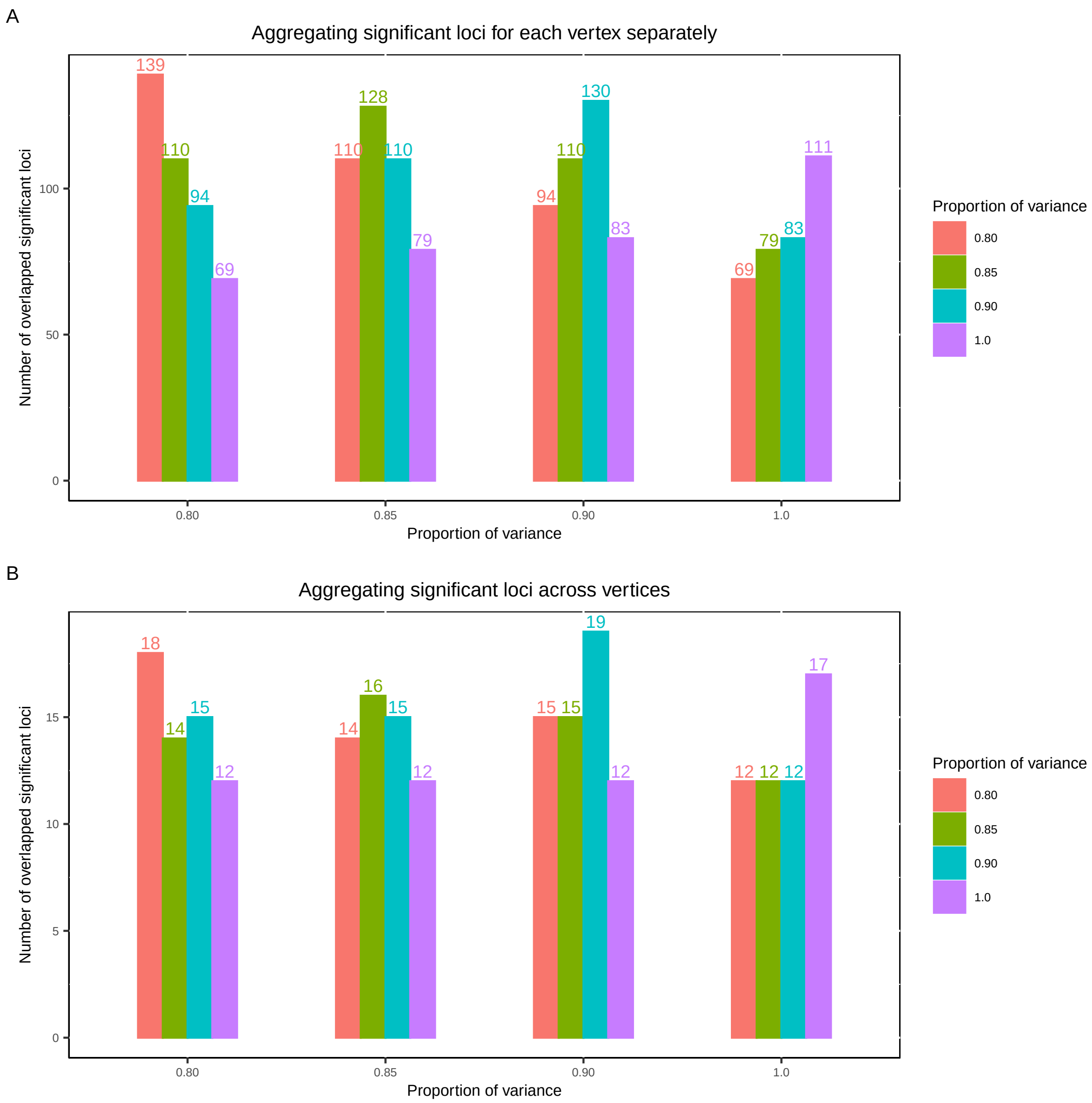

Figure S28: RVGA GWAS sensitivity analysis using 100 vertices in the left hippocampus by preserving varying proportions of variance. Each bar shows the number of overlapped significant loci ($P < 5\times10^{-8}$) between two setups. For example in A, 139 loci were identified for 80% of variance (overlapped with itself), and there were 110 shared loci with using 85% of variance. A, Significant variant-vertex pairs were aggregated for each vertex separately using the Peaks algorithm. B, Significant variant-vertex pairs were aggregated across vertices.

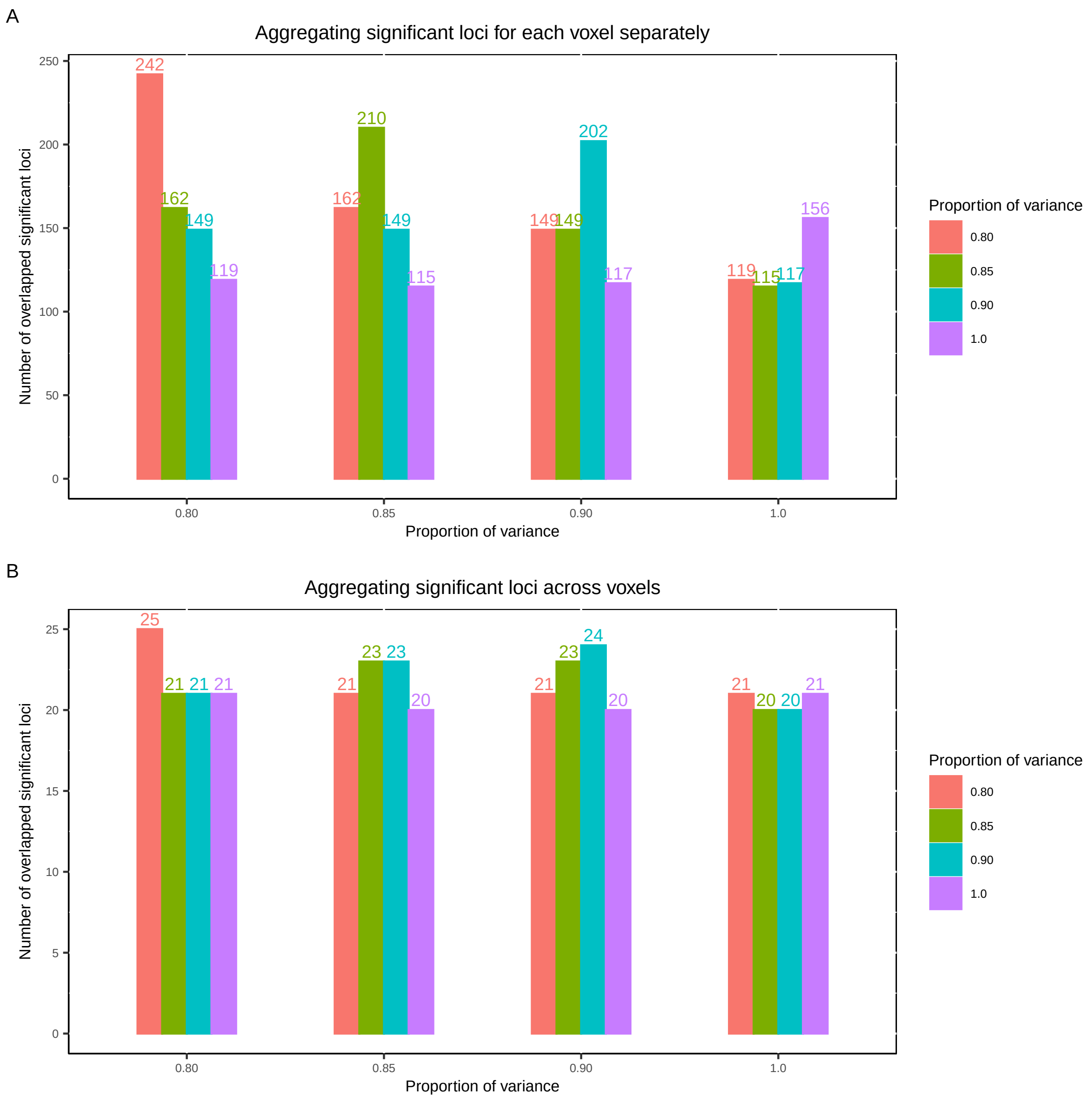

Figure S29: RVGA GWAS sensitivity analysis using 100 voxels in the the superior fronto-occipital fasciculus by preserving varying proportions of variance. Each bar shows the number of overlapped significant loci ($P < 5 \times 10^{-8}$) between two setups. For example in A, 242 loci were identified for 80% of variance (overlapped with itself), and there were 162 shared loci with using 85% of variance. A, Significant variant-voxel pairs were aggregated for each voxel separately using the Peaks algorithm. B, Significant variant-voxel pairs were aggregated across voxels.

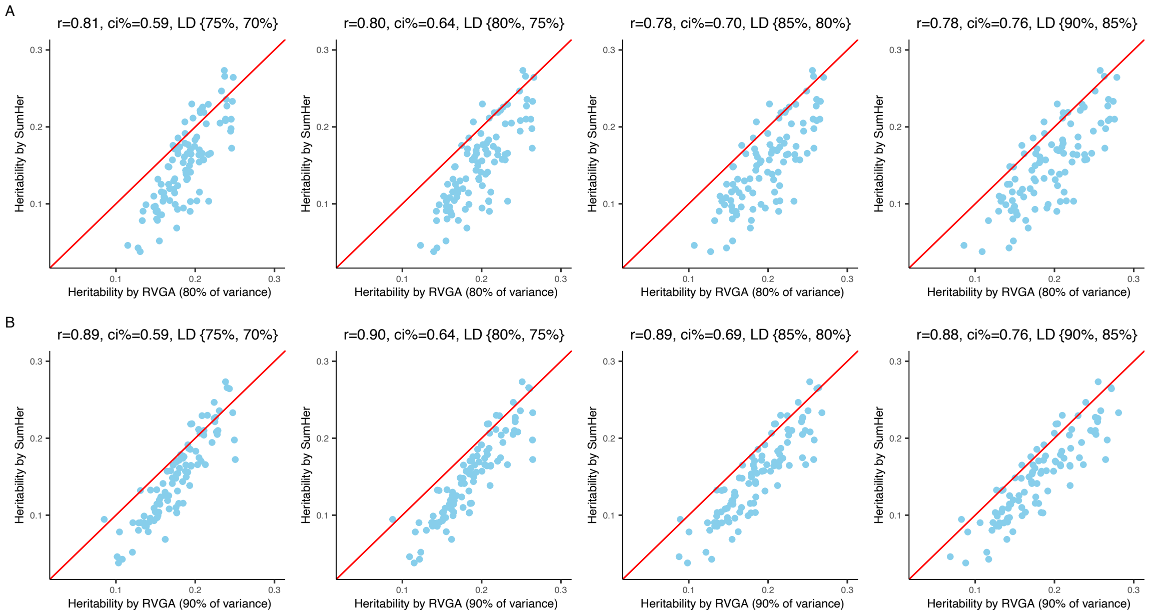

Figure S30: Sensitivity analysis of heritability using genotyped SNPs. We compared RVGA with SumHer, focusing on 100 randomly selected vertices from the left hippocampus. r: Pearson correlation coefficient. ci %: relative confidence interval width, defined as the mean standard error of RVGA estimates to that of SumHer estimates. A, Preserving 80% of variance. B, Preserving 90% of variance.

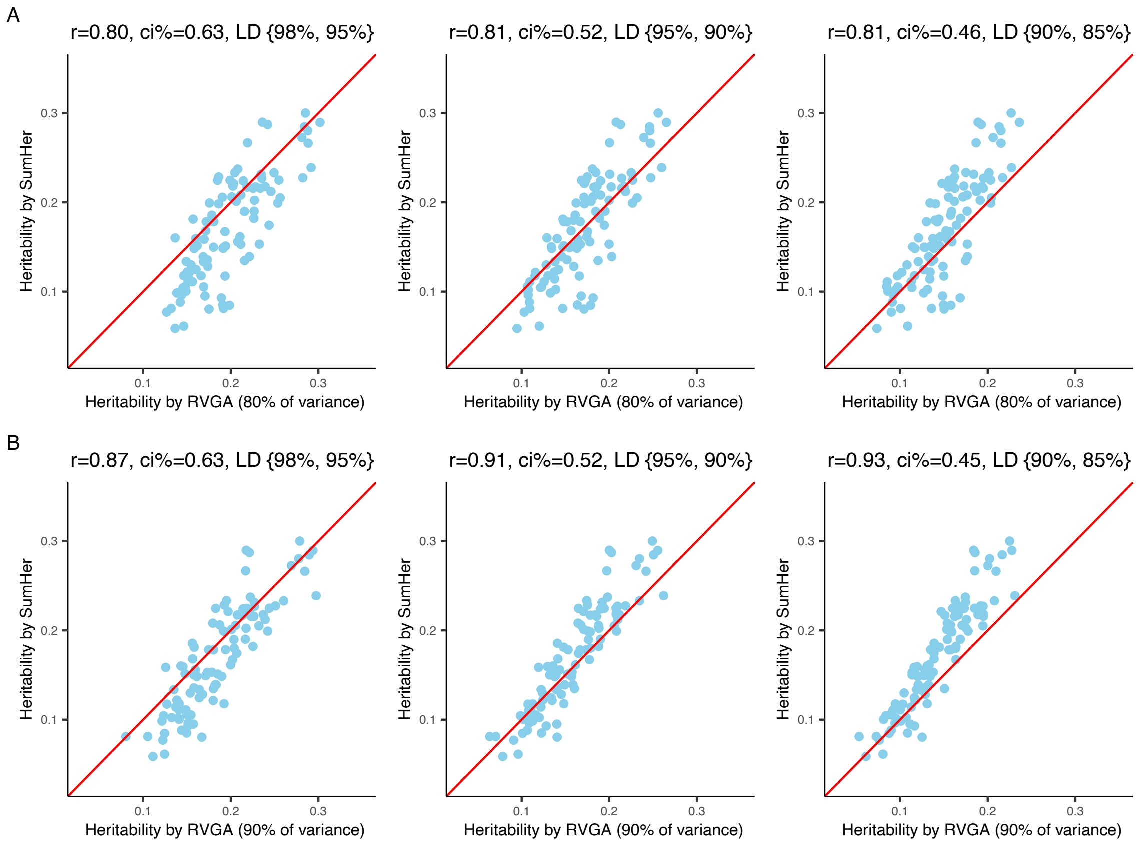

Figure S31: Sensitivity analysis of heritability using imputed HapMap3 SNPs. We compared RVGA with SumHer, focusing on 100 randomly selected vertices from the left hippocampus. r: Pearson correlation coefficient. ci %: relative confidence interval width, defined as the mean standard error of RVGA estimates to that of SumHer estimates. A, Preserving 80% of variance. B, Preserving 90% of variance.

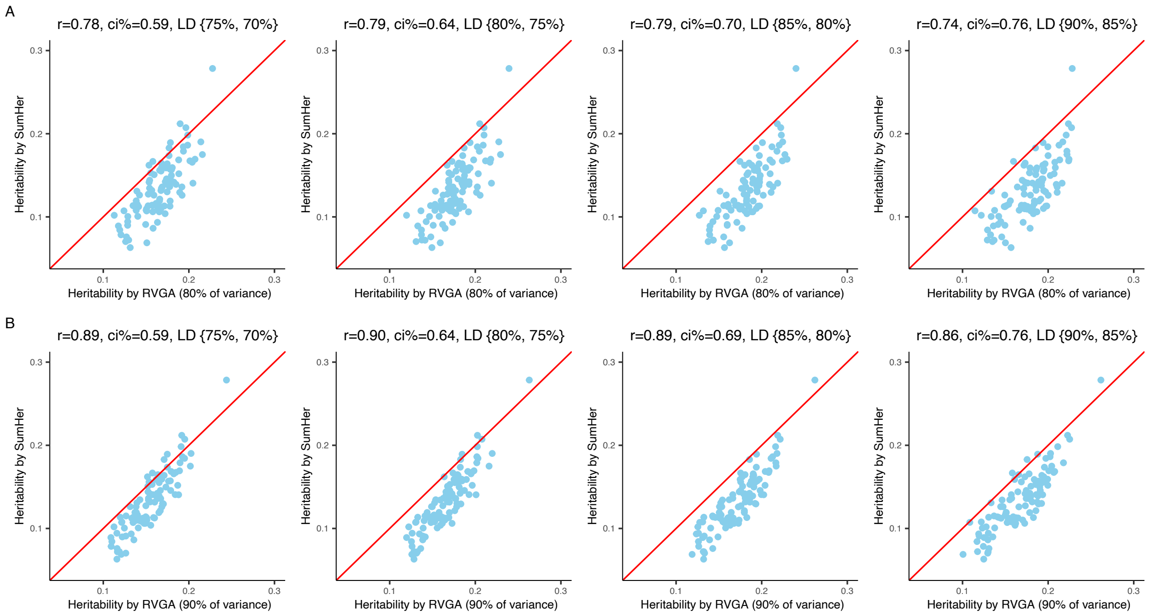

Figure S32: Sensitivity analysis of heritability using genotyped SNPs. We compared RVGA with SumHer, focusing on 100 randomly selected voxels from the superior fronto-occipital fasciculus. r: Pearson correlation coefficient. ci %: relative confidence interval width, defined as the mean standard error of RVGA estimates to that of SumHer estimates. A, Preserving 80% of variance. B, Preserving 90% of variance.

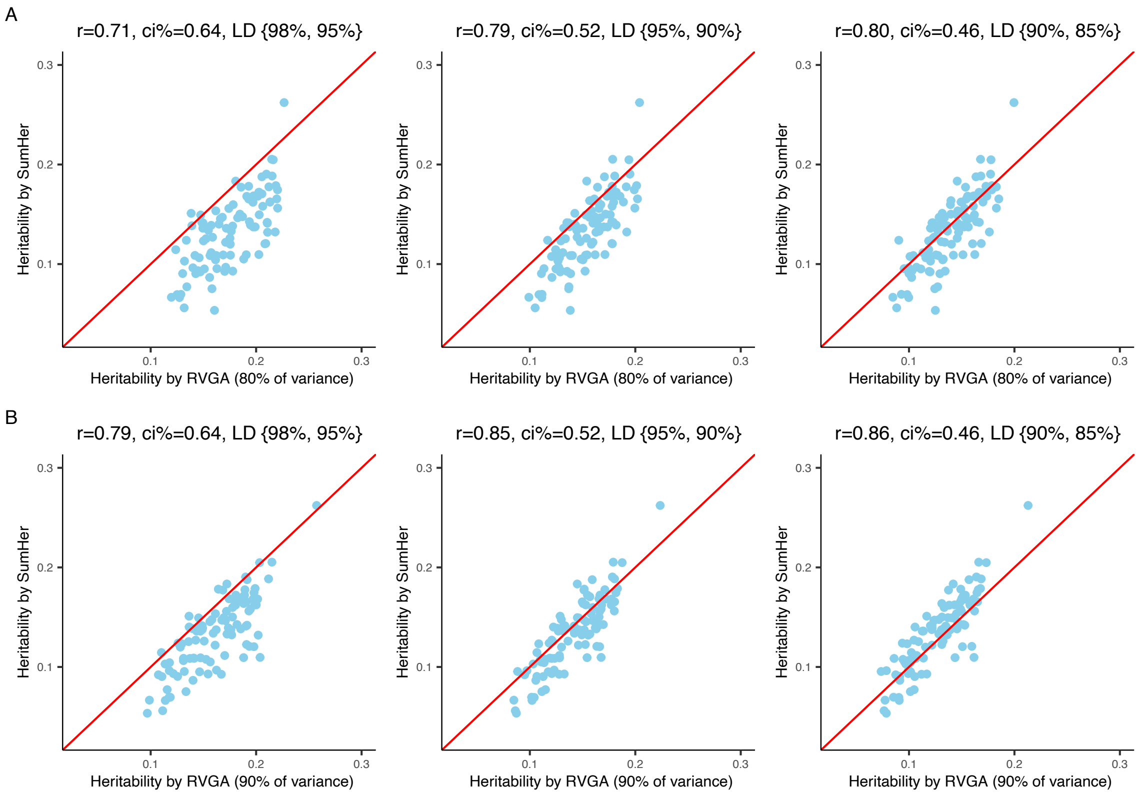

Figure S33: Sensitivity analysis of heritability using imputed HapMap3 SNPs. We compared RVGA with SumHer, focusing on 100 randomly selected voxels from the superior fronto-occipital fasciculus. r: Pearson correlation coefficient. ci %: relative confidence interval width, defined as the mean standard error of RVGA estimates to that of SumHer estimates. A, Preserving 80% of variance. B, Preserving 90% of variance.

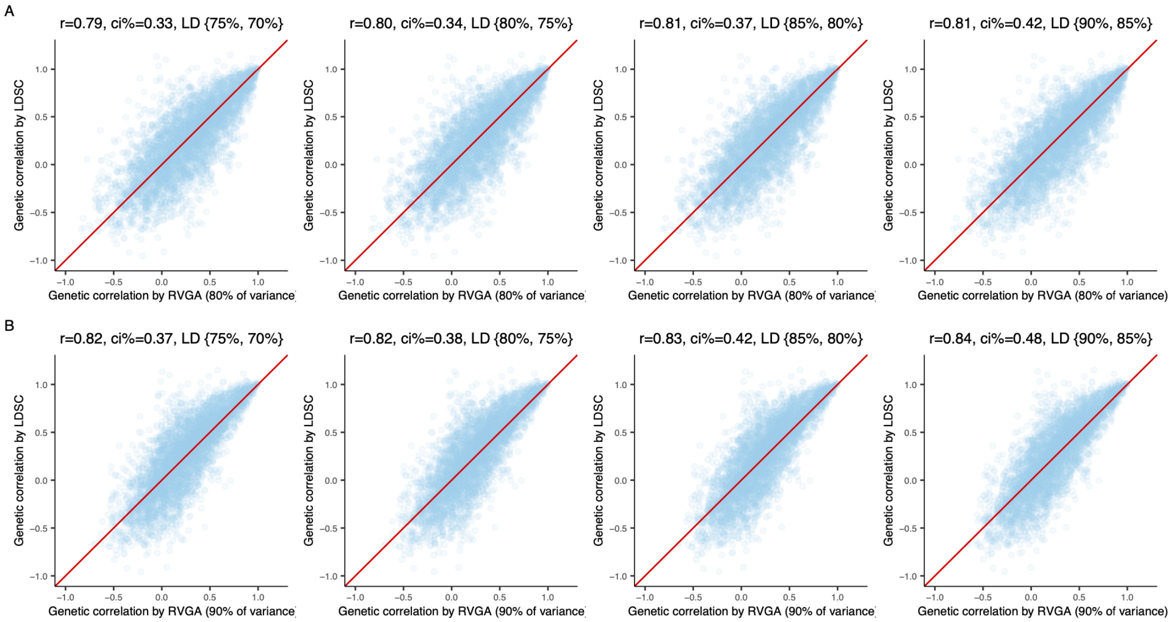

Figure S34: Sensitivity analysis of genetic correlation within images using genotyped SNPs. We compared RVGA with LDSC, focusing on 100 randomly selected vertices from the left hippocampus. r: Pearson correlation coefficient. ci %: relative confidence interval width, defined as the mean standard error of RVGA estimates to that of LDSC estimates. A, Preserving 80% of variance. B, Preserving 90% of variance.

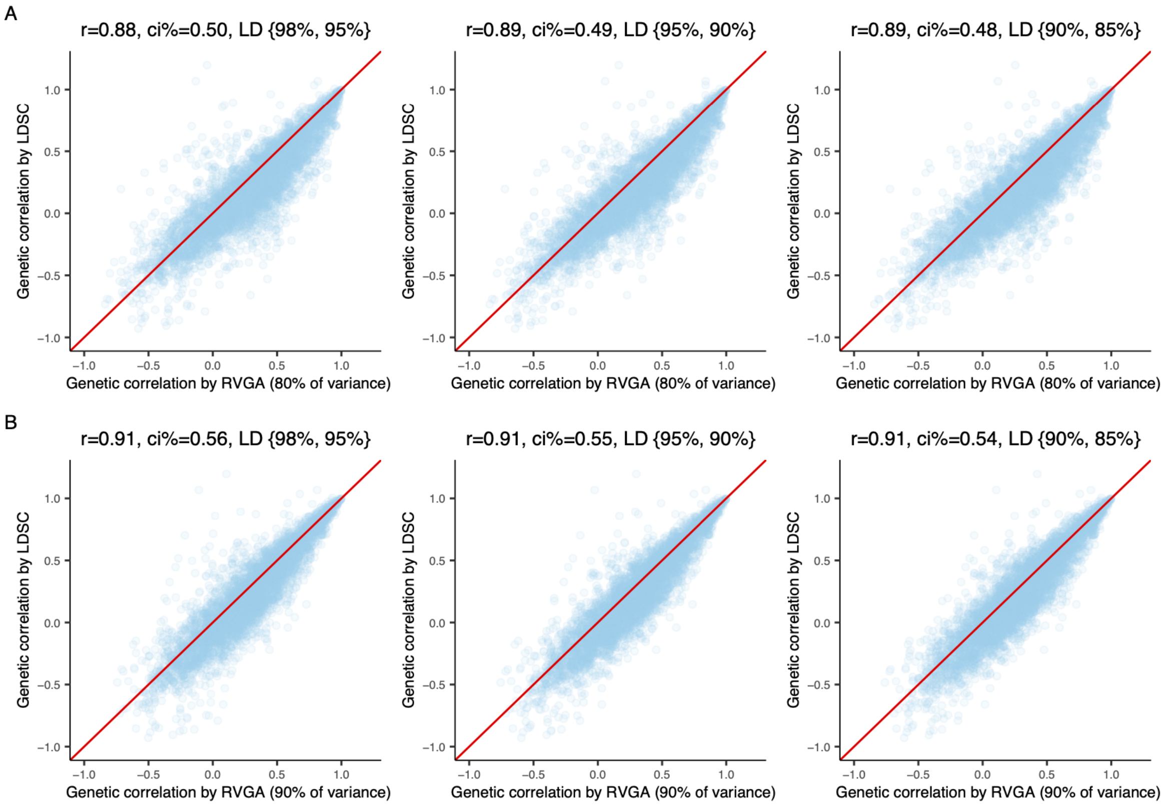

Figure S35: Sensitivity analysis of genetic correlation within images using imputed HapMap3 SNPs. We compared RVGA with LDSC, focusing on 100 randomly selected vertices from the left hippocampus. r: Pearson correlation coefficient. ci %: relative confidence interval width, defined as the mean standard error of RVGA estimates to that of LDSC estimates. A, Preserving 80% of variance. B, Preserving 90% of variance.

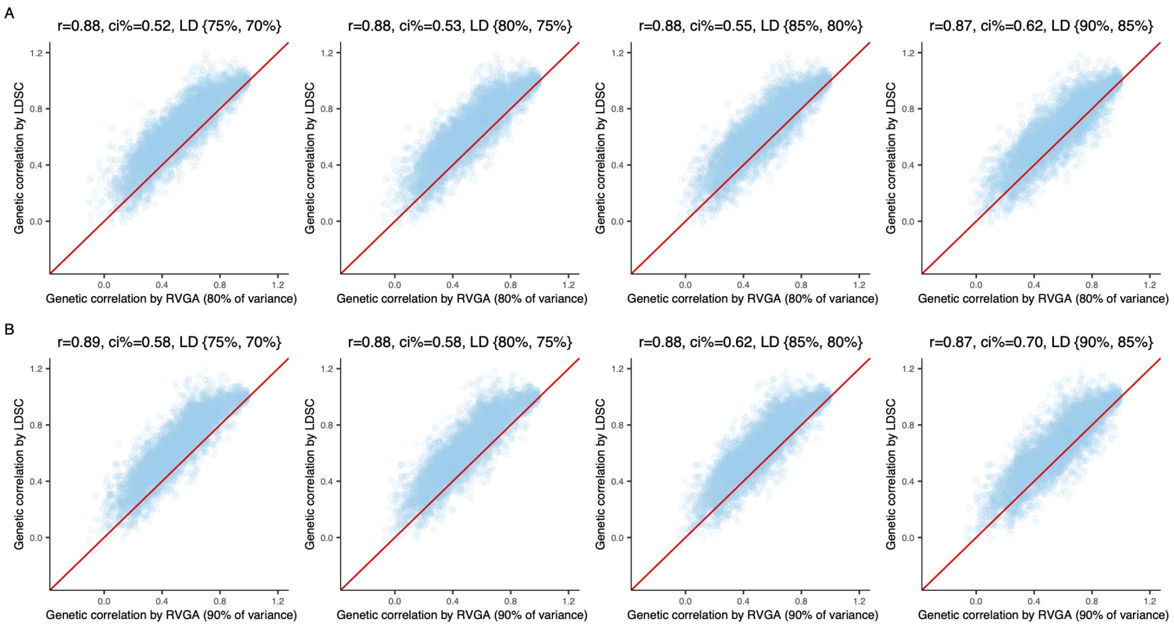

Figure S36: Sensitivity analysis of genetic correlation within images using genotyped SNPs. We compared RVGA with LDSC, focusing on 100 randomly selected voxels from the superior fronto-occipital fasciculus. r: Pearson correlation coefficient. ci %: relative confidence interval width, defined as the mean standard error of RVGA estimates to that of LDSC estimates. A, Preserving 80% of variance. B, Preserving 90% of variance.

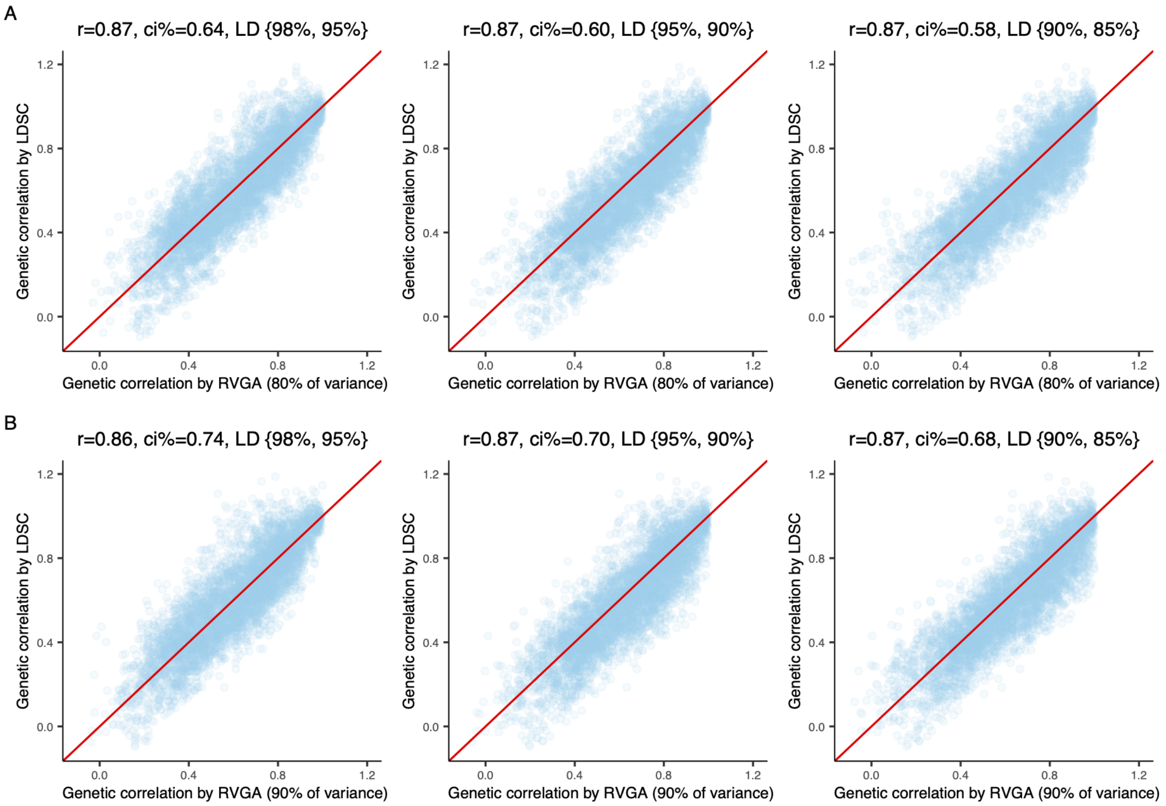

Figure S37: Sensitivity analysis of genetic correlation within images using imputed HapMap3 SNPs. We compared RVGA with LDSC, focusing on 100 randomly selected voxels from the superior fronto-occipital fasciculus. r: Pearson correlation coefficient. ci %: relative confidence interval width, defined as the mean standard error of RVGA estimates to that of LDSC estimates. A, Preserving 80% of variance. B, Preserving 90% of variance.

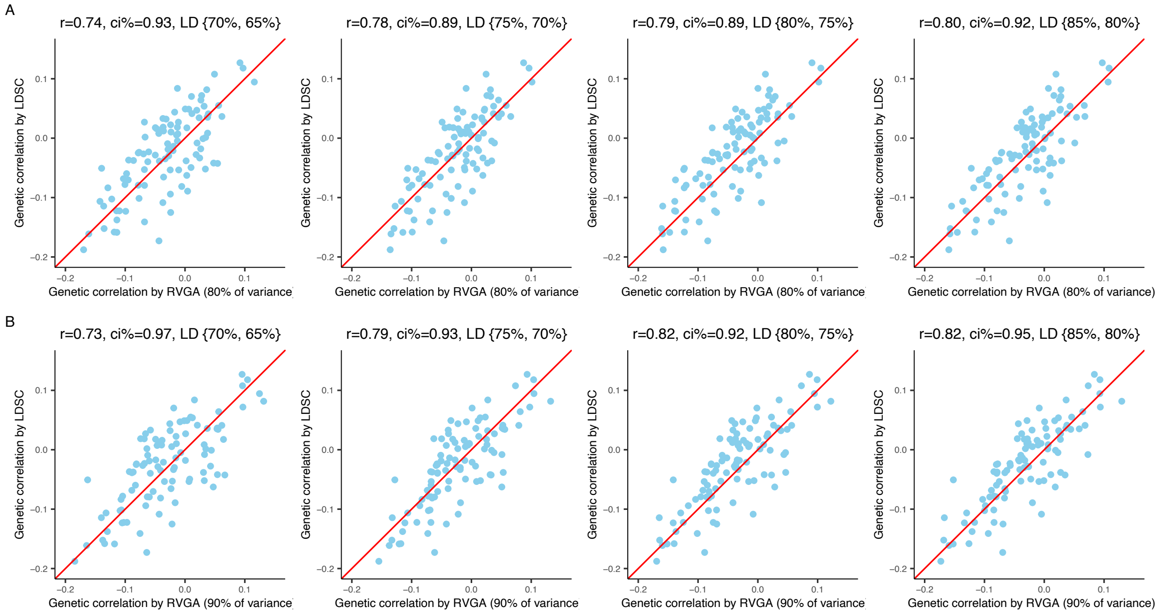

Figure S38: Sensitivity analysis of cross-trait genetic correlation between the left hippocampus and Alzheimer's disease using genotyped SNPs. We compared RVGA with LDSC, focusing on 100 randomly selected vertices from the left hippocampus. r: Pearson correlation coefficient. ci %: relative confidence interval width, defined as the mean standard error of RVGA estimates to that of LDSC estimates. A, Preserving 80% of variance. B, Preserving 90% of variance.

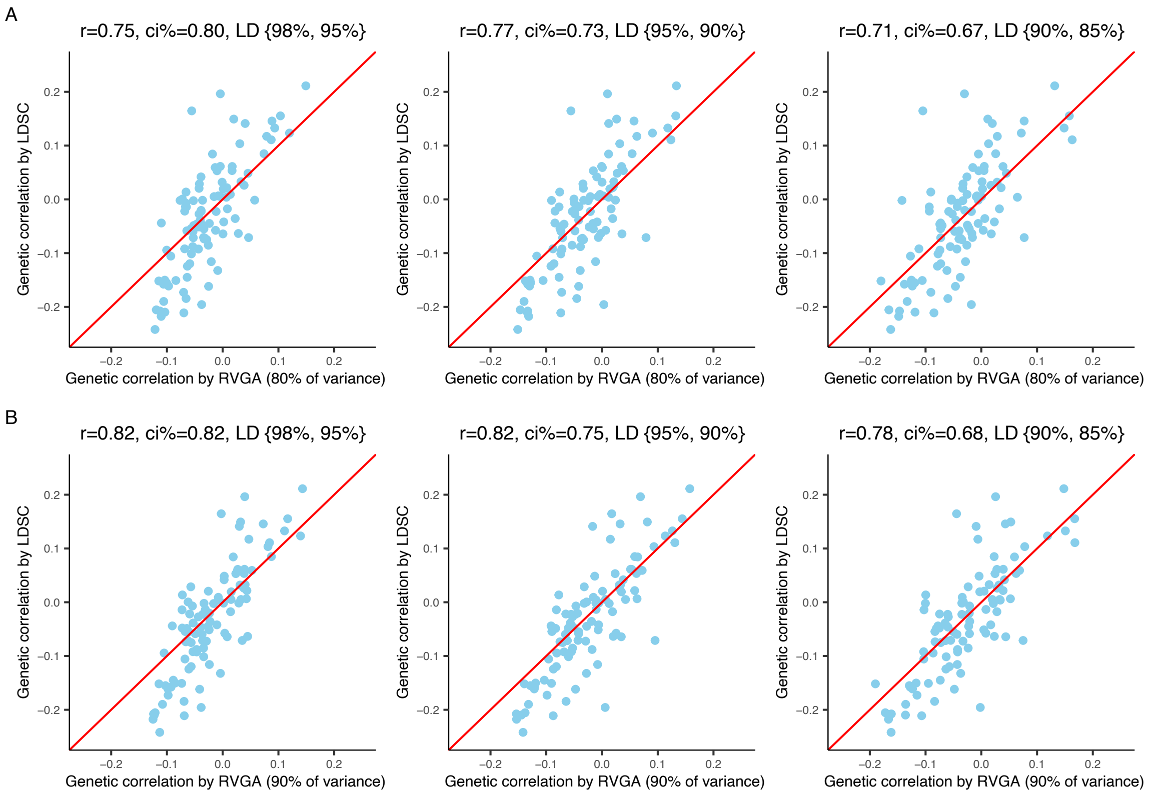

Figure S39: Sensitivity analysis of cross-trait genetic correlation between the left hippocampus and Alzheimer's disease using imputed HapMap3 SNPs. We compared RVGA with LDSC, focusing on 100 randomly selected vertices from the left hippocampus. r: Pearson correlation coefficient. ci %: relative confidence interval width, defined as the mean standard error of RVGA estimates to that of LDSC estimates. A, Preserving 80% of variance. B, Preserving 90% of variance.

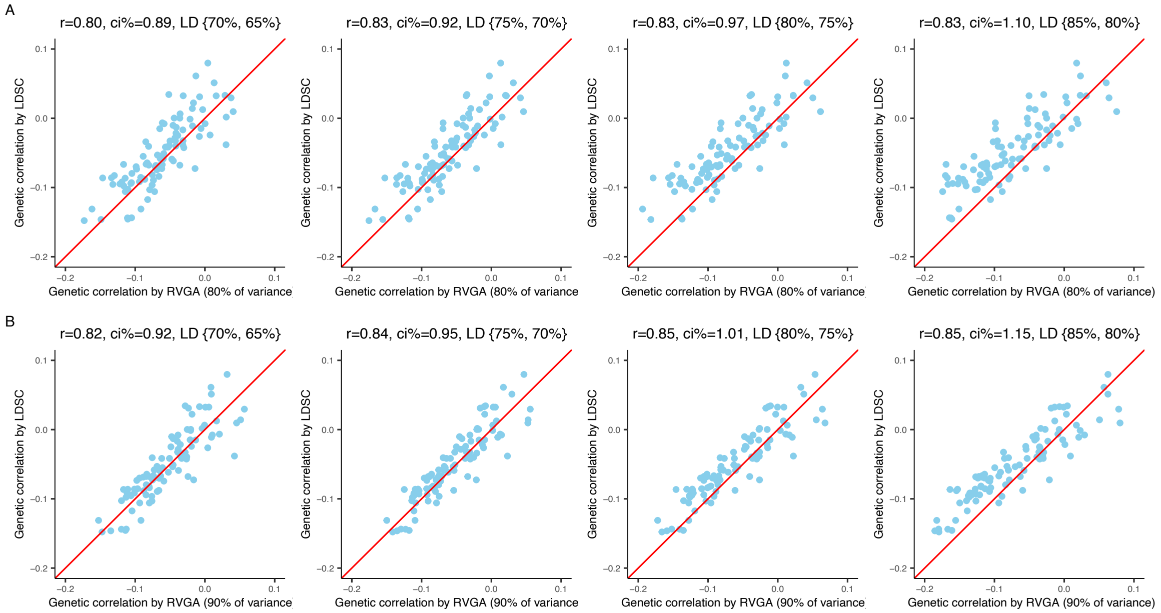

Figure S40: Sensitivity analysis of cross-trait genetic correlation between the left hippocampus and educational attainment using genotyped SNPs. We compared RVGA with LDSC, focusing on 100 randomly selected vertices from the left hippocampus. r: Pearson correlation coefficient. ci %: relative confidence interval width, defined as the mean standard error of RVGA estimates to that of LDSC estimates. A, Preserving 80% of variance. B, Preserving 90% of variance.

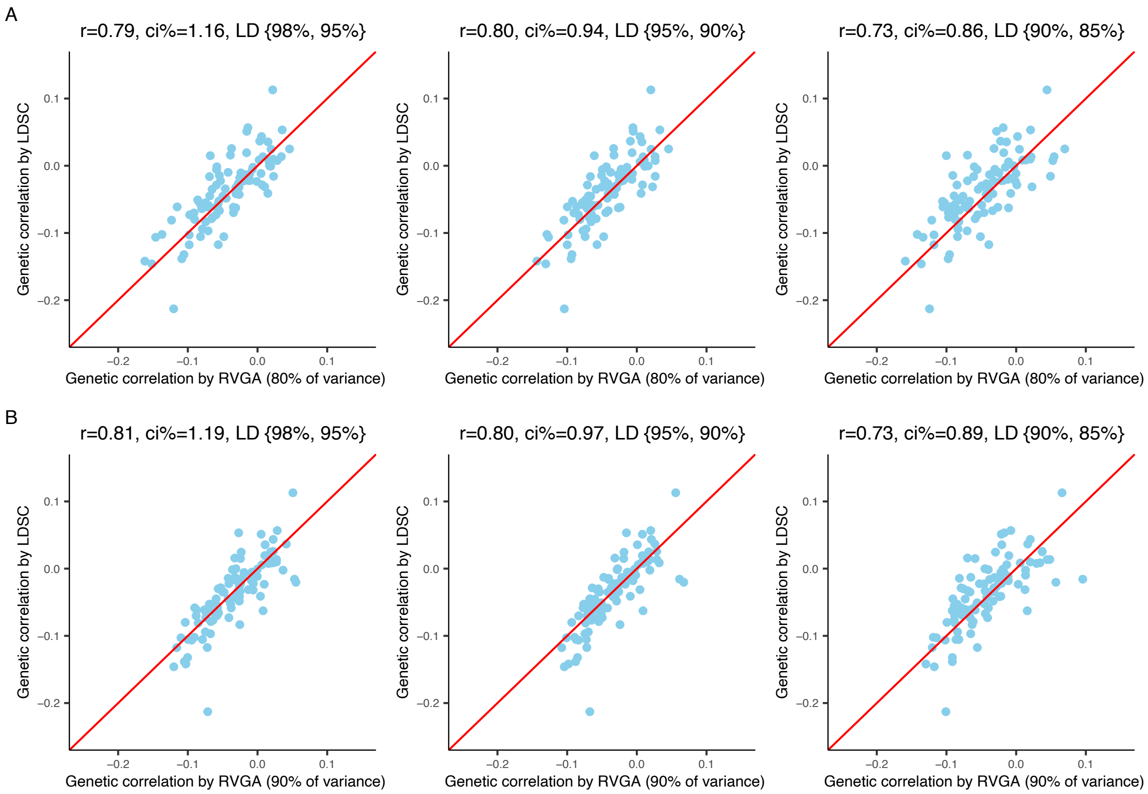

Figure S41: Sensitivity analysis of cross-trait genetic correlation between the left hippocampus and educational attainment using imputed HapMap3 SNPs. We compared RVGA with LDSC, focusing on 100 randomly selected vertices from the left hippocampus. r: Pearson correlation coefficient. ci %: relative confidence interval width, defined as the mean standard error of RVGA estimates to that of LDSC estimates. A, Preserving 80% of variance. B, Preserving 90% of variance.

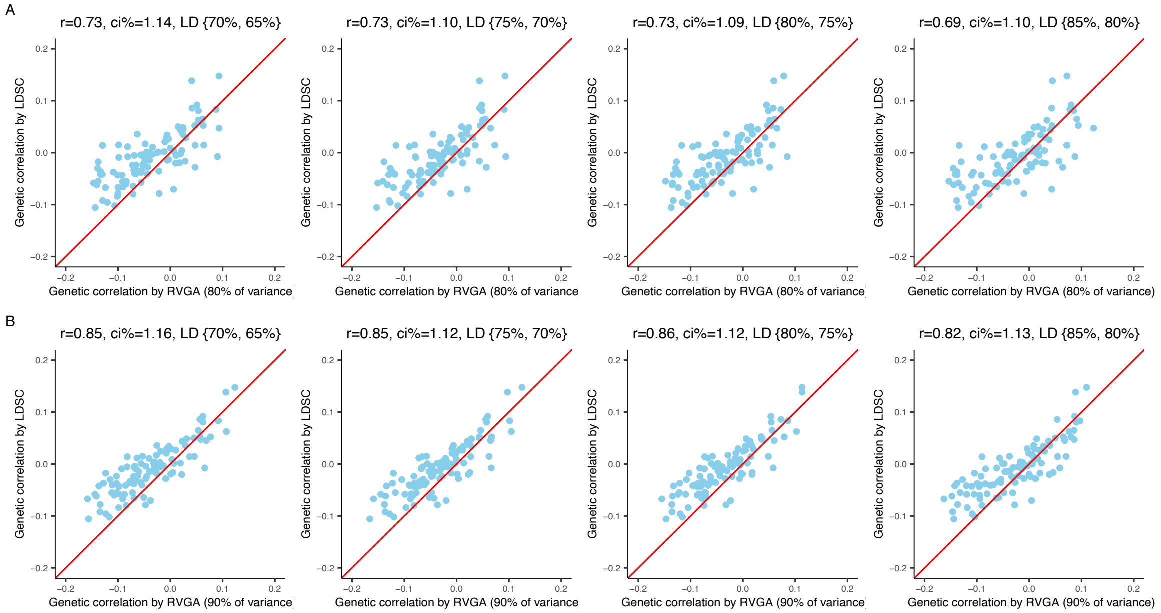

Figure S42: Sensitivity analysis of cross-trait genetic correlation between the superior fronto-occipital fasciculus and Alzheimer's disease using genotyped SNPs. We compared RVGA with LDSC, focusing on 100 randomly selected voxels from the superior fronto-occipital fasciculus. r: Pearson correlation coefficient. ci %: relative confidence interval width, defined as the mean standard error of RVGA estimates to that of LDSC estimates. A, Preserving 80% of variance. B, Preserving 90% of variance.

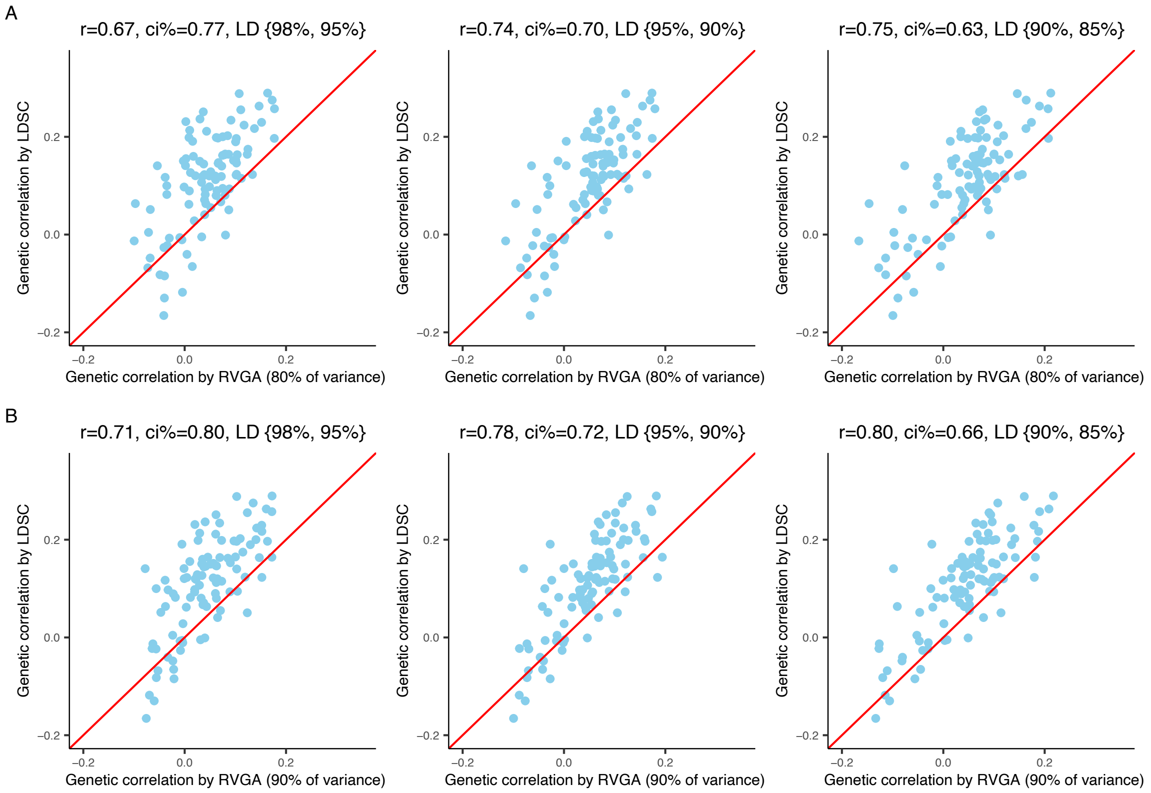

Figure S43: Sensitivity analysis of cross-trait genetic correlation between the superior fronto-occipital fasciculus and Alzheimer's disease using imputed HapMap3 SNPs. We compared RVGA with LDSC, focusing on 100 randomly selected voxels from the superior fronto-occipital fasciculus. r: Pearson correlation coefficient. ci %: relative confidence interval width, defined as the mean standard error of RVGA estimates to that of LDSC estimates. A, Preserving 80% of variance. B, Preserving 90% of variance.

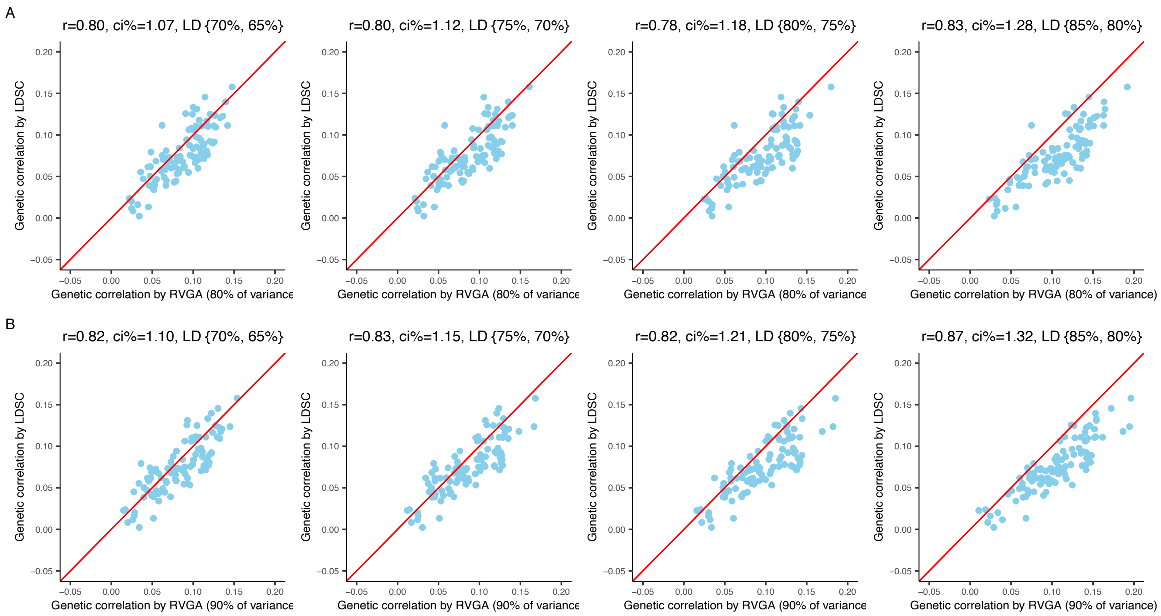

Figure S44: Sensitivity analysis of cross-trait genetic correlation between the superior fronto-occipital fasciculus and educational attainment using genotyped SNPs. We compared RVGA with LDSC, focusing on 100 randomly selected voxels from the superior fronto-occipital fasciculus. r: Pearson correlation coefficient. ci %: relative confidence interval width, defined as the mean standard error of RVGA estimates to that of LDSC estimates. A, Preserving 80% of variance. B, Preserving 90% of variance.

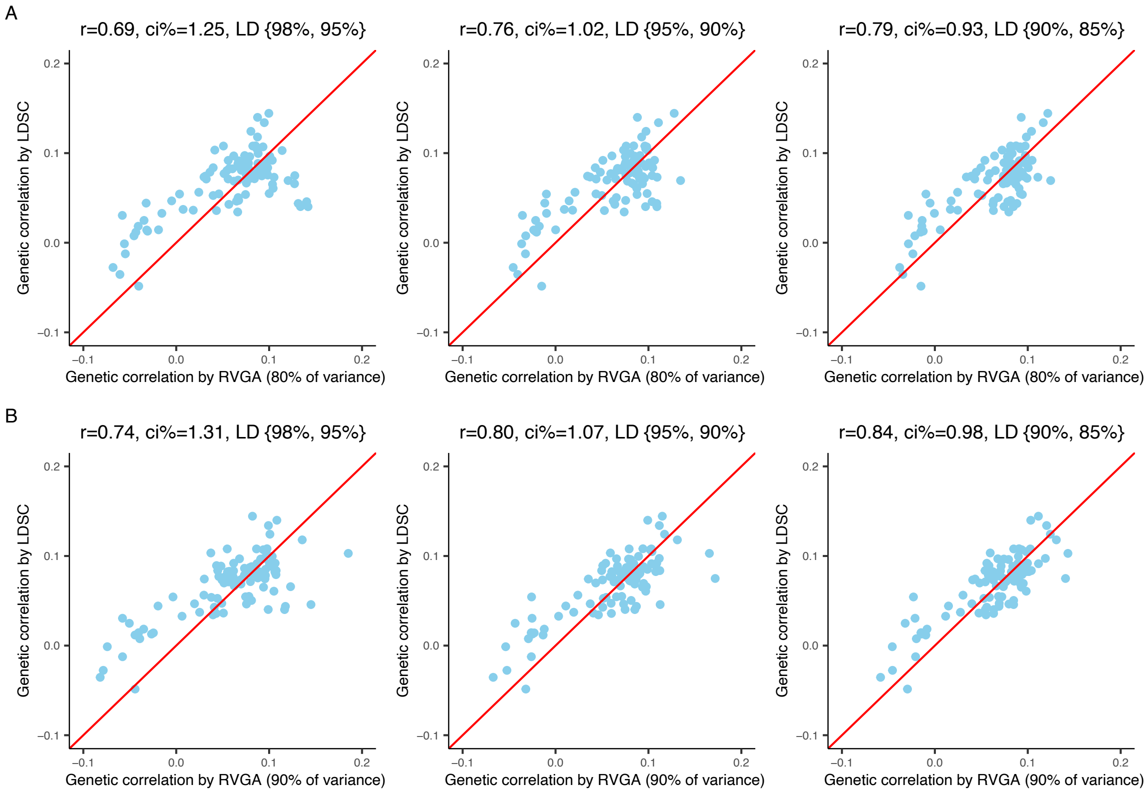

Figure S45: Sensitivity analysis of cross-trait genetic correlation between the superior fronto-occipital fasciculus and educational attainment using imputed HapMap3 SNPs. We compared RVGA with LDSC, focusing on 100 randomly selected voxels from the superior fronto-occipital fasciculus. r: Pearson correlation coefficient. ci %: relative confidence interval width, defined as the mean standard error of RVGA estimates to that of LDSC estimates. A, Preserving 80% of variance. B, Preserving 90% of variance.

### 3 Supplementary Tables

All tables are in a separate .xlsx file.

Table S1: Abbreviations and full names for white matter tracts.

Table S2: The number of LDRs for preserving a specific proportion of variance. Effective number: the number of approximately independent voxels (vertices) in the ROI. Threshold: a Bonferroni genome-wide significance threshold using the effective number. Hippocampus and white matter tracts are considered separately. Refer to Table S1 for the full name of white matter tracts.

Table S3: The identified genetic loci for FA of white matter tracts in the discovery study (n = 33,324). Each locus was constructed by grouping significant voxel-variant pairs ($P < 1.91 \times 10^{-10}$) within an ROI such that the longest distance from any variant to the most significant variant was less than 0.25 cM. Each voxel-variant pair can be included in one and only one locus. A previous locus was replicated by our study if the most significant variant in that locus was within 0.25 cM from any of the most significant variants in our loci. INDEX: the one-based index of voxel in the image. P: $-\log_{10}$ $p$-values. otherSNPs: other significant SNPs within the same locus, may be associated with other voxels in the image. CM: genetic distance (1000 Genome, GRCh37). Replicated_Zhao_Smith: if the locus replicated the locus in Zhao et al (2021) and/or Smith et al (2021). Replicated: if the locus replicated by using UKB phases 4 to 6 unrelated white subjects (n = 20,130), adjusting for the number of significant loci in the discovery study ($P < 0.05/526$). Replicated_rigorous: if the locus replicated using a more rigorous threshold accounting for the number of significant loci and the effective number of independent voxels ($P < 0.05/526/261.8$).

Table S4: The identified genetic loci for hippocampus shape in the discovery study (n = 33,324). Each locus was constructed by grouping significant vertex-variant pairs ($P < 4.94 \times 10^{-9}$) within an ROI such that the longest distance from any variant to the most significant variant was less than 0.25 cM. Each vertex-variant pair can be included in one and only one locus. A previous locus was replicated by our study if the most significant variant in that locus was within 0.25 cM from any of the most significant variants in our loci. INDEX: the one-based index of vertex in the image. P: $-\log_{10}$ $p$-values. OtherSNPs: other significant SNPs within the same locus, may be associated with other vertices in the image. CM: genetic distance (1000 Genome, GRCh37). Replicated_cataglog: if the locus replicated loci in our previous study and/or any hippocampus-related locus reported on NHGRI-EBI GWAS catalog. Replicated: if the locus replicated by using UKB phases 4 to 6 unrelated white subjects (n = 20,130), adjusting for the number of significant loci in the discovery study ($P < 0.05/72$). Replicated_rigorous: if the locus replicated using a more rigorous threshold accounting for the number of significant loci and the effective number of independent vertices ($P < 0.05/72/10.1$).

Table S5: The identified genetic loci for FA of white matter tracts in the discovery phase reproduced in the replication study (n = 20,130). We extracted all the variants in significant loci ($P < 1.91 \times 10^{-10}$) in the discovery phase and conducted association analysis for those variants across all voxels in the replication study, saving associations with a $p$-value less than 0.05/526. Each locus was constructed by grouping significant voxel-variant pairs within an ROI such that the longest distance from any variant to the most significant variant was less than 0.25 cM. Each voxel-variant pair can be included in one and only one locus. INDEX: the one-based index of voxel in the image. P: -log10 $p$-values. otherSNPs: other significant SNPs within the same locus, may be associated with other voxels in the image. CM: genetic distance (1000 Genome, GRCh37).

Table S6: The identified genetic loci for hippocampus shape in the discovery phase reproduced in the replication study (n = 20,130). We extracted all the variants in significant loci ($P < 4.94 \times 10^{-9}$) in the discovery phase and conducted association analysis for those variants across all vertices in the replication study, saving associations with a $p$-value less than 0.05/72. Each locus was constructed by grouping significant vertex-variant pairs ($P < 0.05/72$) within an ROI such that the longest distance from any variant to the most significant variant was less than 0.25 cM. Each vertex-variant pair can be included in one and only one locus. INDEX: the one-based index of vertex in the image. P: $-\log_{10}$ $p$-values. otherSNPs: other significant SNPs within the same locus, may be associated with other vertices in the image. CM: genetic distance (1000 Genome, GRCh37).

Table S7: Heritability analysis for the hippocampus and white matter tracts. The heritability was estimated using Hapmap3 SNPs. Standard deviation: standard deviation of all estimates across voxels. Mean standard error: average of standard error estimate of all voxels.

Table S8: Genetic correlation analysis within the hippocampus and white matter tracts. The genetic correlation was estimated using Hapmap3 SNPs. Standard deviation: standard deviation of all genetic correlation estimates across voxels. Mean standard error: average of standard error estimate of all voxel pairs.

Table S9: Summary statistics of brain-related phenotypes used in the study.

Table S10: Cross-trait genetic correlation analysis between the hippocampus shape and 14 brain-related phenotypes. The genetic correlation was estimated using Hapmap3 SNPs. Global significant results (FDR $< 0.05$) are highlighted. Standard deviation: standard deviation of genetic correlation estimates aross all vertices. Mean standard error: average of standard error estimate of all vertices. Cauchy $p$-value: a $p$-value generated by meta-analyzing the $p$-value of each vertex using the Cauchy combination strategy (Liu et al. 2019).

Table S11: Cross-trait genetic correlation analysis between FA of white matter tracts and 14 brain-related phenotypes. The genetic correlation was estimated using Hapmap3 SNPs. The full name of tracts can be found the Table S1. Global significant results (FDR $< 0.05$) are highlighted. Standard deviation: standard deviation of genetic correlation estimates aross all voxels. Mean standard error: average of standard error estimate of all voxels. Cauchy $p$-value: a $p$-value generated by meta-analyzing the $p$-value of each voxel using the Cauchy combination strategy (Liu et al. 2019). Replicated: if the significant signal replicates the results in Table S15 in Zhao et al. (2021).

Table S12: The identified genetic loci for cortical surface curvature in the discovery study (n = 15,752). Each locus was constructed by grouping significant vertex-variant pairs ($P < 3.15 \times 10^{-11}$) within the whole cortical surface such that the longest distance from any variant to the most significant variant was less than 0.25 cM. Each vertex-variant pair can be included in one and only one locus. A locus was replicated by the replication study if the most significant variant in the locus was within 0.25 cM from any of the most significant variants in loci from the replication study. INDEX: the one-based index of vertex in the image. P: -log10 $p$-values. OtherSNPs: other significant SNPs within the same locus, may be associated with other vertices in the image. CM: genetic distance (1000 Genome, GRCh37). Replicated: if the locus replicated by using UKB phase 1 and 2 unrelated white subjects (n = 12,431), adjusting for the number of significant loci in the discovery study ($P < 0.05/35$). Replicated_rigorous: if the locus replicated using a more rigorous threshold accounting for the number of significant loci and the effective number of independent vertices ($P < 0.05/35/1585.3$).

Table S13: The identified genetic loci for curtical surface curvature in the discovery phase reproduced in the replication study (n = 12,431). We extracted variants with at least one nominally significant association ($P < 3.15 \times 10^{-11}$) in the discovery phase and conducted association analysis for those variants across all vertices in the replication study, saving associations with a $p$-value less than 0.05/35. Each vertex-variant pair can be included in one and only one locus. INDEX: the one-based index of vertex in the image. P: -log10 $p$-values. otherSNPs: other significant SNPs within the same locus, may be associated with other vertices in the image. CM: genetic distance (1000 Genome, GRCh37).

Table S14: Cross-trait genetic correlation analysis between the cortical surface curvature and 14 brain-related phenotypes. The genetic correlation was estimated using Hapmap3 SNPs. Standard deviation: standard deviation of genetic correlation estimates aross all vertices. Mean standard error: average of standard error estimate of all vertices. Cauchy $p$-value: a $p$-value generated by meta-analyzing the $p$-value of each vertex using the Cauchy combination strategy (Liu et al. 2019).

Table S15: A comparison of voxel-level GWAS using FPCA and PCA. UKB phases 1 to 3 unrelated subjects of European ancestry (n = 33,324) were employed in the investigation. We selected the same number of LDRs for each ROI in FPCA and PCA. We compared the number of significant variant-voxel associations, and then aggregated them into loci, removing suspicious loci with a single variant-voxel association. Two loci are overlapping if the distance between index SNPs is less than 0.25 cM.
